## Supplementary Figures and Tables for "Clonal chromosomal mosaicism and loss of chromosome Y in men are risk factors for SARS-CoV-2 vulnerability in the elderly"

Supplementary material

Luis A. Pérez-Jurado; Alejandro Cáceres; Tonu Esko; Miguel López de Heredia; Inés Quintela; Raquel Cruz; Pablo Lapunzina; Ángel Carracedo; SCOURGE Cohort Group; Juan R. González

### Contents

**Supplementary tables** **1**

**Supplementary figures** **10**

### Supplementary tables

Table S1: Patients with CMEs.

[See supplementary Excel File]

Table S2: Patients with LOY/XCM.

[See supplementary Excel File]

Table S3: Individuals with germinal line.

[See supplementary Excel File]

| | normal N=502 | LOY N=28 | p.overall |
|---|---|---|---|
| WBC | 6.18 [5.24;7.27] | 6.49 [5.25;7.65] | 0.443 |
| RBC | 5.07 [4.82;5.30] | 4.72 [4.50;5.13] | 0.004 |
| PLT | 217 [190;248] | 239 [219;270] | 0.015 |
| EO% | 2.30 [1.50;3.55] | 2.60 [1.60;4.00] | 0.426 |
| BASO% | 0.40 [0.30;0.60] | 0.30 [0.20;0.43] | 0.027 |
| MONO% | 8.50 [7.00;10.1] | 8.70 [7.38;9.62] | 0.720 |
| LYMPH% | 30.6 (7.53) | 27.6 (8.83) | 0.088 |
| NEUT% | 57.4 (8.73) | 59.8 (9.38) | 0.184 |
| EO (total) | 0.15 [0.09;0.23] | 0.15 [0.11;0.28] | 0.300 |
| BASO (total) | 0.03 [0.02;0.04] | 0.02 [0.02;0.02] | 0.016 |
| MONO (total) | 0.52 [0.43;0.65] | 0.58 [0.46;0.75] | 0.146 |
| LYMPH (total) | 1.83 [1.53;2.28] | 1.66 [1.34;2.14] | 0.179 |
| NEUT (total) | 3.46 [2.81;4.39] | 3.99 [2.77;4.90] | 0.247 |
| RDW-SD | 40.4 [38.9;42.4] | 42.5 [41.5;46.0] | 0.007 |
| HGB | 152 [146;159] | 151 [138;156] | 0.401 |
| PDW | 13.1 [11.8;14.4] | 13.6 [12.2;15.2] | 0.596 |
| MPV | 10.9 [10.2;11.5] | 11.1 [10.5;11.6] | 0.523 |
| LCR | 31.8 [26.7;37.2] | 32.4 [29.0;38.5] | 0.587 |
| PCT | 0.24 [0.21;0.27] | 0.25 [0.24;0.27] | 0.096 |
| Hb | 151 (14.5) | 147 (13.2) | 0.247 |
| Hct | 45.0 [43.0;47.0] | 44.0 [42.0;47.0] | 0.454 |
| MCV | 88.0 [85.6;91.2] | 90.6 [89.0;94.6] | <0.001 |
| MCH | 30.1 [29.2;30.9] | 30.2 [29.5;30.9] | 0.384 |
| MCHC | 340 [335;346] | 332 [325;337] | <0.001 |
| RDW-CV | 12.7 [12.3;13.2] | 13.1 [12.7;13.4] | 0.006 |
| Alb | 47.0 [45.0;49.0] | 44.0 [42.0;45.2] | <0.001 |
| ALAT | 23.0 [17.0;32.8] | 25.0 [15.5;36.2] | 0.615 |
| ALP | 66.0 [56.0;80.8] | 73.5 [59.5;81.5] | 0.156 |
| ASAT | 24.0 [20.0;28.0] | 24.0 [18.0;30.2] | 0.778 |
| Bil | 9.00 [6.00;12.0] | 8.00 [7.00;13.2] | 0.951 |
| GGT | 21.0 [15.0;31.0] | 22.5 [15.8;30.0] | 0.738 |
| Hcy | 10.6 [8.43;13.6] | 14.8 [11.6;18.3] | <0.001 |
| Chol | 5.20 [4.40;6.10] | 5.20 [4.80;5.98] | 0.396 |
| HDL-Chol | 1.31 [1.09;1.55] | 1.39 [1.12;1.76] | 0.135 |
| LDL-Chol | 3.33 [2.71;4.25] | 3.25 [2.93;3.73] | 0.912 |
| Trigl | 1.41 [0.98;2.13] | 1.22 [1.01;1.67] | 0.120 |
| Crea | 77.0 [70.0;86.0] | 81.5 [69.8;89.2] | 0.253 |
| UA | 335 [295;383] | 350 [325;388] | 0.182 |
| Urea | 5.40 [4.50;6.40] | 6.15 [5.20;7.25] | 0.002 |
| Fer | 107 [59.0;179] | 103 [54.8;199] | 0.840 |
| Fe | 18.1 [14.1;23.0] | 19.2 [15.0;23.7] | 0.591 |
| Transf | 2.62 [2.38;2.92] | 2.54 [2.32;3.02] | 0.711 |
| Transf-sR | 2.90 [2.40;3.50] | 2.90 [2.48;3.60] | 0.928 |
| Fol | 15.9 [12.8;20.3] | 15.8 [13.8;19.9] | 0.850 |
| B12 | 301 [232;378] | 262 [226;312] | 0.067 |
| TSH | 1.07 [0.75;1.53] | 1.49 [1.09;1.85] | 0.007 |
| CRP | 1.57 [0.97;3.04] | 2.23 [0.87;3.11] | 0.697 |
| Gluc | 5.40 [5.00;6.00] | 5.30 [4.80;5.90] | 0.659 |
| CysC | 1.04 [0.92;1.14] | 1.11 [1.06;1.13] | 0.050 |
| EPO | 9.59 [7.24;11.8] | 9.56 [7.40;10.4] | 0.836 |
| Testo | 15.2 [10.2;20.7] | 14.4 [10.4;16.7] | 0.585 |

*continued on next page*

Table S4 – *continued from previous page*

| | normal N=502 | LOY N=28 | p.overall |
|---|---|---|---|
| Ins | 7.80 [4.80;15.0] | 9.60 [5.20;13.7] | 0.797 |

Table S4: Comparison of clinical data between normal and LOY individuals

| logFC | CI.L | CI.R | AveExpr | t | P.Value | adj.P.Val | B | SE | Chromosome | Symbol |
|---|---|---|---|---|---|---|---|---|---|---|
| 3 | 2.1 | 3.8 | -0.32 | 7.1 | $4.6 \times 10^{-9}$ | 0.00022 | 5.3 | 0.23 | 12 | VWF |
| -2.5 | -3.3 | -1.7 | -0.24 | -6.4 | $5.7 \times 10^{-8}$ | 0.0014 | 4 | 0.15 | Y | CSF2RA |
| 2.3 | 1.5 | 3.1 | 0.25 | 5.5 | $1.3 \times 10^{-6}$ | 0.021 | 2.3 | 0.26 | 20 | MYL9 |
| -2.2 | -3 | -1.3 | -0.22 | -5.4 | $2.3 \times 10^{-6}$ | 0.028 | 2 | 0.21 | Y | CSF2RA |
| 0.71 | 0.43 | 0.98 | -0.017 | 5.2 | $3.9 \times 10^{-6}$ | 0.038 | 1.7 | 0.17 | 2 | SPC25 |
| 1.3 | 0.74 | 1.8 | 0.17 | 4.9 | $1.2 \times 10^{-5}$ | 0.099 | 1 | 0.18 | X | BEND2 |
| 2.5 | 1.4 | 3.5 | 0.089 | 4.7 | $2.1 \times 10^{-5}$ | 0.14 | 0.72 | 0.18 | 17 | ITGA2B |
| 3.3 | 1.9 | 4.8 | 0.11 | 4.7 | $2.4 \times 10^{-5}$ | 0.14 | 0.64 | 0.29 | 4 | PPBP |
| 1.1 | 0.6 | 1.5 | -0.18 | 4.6 | $2.6 \times 10^{-5}$ | 0.14 | 0.6 | 0.19 | 14 | SYNE3 |
| 0.71 | 0.39 | 1 | -0.062 | 4.5 | $4.4 \times 10^{-5}$ | 0.18 | 0.29 | 0.19 | 12 | ANKS1B |
| -1 | -1.5 | -0.55 | 0.02 | -4.5 | $4.5 \times 10^{-5}$ | 0.18 | 0.28 | 0.23 | 11 | POU2AF1 |
| 0.69 | 0.37 | 1 | -0.11 | 4.4 | $6.8 \times 10^{-5}$ | 0.26 | 0.041 | 0.28 | X | GAGE4 |
| -1.6 | -2.4 | -0.87 | -0.034 | -4.3 | $8.2 \times 10^{-5}$ | 0.26 | -0.07 | 0.2 | 17 | KRT23 |
| -0.8 | -1.2 | -0.43 | 0.032 | -4.3 | $8.3 \times 10^{-5}$ | 0.26 | -0.073 | 0.2 | 8 | ENPP2 |
| 0.75 | 0.4 | 1.1 | -0.14 | 4.3 | $8.9 \times 10^{-5}$ | 0.26 | -0.11 | 0.22 | 16 | CDH8 |
| 0.79 | 0.42 | 1.2 | -0.056 | 4.3 | $9.2 \times 10^{-5}$ | 0.26 | -0.14 | 0.18 | 15 | TMOD3 |
| 2.3 | 1.2 | 3.4 | 0.04 | 4.2 | 0.0001 | 0.27 | -0.21 | 0.37 | 11 | JAM3 |
| -0.92 | -1.4 | -0.47 | -0.12 | -4.1 | 0.00017 | 0.41 | -0.49 | 0.19 | Y | SFRS17A |
| -1 | -1.5 | -0.52 | -0.1 | -4 | 0.00019 | 0.44 | -0.56 | 0.17 | 2 | WIPF1 |
| 1.7 | 0.83 | 2.5 | 0.34 | 4 | 0.00023 | 0.48 | -0.68 | 0.3 | 18 | GTSCR1 |
| 1.1 | 0.55 | 1.7 | -0.14 | 4 | 0.00024 | 0.48 | -0.71 | 0.22 | 1 | ADAM15 |
| -1.4 | -2.1 | -0.7 | 0.25 | -4 | 0.00024 | 0.48 | -0.71 | 0.24 | Y | TMSB4Y |
| 1.5 | 0.75 | 2.3 | 0.054 | 4 | 0.00025 | 0.48 | -0.71 | 0.2 | 4 | GUCY1A3 |
| 0.78 | 0.38 | 1.2 | -0.033 | 3.9 | 0.00026 | 0.5 | -0.76 | 0.22 | 5 | PIK3R1 |
| 0.87 | 0.42 | 1.3 | 0.035 | 3.9 | 0.00029 | 0.5 | -0.81 | 0.22 | 17 | MEOX1 |
| 1 | 0.5 | 1.6 | 0.012 | 3.9 | 0.00031 | 0.5 | -0.85 | 0.19 | 15 | ACSBG1 |
| 0.92 | 0.44 | 1.4 | -0.04 | 3.8 | 0.00035 | 0.5 | -0.92 | 0.28 | 19 | ZNF266 |
| 2.6 | 1.3 | 4 | -0.73 | 3.8 | 0.00036 | 0.5 | -0.94 | 0.17 | 8 | DEFCR24 |
| 0.55 | 0.26 | 0.84 | 0.038 | 3.8 | 0.00037 | 0.5 | -0.96 | 0.17 | 10 | CYP26A1 |
| -0.73 | -1.1 | -0.34 | 0.13 | -3.8 | 0.00041 | 0.5 | -1 | 0.2 | 16 | TAOK2 |

Table S5: Top-30 DE genes in LOY individuals

| logFC | CI.L | CI.R | AveExpr | t | P.Value | adj.P.Val | B | SE | Chromosome | Symbol |
|---|---|---|---|---|---|---|---|---|---|---|
| -2.5 | -3.3 | -1.7 | -0.24 | -6.4 | $5.7 \times 10^{-8}$ | 0.0014 | 4 | 0.15 | Y | CSF2RA |
| -2.2 | -3 | -1.3 | -0.22 | -5.4 | $2.3 \times 10^{-6}$ | 0.028 | 2 | 0.21 | Y | CSF2RA |
| -0.92 | -1.4 | -0.47 | -0.12 | -4.1 | 0.00017 | 0.41 | -0.49 | 0.19 | Y | SFRS17A |
| -1.4 | -2.1 | -0.7 | 0.25 | -4 | 0.00024 | 0.48 | -0.71 | 0.24 | Y | TMSB4Y |
| -2.3 | -3.5 | -1.1 | 0.11 | -3.7 | 0.00049 | 0.5 | -1.1 | 0.2 | Y | EIF1AY |
| -5.2 | -8 | -2.3 | 1.1 | -3.6 | 0.00074 | 0.53 | -1.4 | 0.18 | Y | EIF1AY |
| -0.8 | -1.3 | -0.31 | 0.036 | -3.3 | 0.0018 | 0.69 | -1.9 | 0.21 | XY | ASMTL |
| -0.89 | -1.5 | -0.33 | 0.079 | -3.2 | 0.0024 | 0.77 | -2.1 | 0.2 | Y | TLNGY |
| -0.66 | -1.1 | -0.22 | -0.044 | -3 | 0.0041 | 0.89 | -2.4 | 0.25 | Y | BCORL2 |
| -1.9 | -3.3 | -0.53 | 0.43 | -2.8 | 0.0074 | 0.99 | -2.7 | 0.17 | Y | KDM5D |
| -0.88 | -1.5 | -0.24 | -0.15 | -2.8 | 0.0076 | 0.99 | -2.7 | 0.24 | Y | SFRS17A |
| -1.7 | -3 | -0.46 | 0.48 | -2.7 | 0.0087 | 1 | -2.8 | 0.19 | Y | TXLNGY |
| -5.5 | -9.7 | -1.3 | 2 | -2.6 | 0.011 | 1 | -3 | 0.21 | Y | RPS4Y1 |
| -1.7 | -3.1 | -0.38 | 0.2 | -2.6 | 0.013 | 1 | -3 | 0.36 | Y | RPS4Y2 |
| -0.75 | -1.3 | -0.17 | -0.086 | -2.6 | 0.013 | 1 | -3 | 0.15 | Y | ZBED1 |
| -0.77 | -1.4 | -0.17 | 0.01 | -2.6 | 0.013 | 1 | -3.1 | 0.21 | Y | ZFY |
| 0.49 | 0.1 | 0.87 | -0.0077 | 2.5 | 0.014 | 1 | -3.1 | 0.23 | Y | SHOX |
| -0.38 | -0.68 | -0.073 | 0.058 | -2.5 | 0.016 | 1 | -3.2 | 0.18 | Y | TTTY14 |
| 0.39 | 0.054 | 0.73 | -0.018 | 2.3 | 0.024 | 1 | -3.4 | 0.33 | Y | TTTY8 |
| -1 | -2 | -0.11 | 0.29 | -2.2 | 0.029 | 1 | -3.5 | 0.27 | Y | PRKY |
| -0.48 | -0.91 | -0.04 | 0.038 | -2.2 | 0.033 | 1 | -3.6 | 0.21 | Y | UTY |
| 0.41 | 0.022 | 0.81 | -0.095 | 2.1 | 0.039 | 1 | -3.7 | 0.18 | Y | PCDH11Y |
| -0.41 | -0.79 | -0.019 | 0.011 | -2.1 | 0.04 | 1 | -3.7 | 0.2 | Y | RBMY1A1 |
| 0.45 | 0.0065 | 0.9 | -0.068 | 2 | 0.047 | 1 | -3.8 | 0.25 | XY | SHOX |
| -0.4 | -0.82 | 0.028 | 0.16 | -1.9 | 0.066 | 1 | -4 | 0.21 | Y | PPP2R3B |
| -0.65 | -1.4 | 0.083 | 0.2 | -1.8 | 0.081 | 1 | -4.1 | 0.29 | Y | UTY |
| 0.32 | -0.042 | 0.68 | -0.034 | 1.8 | 0.082 | 1 | -4.1 | 0.32 | Y | BPY2B |
| -0.48 | -1 | 0.079 | -0.066 | -1.7 | 0.09 | 1 | -4.2 | 0.15 | Y | ASMTL |
| -0.28 | -0.61 | 0.05 | -0.085 | -1.7 | 0.094 | 1 | -4.2 | 0.21 | Y | TTTY2 |
| -0.52 | -1.2 | 0.12 | -0.19 | -1.6 | 0.11 | 1 | -4.3 | 0.19 | Y | GTPBP6 |

Table S6: Top-30 DE genes in LOY individuals located in gonosomes.

Table S7: Genes located on the human Y chromosome with homolog on X and a possible role in immunity.
[See supplementary Excel File]

| | effect | inf | sup | pvalue |
|---|---|---|---|---|
| B cell naive | -1.57 | -2.97 | -0.16 | 0.03454 |
| Neutrophil | -1.15 | -2.23 | -0.06 | 0.04477 |
| NK cell | 0.05 | -0.01 | 0.10 | 0.08817 |
| T cell NK | -0.85 | -1.86 | 0.16 | 0.1077 |
| Monocyte | 0.97 | -0.20 | 2.15 | 0.1115 |
| Macrophage | 1.05 | -0.27 | 2.37 | 0.1258 |
| T cell CD8+ naive | 1.00 | -0.39 | 2.38 | 0.1653 |
| T cell regulatory (Tregs) | 1.12 | -0.49 | 2.74 | 0.1809 |
| Eosinophil | -0.75 | -1.89 | 0.40 | 0.207 |
| B cell memory | -0.85 | -2.25 | 0.55 | 0.2414 |
| B cell plasma | -0.53 | -1.43 | 0.37 | 0.258 |
| T cell CD4+ (non-regulatory) | 0.53 | -0.38 | 1.45 | 0.2599 |
| Macrophage M2 | 0.35 | -0.27 | 0.98 | 0.2738 |
| T cell CD4+ central memory | 0.45 | -0.39 | 1.30 | 0.3009 |
| T cell CD4+ Th2 | -0.26 | -0.77 | 0.24 | 0.3128 |
| Plasmacytoid dendritic cell | -0.73 | -2.18 | 0.72 | 0.3304 |
| T cell CD4+ effector memory | 0.83 | -0.85 | 2.51 | 0.3401 |
| Myeloid dendritic cell | 0.53 | -0.60 | 1.66 | 0.365 |
| T cell CD8+ | -0.51 | -1.79 | 0.76 | 0.4345 |
| Mast cell | 0.27 | -0.41 | 0.95 | 0.4372 |
| Common lymphoid progenitor | 0.18 | -0.31 | 0.67 | 0.4699 |
| Macrophage M1 | 0.33 | -0.72 | 1.38 | 0.5399 |
| Cancer associated fibroblast | -0.43 | -1.83 | 0.98 | 0.5559 |
| T cell CD8+ central memory | 0.35 | -0.89 | 1.59 | 0.5845 |
| T cell gamma delta | 0.00 | -0.00 | 0.00 | 0.6374 |
| Class-switched memory B cell | -0.27 | -1.74 | 1.20 | 0.7227 |
| T cell CD4+ naive | 0.15 | -1.35 | 1.65 | 0.8456 |
| Common myeloid progenitor | 0.11 | -1.18 | 1.39 | 0.8695 |
| Hematopoietic stem cell | -0.04 | -0.52 | 0.45 | 0.884 |
| T cell CD4+ Th1 | 0.11 | -1.59 | 1.81 | 0.9036 |
| T cell CD8+ effector memory | -0.00 | -0.00 | 0.00 | 0.9219 |
| Granulocyte-monocyte progenitor | -2.22 | -3.09 | -1.35 | 1.085e-05 |
| Endothelial cell | 1.40 | 0.77 | 2.02 | 7.856e-05 |

Table S8: Association between cell type composition estimated using bulk transcriptomic data (immunedecov R package) and LOY estatus

| Description | pvalue | p.adjust |
|---|---|---|
| blood coagulation | $6.3 \times 10^{-9}$ | $2.6 \times 10^{-6}$ |
| hemostasis | $7.3 \times 10^{-9}$ | $2.6 \times 10^{-6}$ |
| coagulation | $7.6 \times 10^{-9}$ | $2.6 \times 10^{-6}$ |
| platelet degranulation | $1.6 \times 10^{-7}$ | $4.1 \times 10^{-5}$ |
| regulation of body fluid levels | $4.1 \times 10^{-7}$ | $8.3 \times 10^{-5}$ |
| leukocyte migration | $1.9 \times 10^{-6}$ | 0.00032 |
| platelet activation | $8.6 \times 10^{-6}$ | 0.0013 |
| oxygen transport | $1.4 \times 10^{-5}$ | 0.0017 |
| gas transport | $2.9 \times 10^{-5}$ | 0.0033 |
| platelet aggregation | $3.4 \times 10^{-5}$ | 0.0035 |
| blood coagulation, fibrin clot formation | $8.5 \times 10^{-5}$ | 0.0079 |
| homotypic cell-cell adhesion | 0.0001 | 0.009 |
| cellular oxidant detoxification | 0.00028 | 0.022 |
| cellular detoxification | 0.00037 | 0.027 |
| detoxification | 0.00046 | 0.031 |
| cell-substrate adhesion | 0.00054 | 0.035 |
| cell-matrix adhesion | 0.00059 | 0.035 |

Table S9: GO enrichment analysis of DE genes in individuals with LOY. GO terms significant at 5% FDR.

| Description | pvalue | p.adjust |
|---|---|---|
| ECM-receptor interaction | $3.5 \times 10^{-5}$ | 0.0024 |
| Platelet activation | 0.00018 | 0.0063 |
| Hematopoietic cell lineage | 0.00085 | 0.015 |
| Viral protein interaction with cytokine and cytokine receptor | 0.00088 | 0.015 |
| Malaria | 0.0013 | 0.016 |
| Cytokine-cytokine receptor interaction | 0.0015 | 0.016 |
| Focal adhesion | 0.0016 | 0.016 |

Table S10: KEGG enrichment analysis of DE genes in individuals with LOY. KEGG terms significant at 5% FDR.

| Description | pvalue | p.adjust |
|---|---|---|
| response to virus | $1.7 \times 10^{-21}$ | $5.9 \times 10^{-18}$ |
| defense response to other organism | $1.5 \times 10^{-19}$ | $2.6 \times 10^{-16}$ |
| type I interferon signaling pathway | $8.8 \times 10^{-17}$ | $7.5 \times 10^{-14}$ |
| cellular response to type I interferon | $8.8 \times 10^{-17}$ | $7.5 \times 10^{-14}$ |
| response to type I interferon | $2 \times 10^{-16}$ | $1.4 \times 10^{-13}$ |
| defense response to virus | $2.6 \times 10^{-16}$ | $1.5 \times 10^{-13}$ |
| response to molecule of bacterial origin | $3.9 \times 10^{-16}$ | $1.9 \times 10^{-13}$ |
| response to lipopolysaccharide | $1.1 \times 10^{-15}$ | $4.6 \times 10^{-13}$ |
| response to interferon-gamma | $8 \times 10^{-14}$ | $3 \times 10^{-11}$ |
| regulation of inflammatory response | $6.3 \times 10^{-13}$ | $2.2 \times 10^{-10}$ |
| positive regulation of response to external stimulus | $2.5 \times 10^{-12}$ | $7.8 \times 10^{-10}$ |
| negative regulation of viral genome replication | $9.4 \times 10^{-12}$ | $2.7 \times 10^{-9}$ |
| acute-phase response | $1.4 \times 10^{-11}$ | $3.6 \times 10^{-9}$ |
| acute inflammatory response | $6.2 \times 10^{-11}$ | $1.5 \times 10^{-8}$ |
| regulation of innate immune response | $8.8 \times 10^{-11}$ | $2 \times 10^{-8}$ |
| cellular response to lipopolysaccharide | $1.1 \times 10^{-10}$ | $2.4 \times 10^{-8}$ |
| positive regulation of inflammatory response | $1.2 \times 10^{-10}$ | $2.5 \times 10^{-8}$ |
| cellular response to molecule of bacterial origin | $2.1 \times 10^{-10}$ | $4 \times 10^{-8}$ |
| regulation of viral genome replication | $2.4 \times 10^{-10}$ | $4.3 \times 10^{-8}$ |
| neutrophil mediated immunity | $2.7 \times 10^{-10}$ | $4.6 \times 10^{-8}$ |
| negative regulation of viral process | $4.1 \times 10^{-10}$ | $6.6 \times 10^{-8}$ |
| negative regulation of viral life cycle | $8.3 \times 10^{-10}$ | $1.3 \times 10^{-7}$ |
| cellular response to biotic stimulus | $1.3 \times 10^{-9}$ | $1.8 \times 10^{-7}$ |
| neutrophil activation | $1.3 \times 10^{-9}$ | $1.8 \times 10^{-7}$ |
| positive regulation of MAPK cascade | $1.3 \times 10^{-9}$ | $1.8 \times 10^{-7}$ |
| regulation of response to cytokine stimulus | $1.8 \times 10^{-9}$ | $2.3 \times 10^{-7}$ |
| positive regulation of peptidyl-tyrosine phosphorylation | $1.9 \times 10^{-9}$ | $2.4 \times 10^{-7}$ |
| cornification | $1.9 \times 10^{-9}$ | $2.4 \times 10^{-7}$ |
| positive regulation of innate immune response | $2.8 \times 10^{-9}$ | $3.2 \times 10^{-7}$ |
| regulation of multi-organism process | $2.8 \times 10^{-9}$ | $3.2 \times 10^{-7}$ |
| humoral immune response | $2.9 \times 10^{-9}$ | $3.2 \times 10^{-7}$ |
| regulation of lipid storage | $3 \times 10^{-9}$ | $3.2 \times 10^{-7}$ |
| positive regulation of cytokine production | $3.2 \times 10^{-9}$ | $3.3 \times 10^{-7}$ |
| neutrophil degranulation | $3.5 \times 10^{-9}$ | $3.5 \times 10^{-7}$ |
| neutrophil activation involved in immune response | $4 \times 10^{-9}$ | $3.9 \times 10^{-7}$ |
| viral genome replication | $4.5 \times 10^{-9}$ | $4.2 \times 10^{-7}$ |
| cell chemotaxis | $4.5 \times 10^{-9}$ | $4.2 \times 10^{-7}$ |
| positive regulation of DNA-binding transcription factor activity | $5.8 \times 10^{-9}$ | $5.3 \times 10^{-7}$ |
| positive regulation of smooth muscle cell proliferation | $6.3 \times 10^{-9}$ | $5.5 \times 10^{-7}$ |
| leukocyte migration | $7.8 \times 10^{-9}$ | $6.7 \times 10^{-7}$ |

Table S11: Top GO enrichment of DE genes in the two cell lines infected with SARS-CoV-2.

| Gene | Chr | logFC (LOY) | P.value | logFC (NHBE) | P.value | logFC (A549) | P.value |
|---|---|---|---|---|---|---|---|
| CXCL5 | 4 | 1.7 | 0.014 | 3.5 | $7.1 \times 10^{-31}$ | 0.77 | $1.7 \times 10^{-17}$ |
| IFI44L | 1 | -1.6 | 0.021 | 2.3 | 0.00028 | 5.7 | $2.5 \times 10^{-5}$ |
| IFI6 | 1 | -1 | 0.049 | 2.3 | $3.3 \times 10^{-5}$ | 4.3 | $2.3 \times 10^{-261}$ |
| IFIT1 | 10 | -1.2 | 0.032 | 0.82 | 0.00025 | 4.3 | $3 \times 10^{-141}$ |
| IFIT3 | 10 | -1.4 | 0.017 | 0.73 | $5.1 \times 10^{-5}$ | 2 | $1 \times 10^{-32}$ |
| ITGB3 | 17 | 1.7 | 0.013 | 1.2 | $9.1 \times 10^{-7}$ | -0.24 | 0.42 |
| KRT23 | 17 | -1.6 | $8.2 \times 10^{-5}$ | 0.59 | $4.5 \times 10^{-7}$ | 1.6 | 0.37 |
| KYNU | 2 | 1.2 | 0.0058 | 0.84 | $7.8 \times 10^{-10}$ | 0.48 | $1.2 \times 10^{-5}$ |
| PROS1 | 3 | 1.5 | 0.0022 | -0.66 | $4.2 \times 10^{-6}$ | 0.3 | 0.027 |
| S100P | 4 | 2.4 | 0.033 | 0.69 | $3.2 \times 10^{-10}$ | -0.11 | 0.51 |
| SLPI | 20 | -1.2 | 0.037 | 0.58 | $7.3 \times 10^{-9}$ | 0.42 | 0.018 |
| TSC22D3 | X | 1 | 0.024 | -0.72 | $6.9 \times 10^{-10}$ | -0.17 | 0.2 |
| VNN1 | 6 | 1.6 | 0.00099 | 1.9 | $3.4 \times 10^{-8}$ | 0.46 | 0.41 |

Table S12: Intersection between significant genes in the analysis of normal vs LOY individuals and in the two cell lines infected with SARS-CoV-2 (NHBE and A549).

### Supplementary figures

Figure S1: Plots of the whole-genome molecular karyotype obtained by SNParray of blood DNA from all 133 individuals of SCOURGE with detectable CMEs.
[See supplementary file FigureS1.pdf]

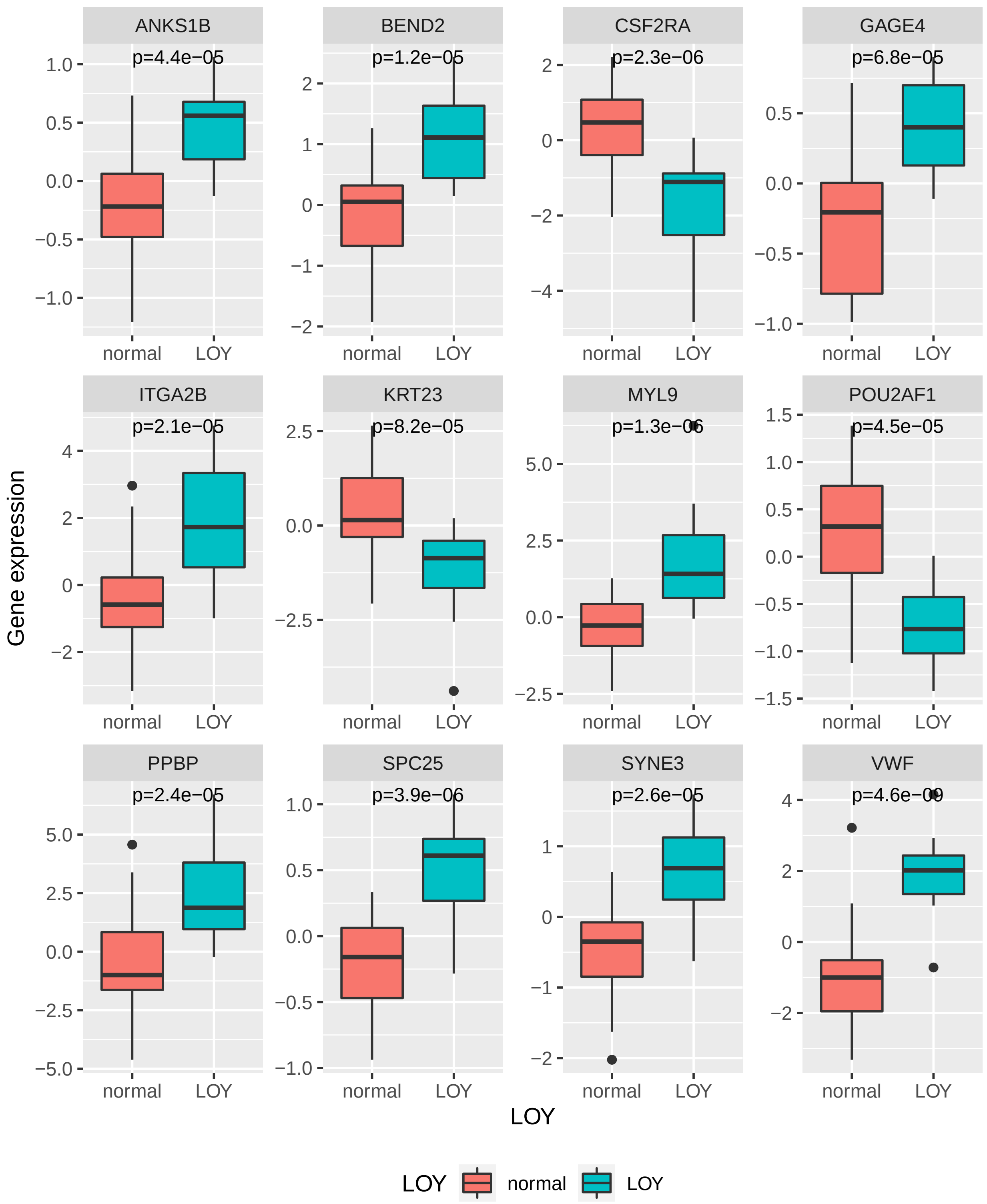


Figure S2: Top differentially expressed genes at genome level. The plots show the gene expression for individuals with (Y) and without (N) LOY. The p-values correspond to a linear model adjusted for age and surrogate variables using limma.

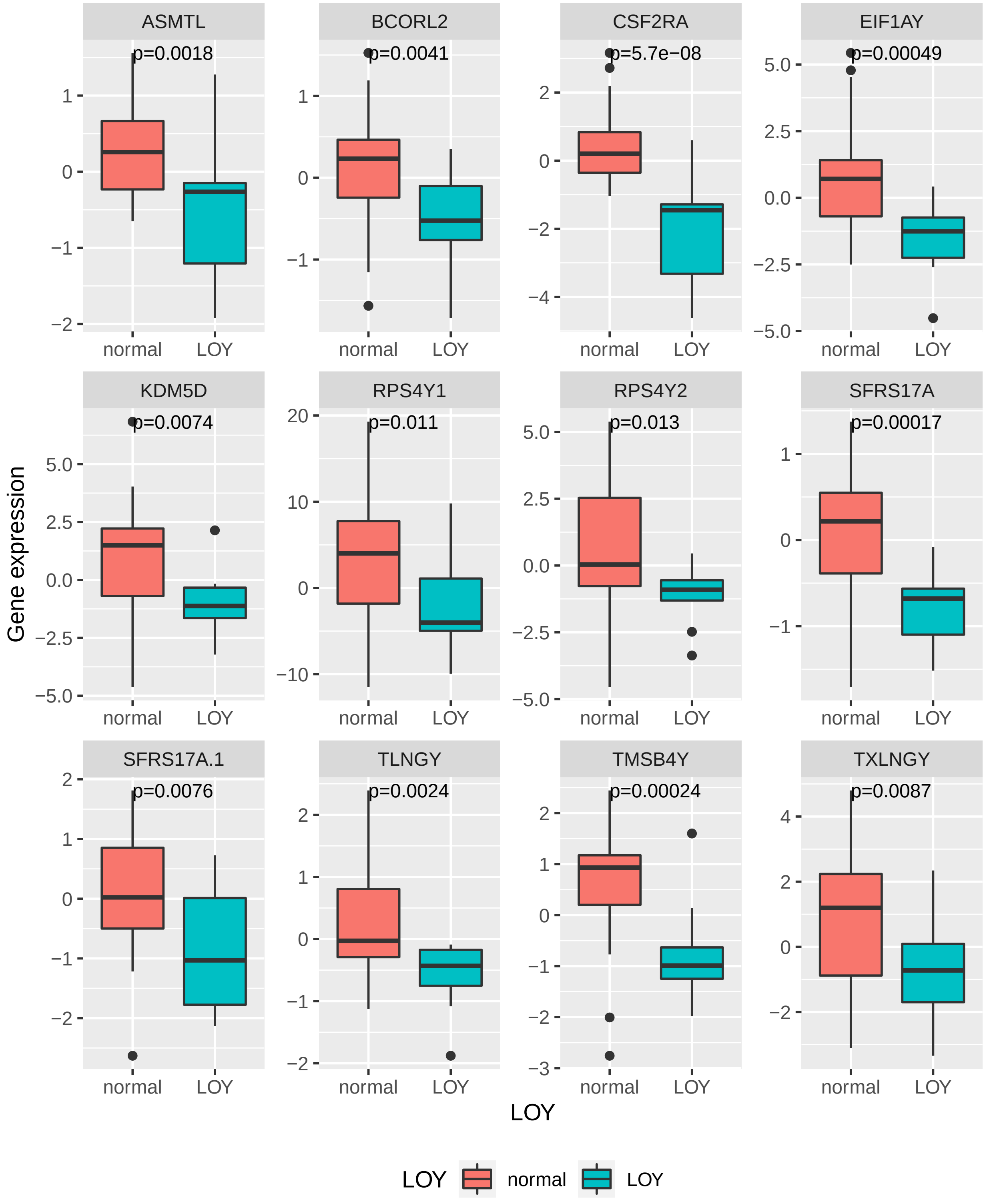


Figure S3: Top differentially expressed genes in chromosome Y. The plots show the gene expression for individuals with (Y) and without (N) LOY. The p-values correspond to a linear model adjusted for age and surrogate variables using limma.
