## Supplementary_FigureS1 for "Clonal chromosomal mosaicism and loss of chromosome Y in men are risk factors for SARS-CoV-2 vulnerability in the elderly"

### CME1

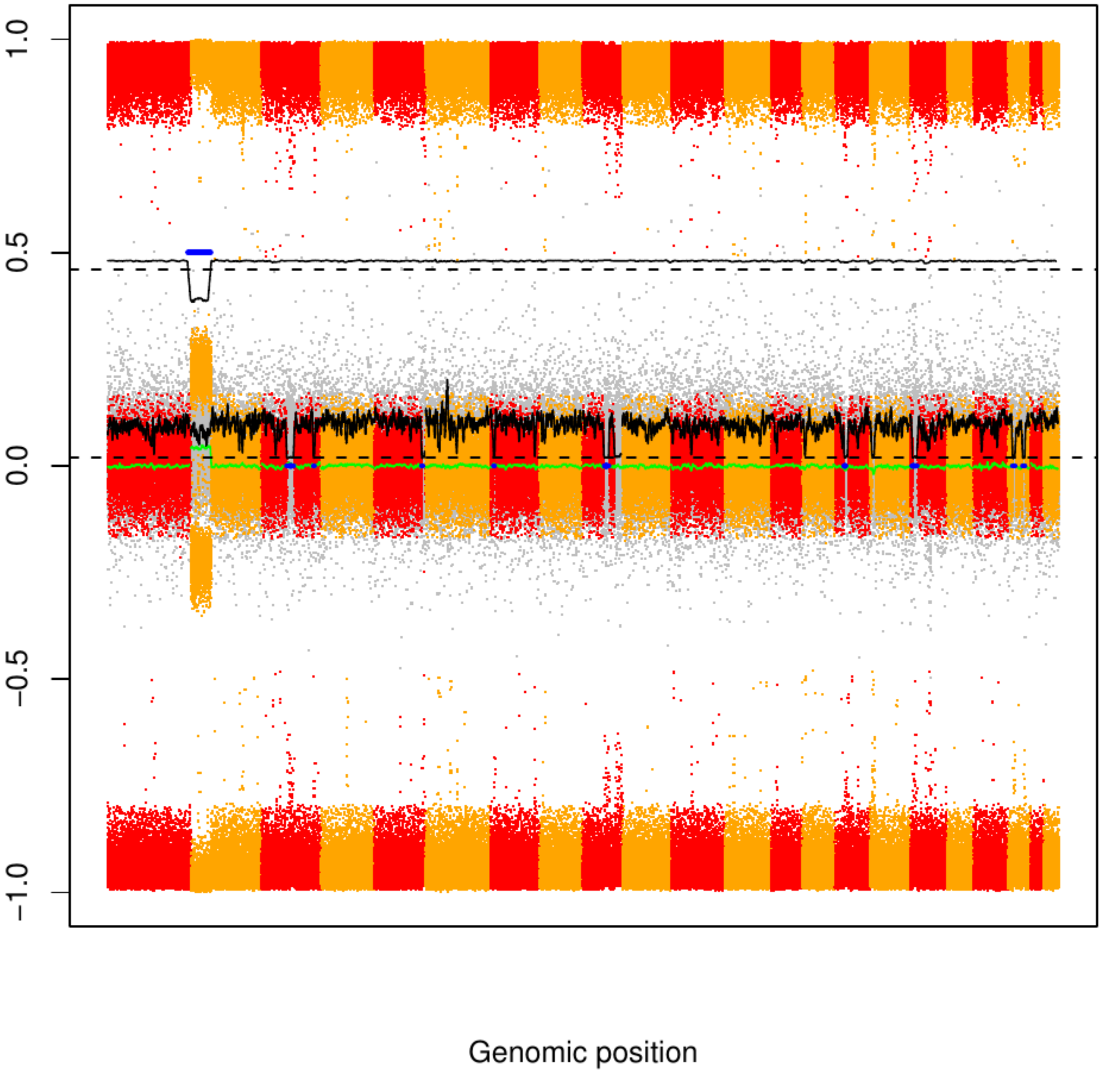

### CME2

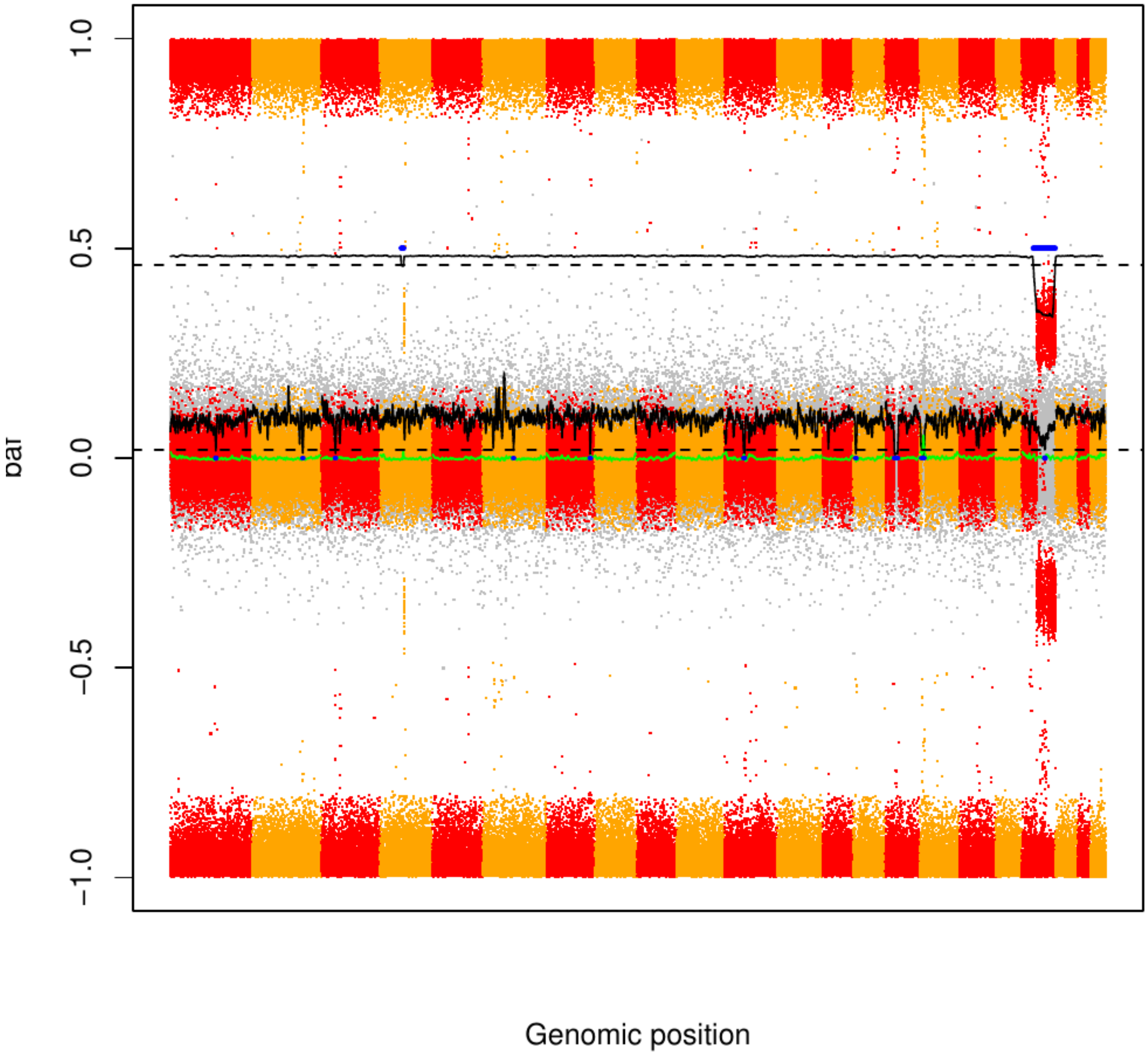

CME3

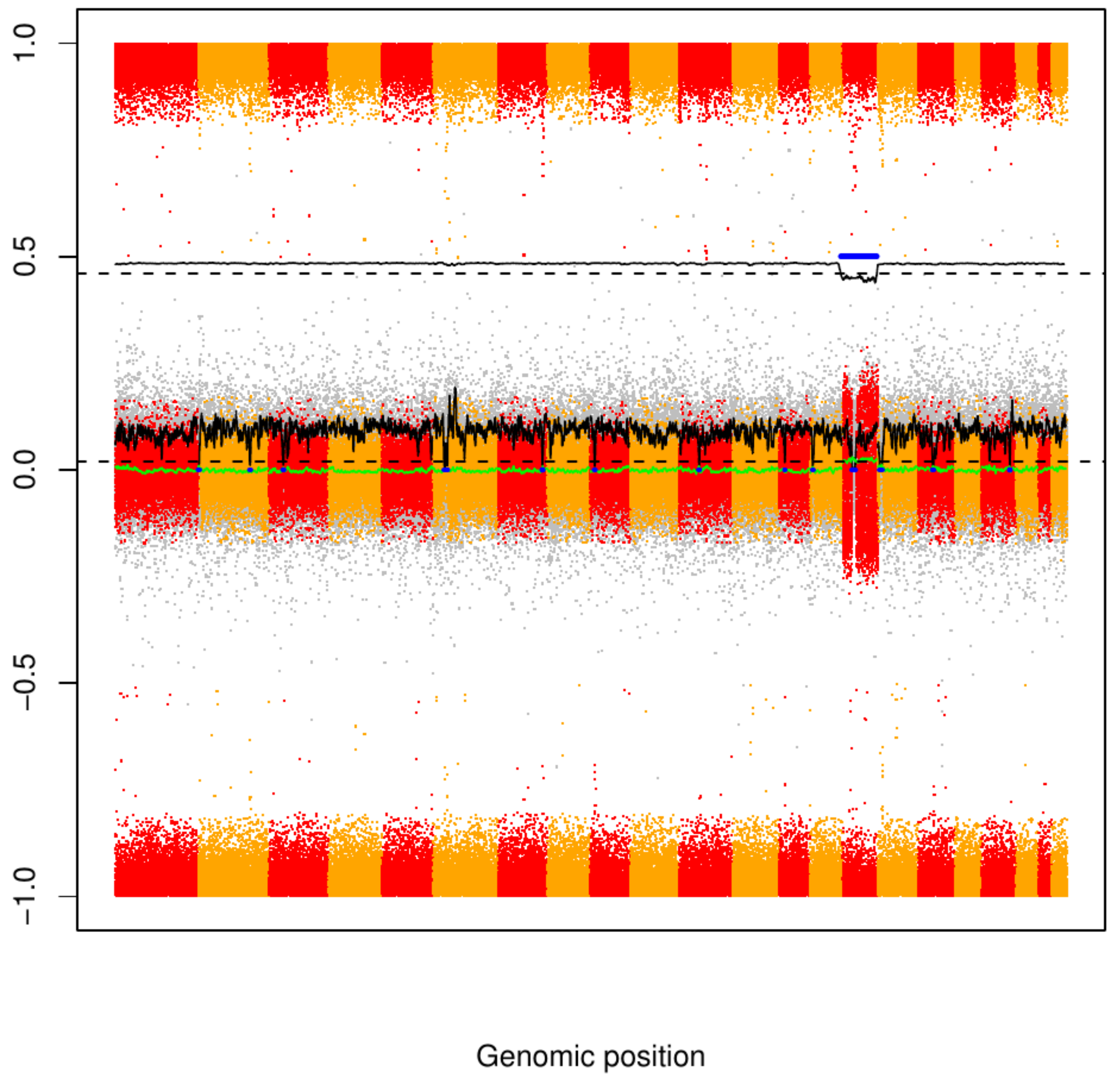

### CME4

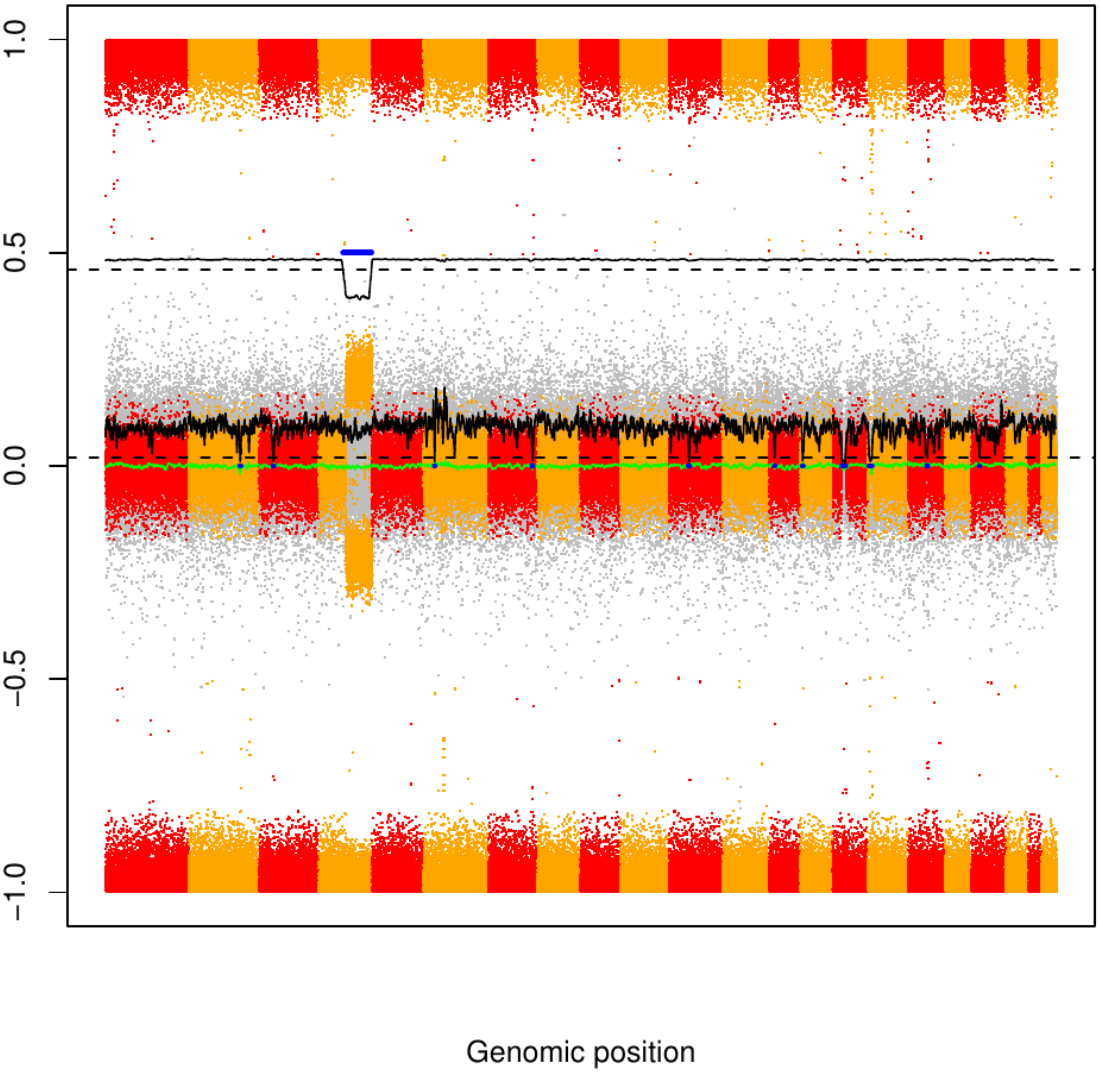

### CME5

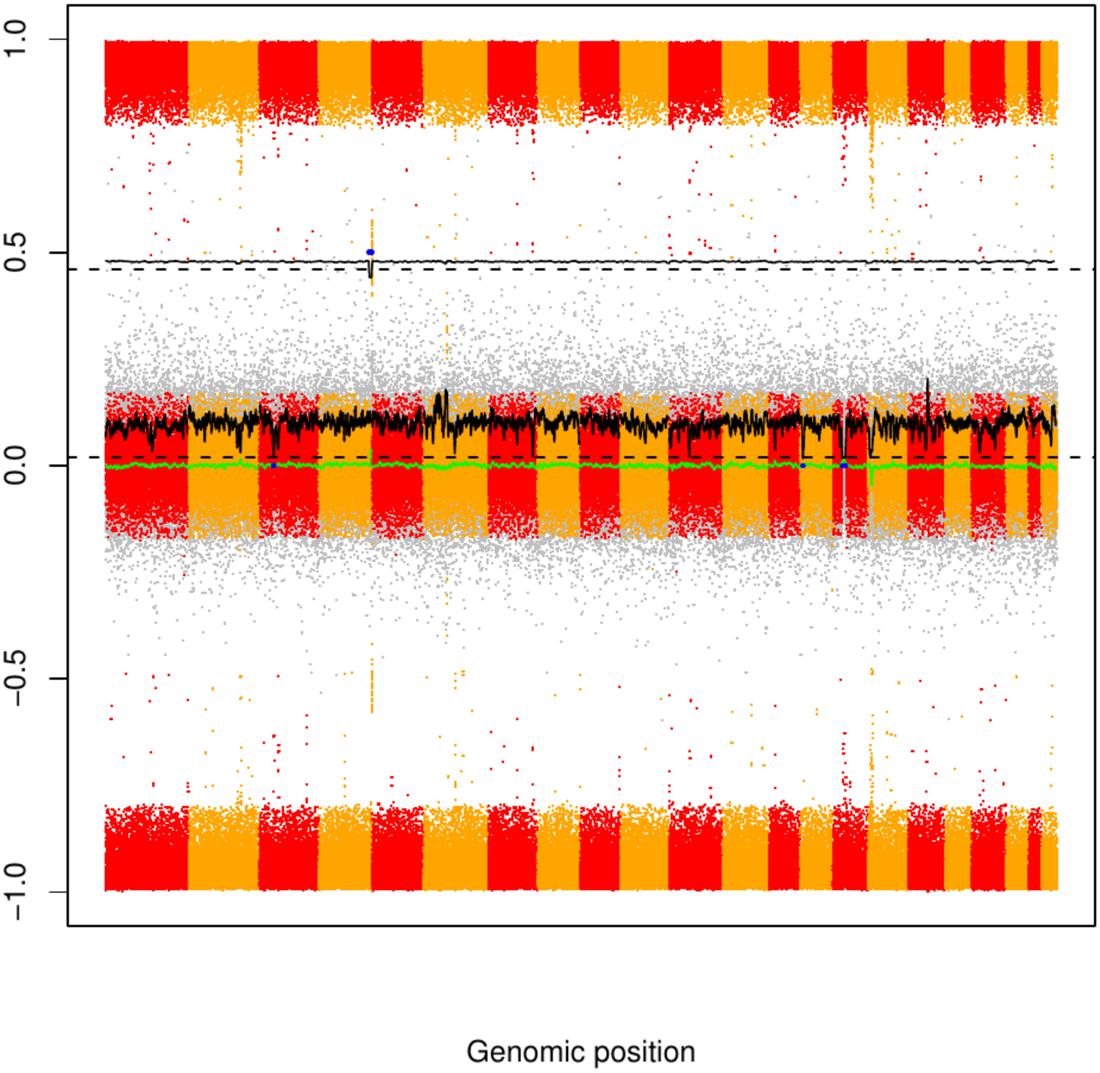

### CME6

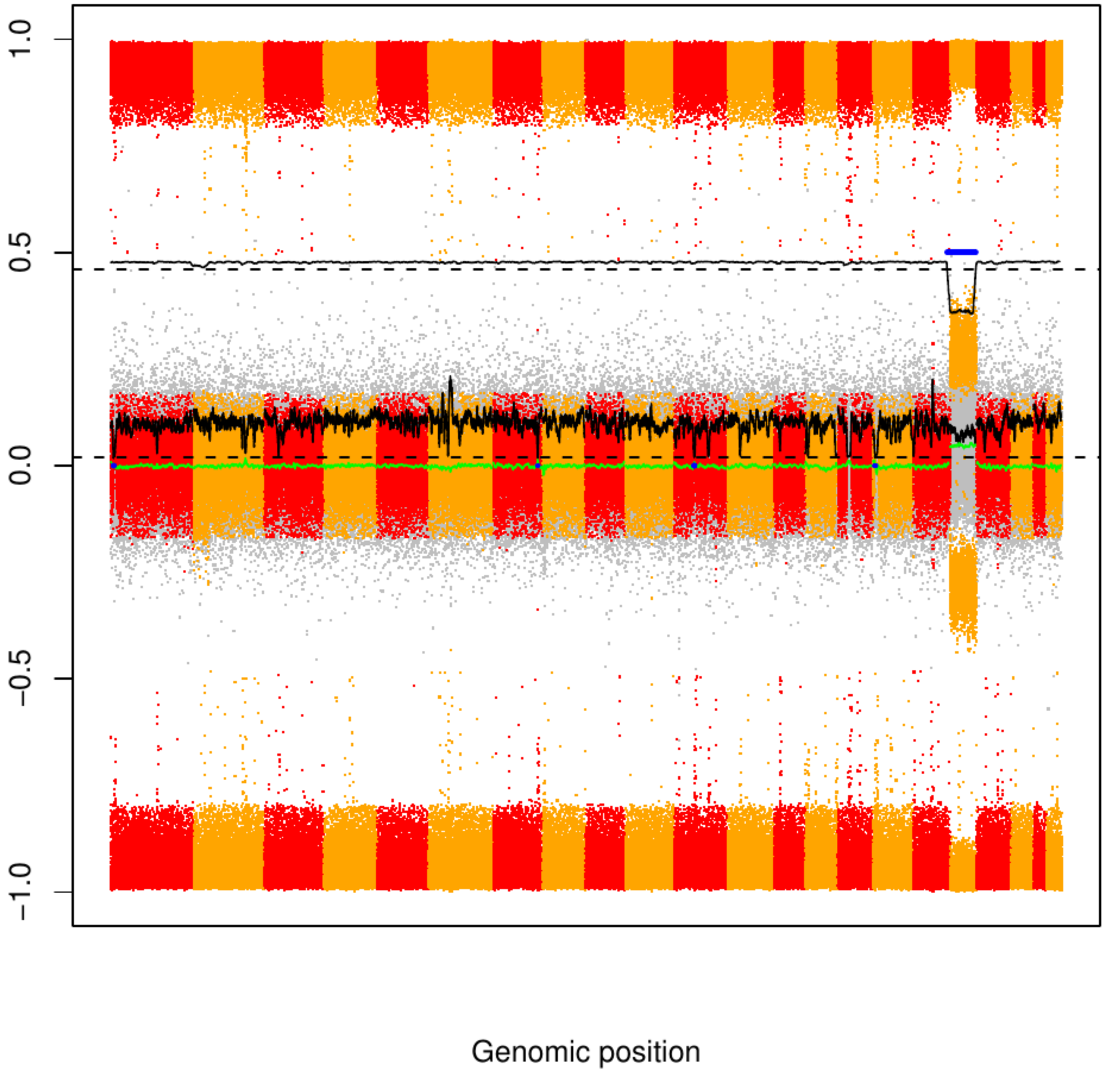

### CME7

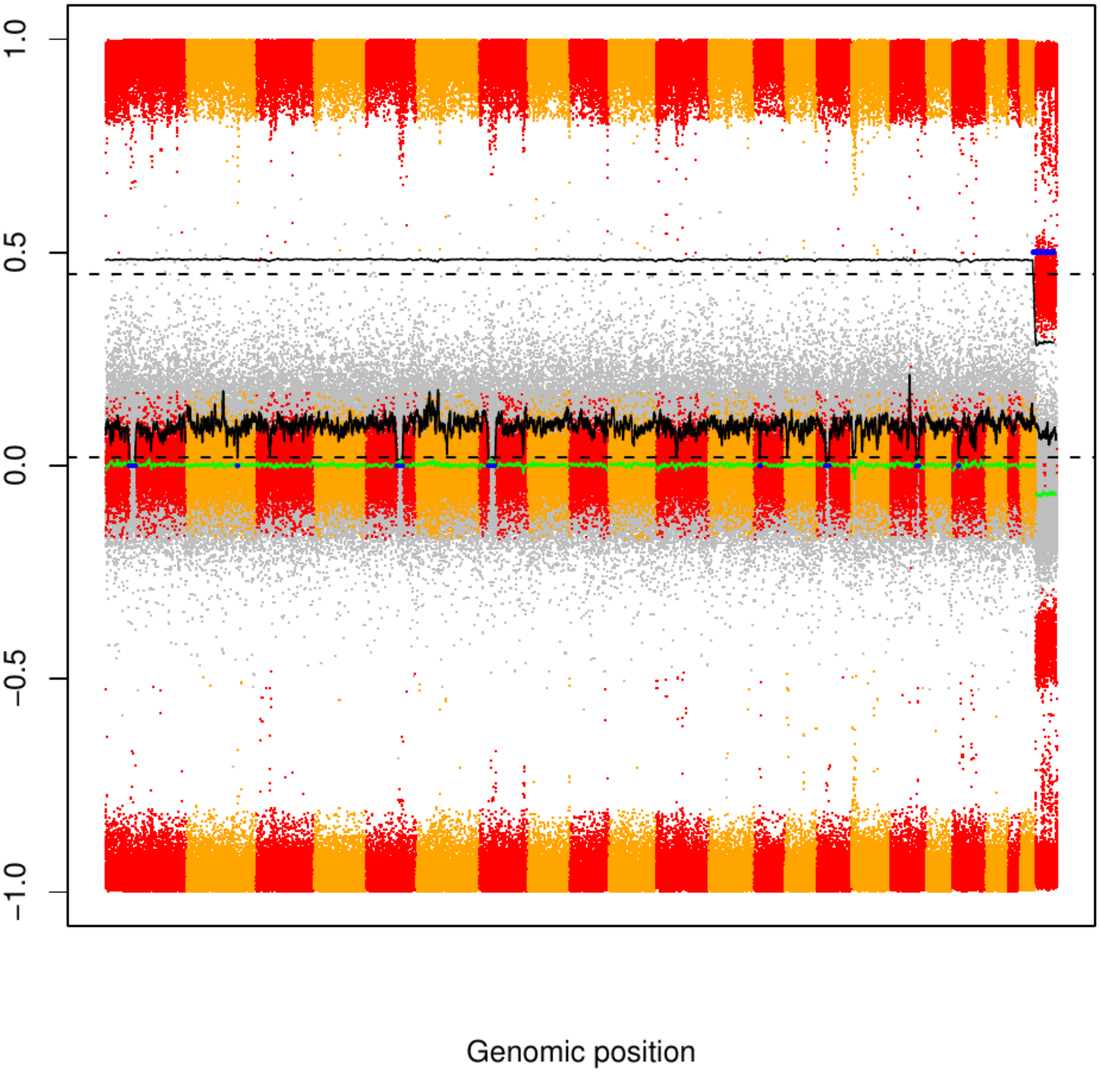

### CME8

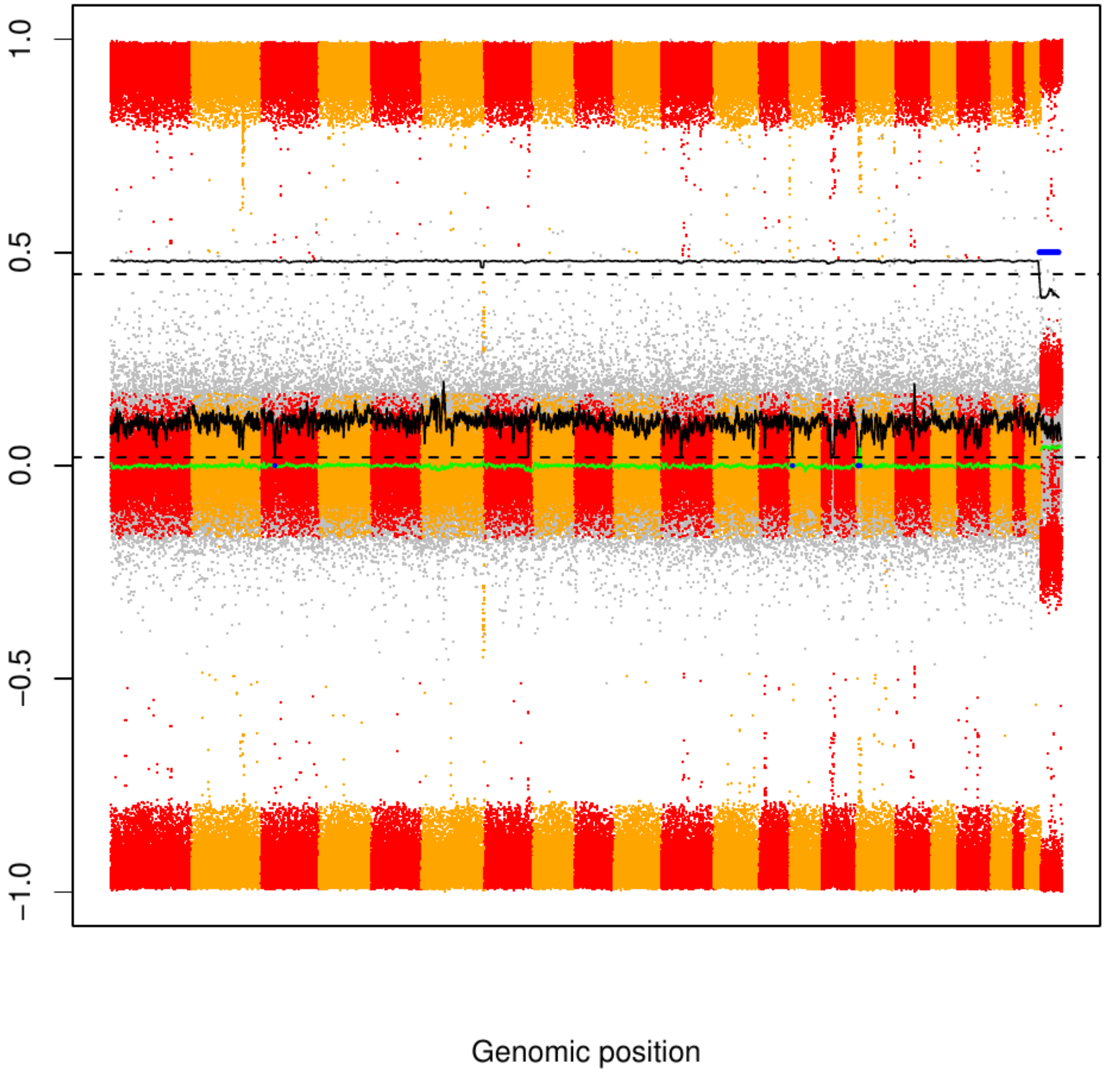

### CME9

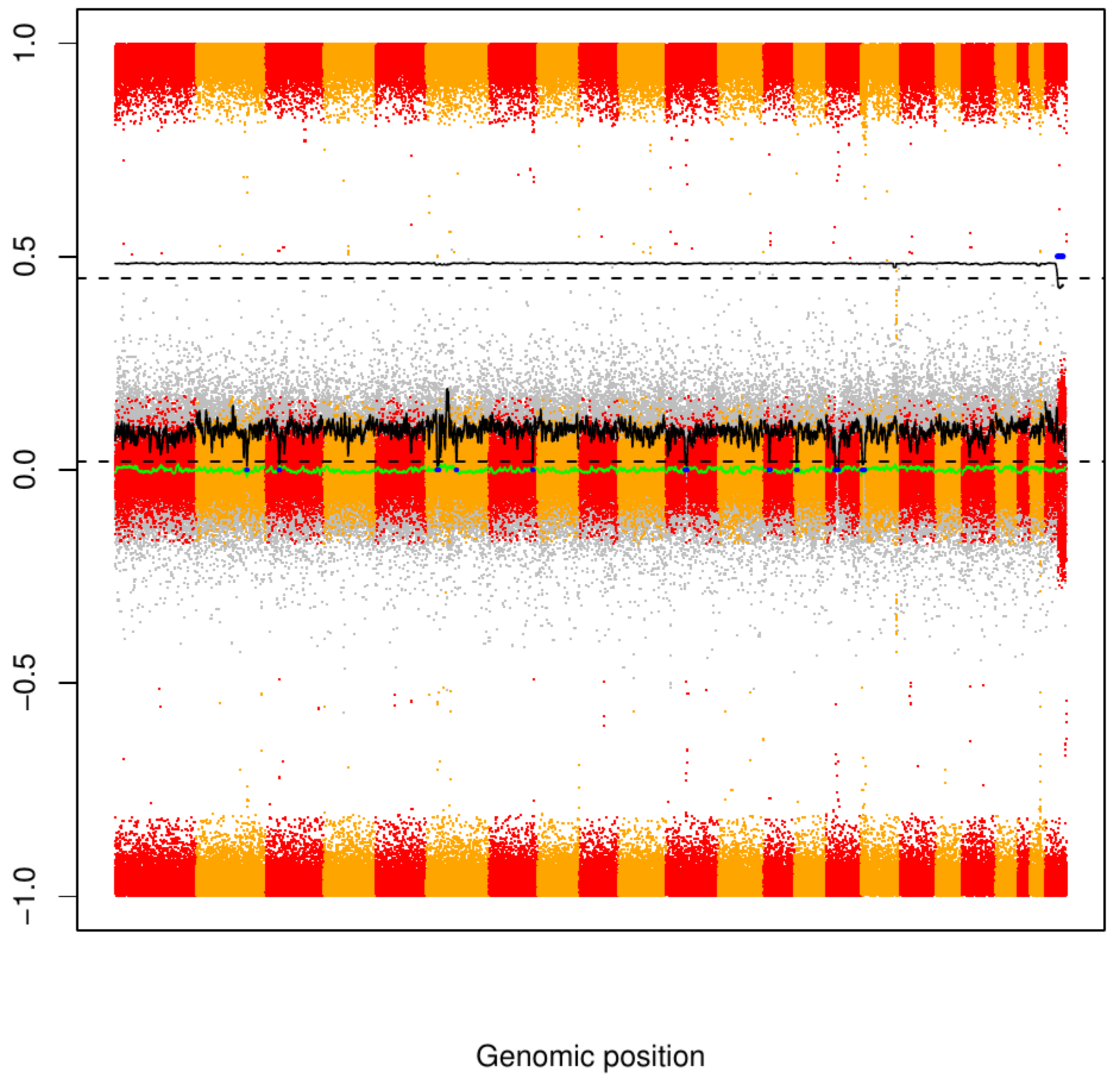

### CME10

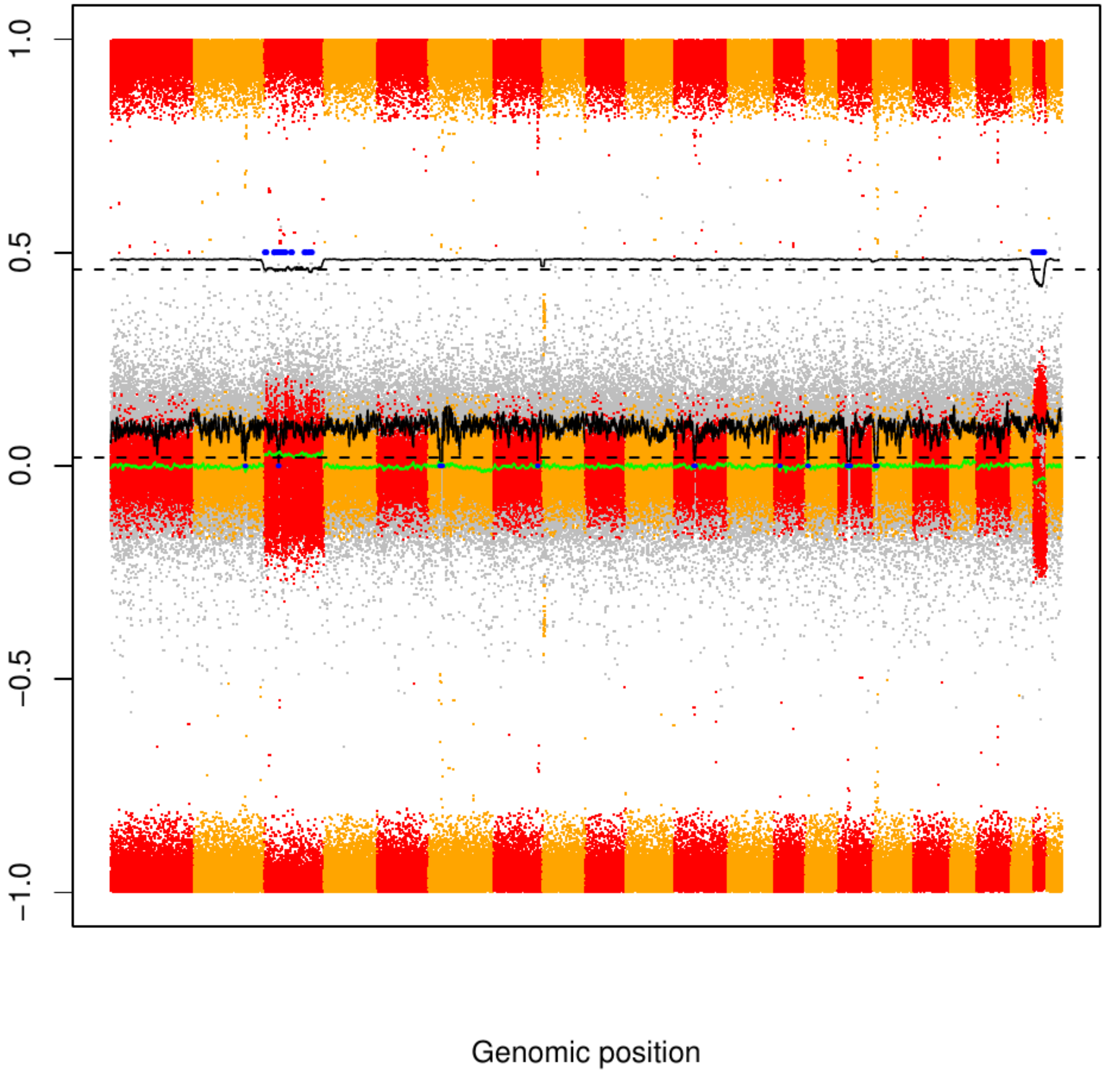

CME11

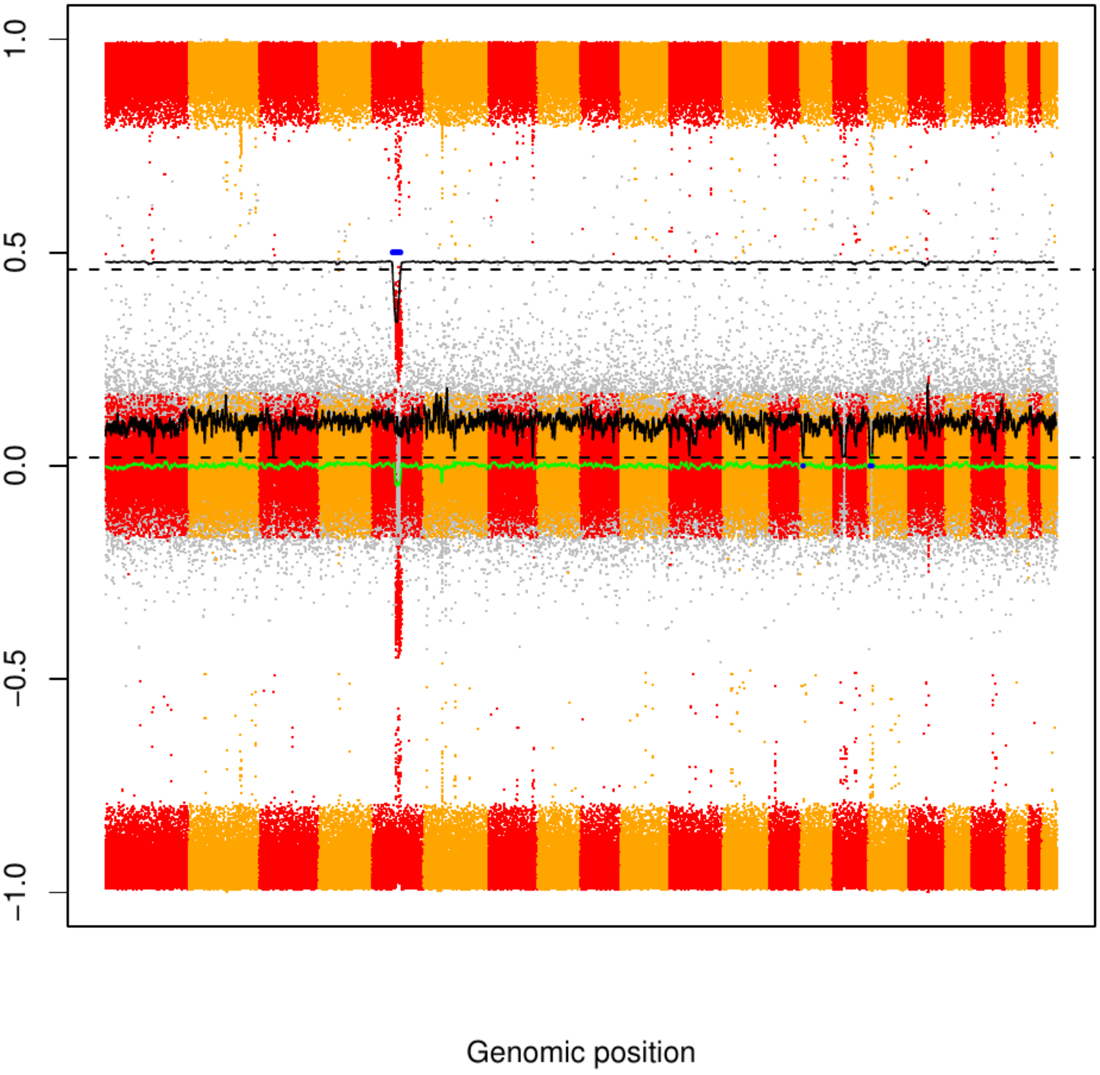

### CME12

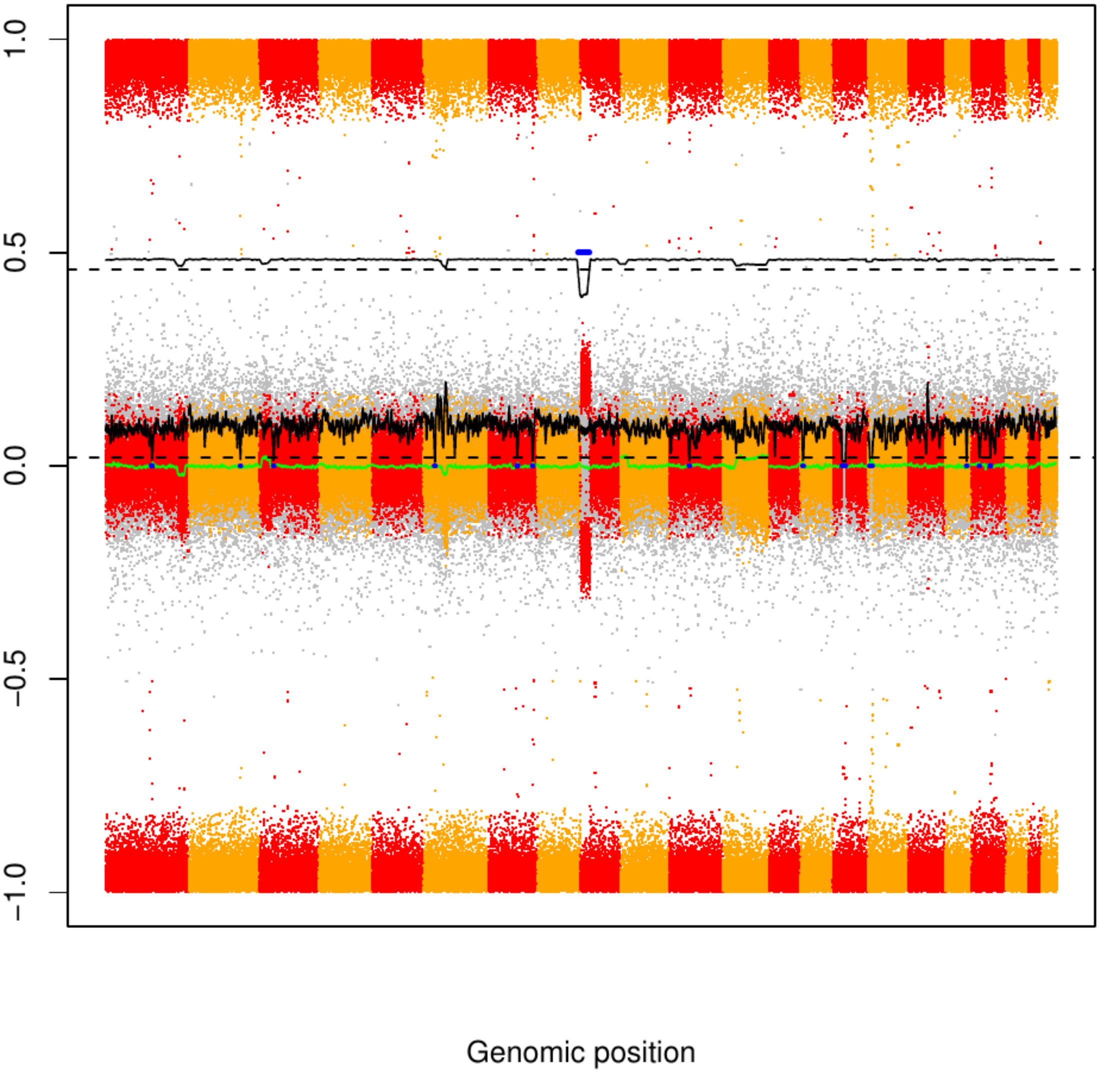

### CME13

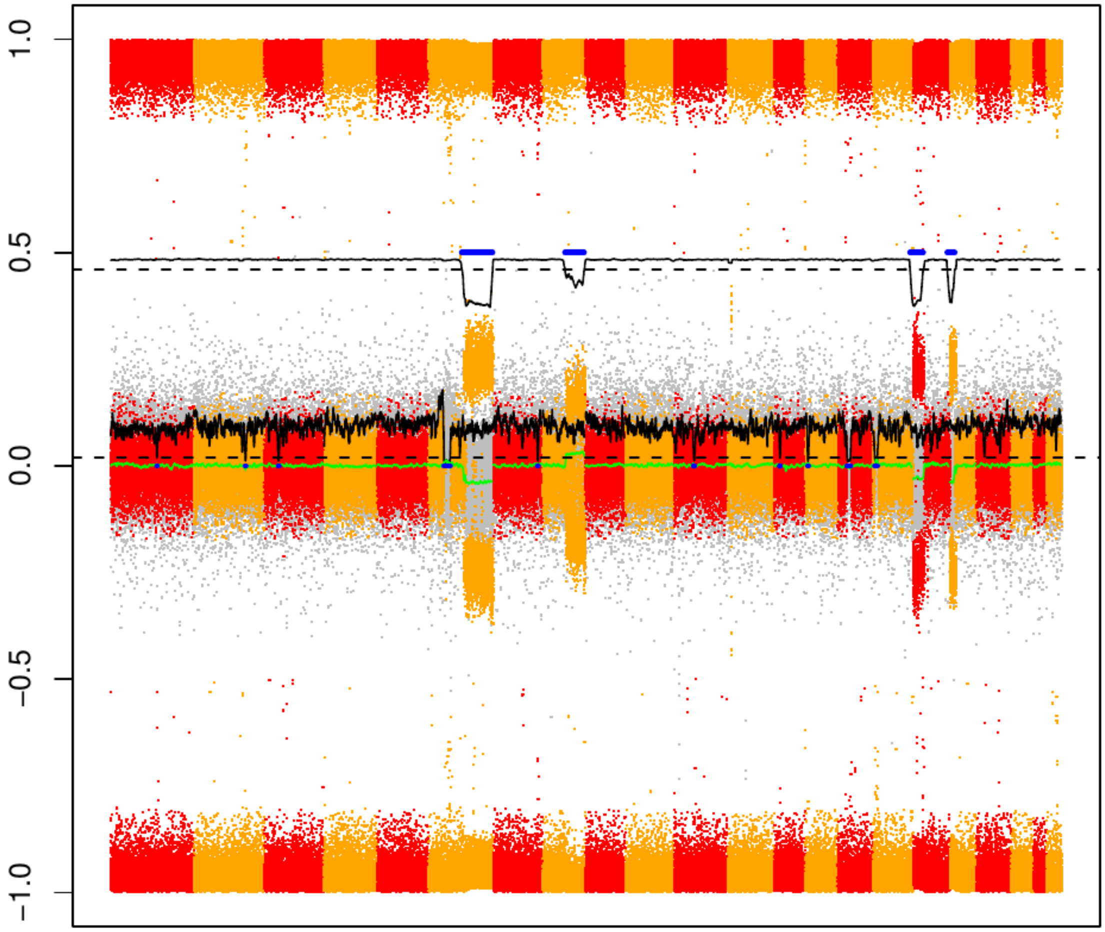

### CME14

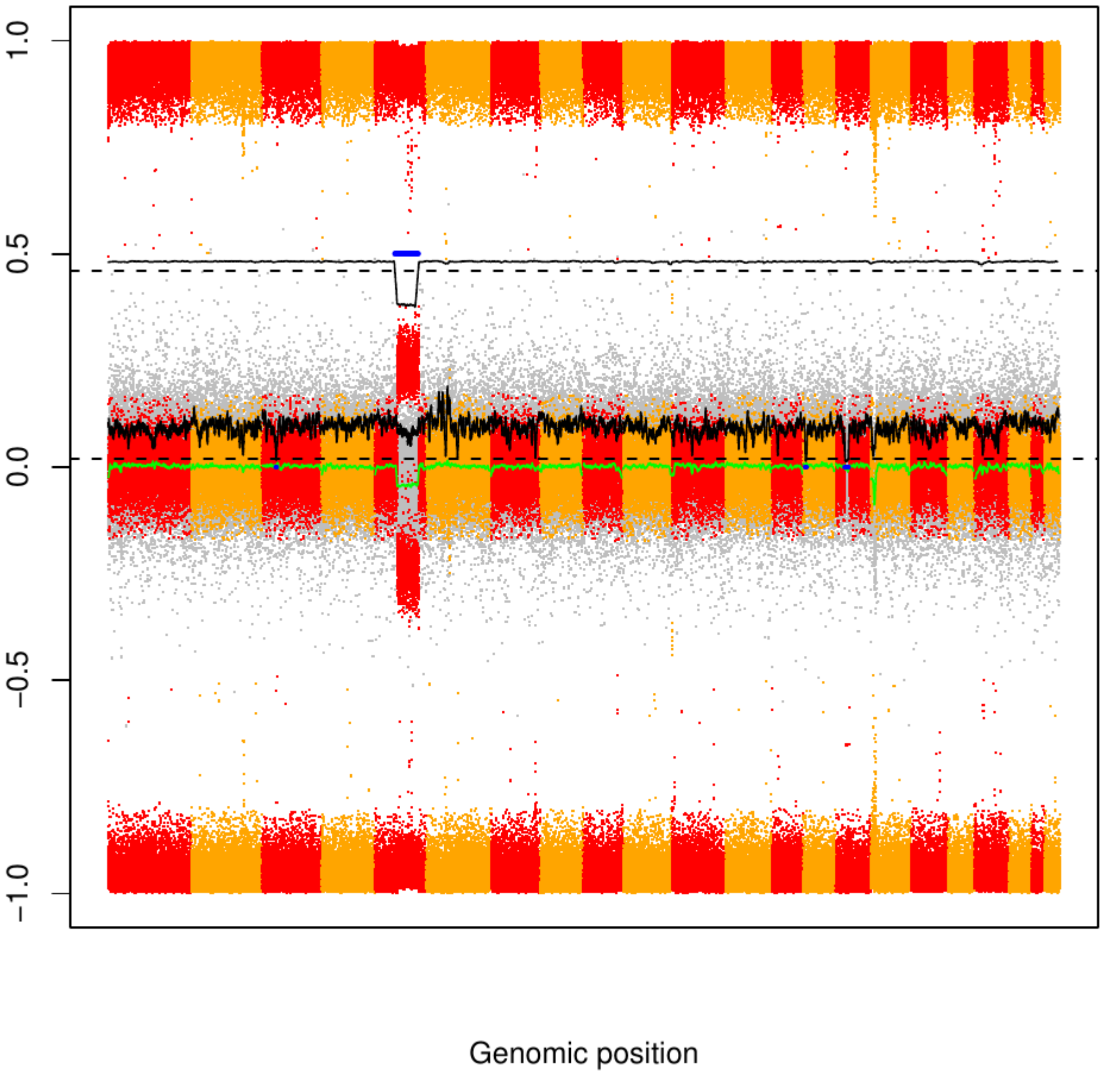

### CME15

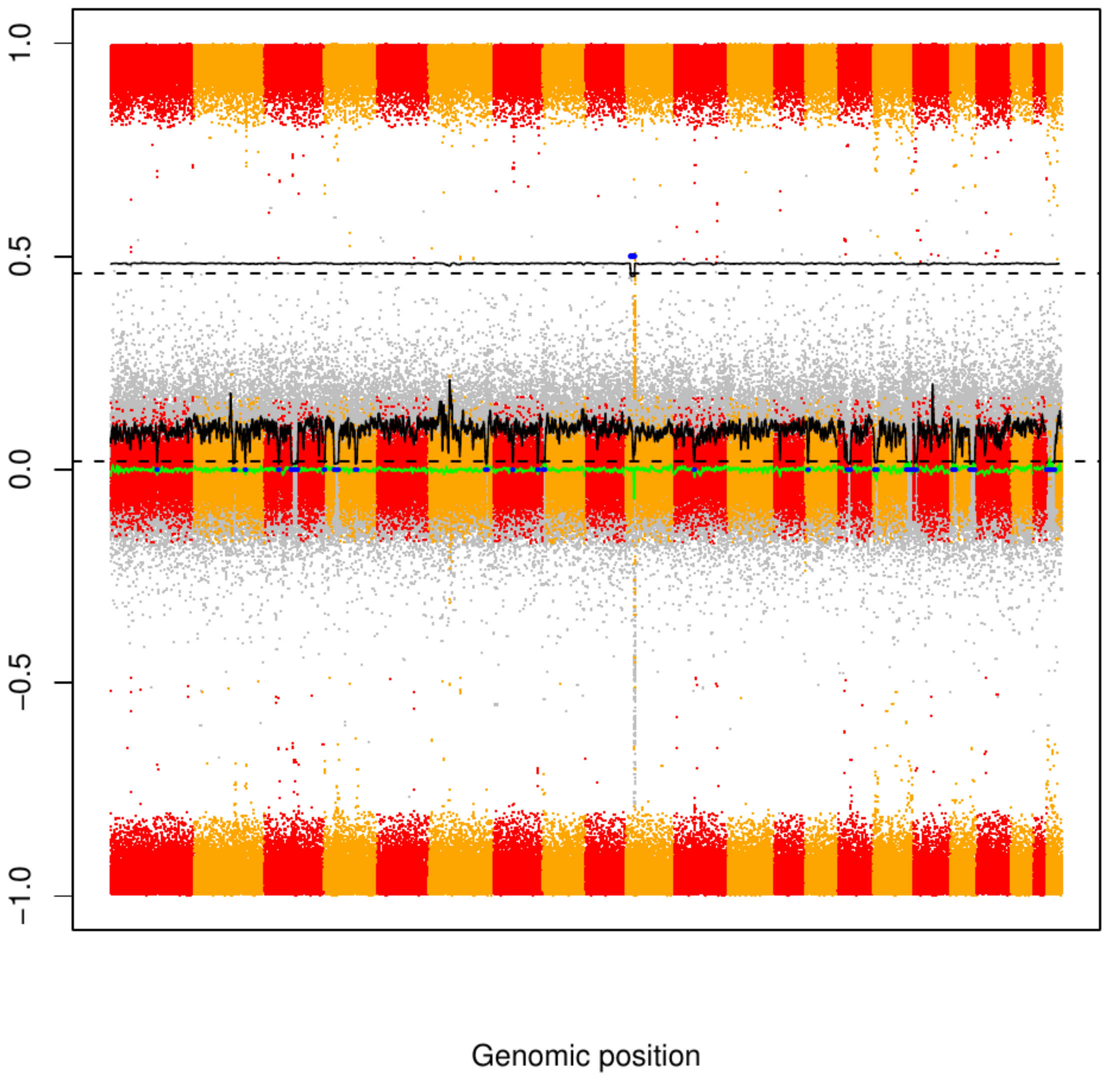

CME16

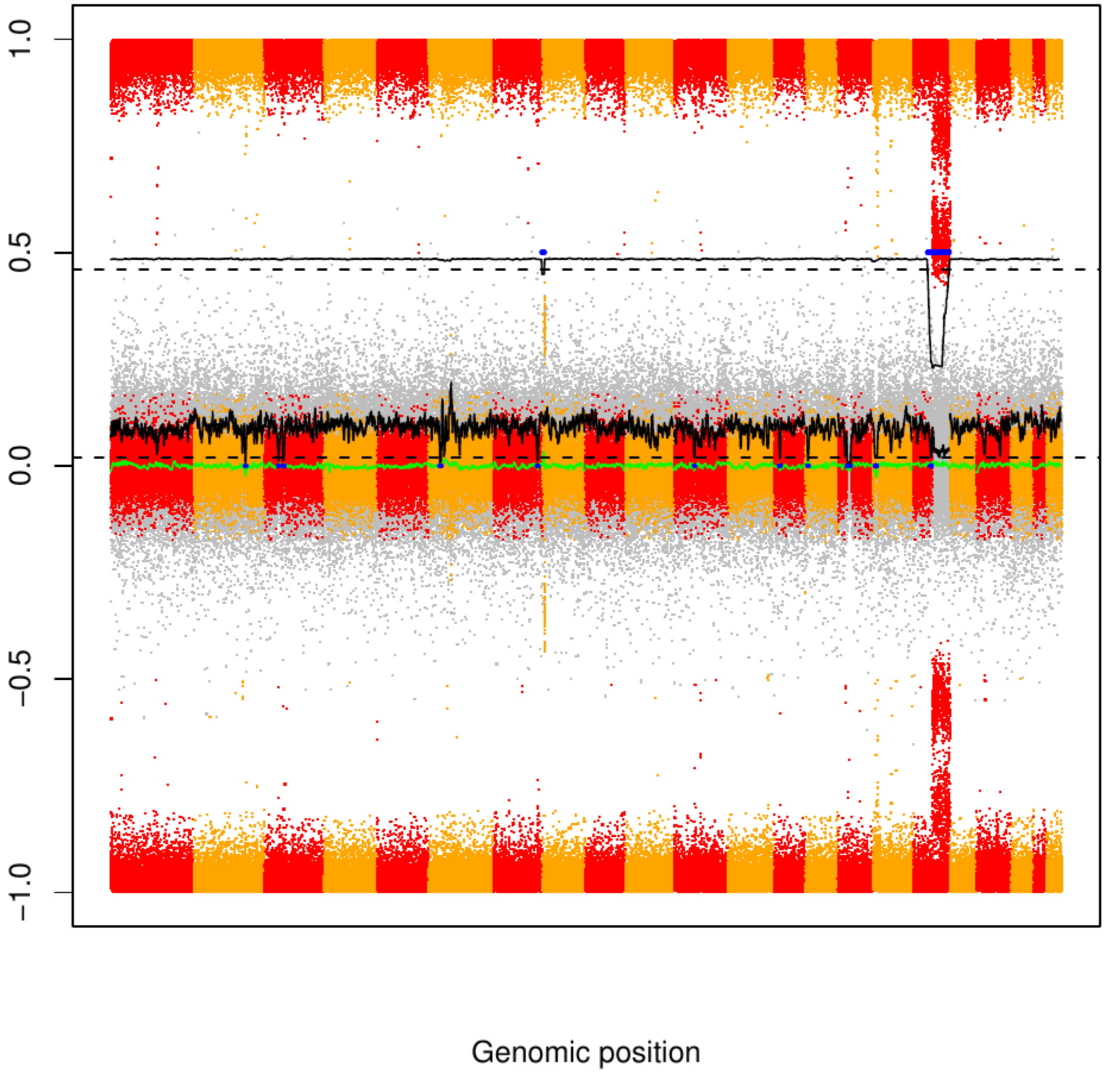

### CME17

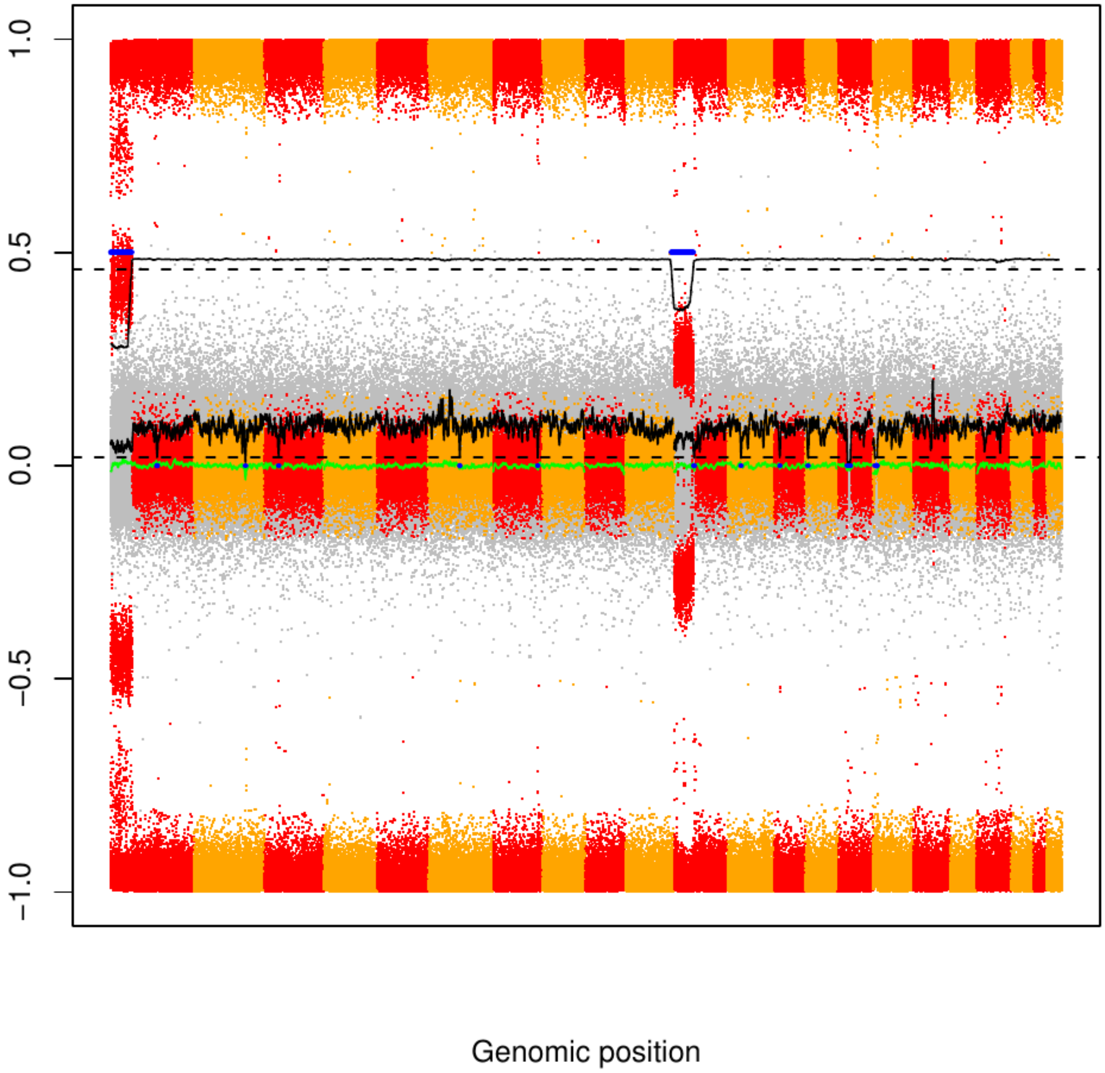

### CME18

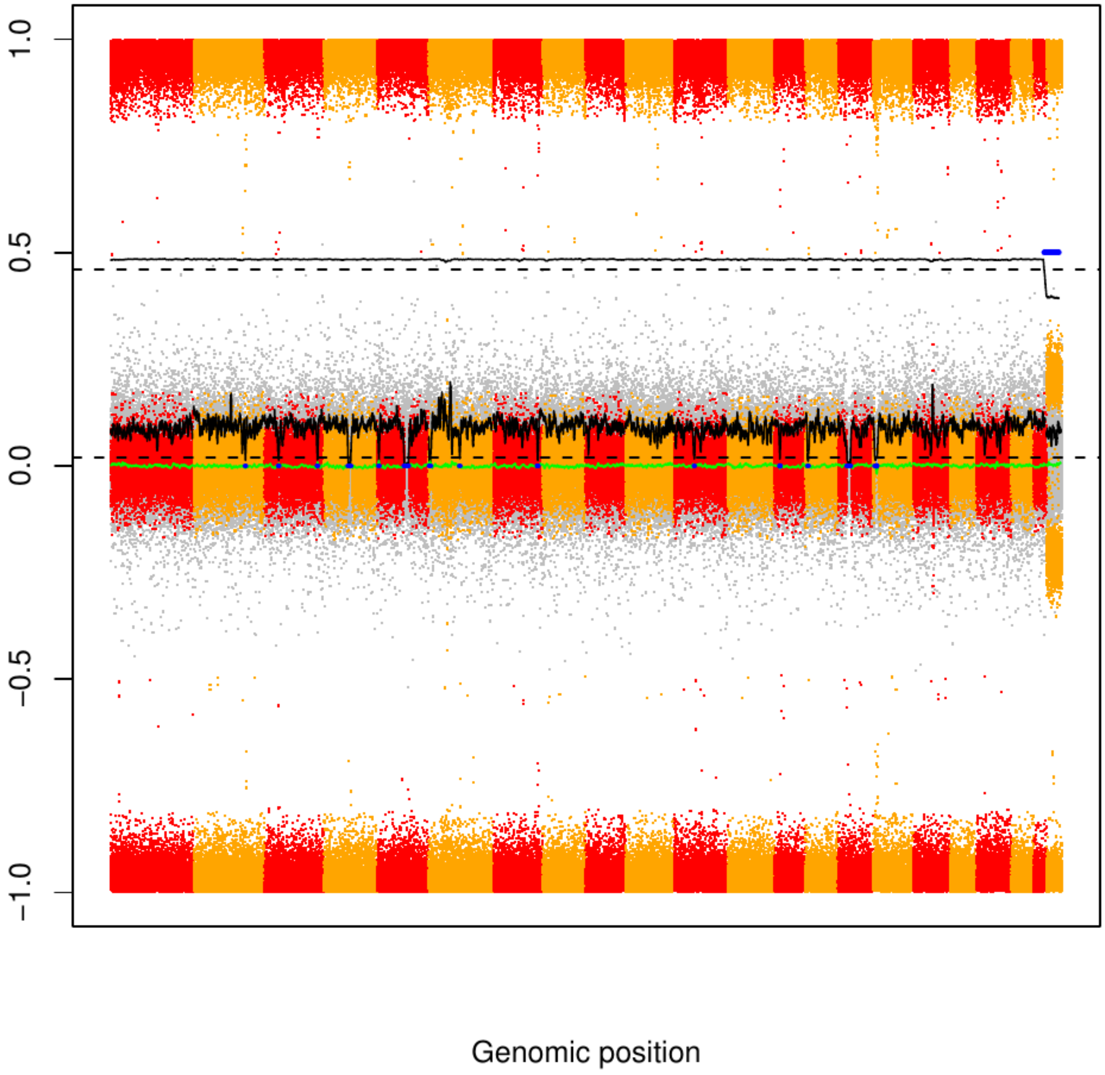

### CME19

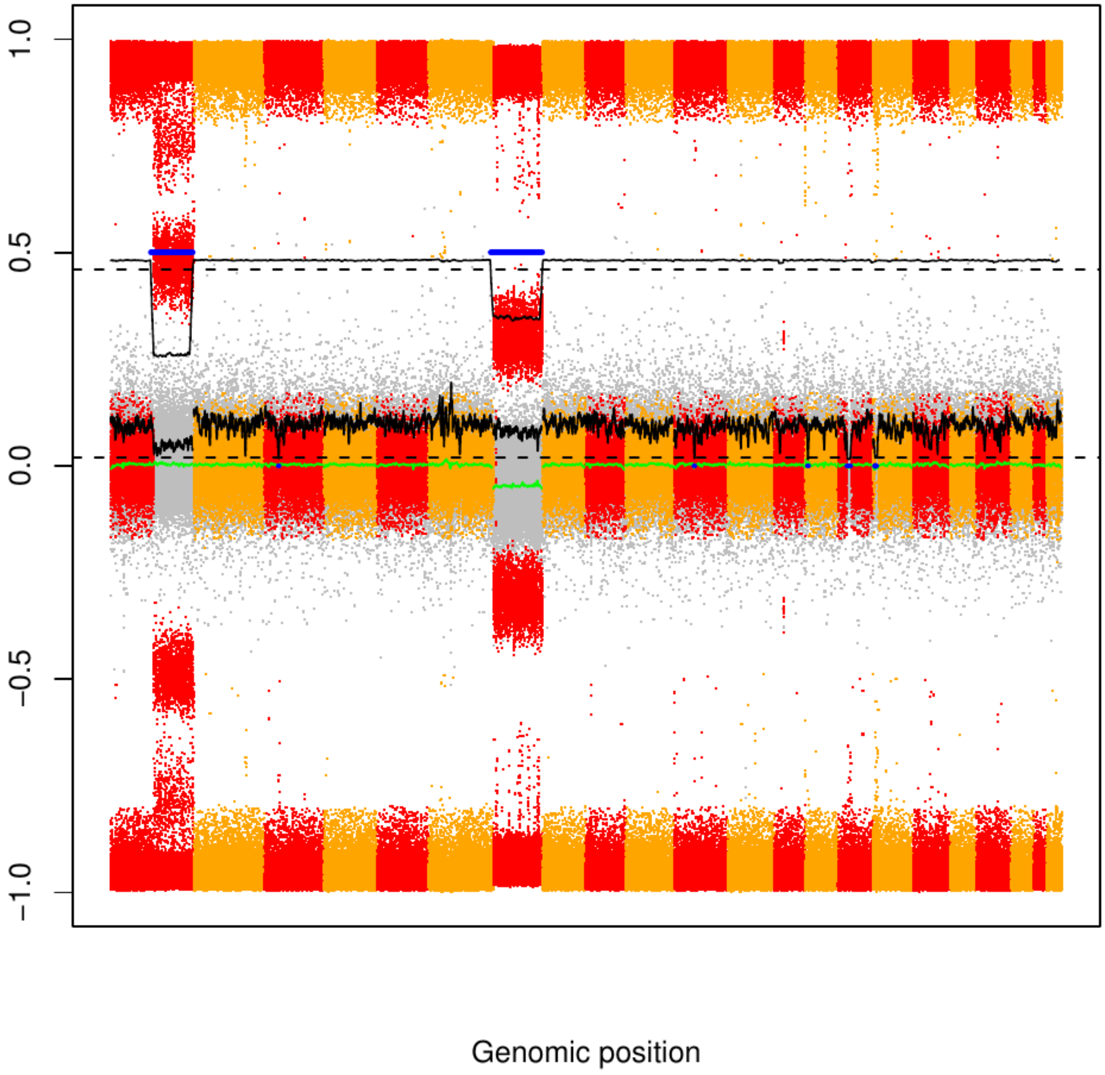

### CME20

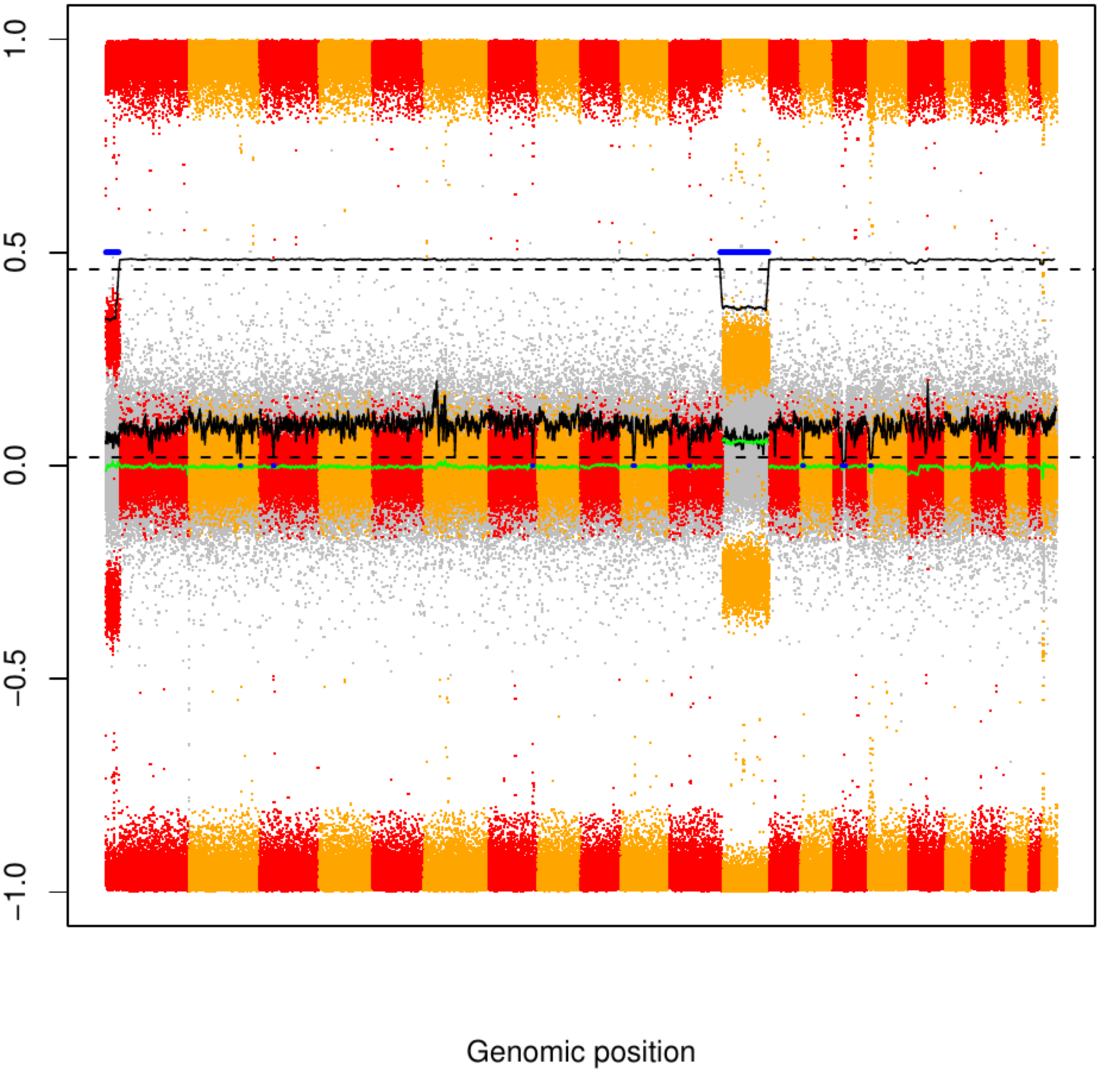

CME21

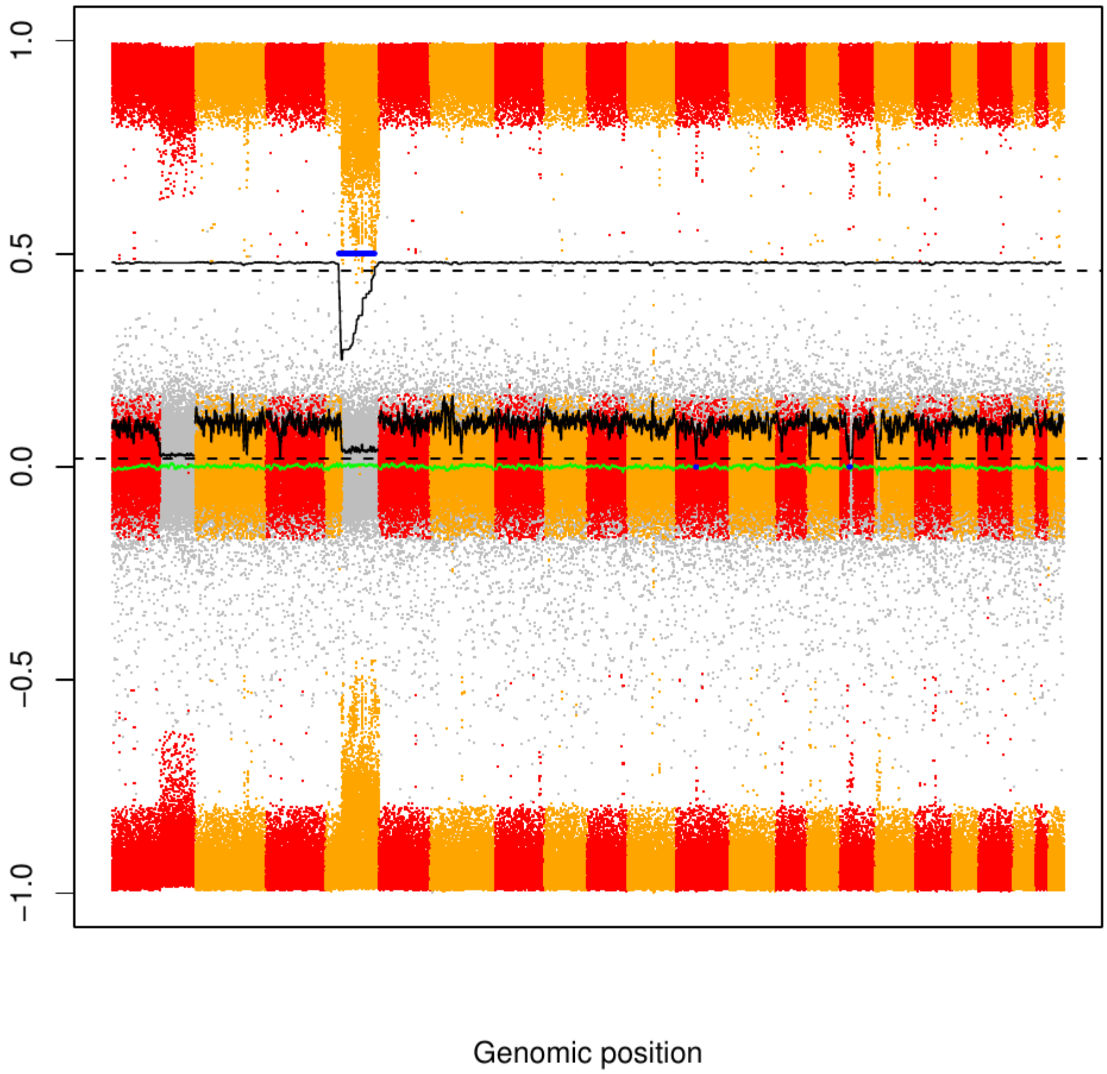

CME22

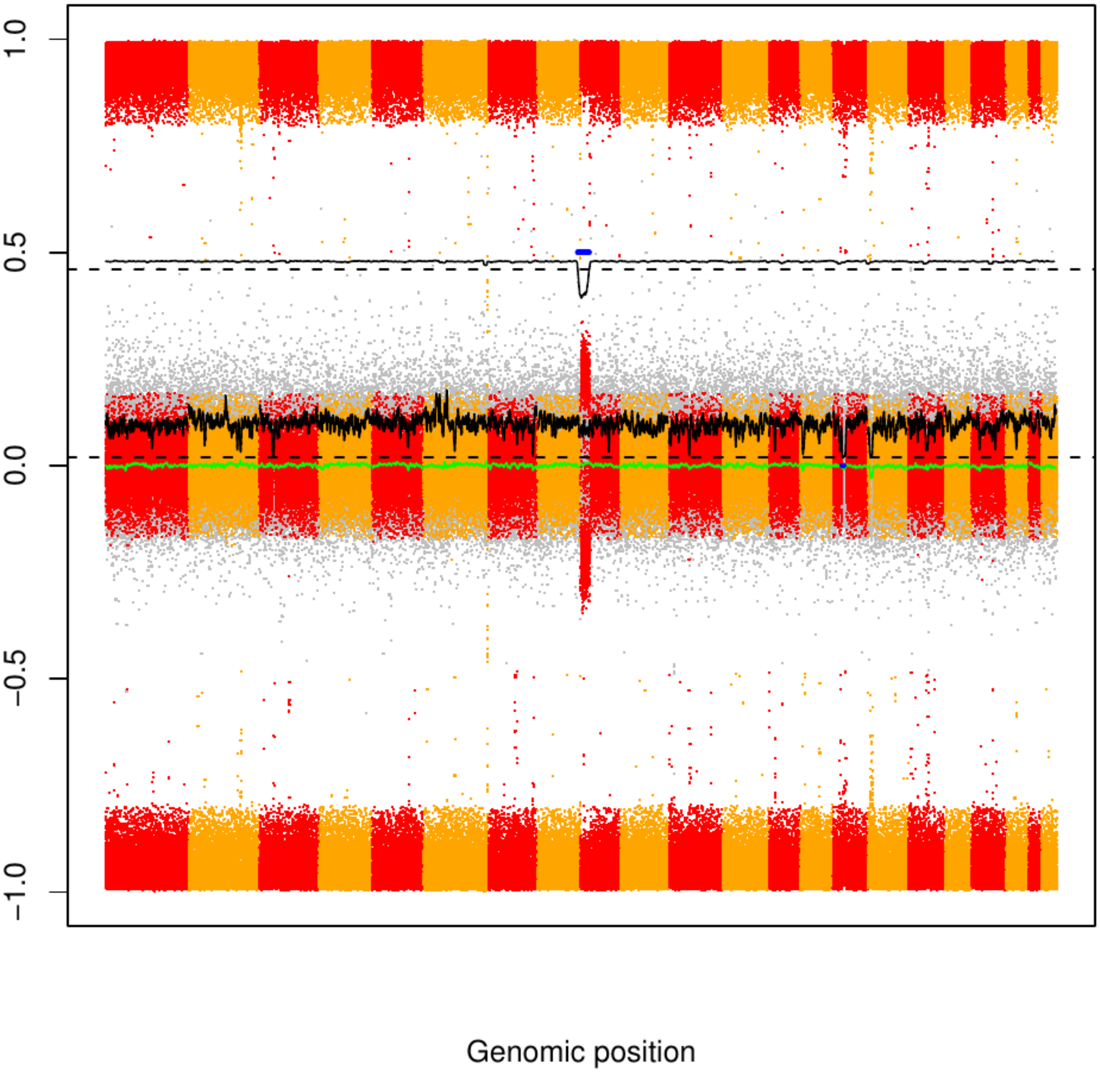

### CME23

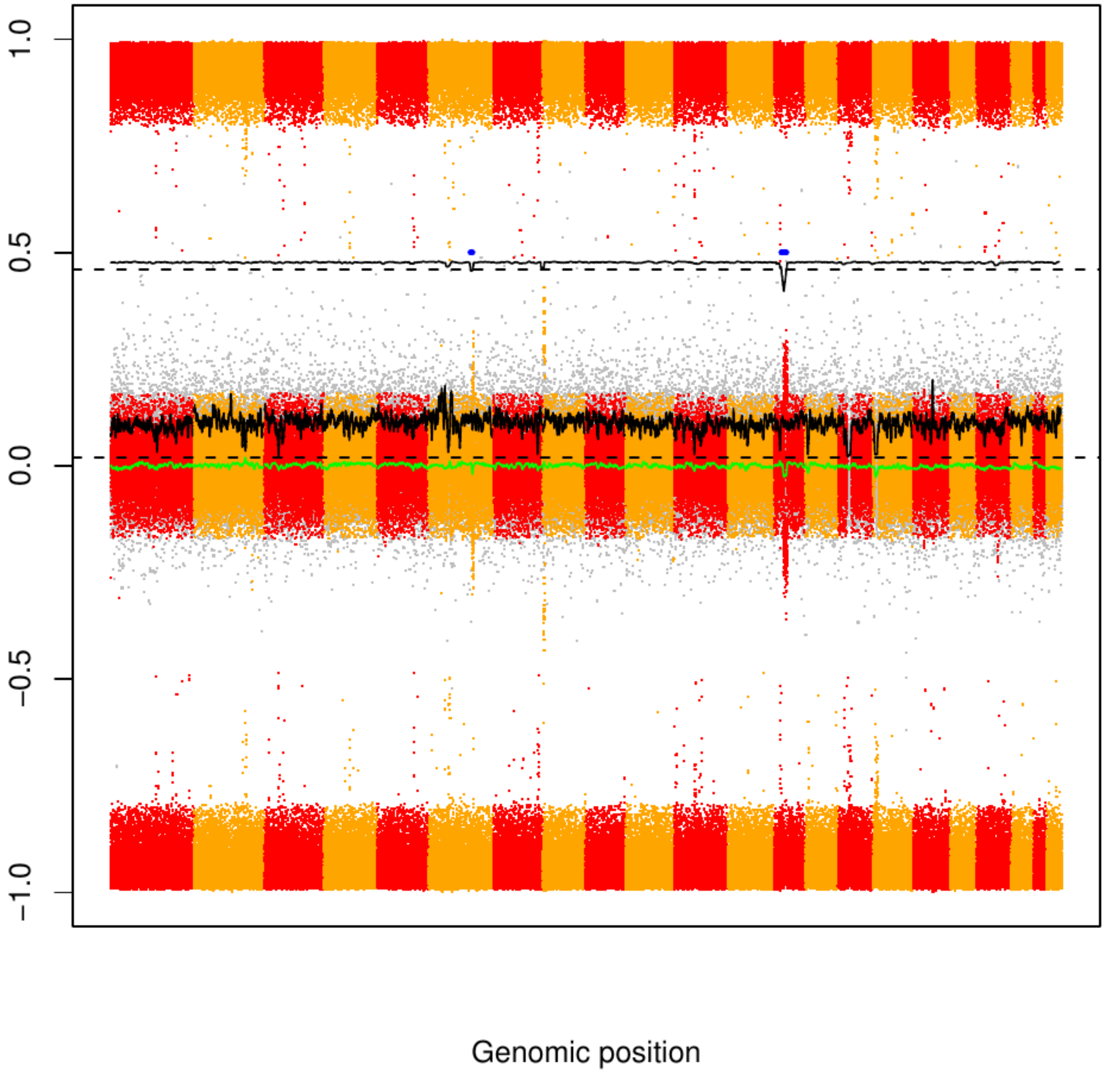

### CME24

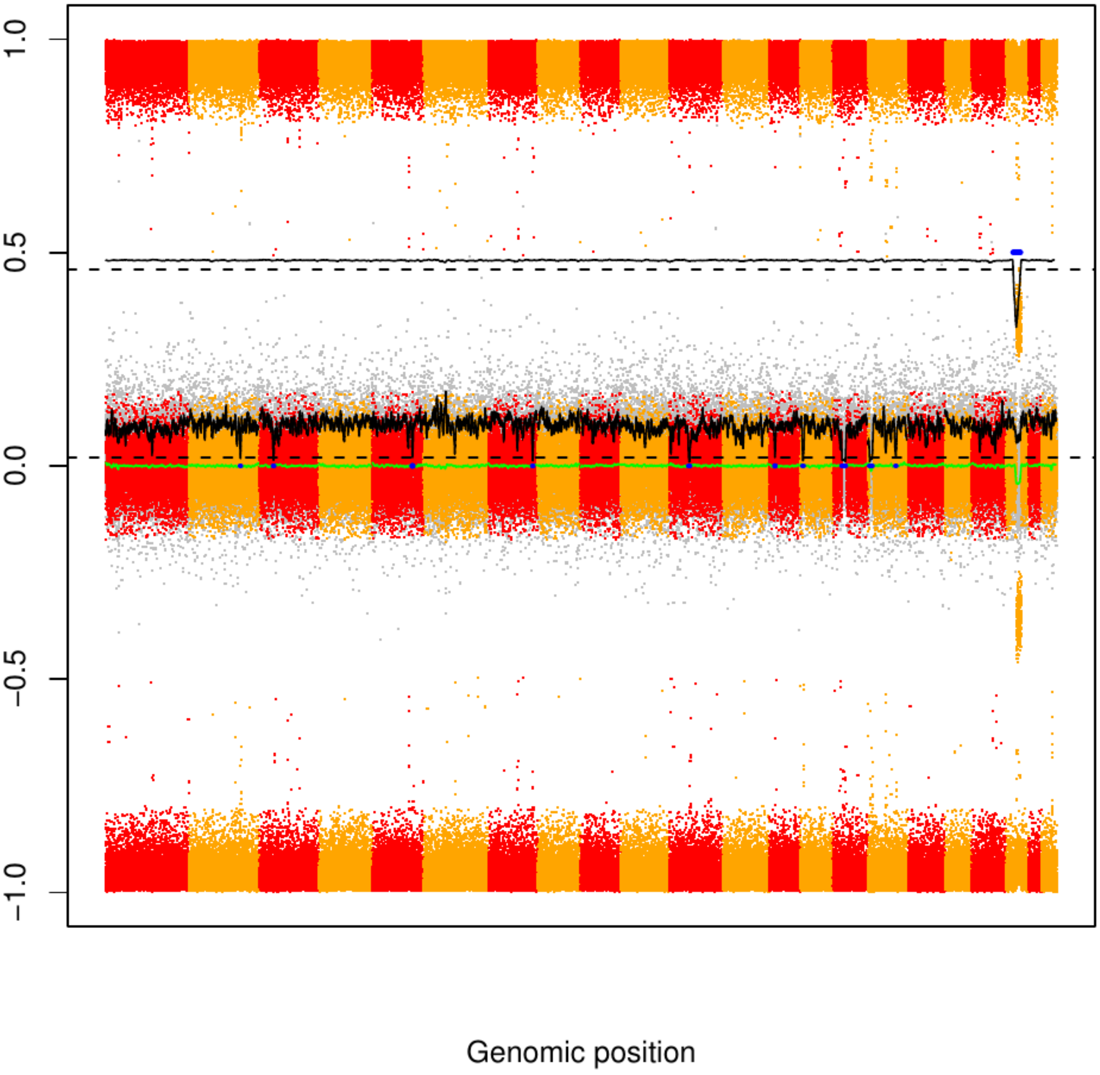

### CME25

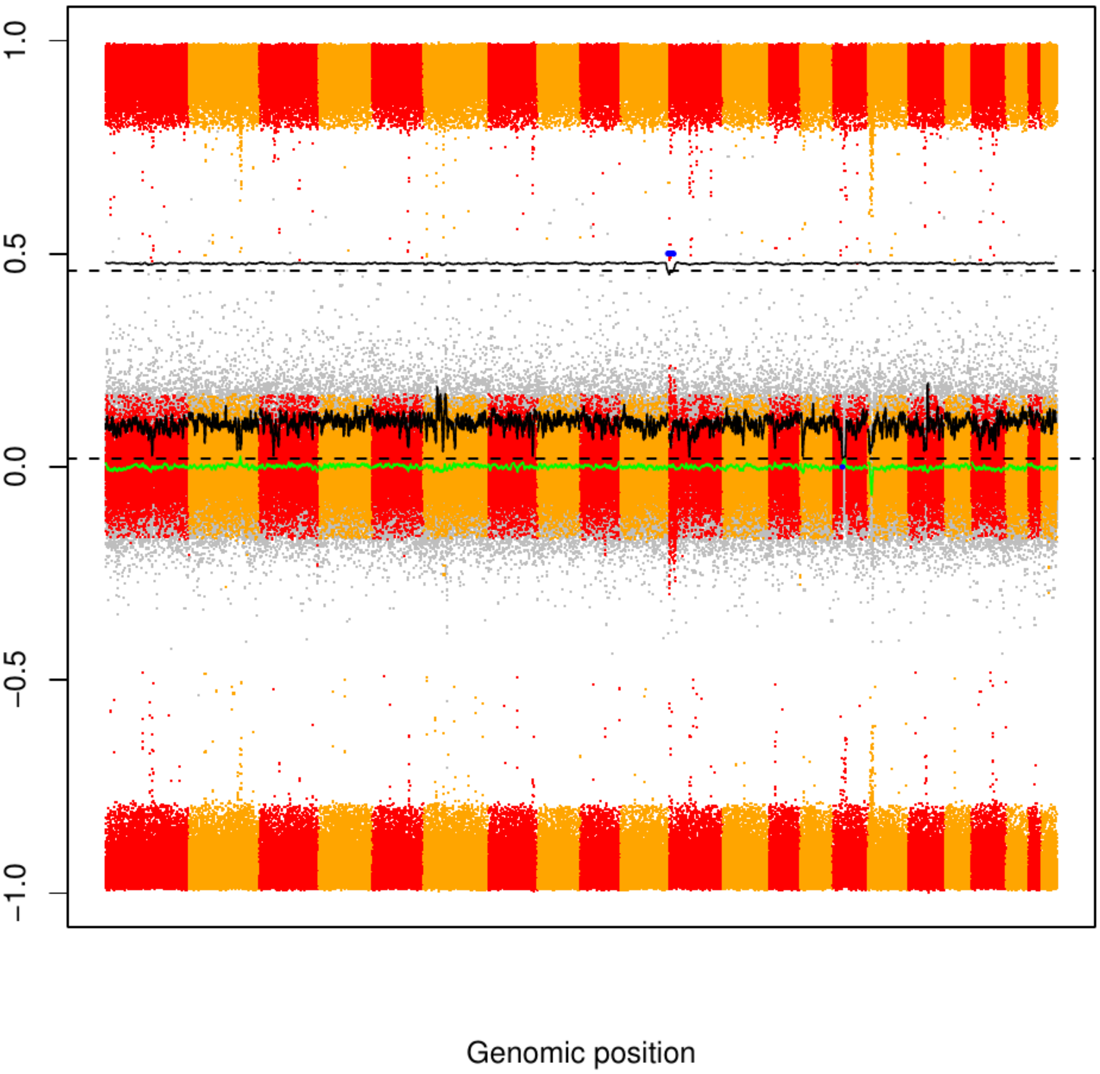

### CME26

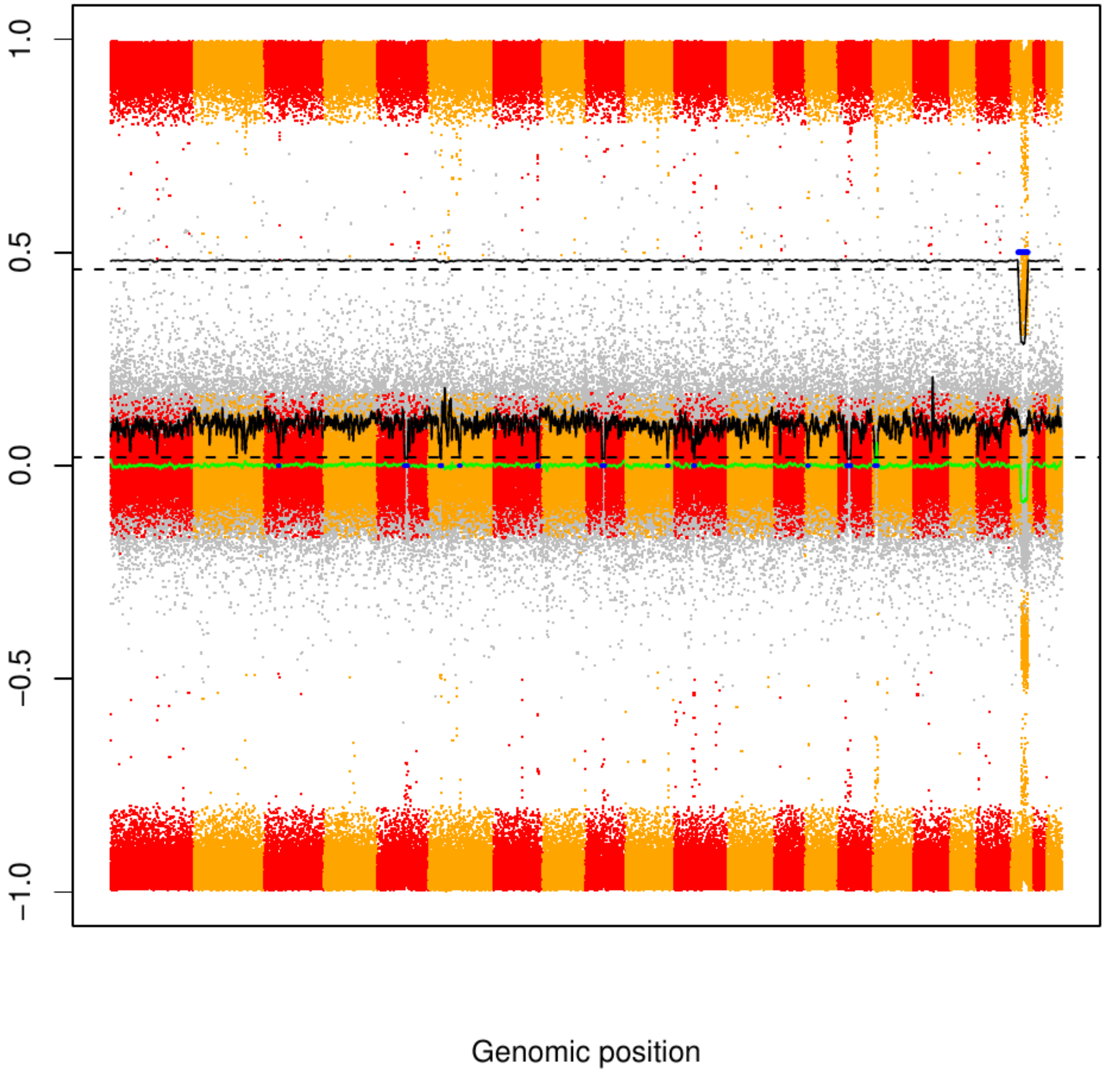

### CME27

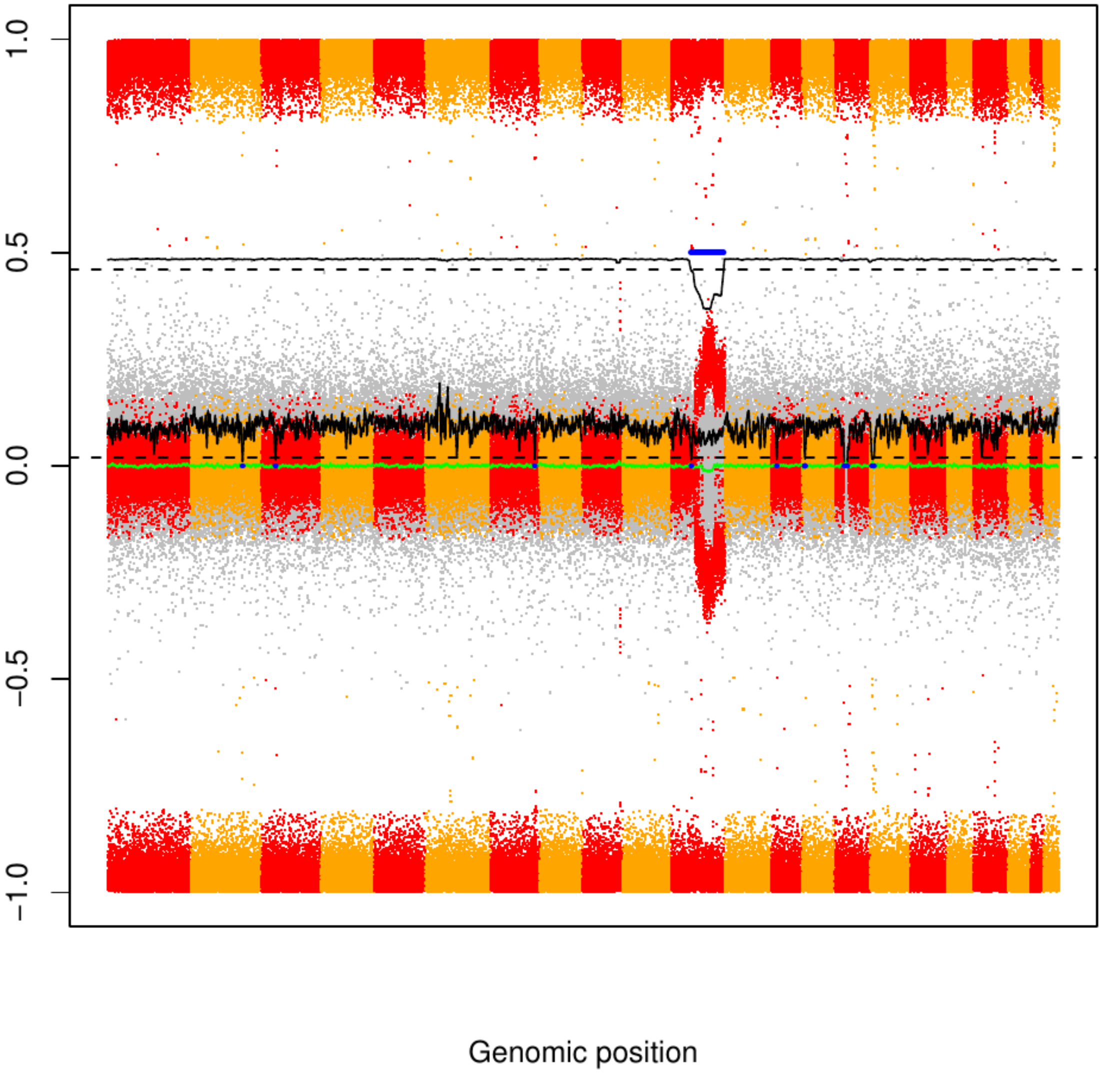

CME28

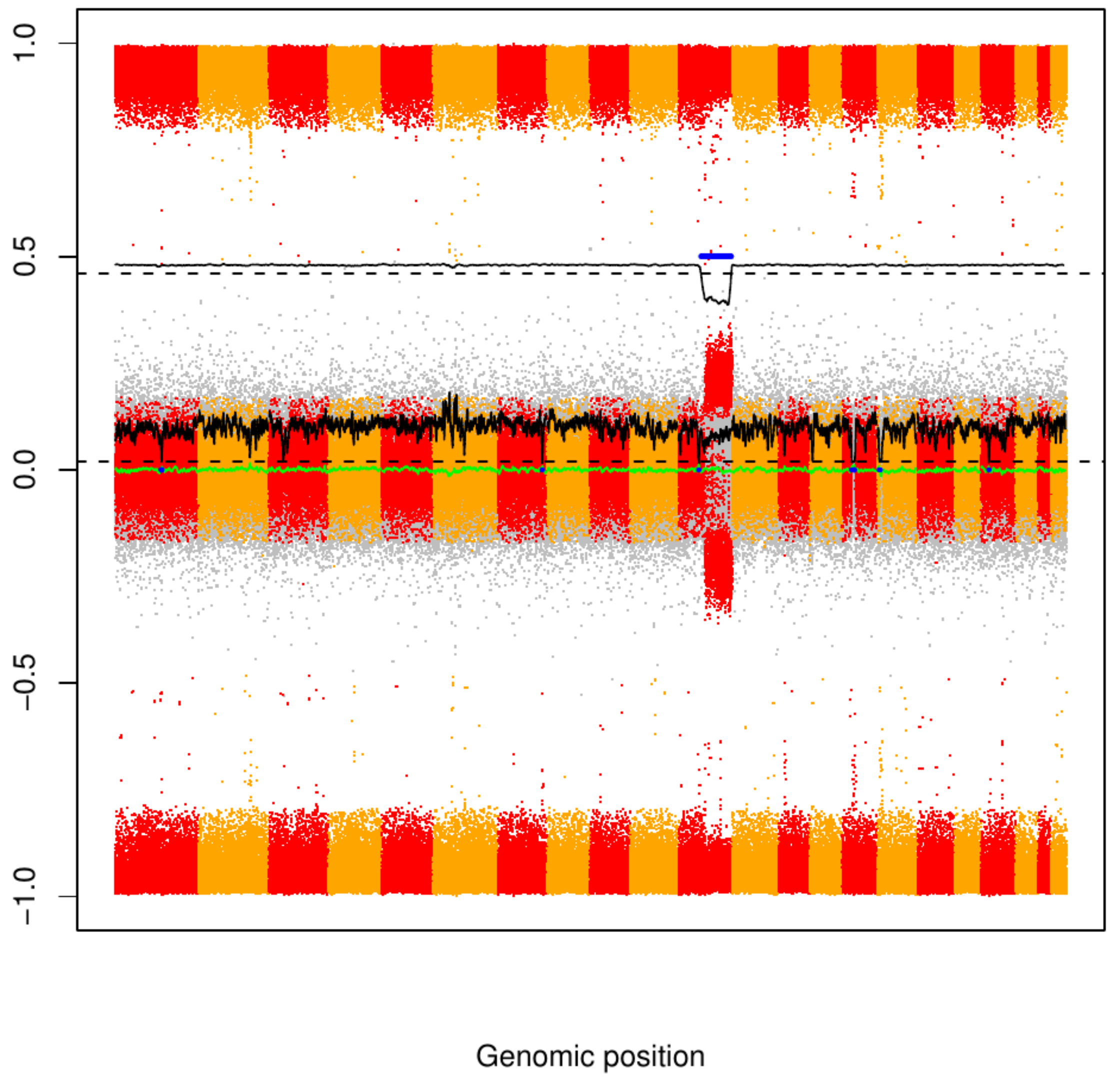

CME29

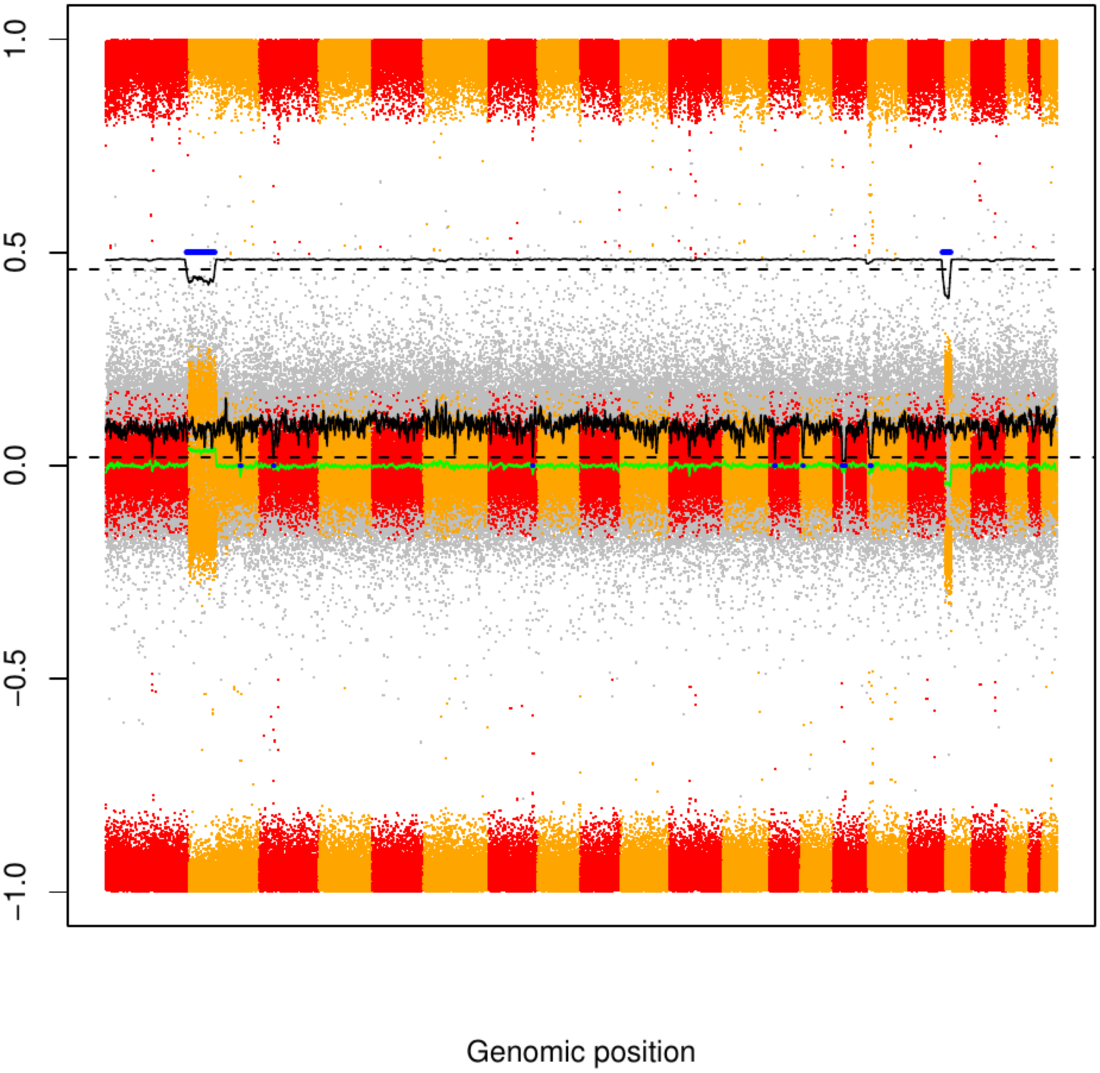

### CME30

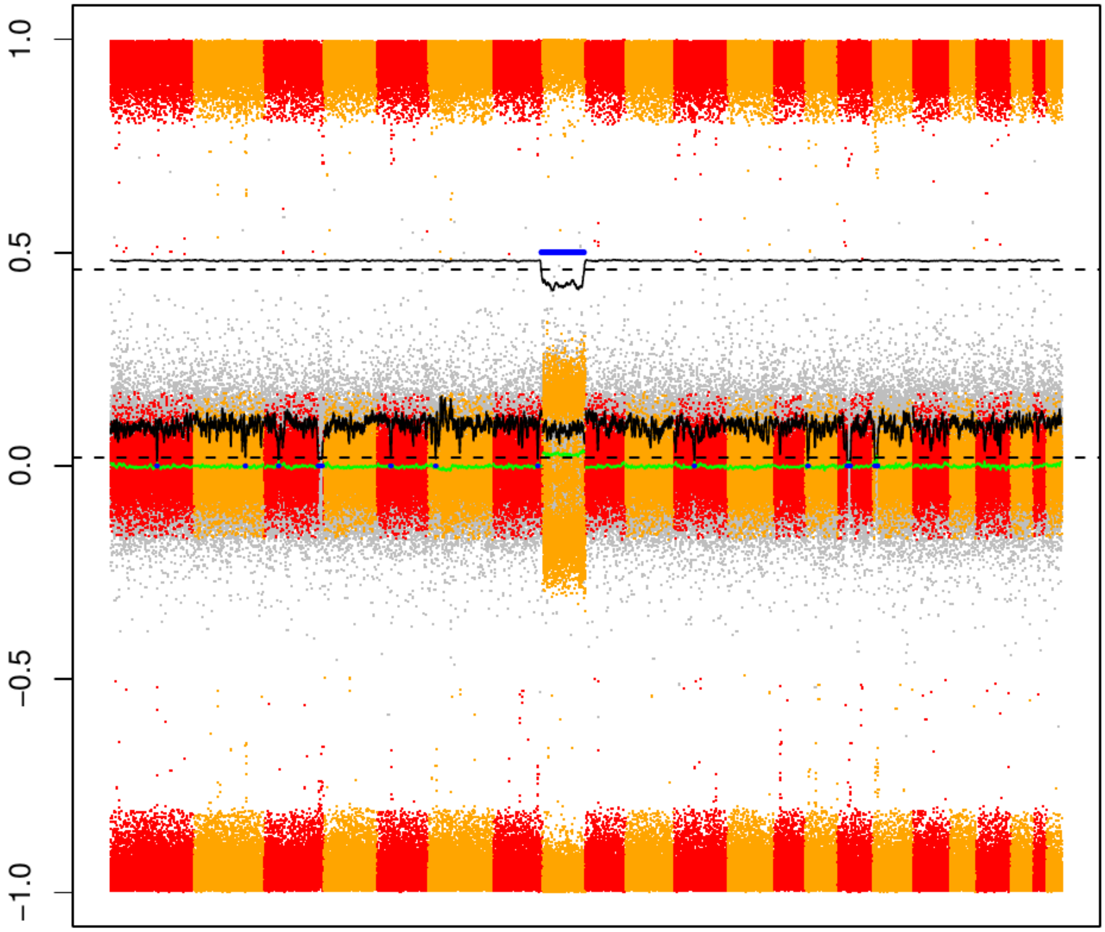


Genomic position

### CME31

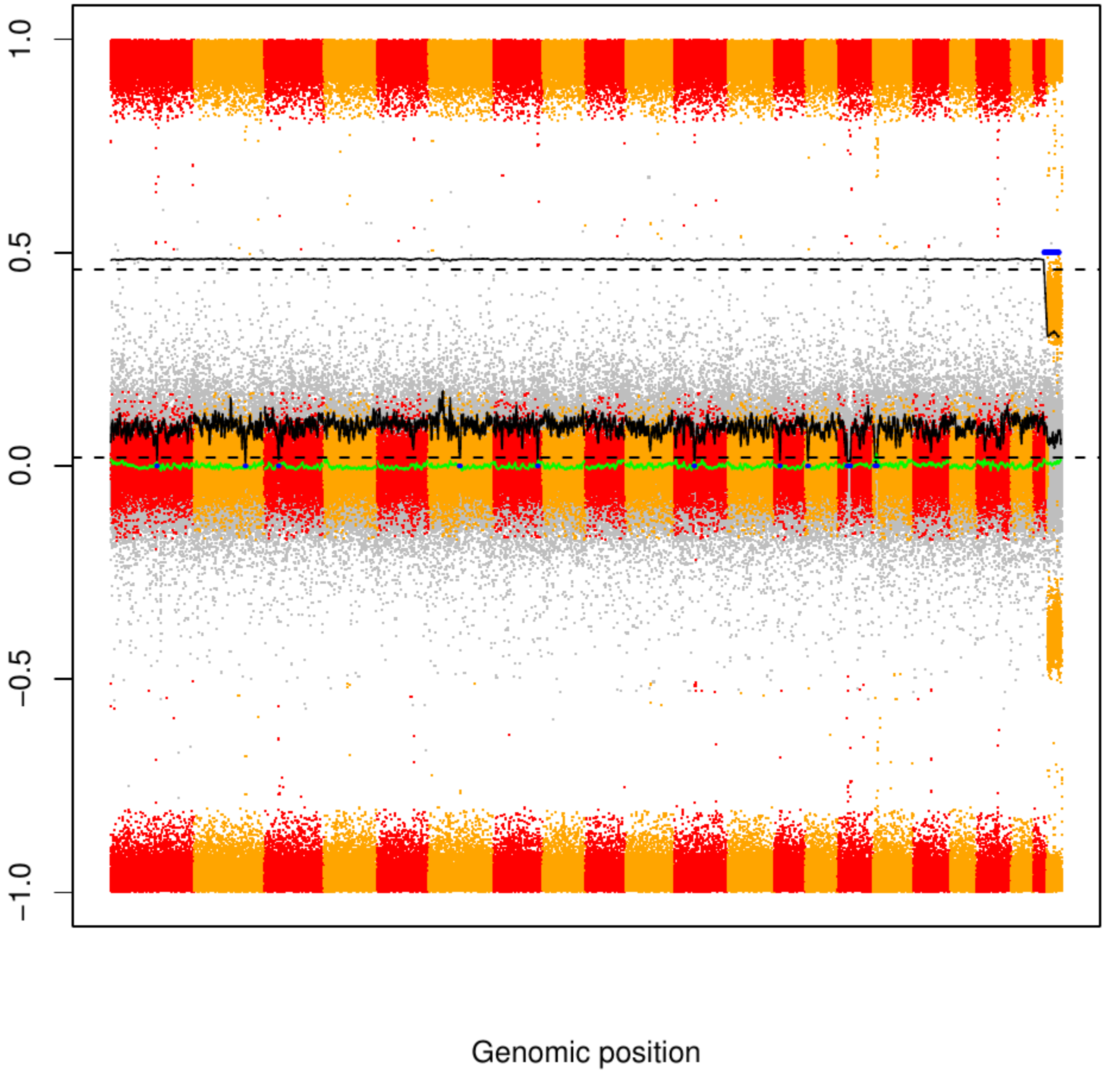

CME32

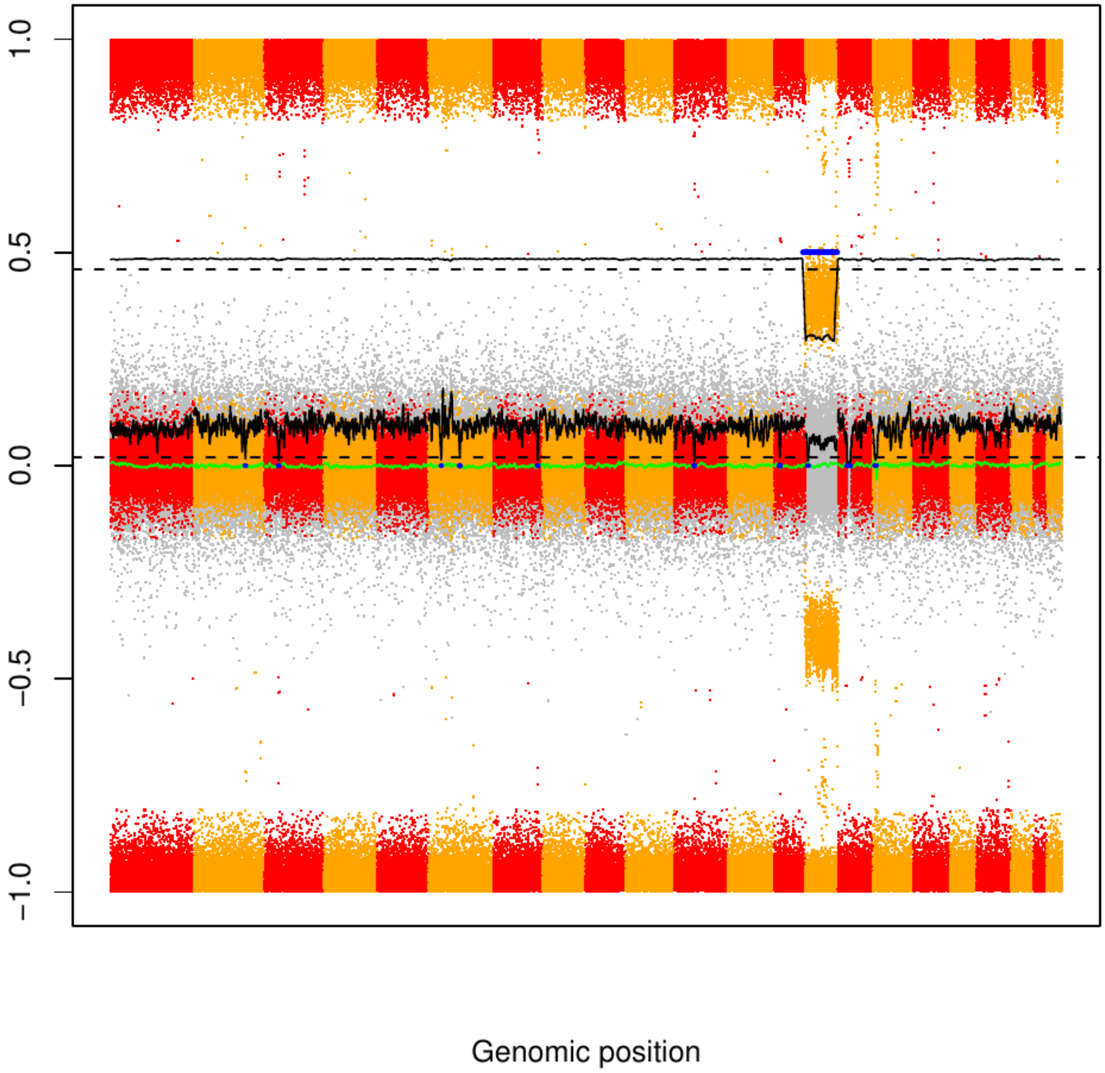

### CME33

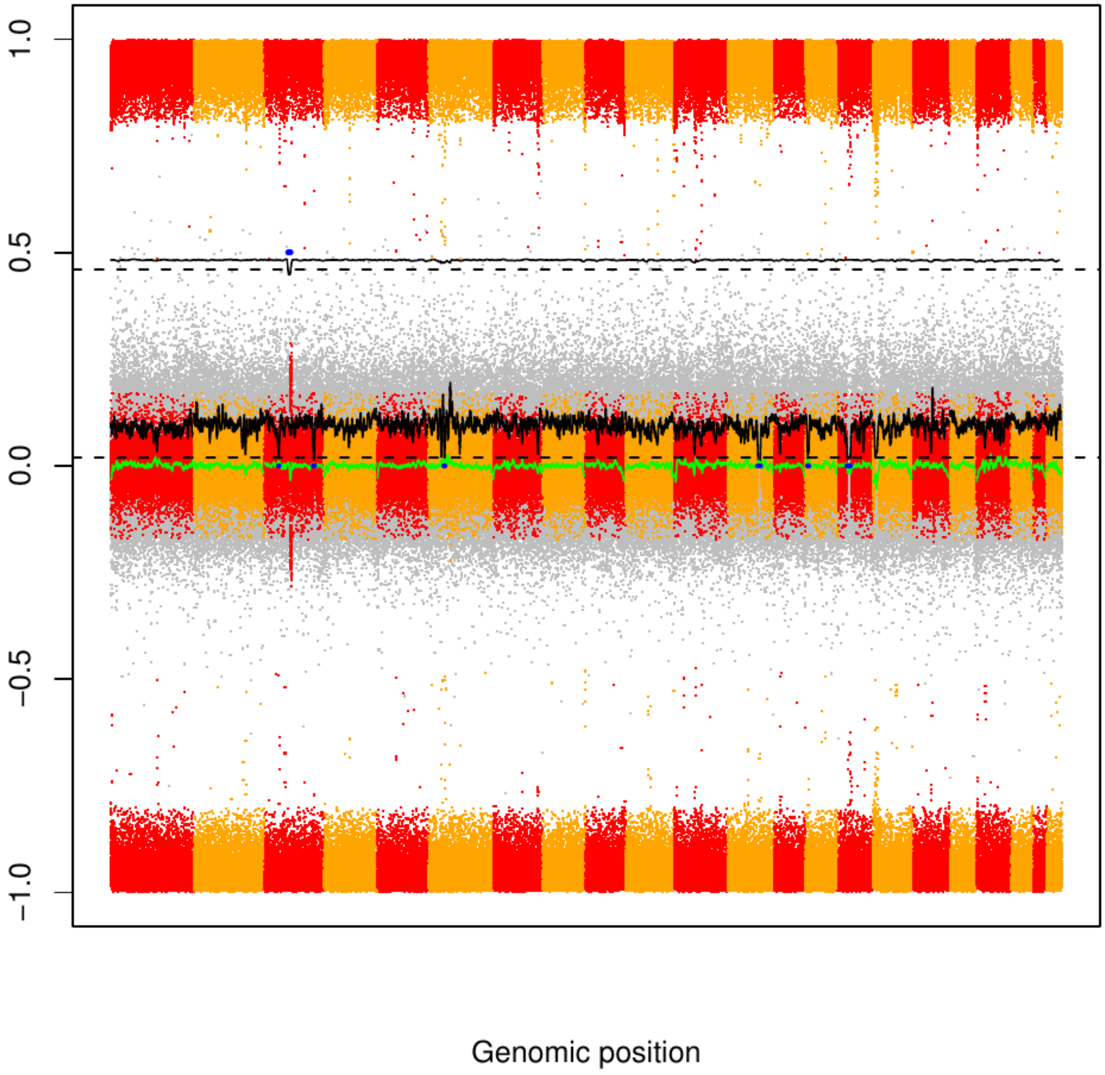

### CME34

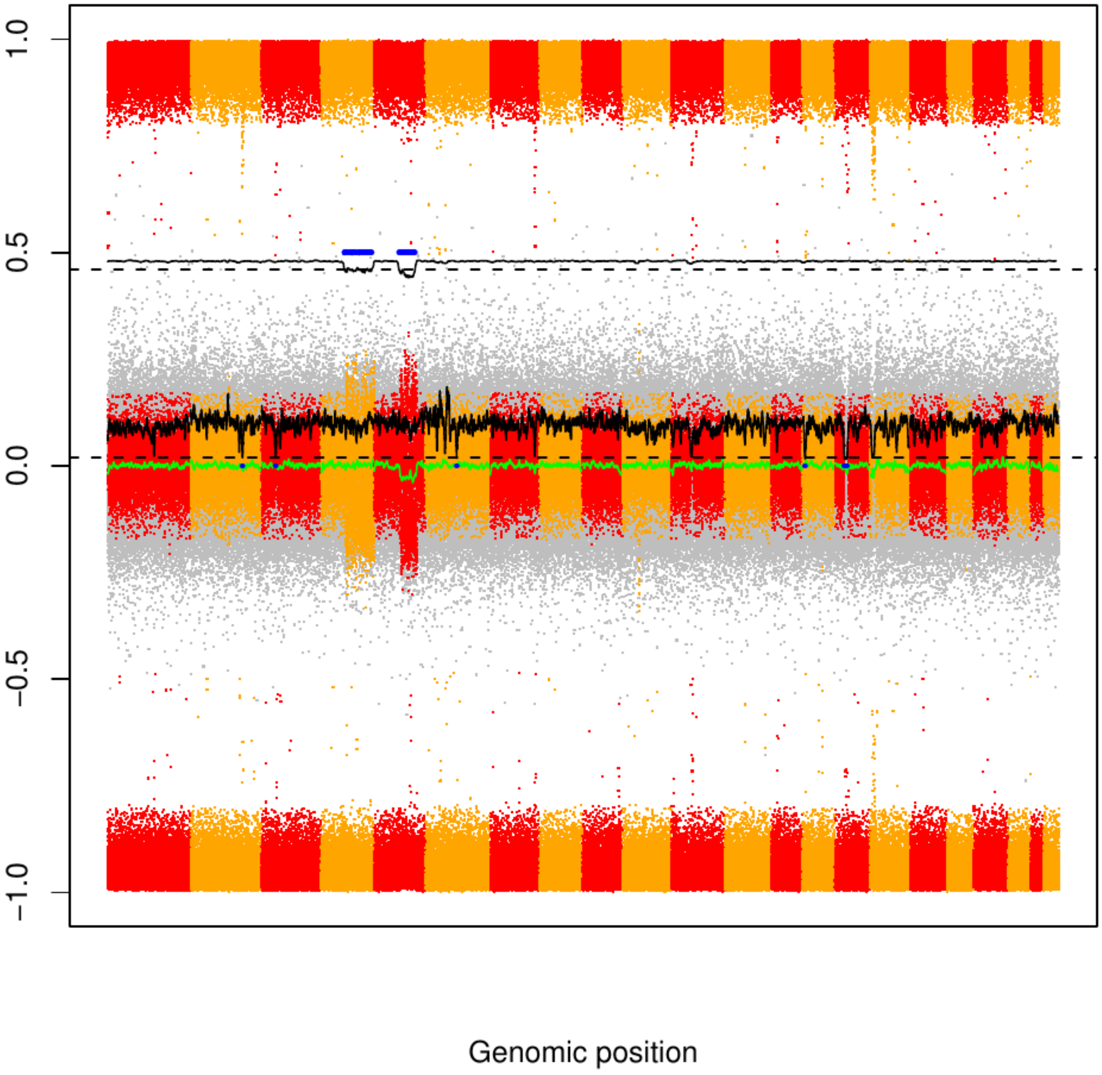

### CME35

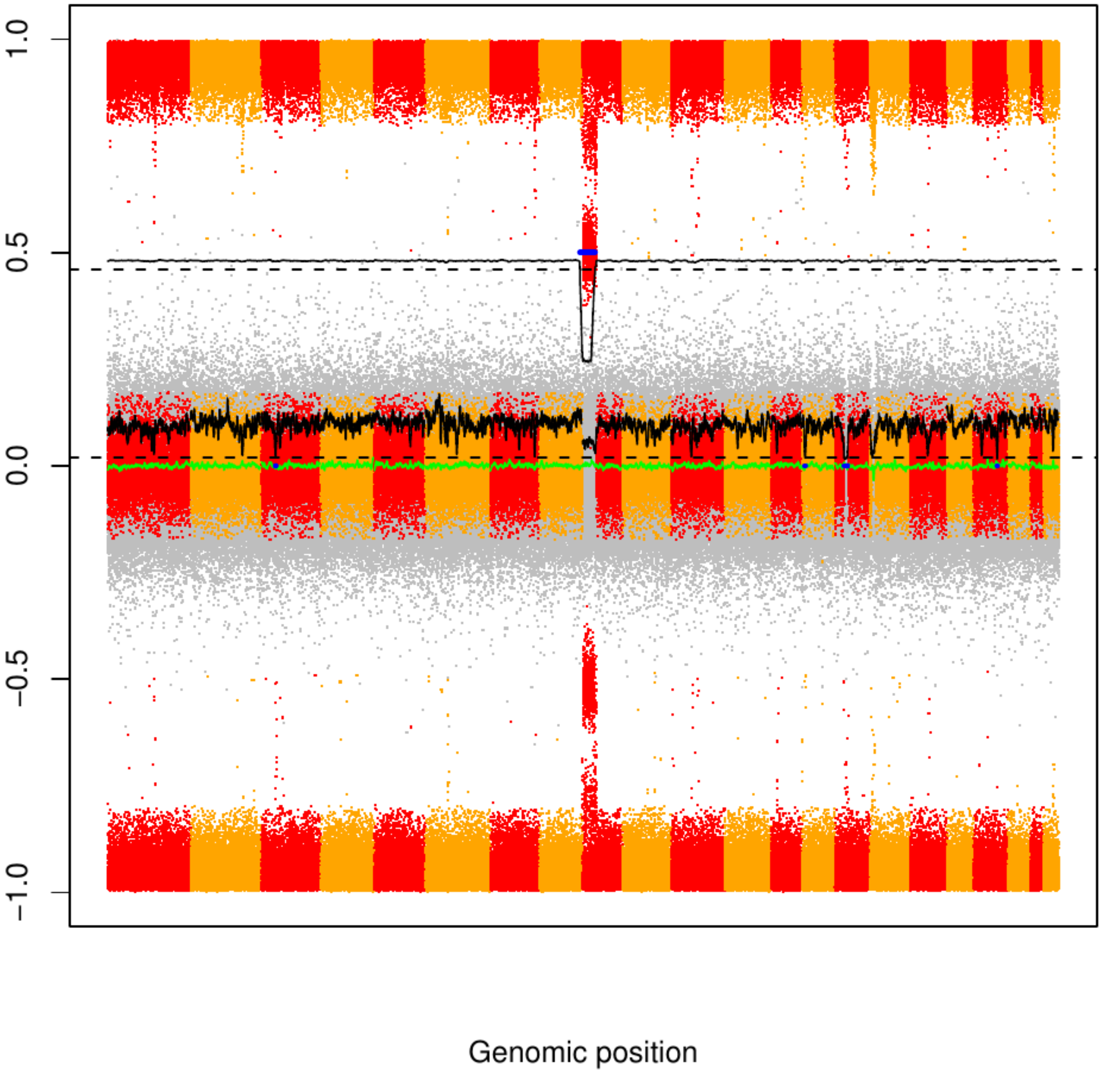

### CME36

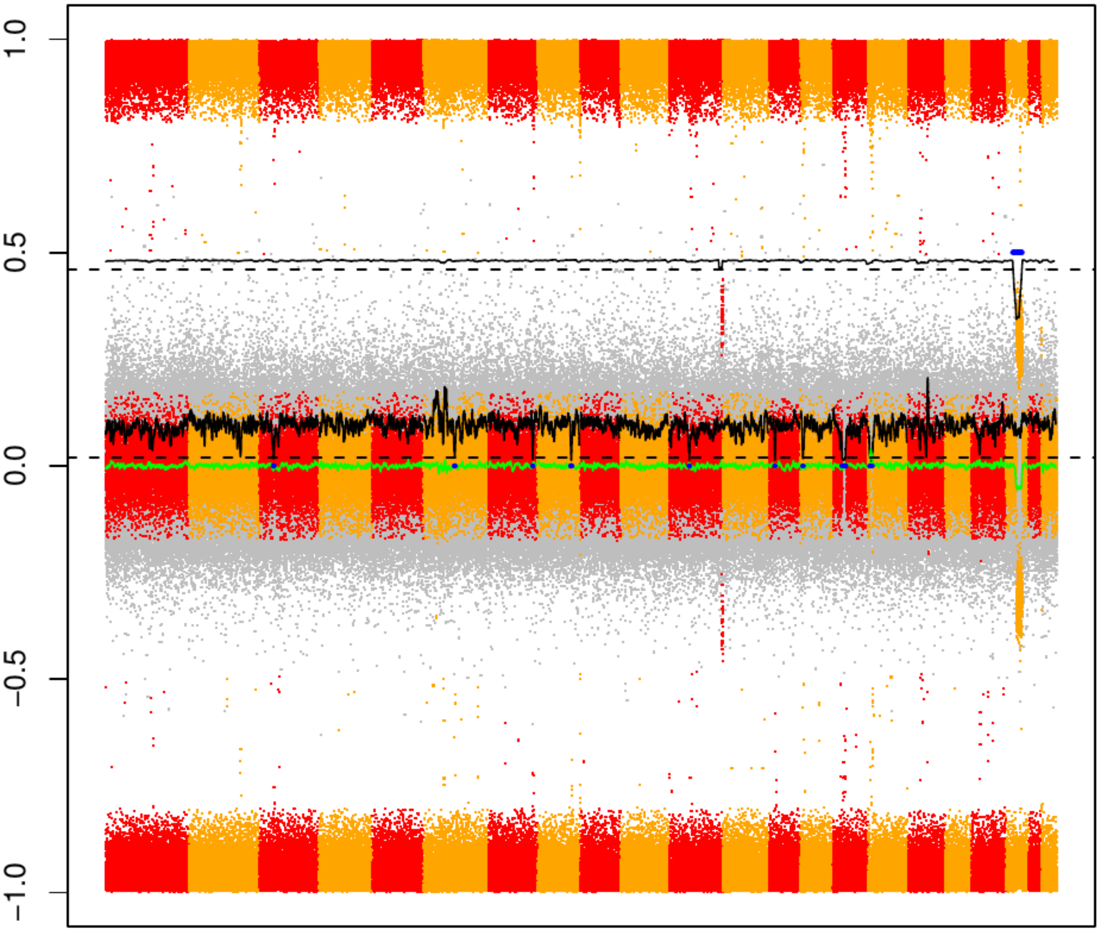

### CME37

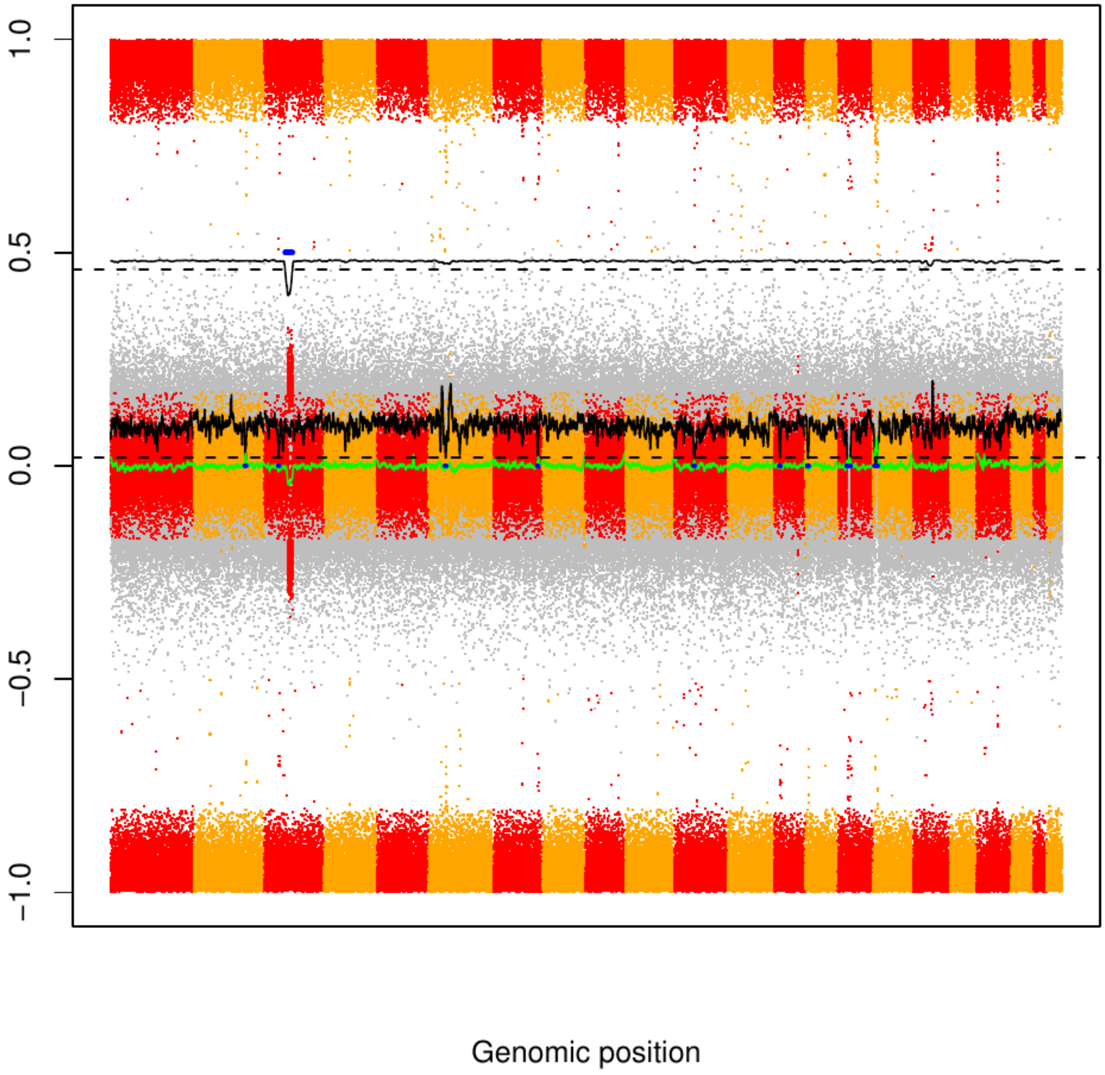

CME38

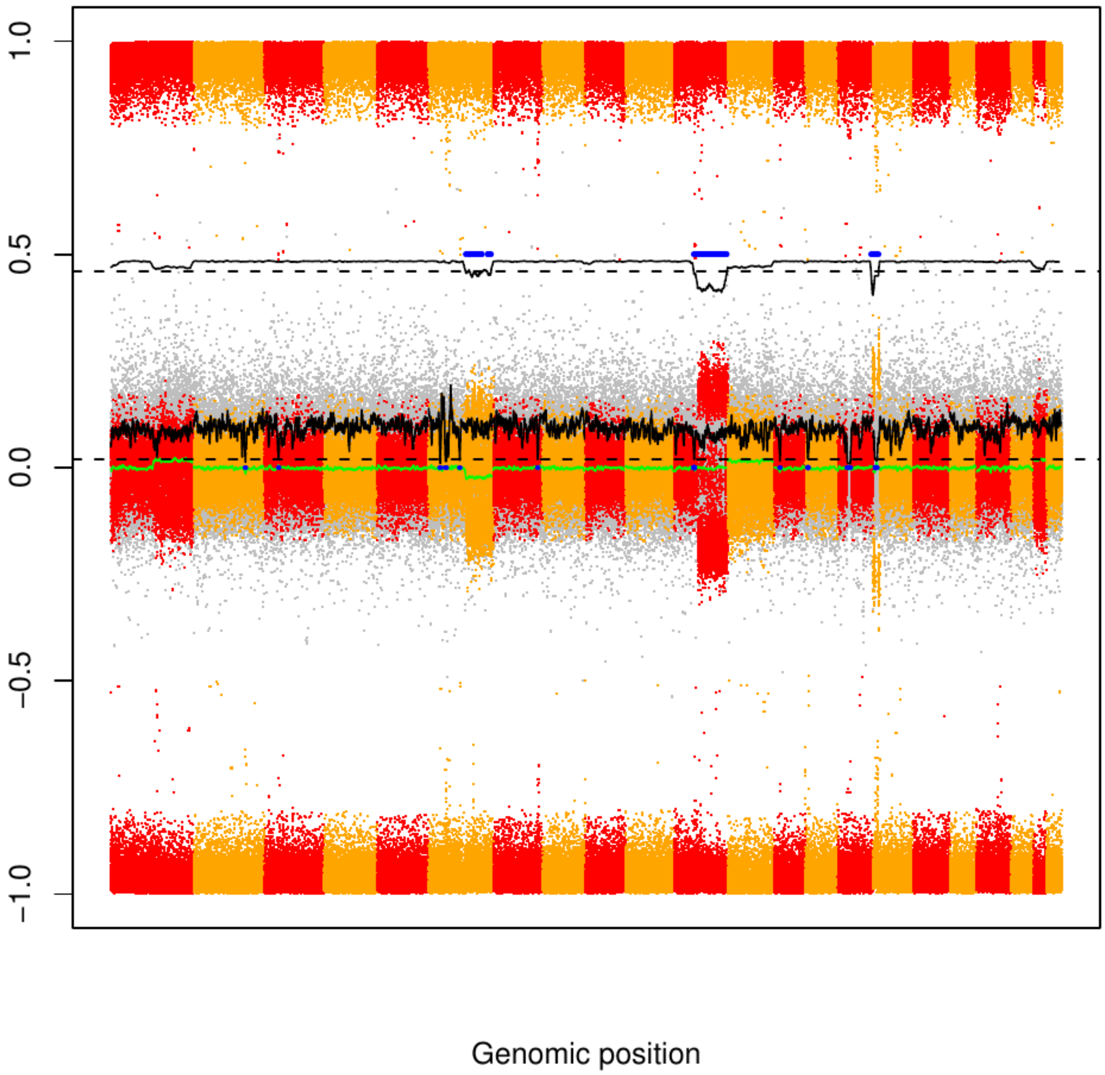

CME39

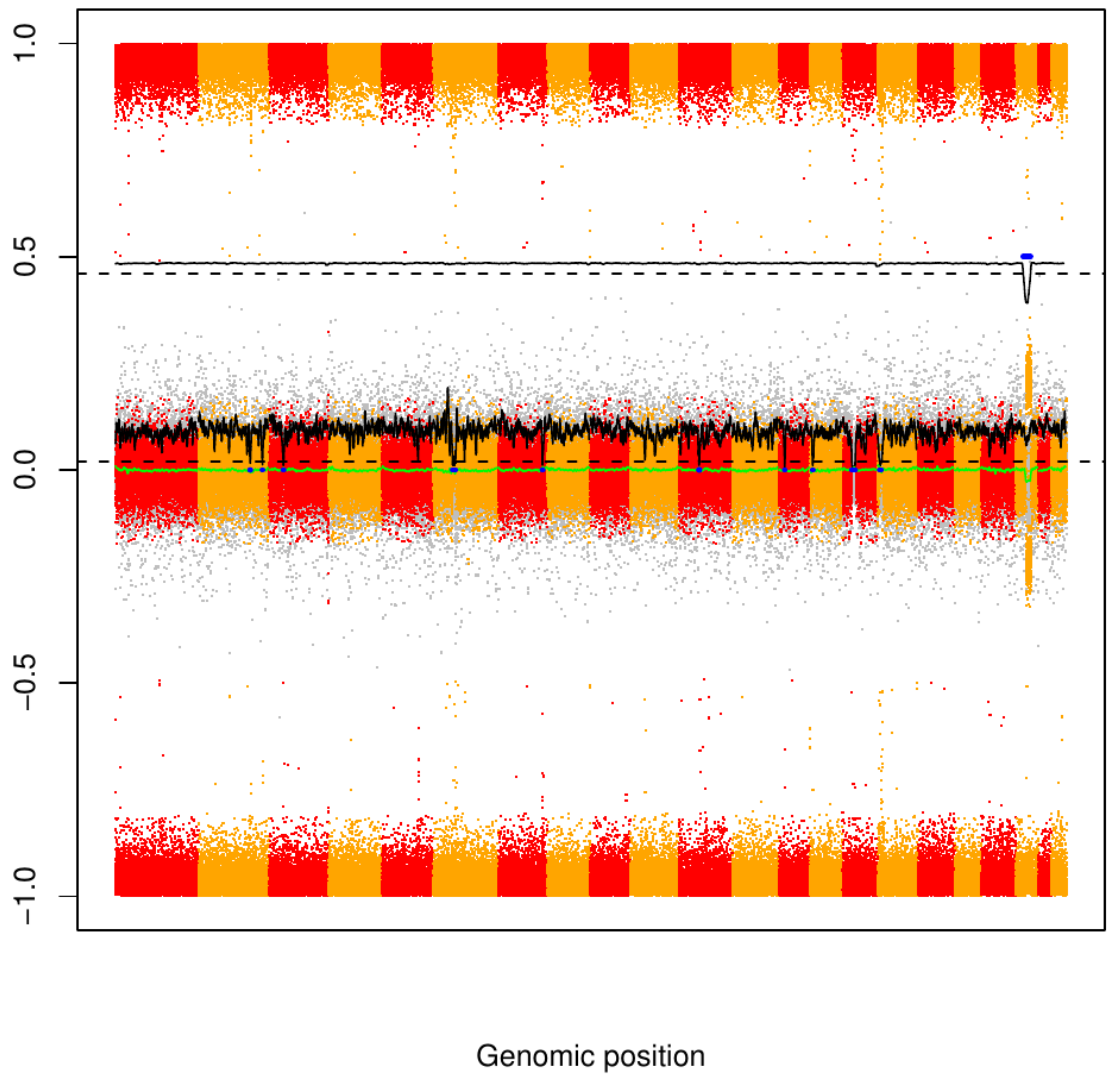

### CME40

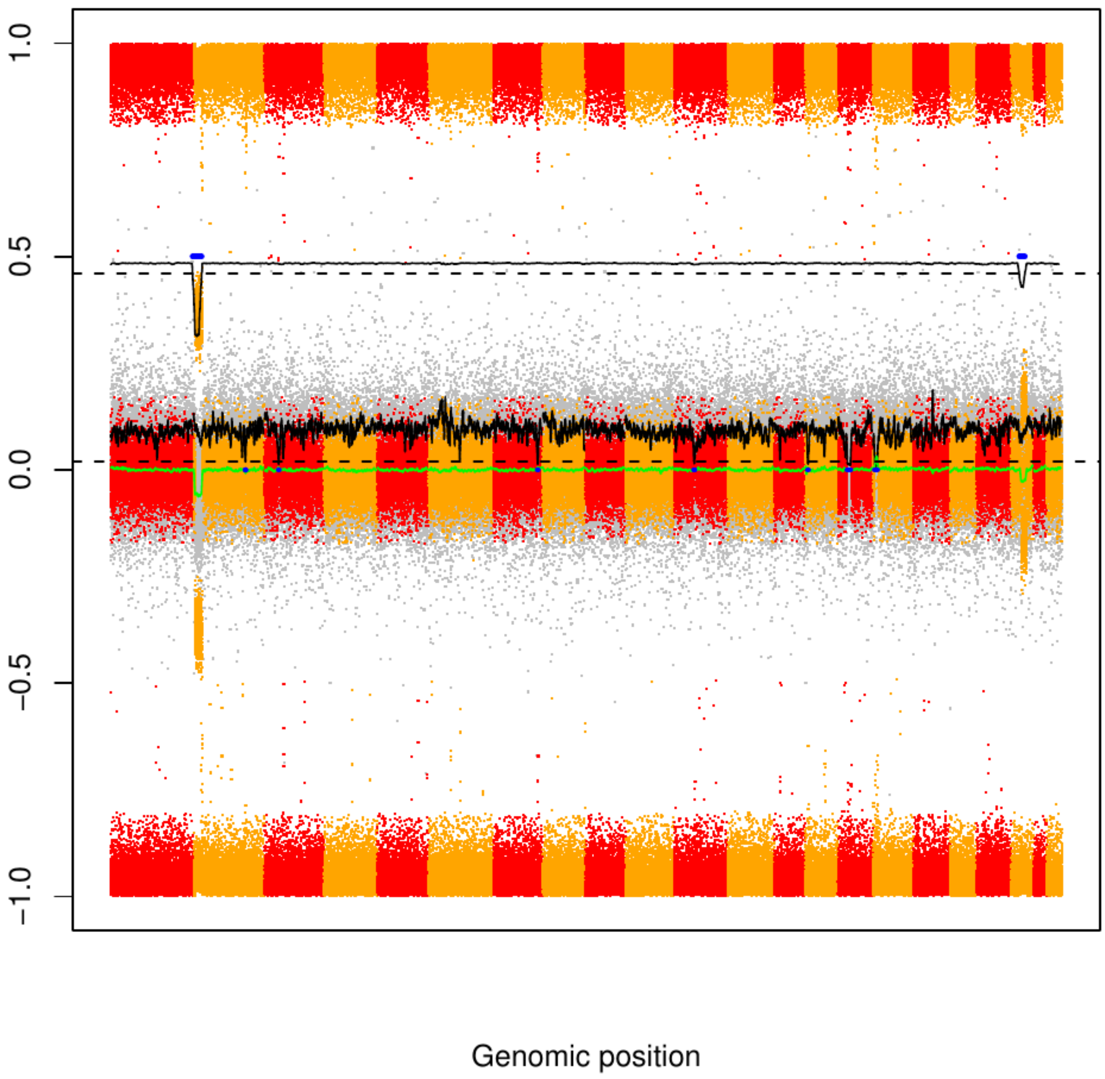

### CME41

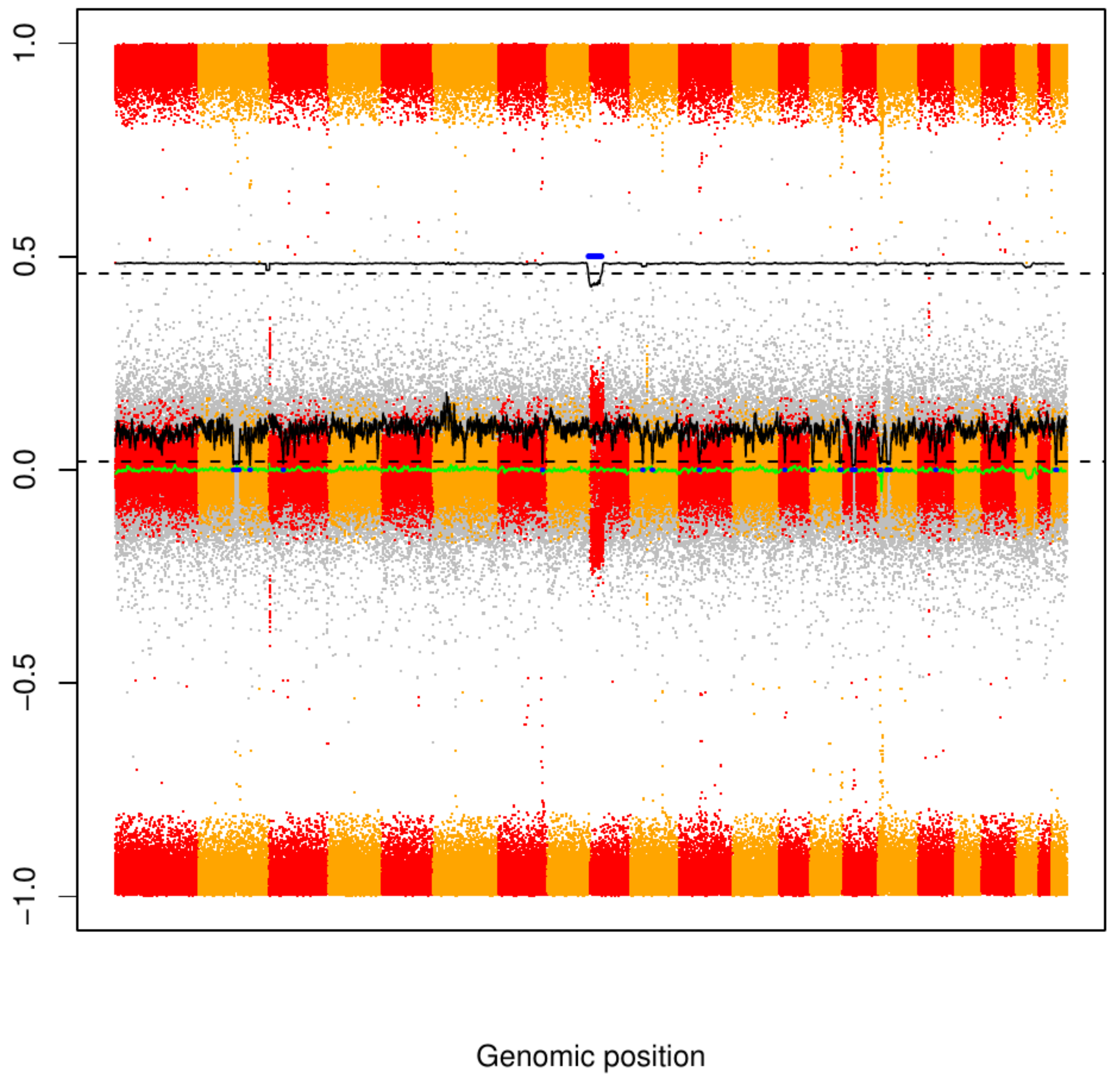

### CME42

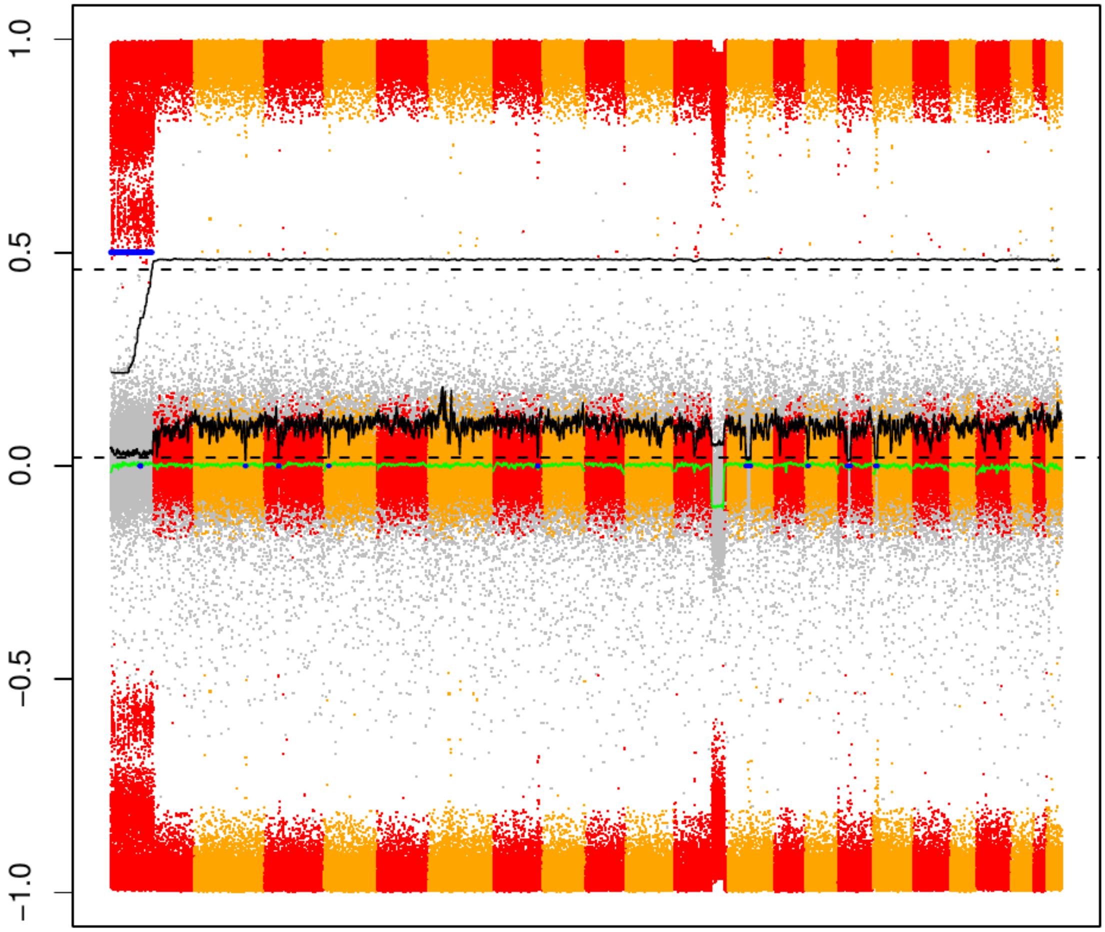

### CME43

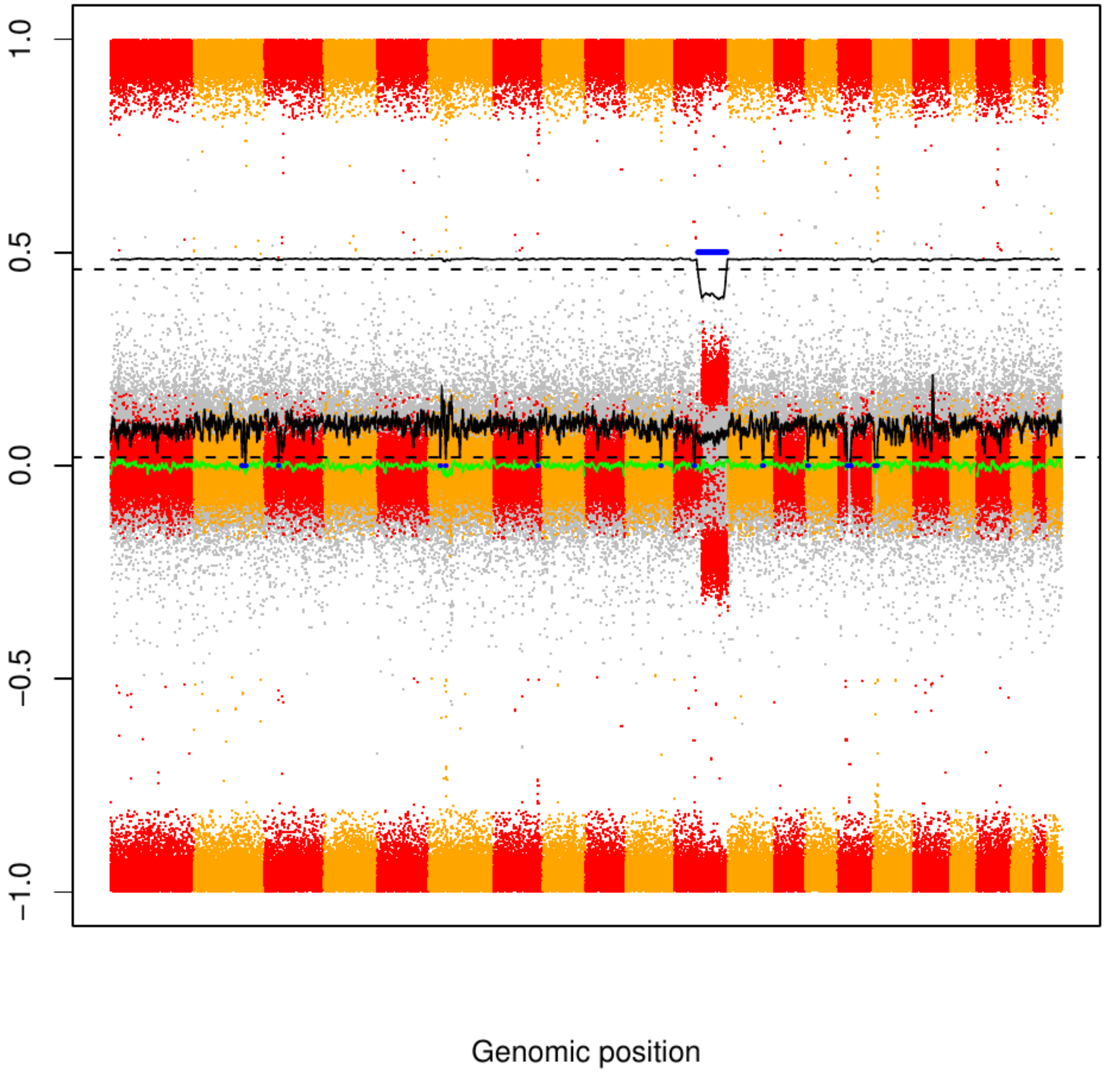

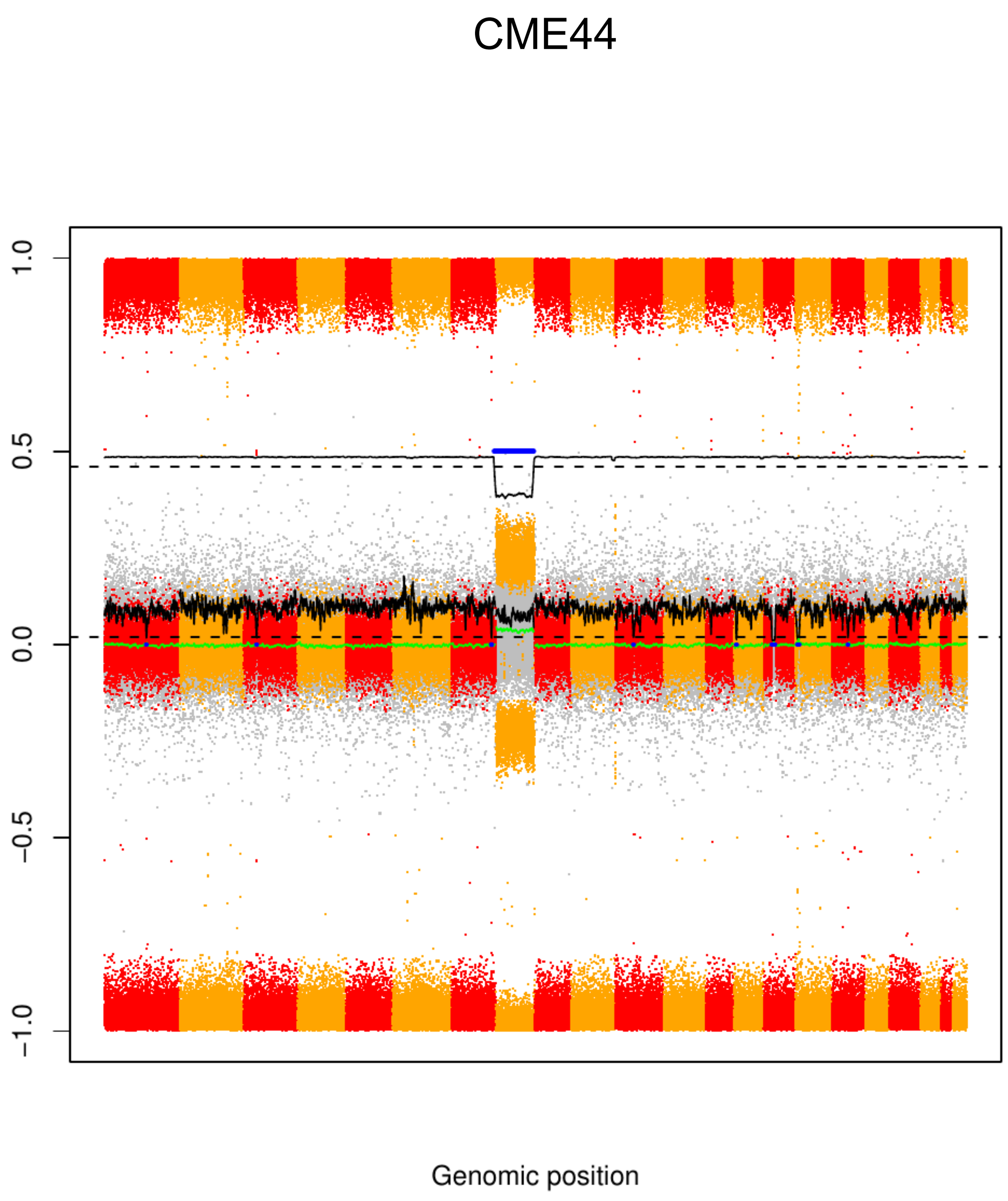
CME44
1.0
0.5
0.0
−0.5
−1.0
Genomic position

CME45

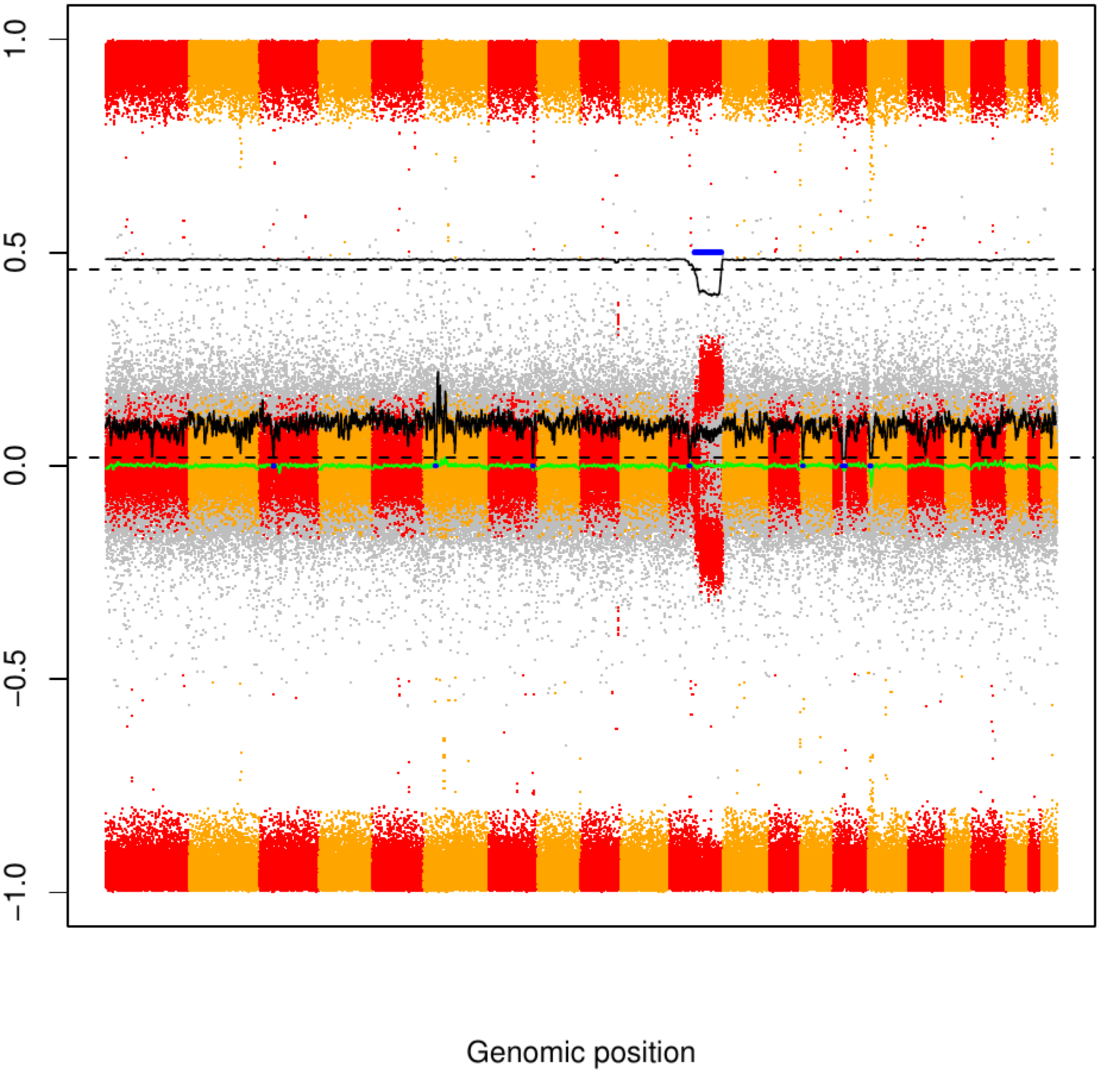

### CME46

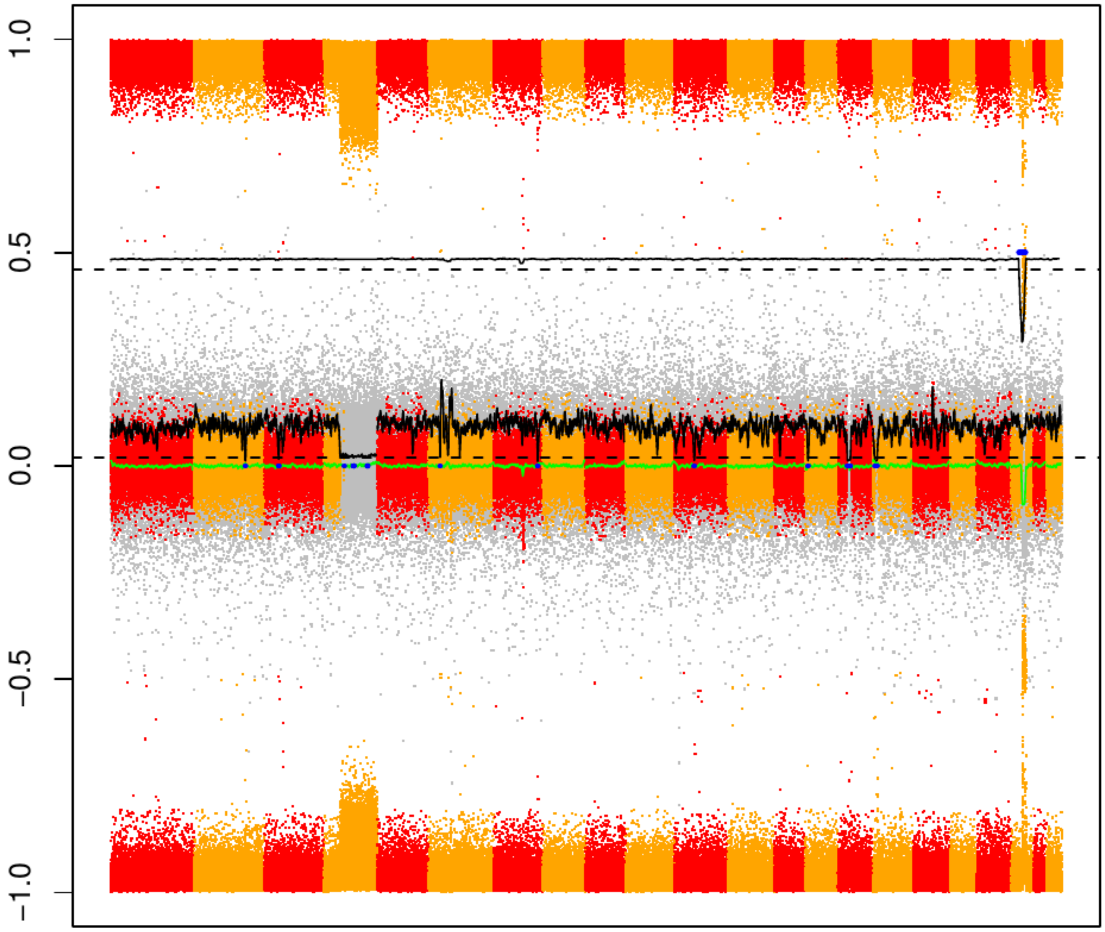

### CME47

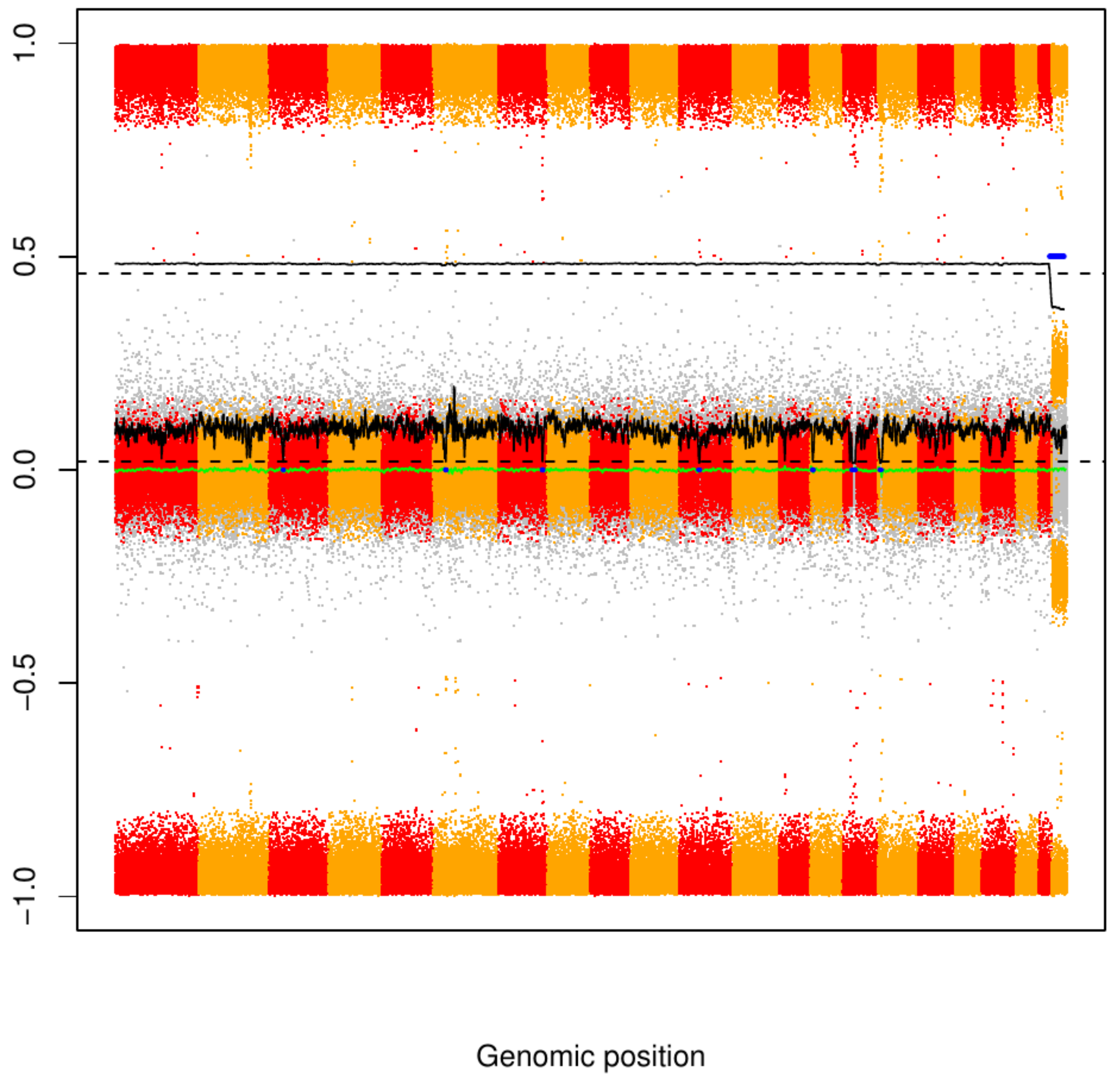

### CME48

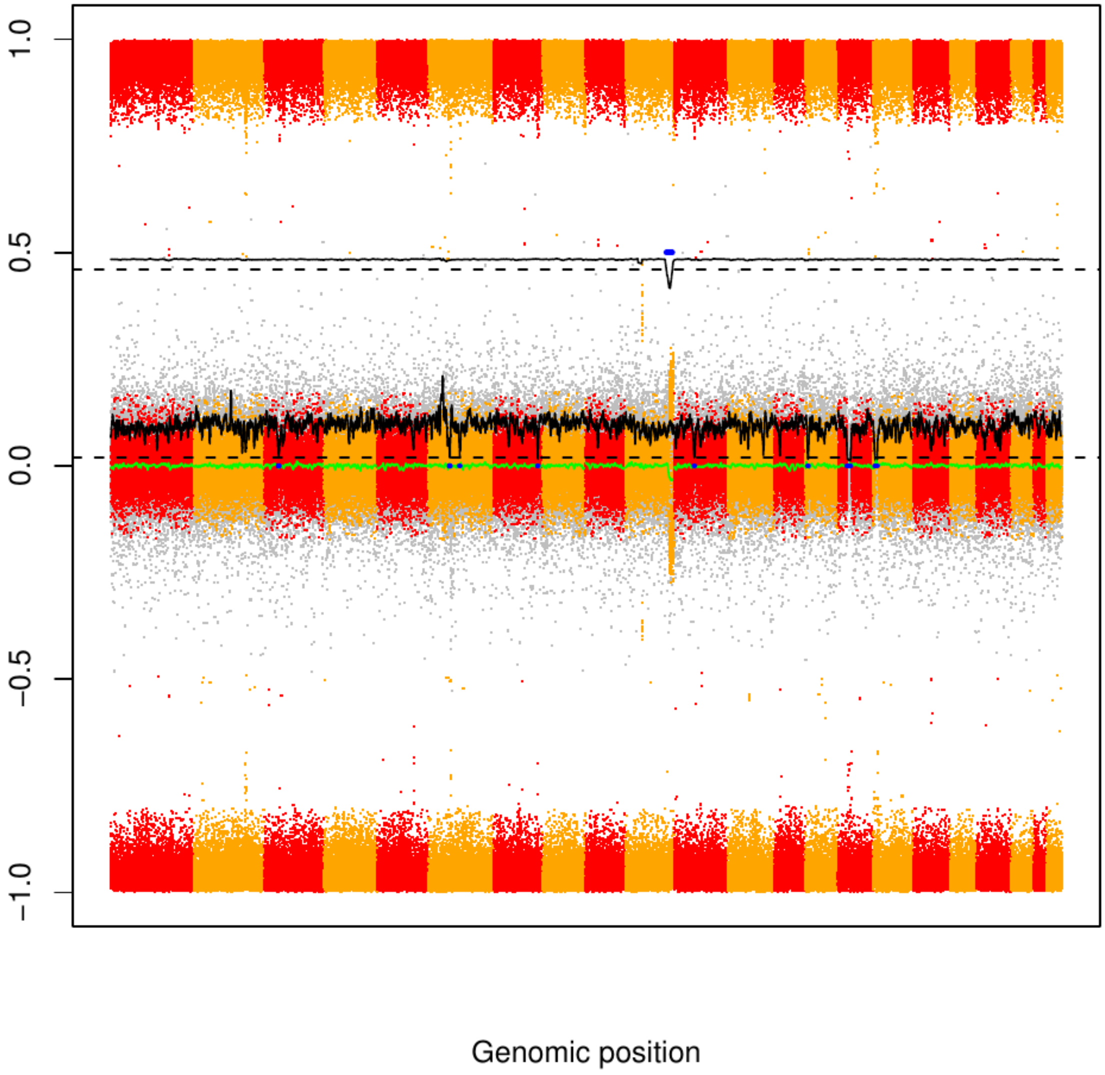

CME49

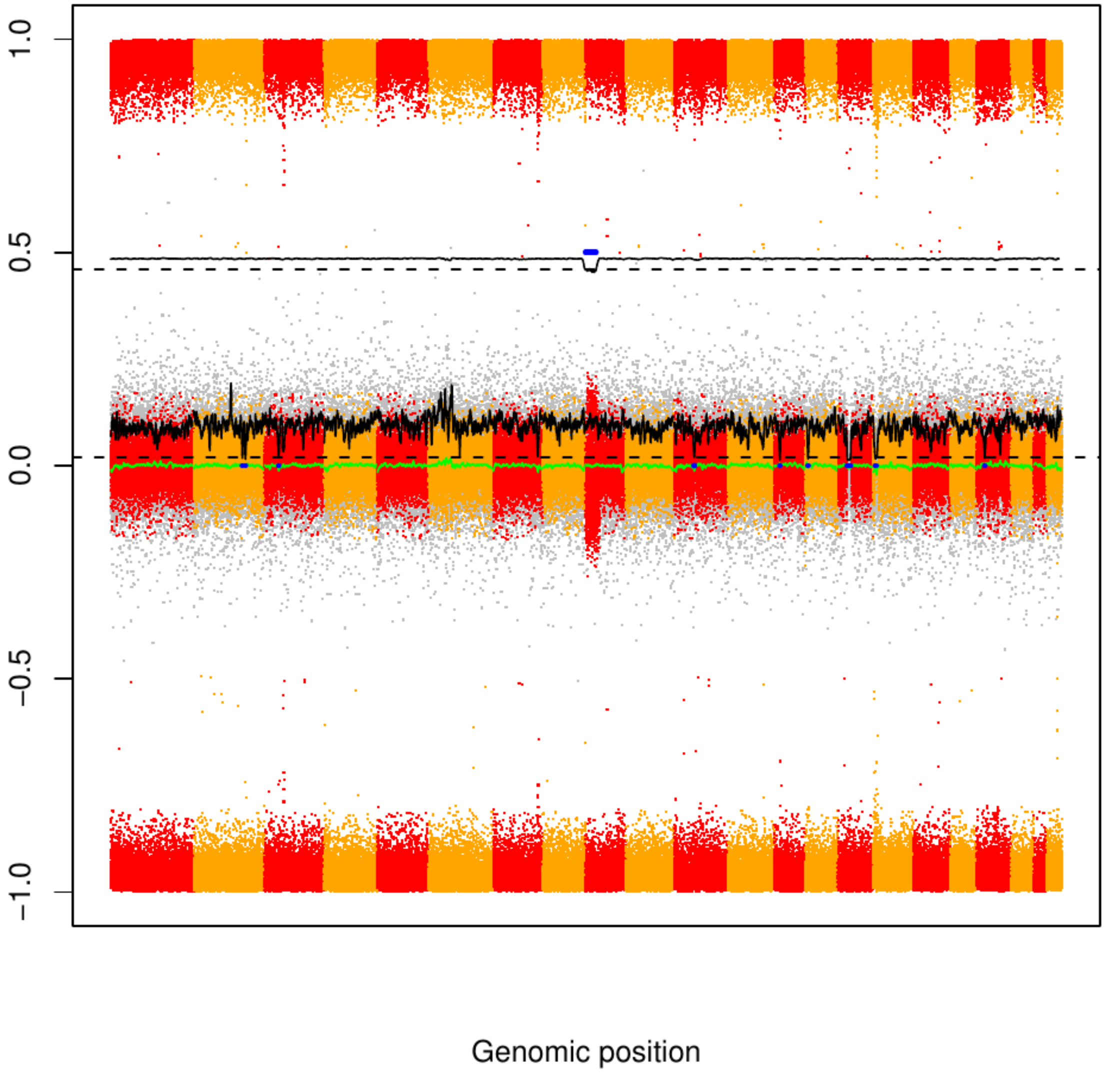

### CME50

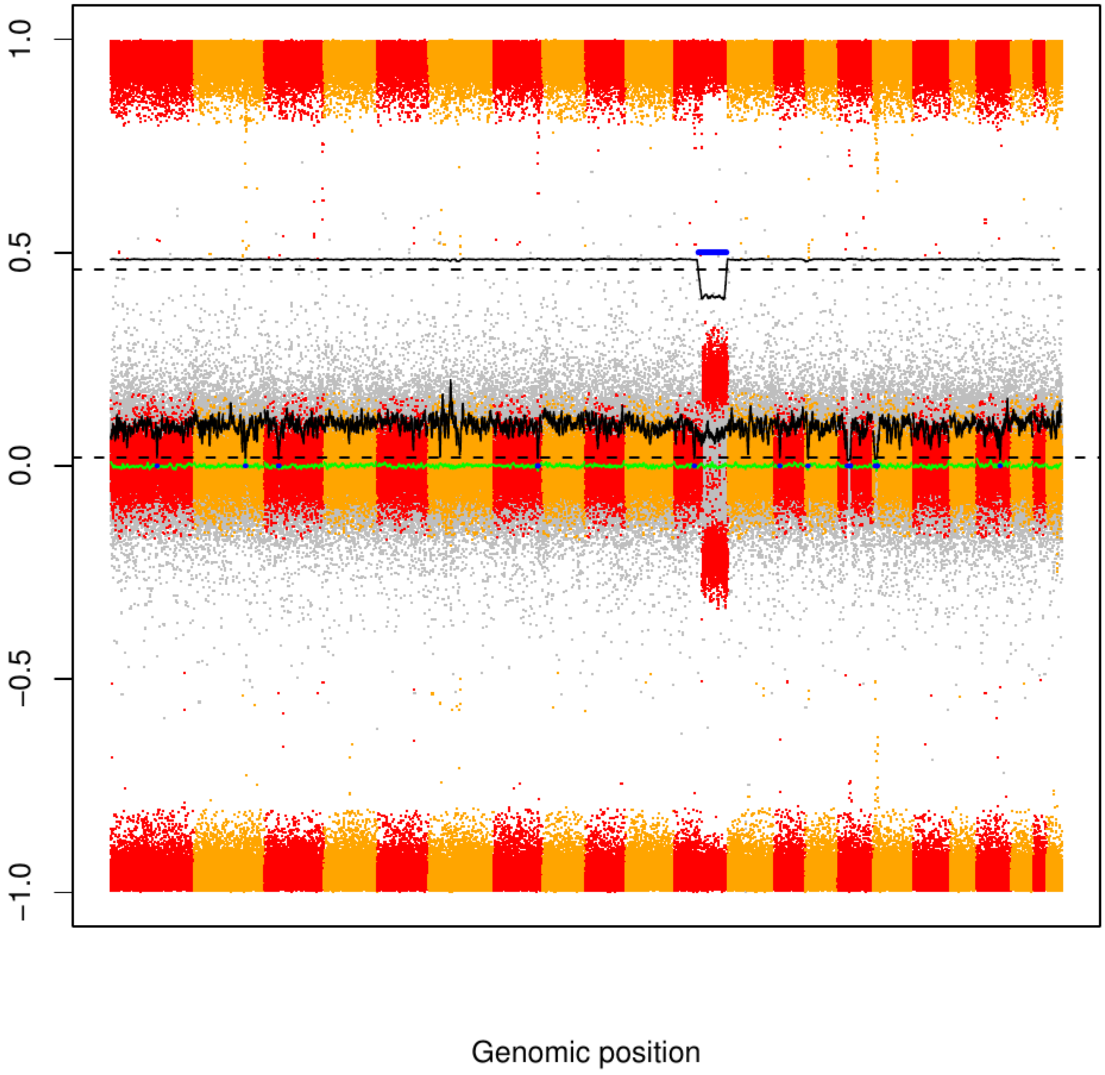

CME51

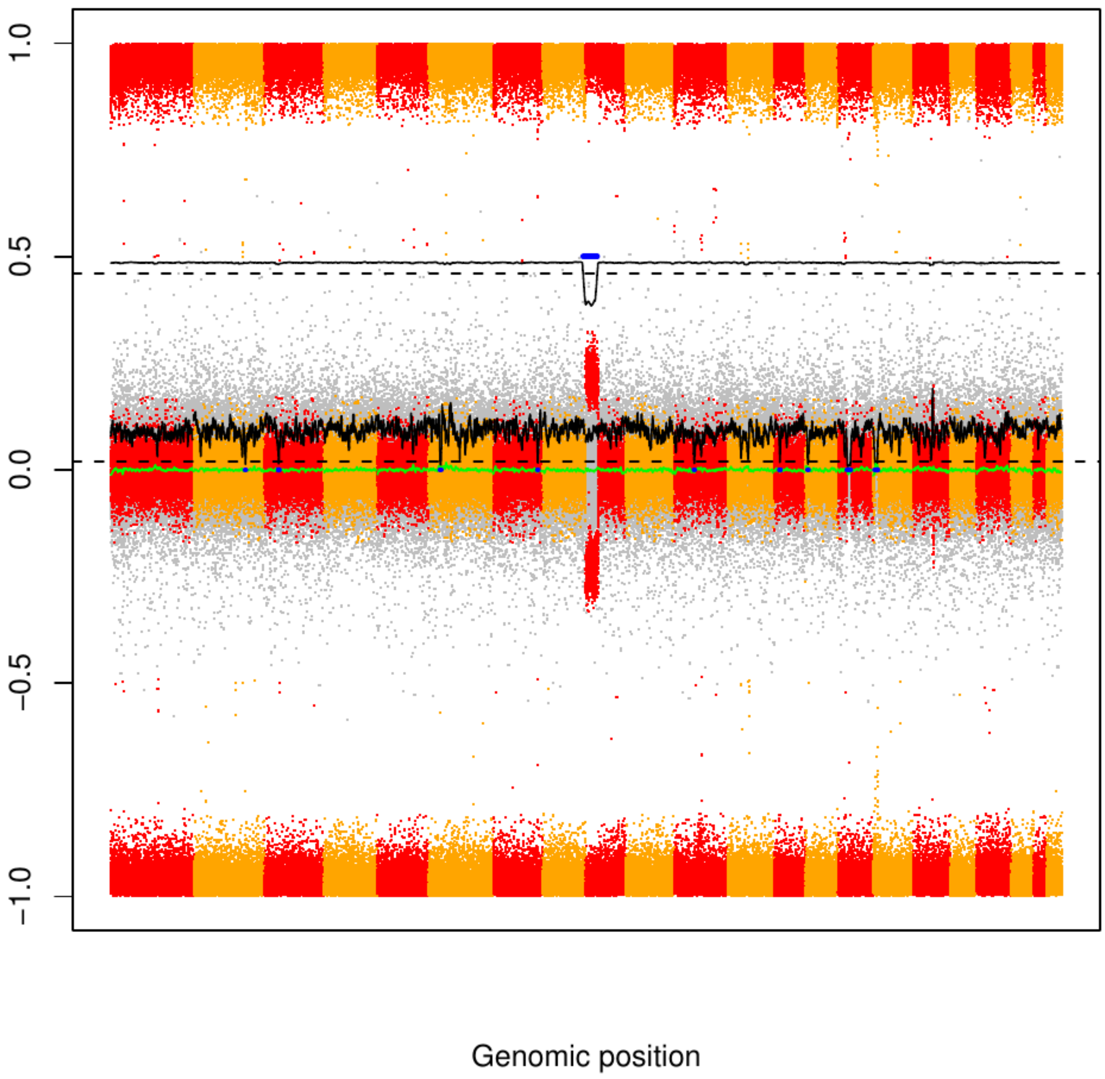

### CME52

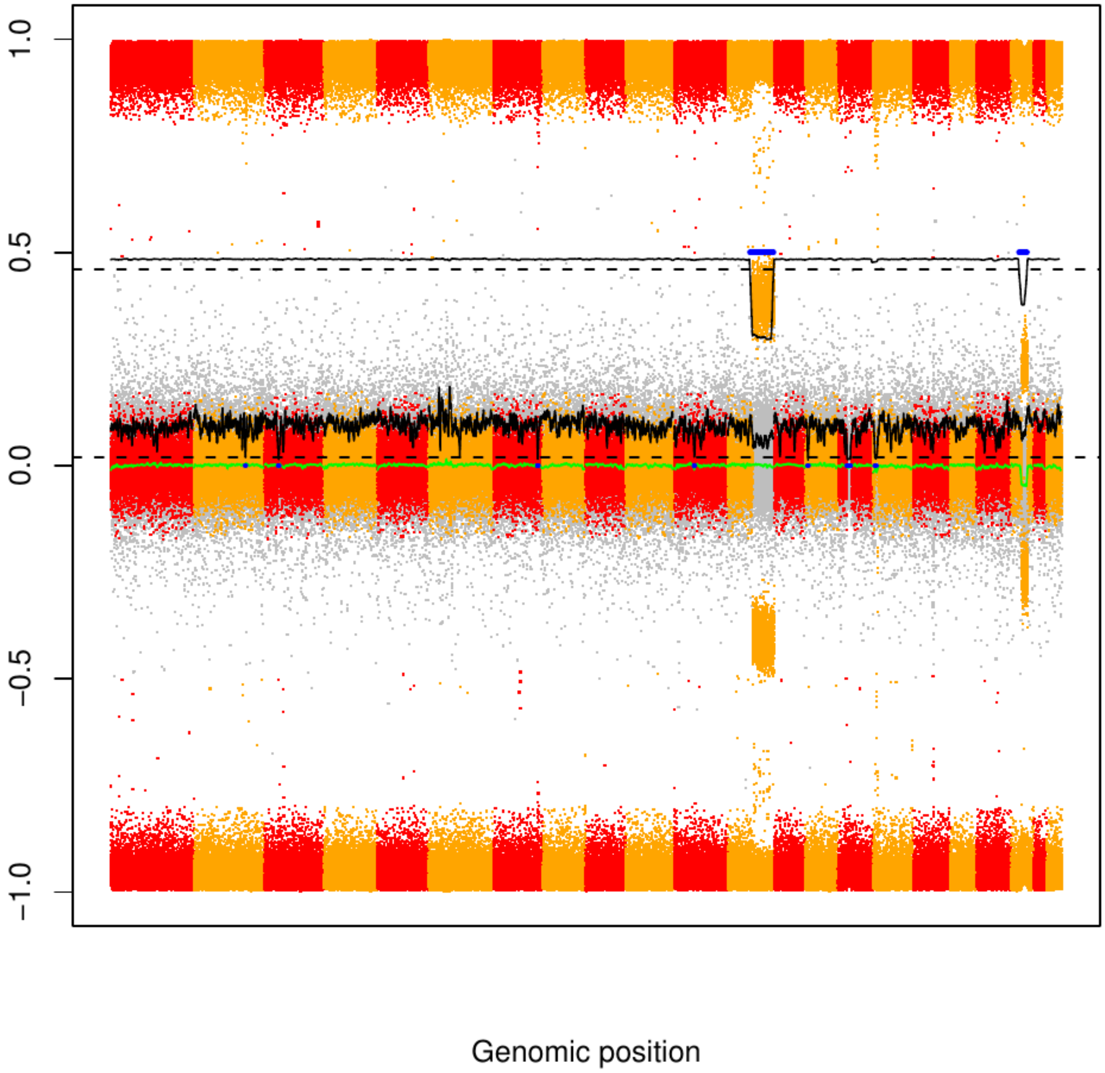

CME53

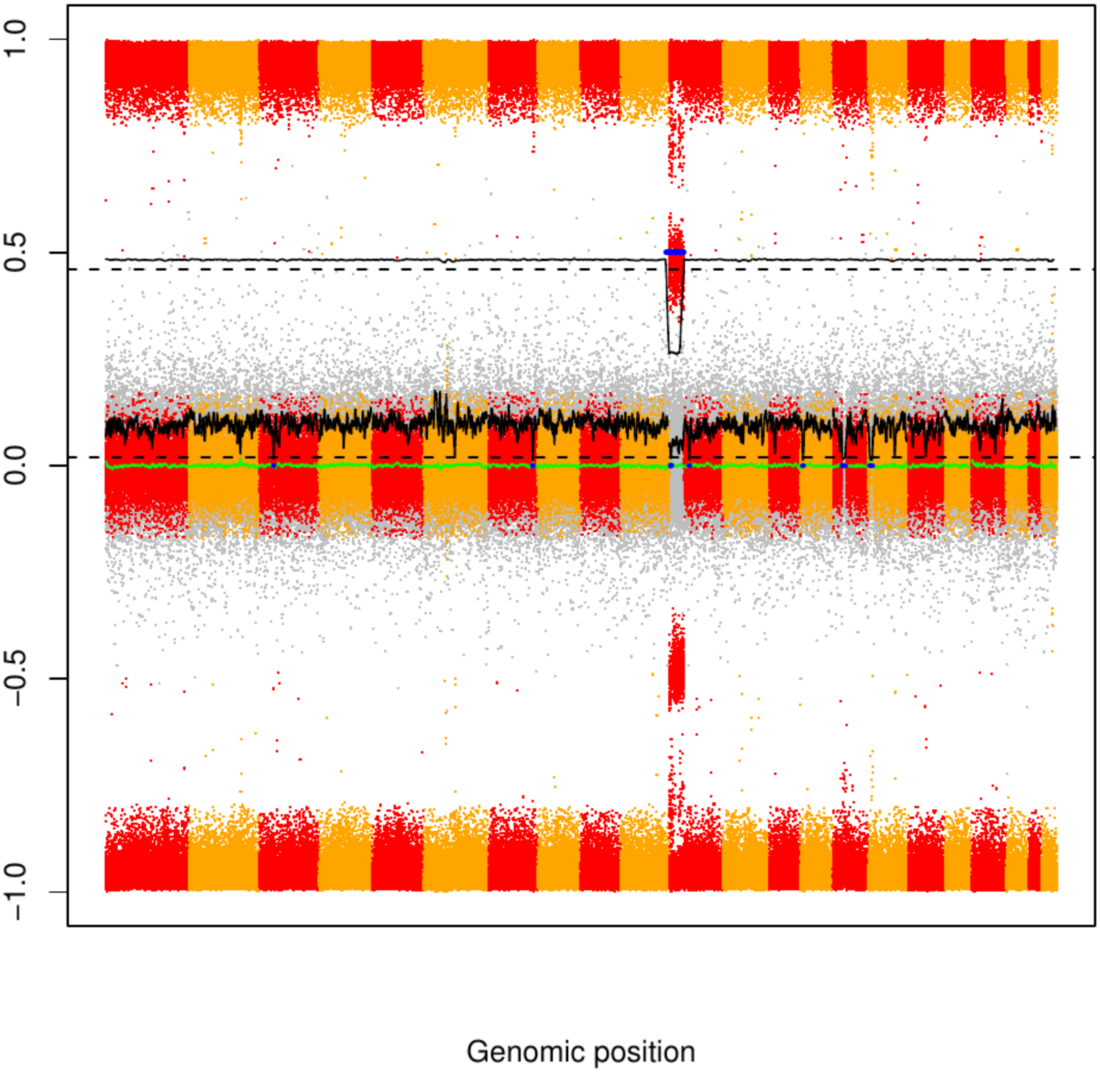

### CME54

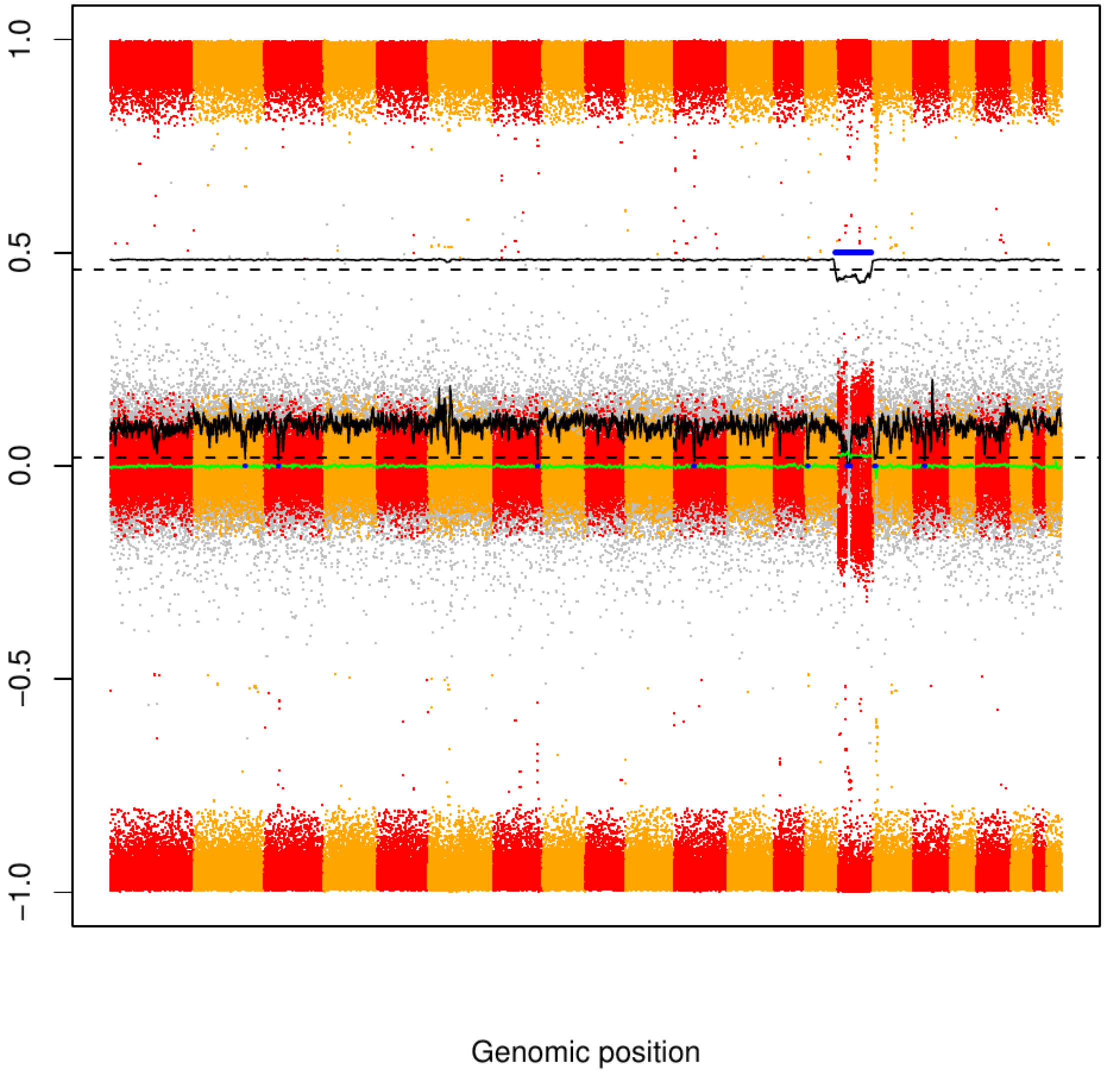

CME55

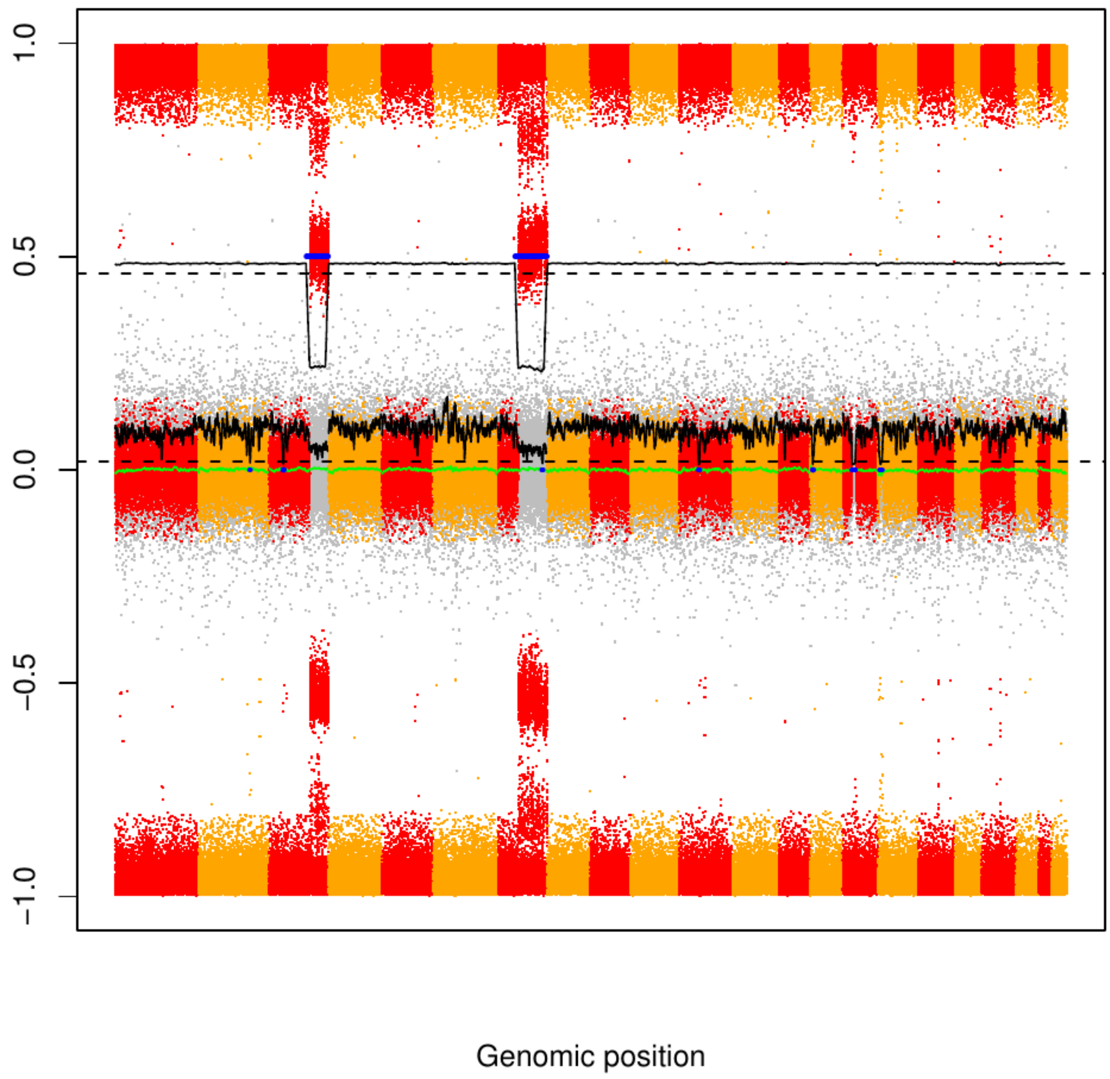

### CME56

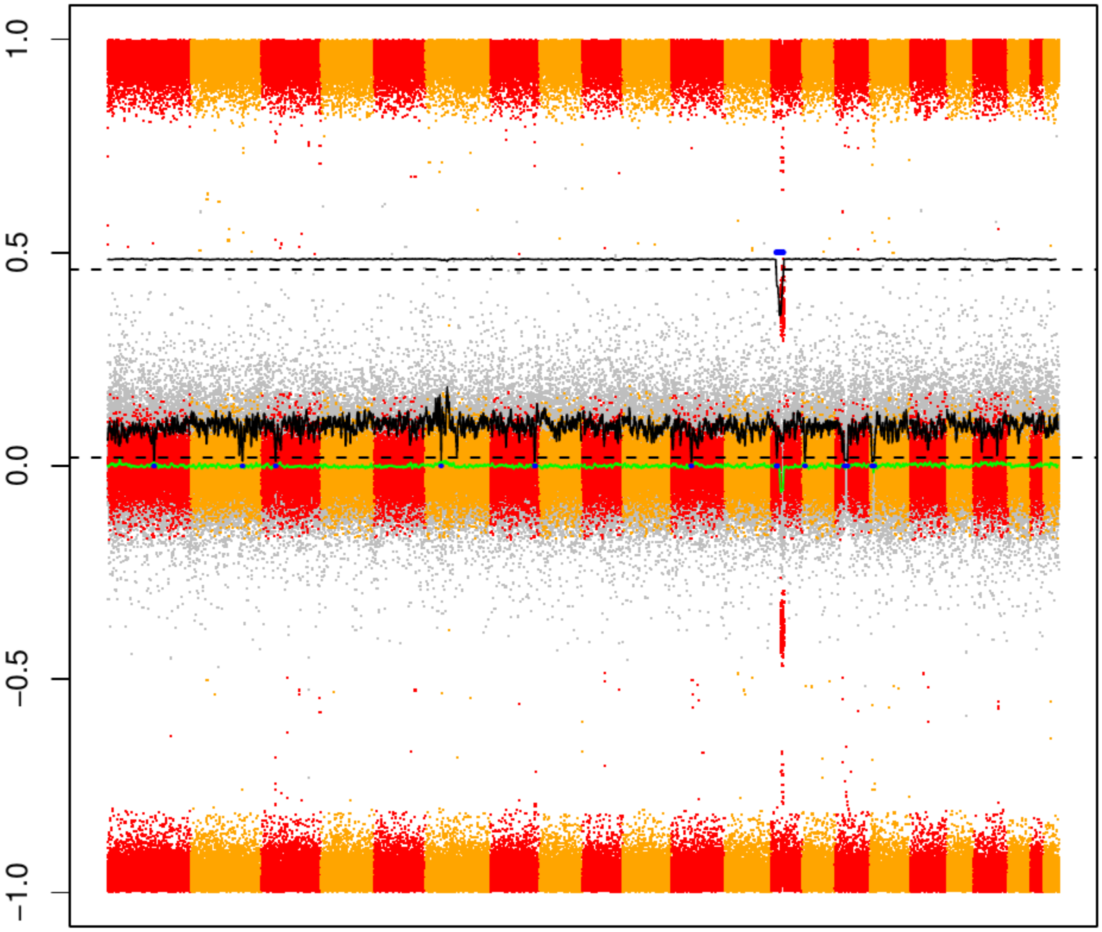


Genomic position

### CME57

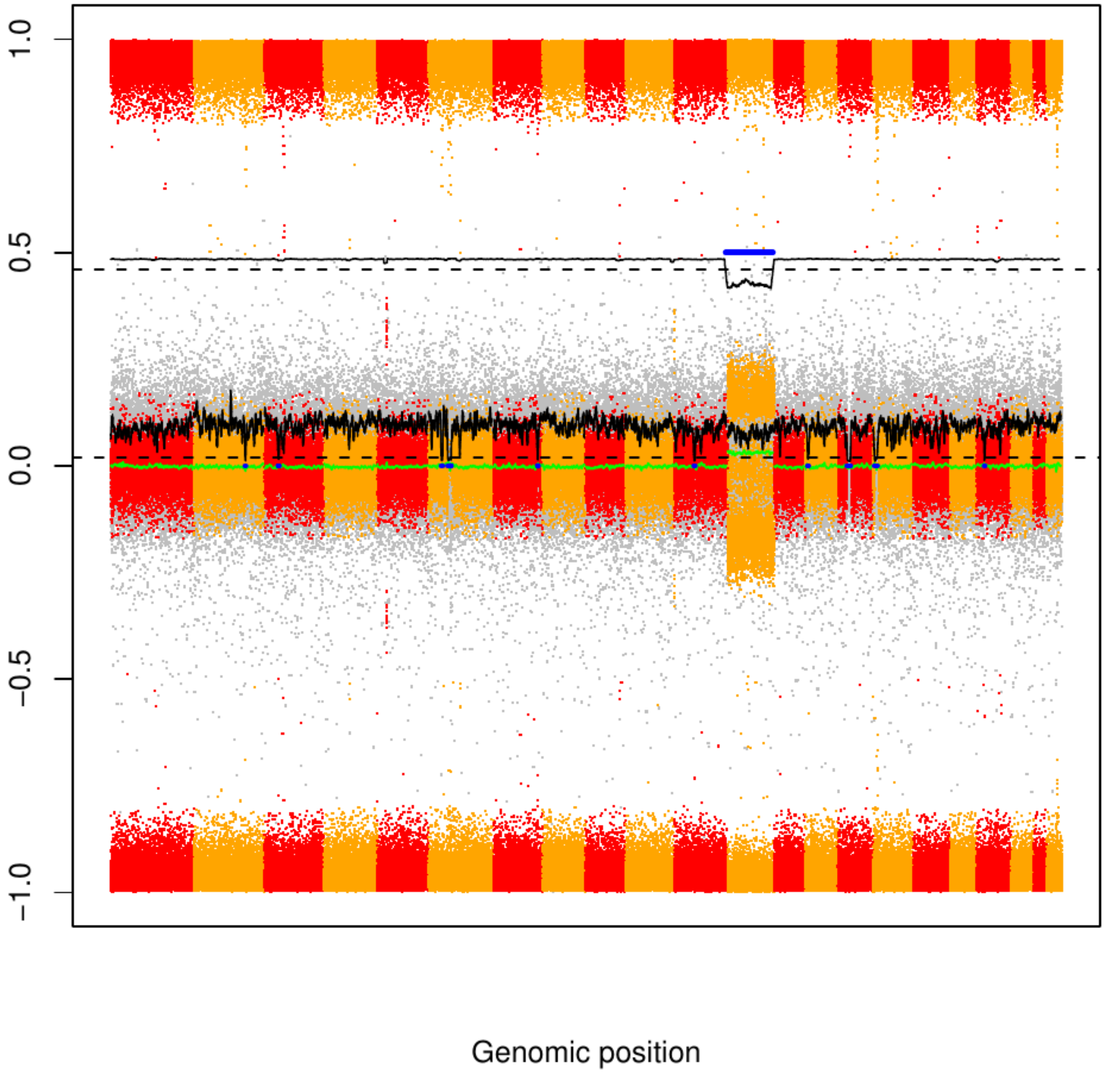

CME58

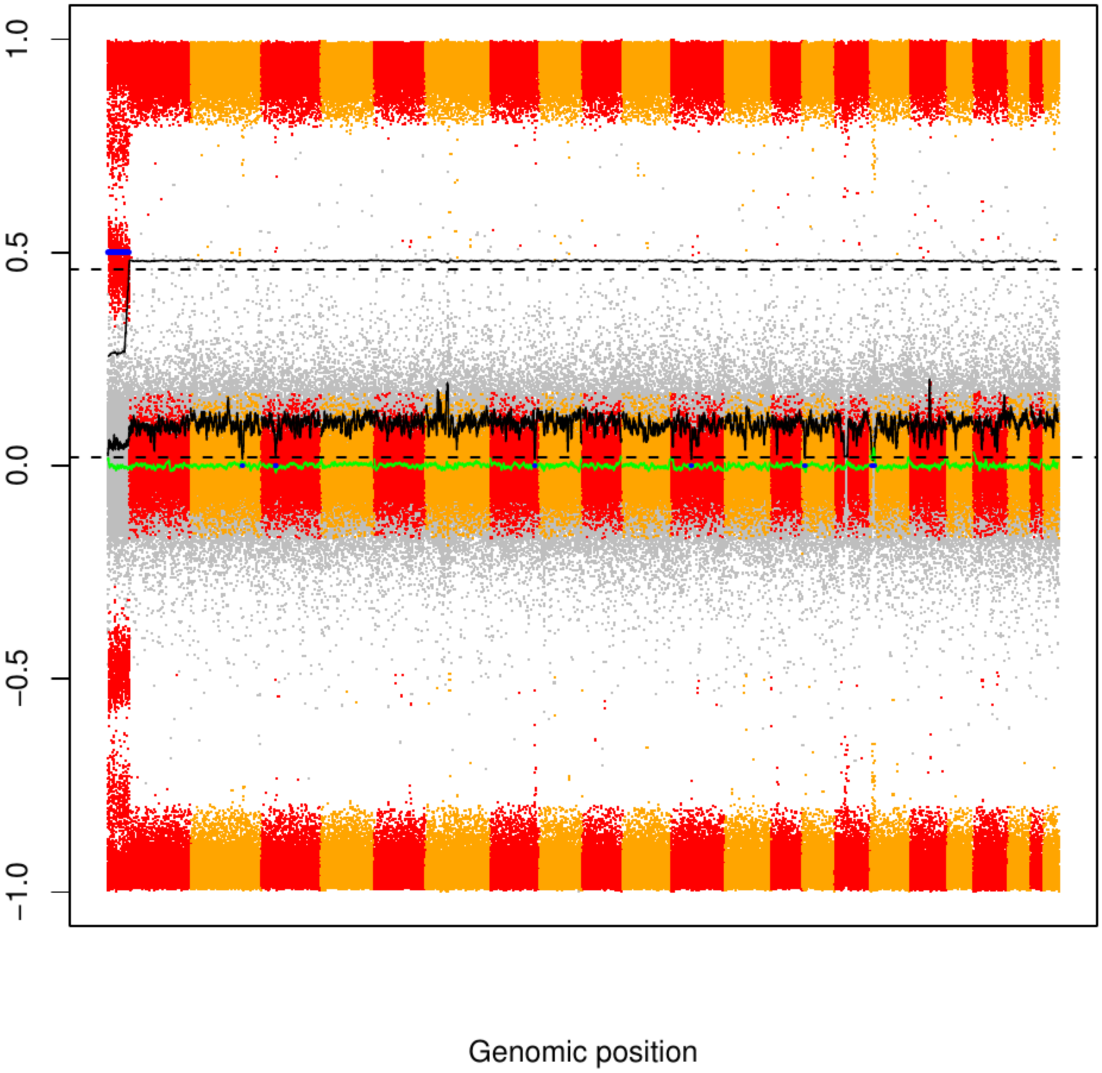

### CME59

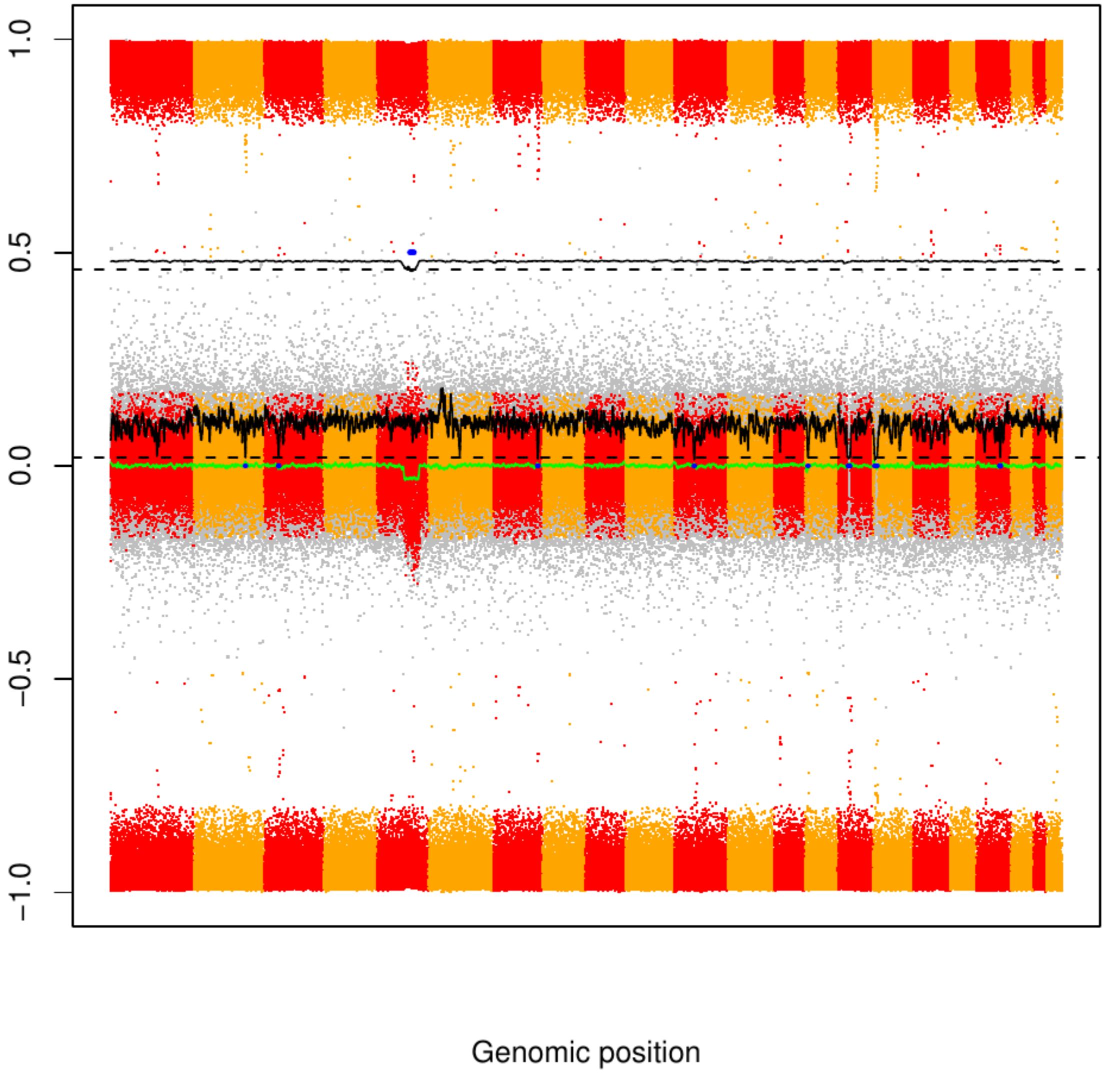

### CME60

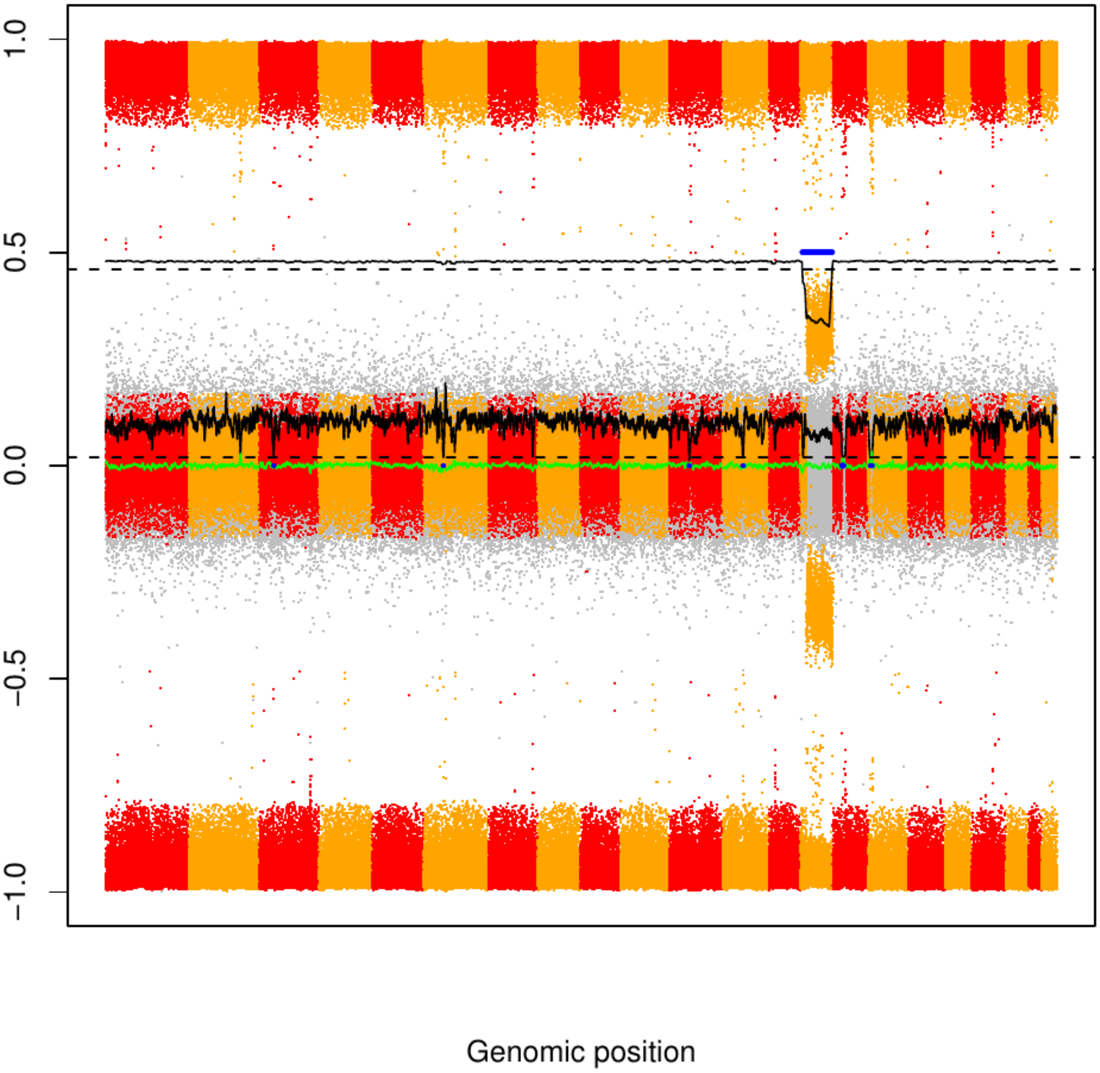

### CME61

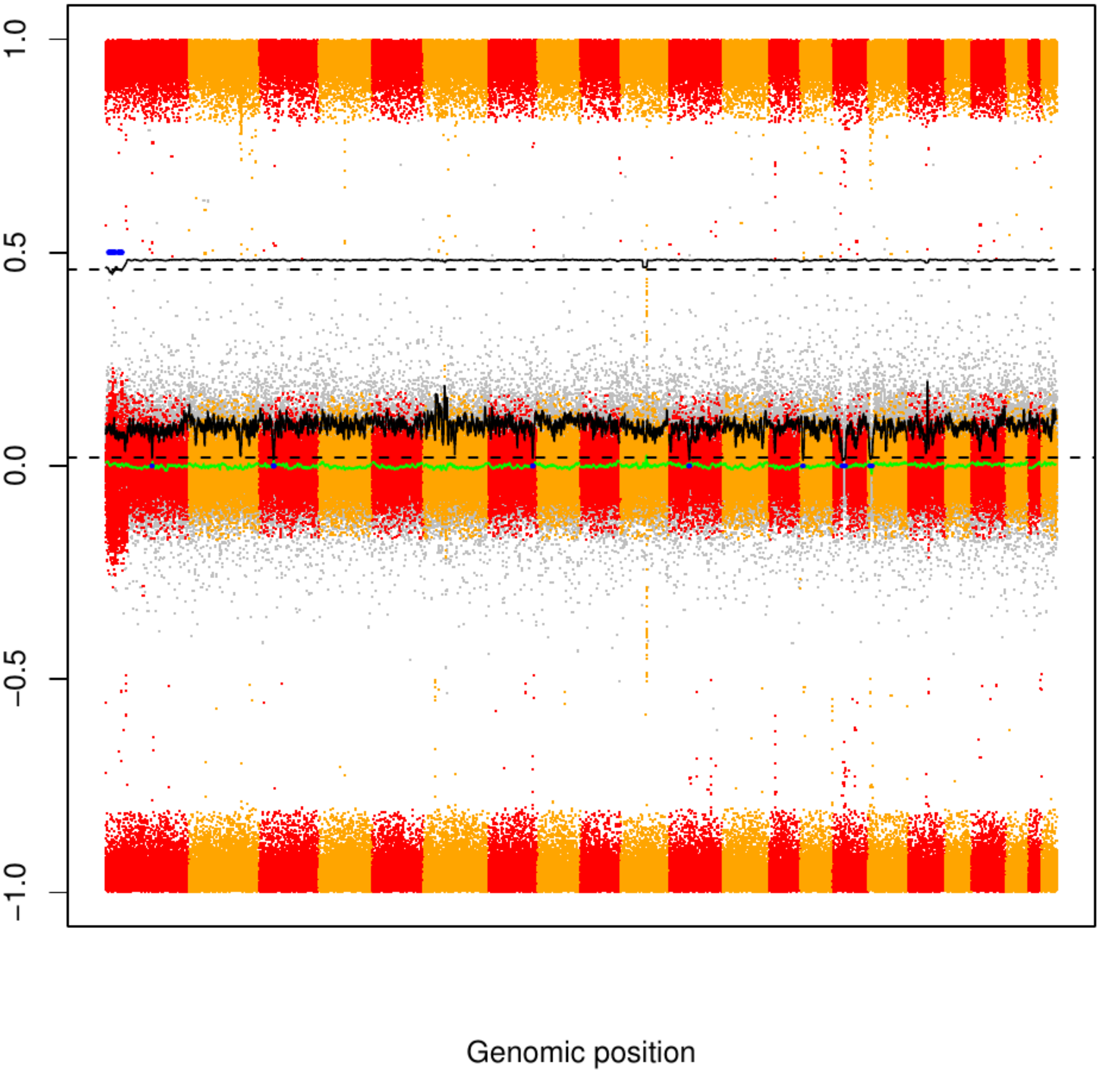

### CME62

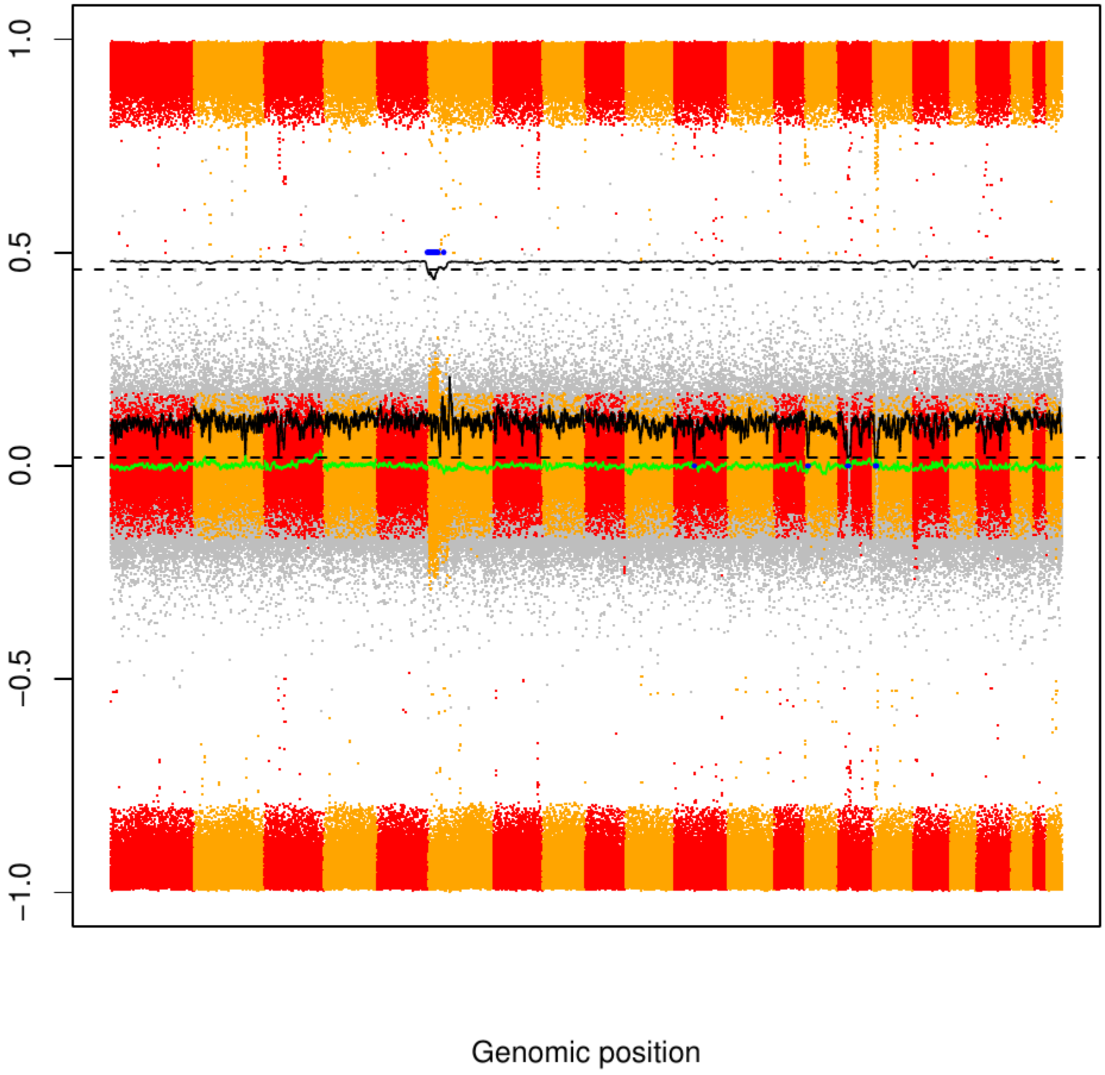

CME63

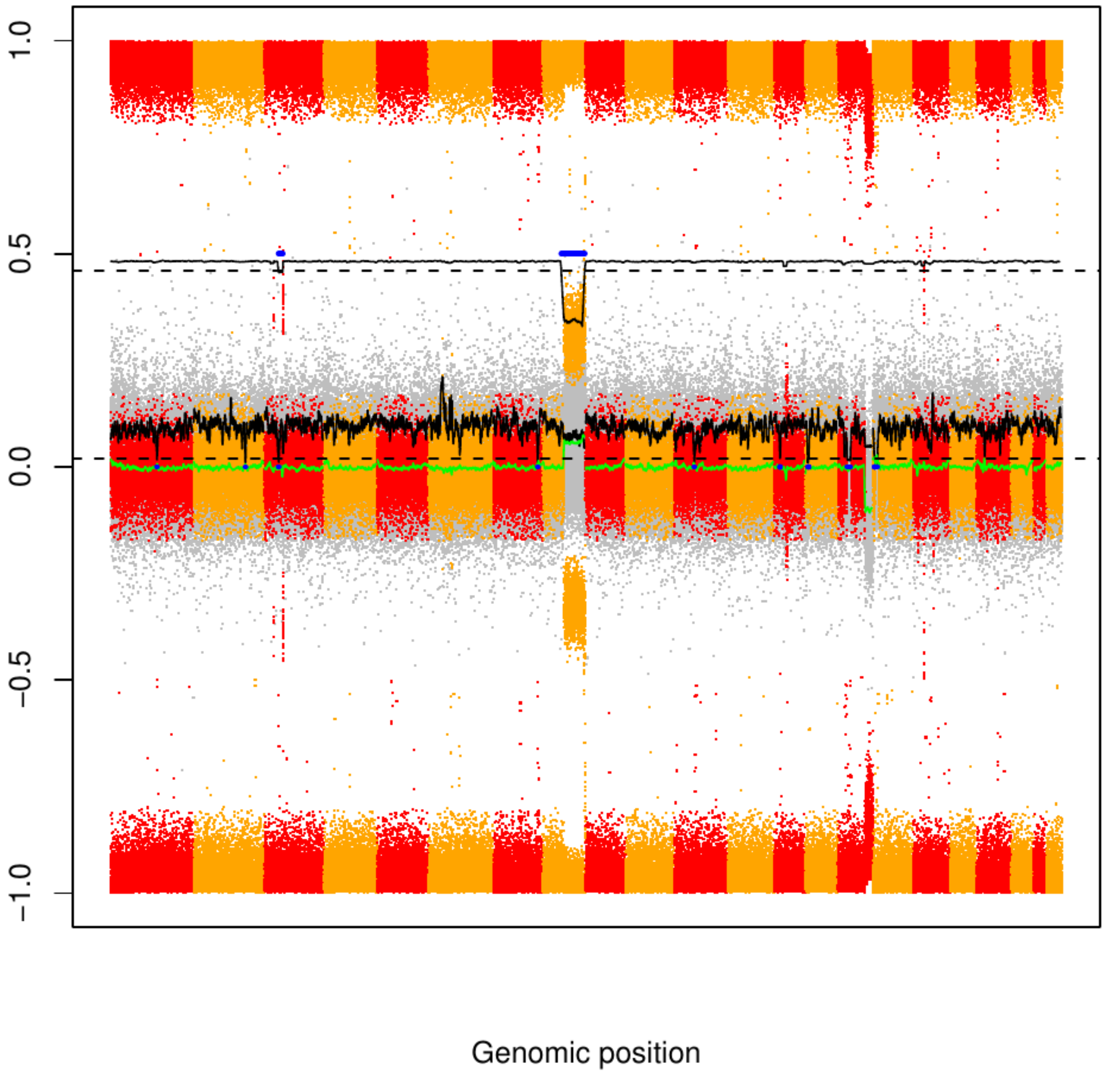

### CME64

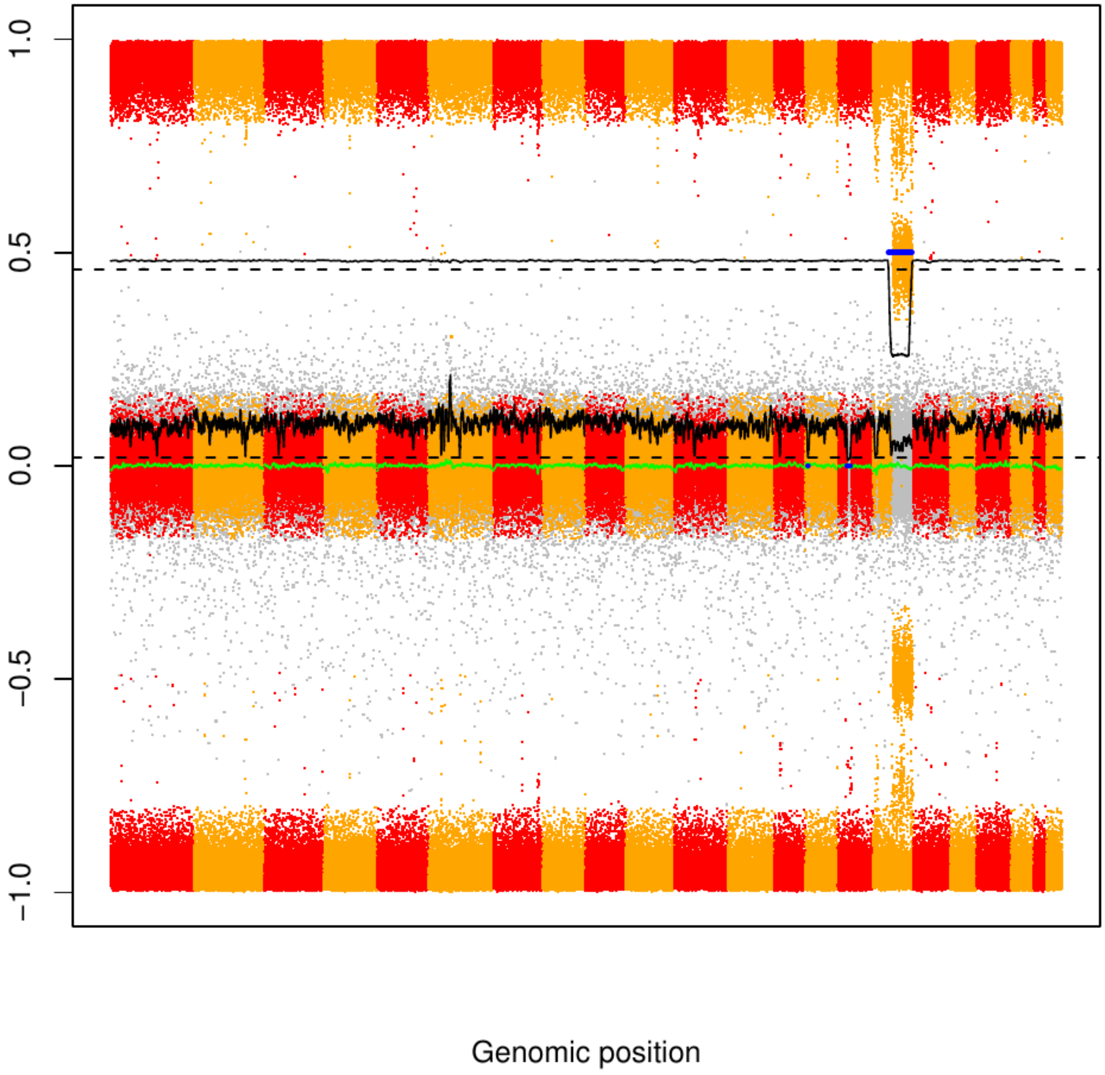

CME65

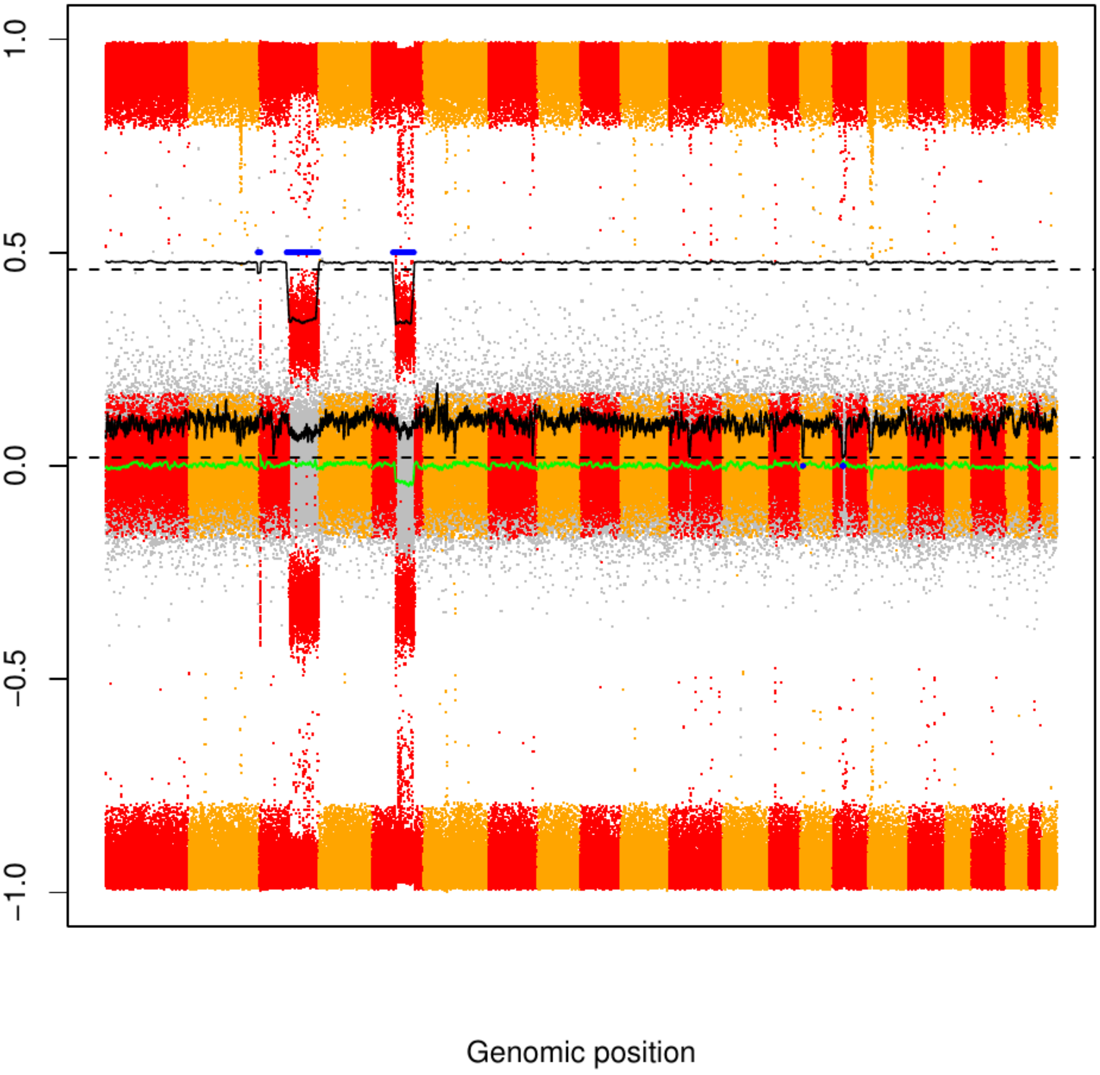

### CME66

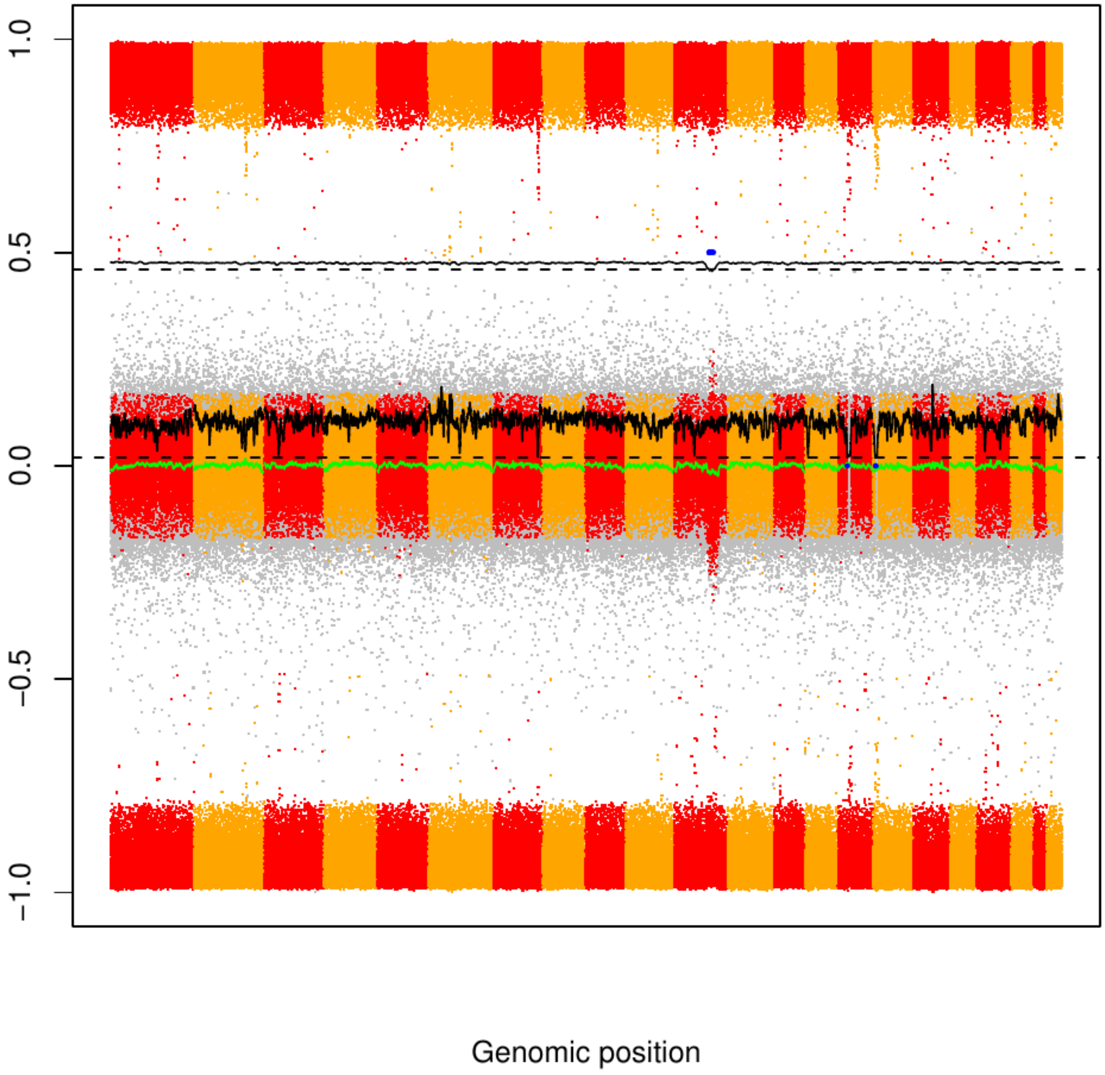

### CME67

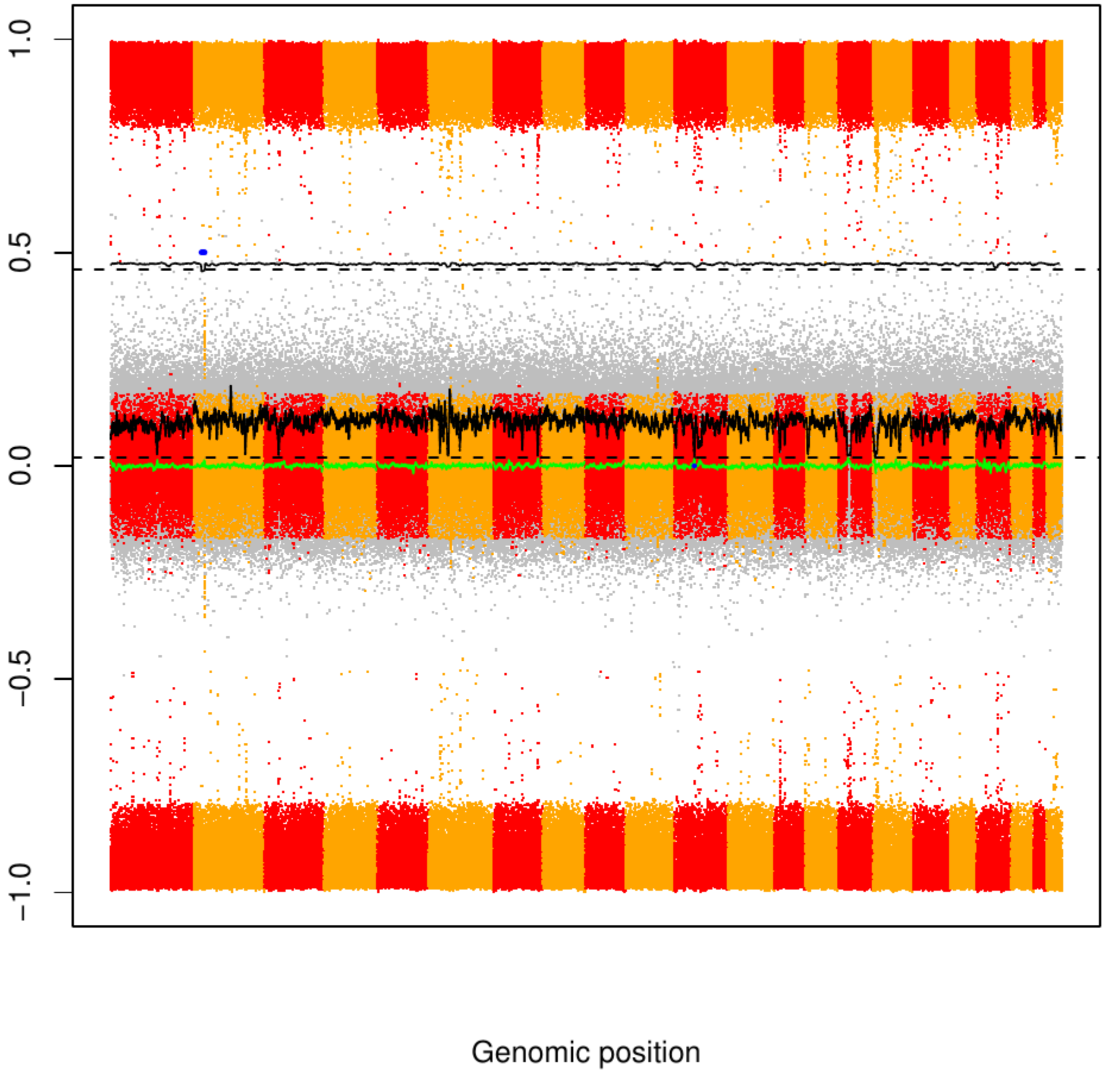

### CME68

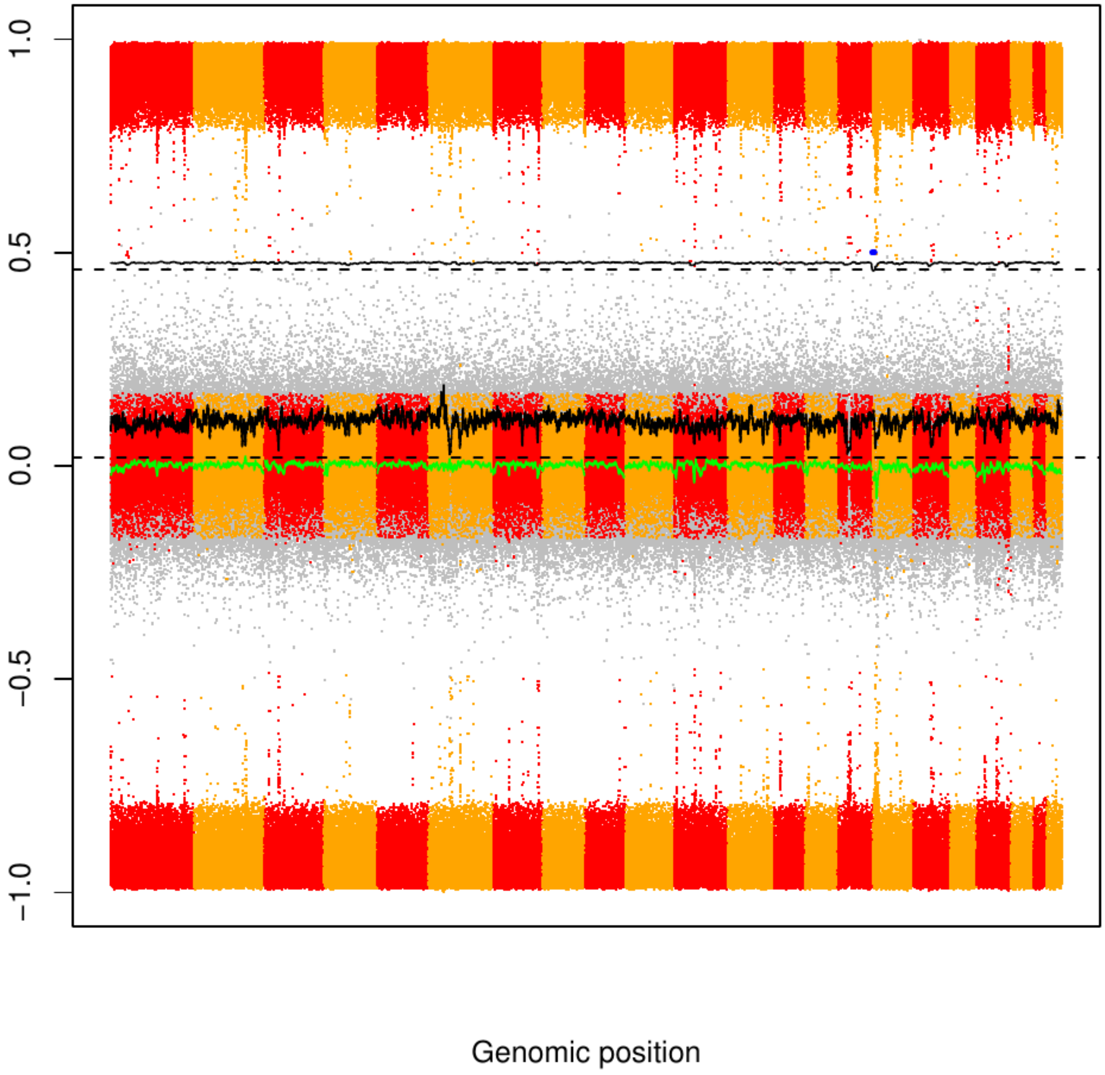

### CME69

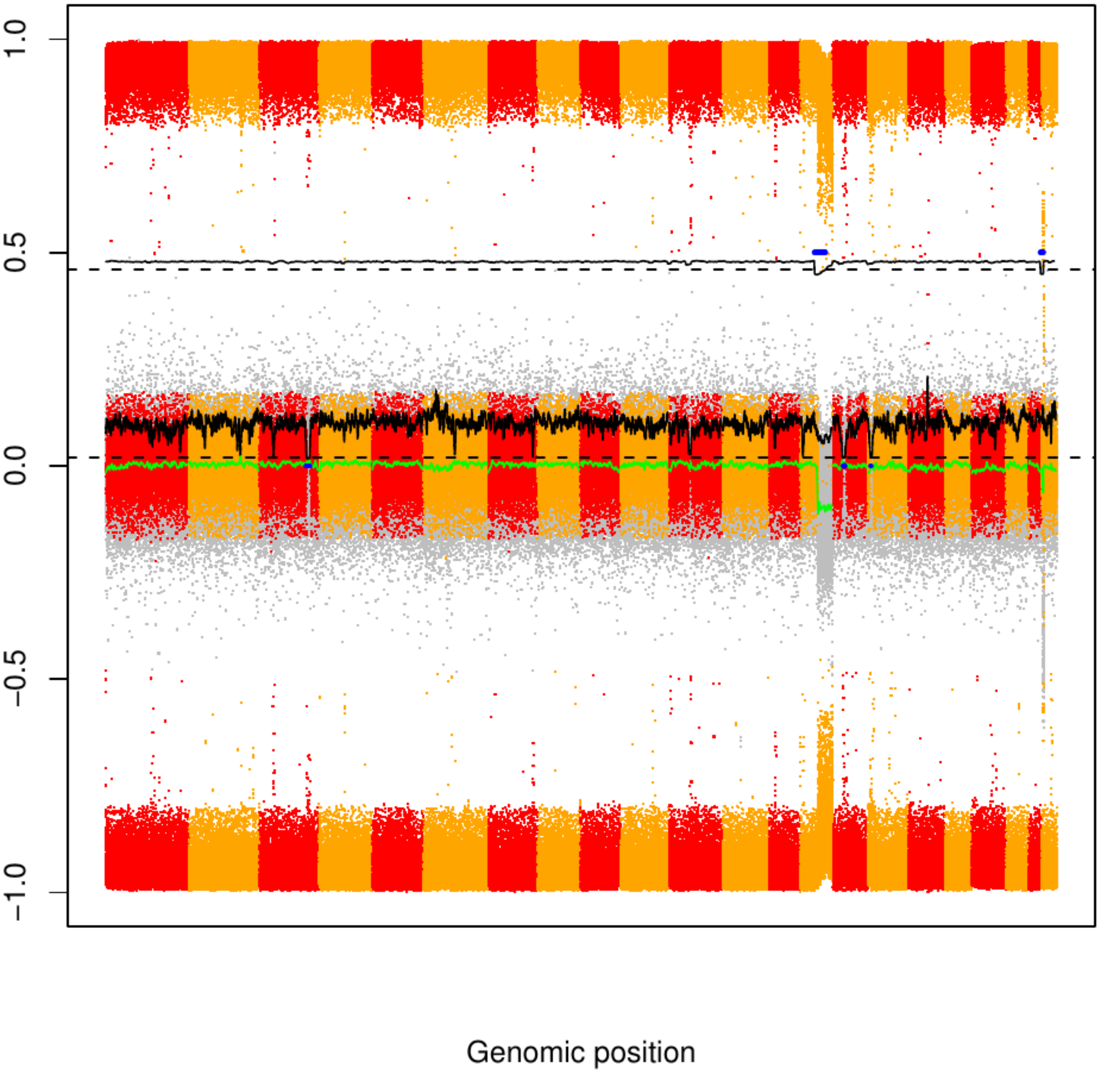

### CME70

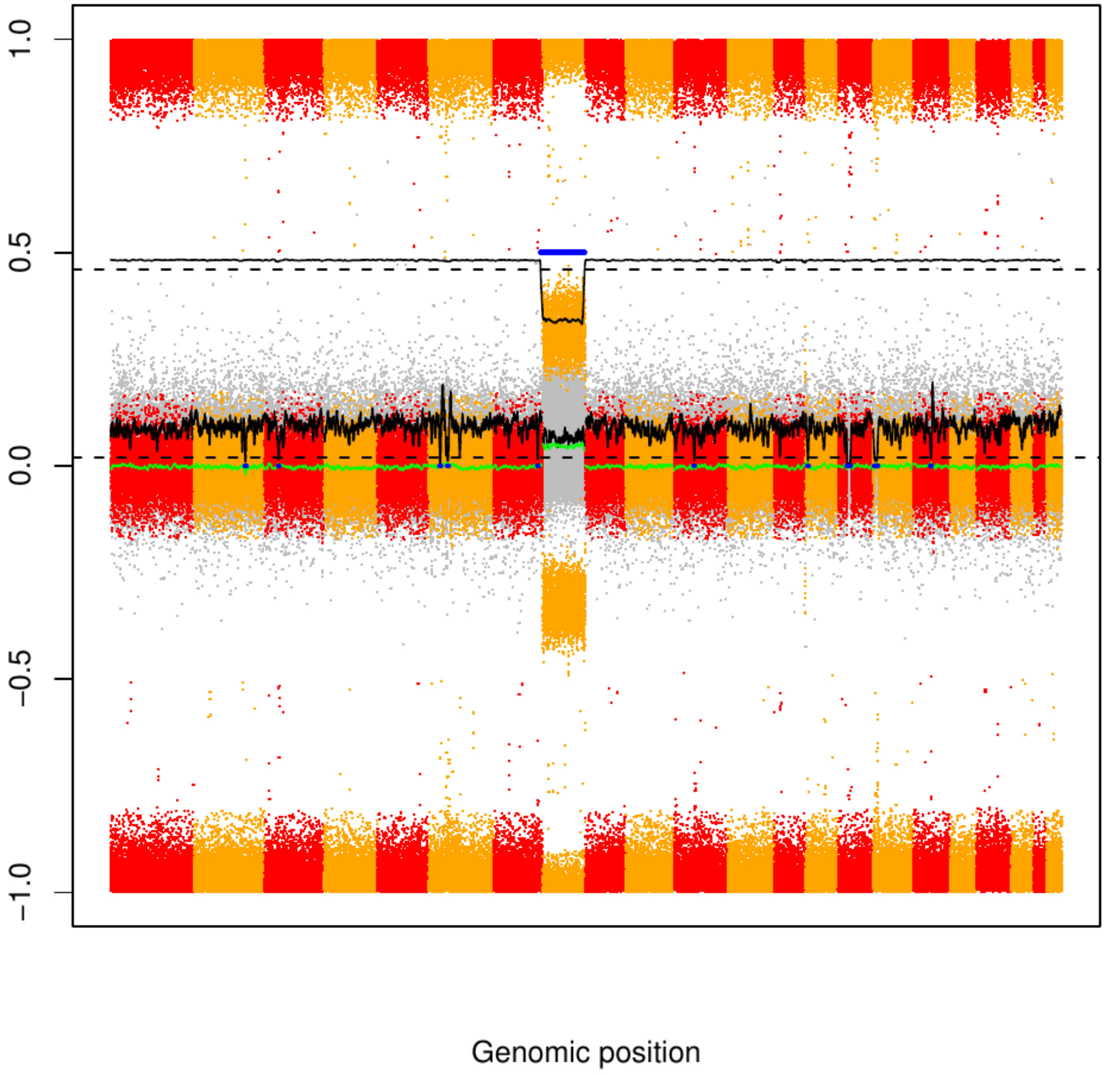

CME71

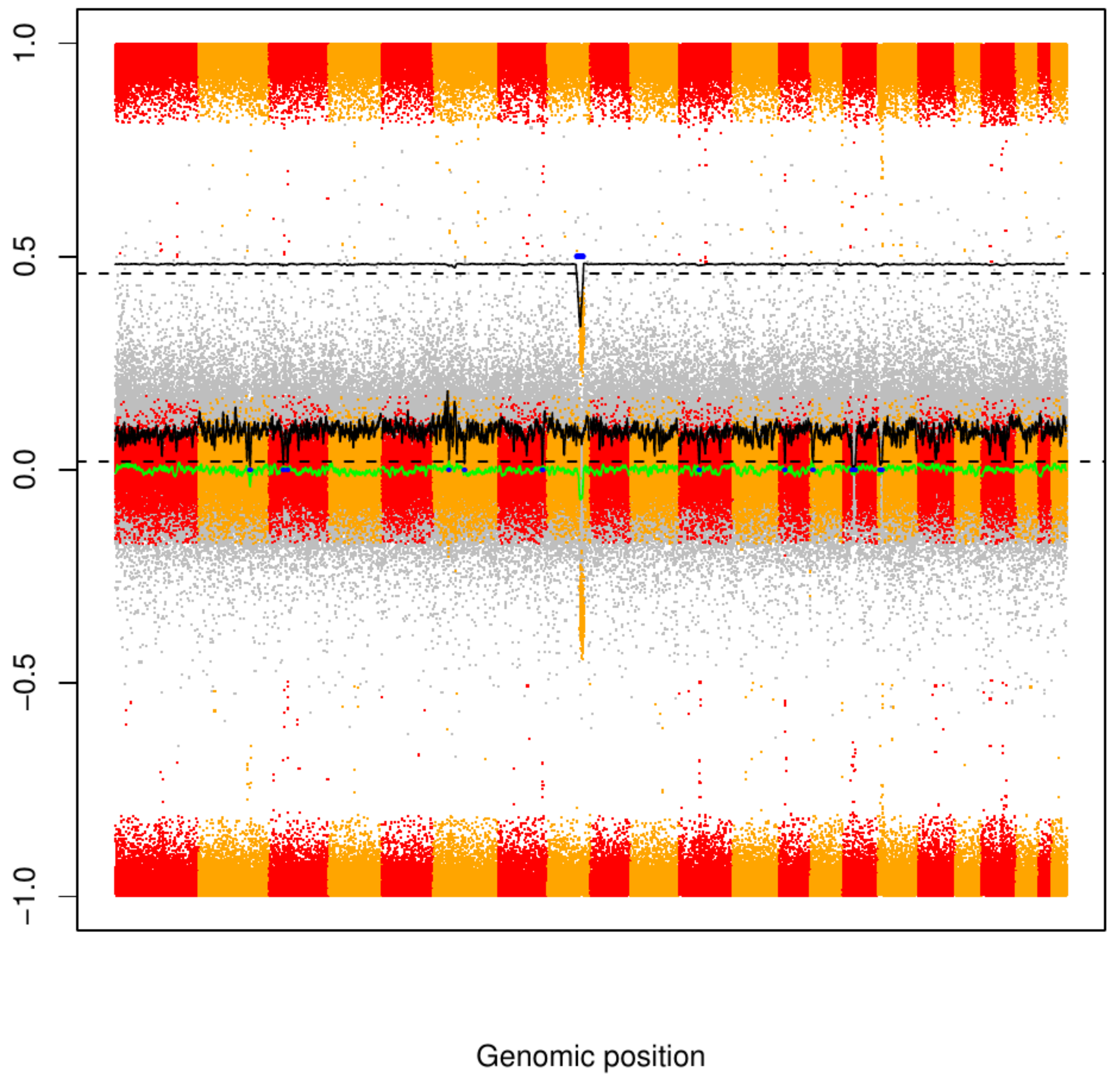

CME72

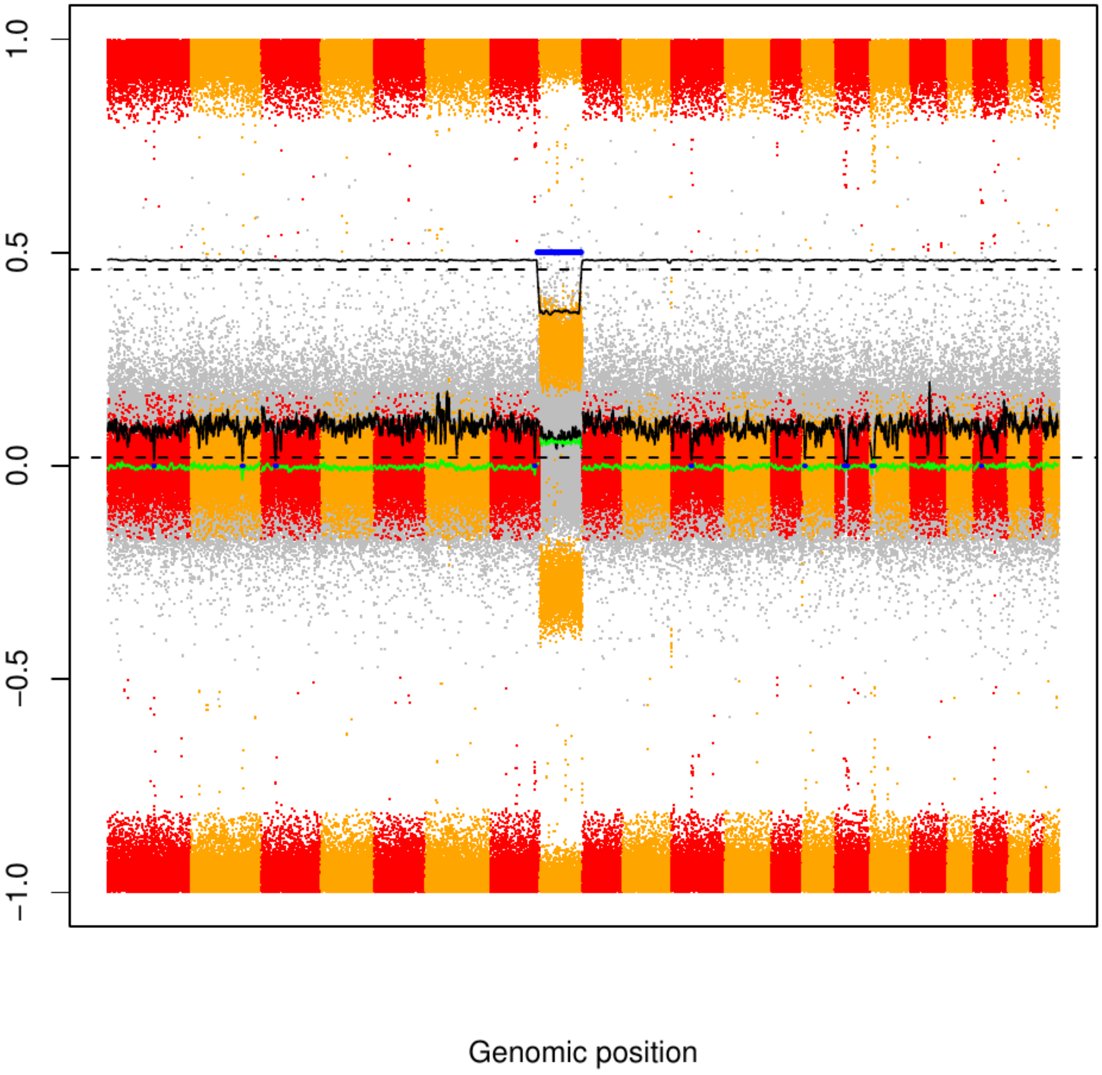

### CME73

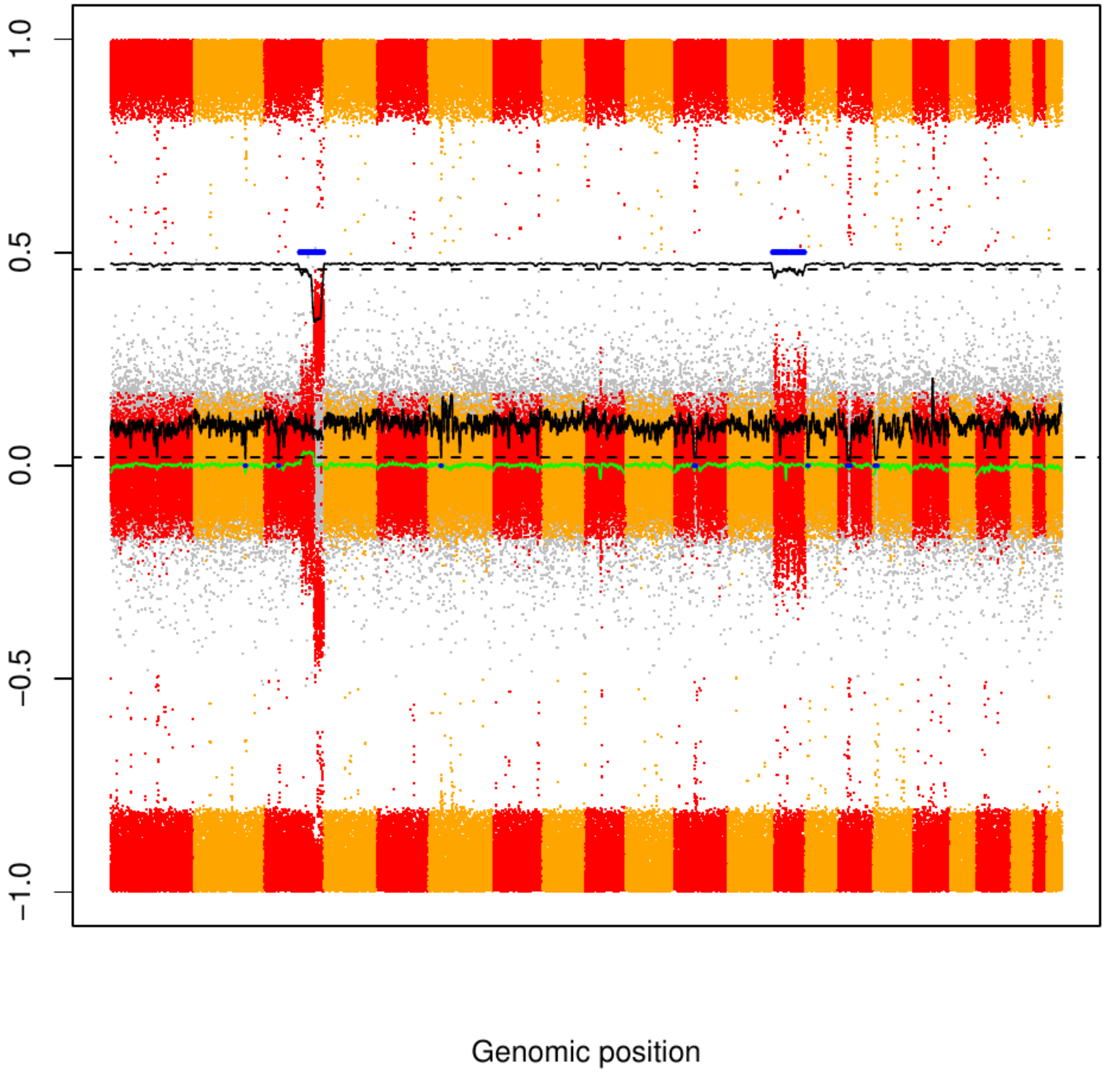

### CME74

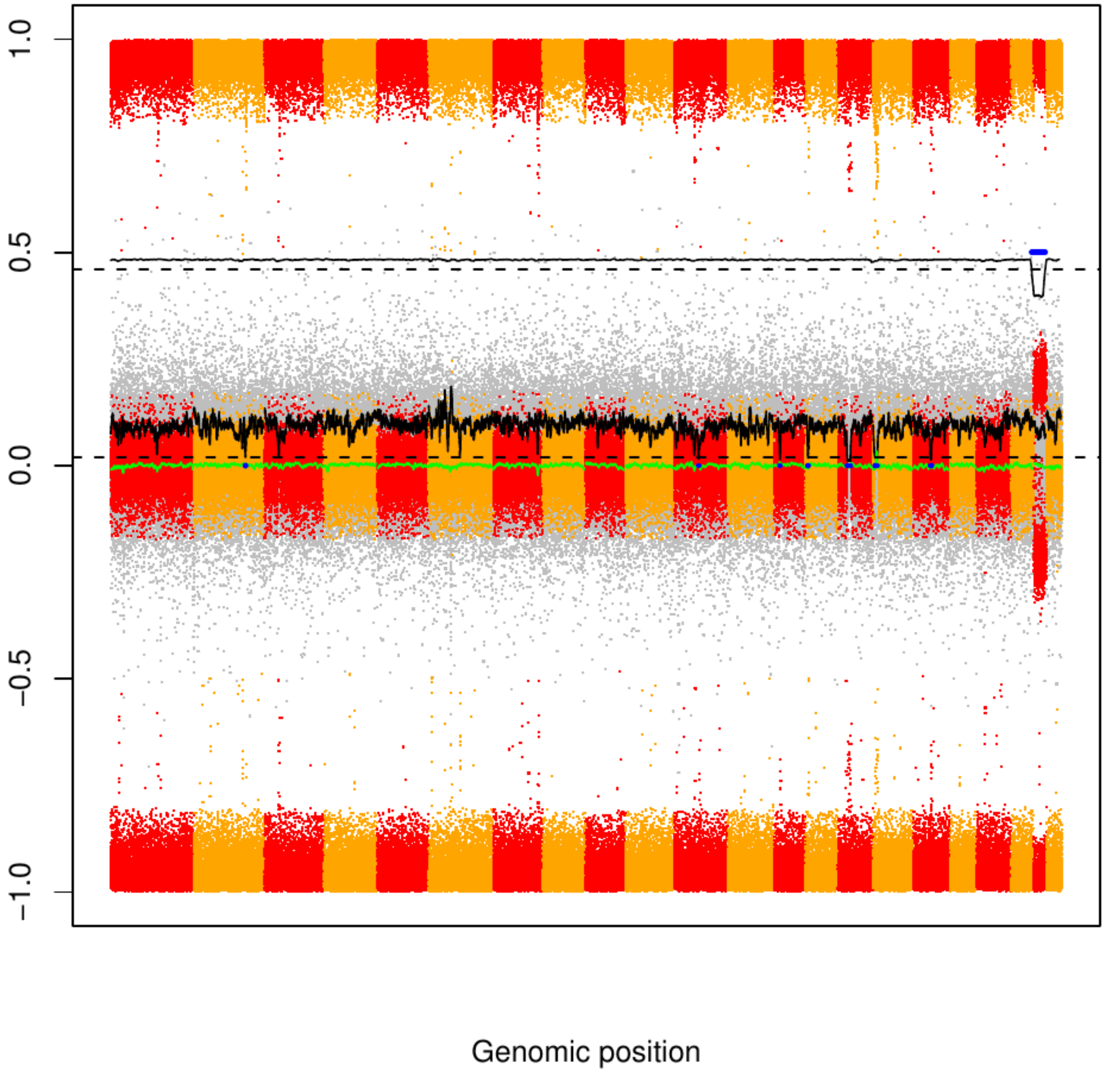

### CME75

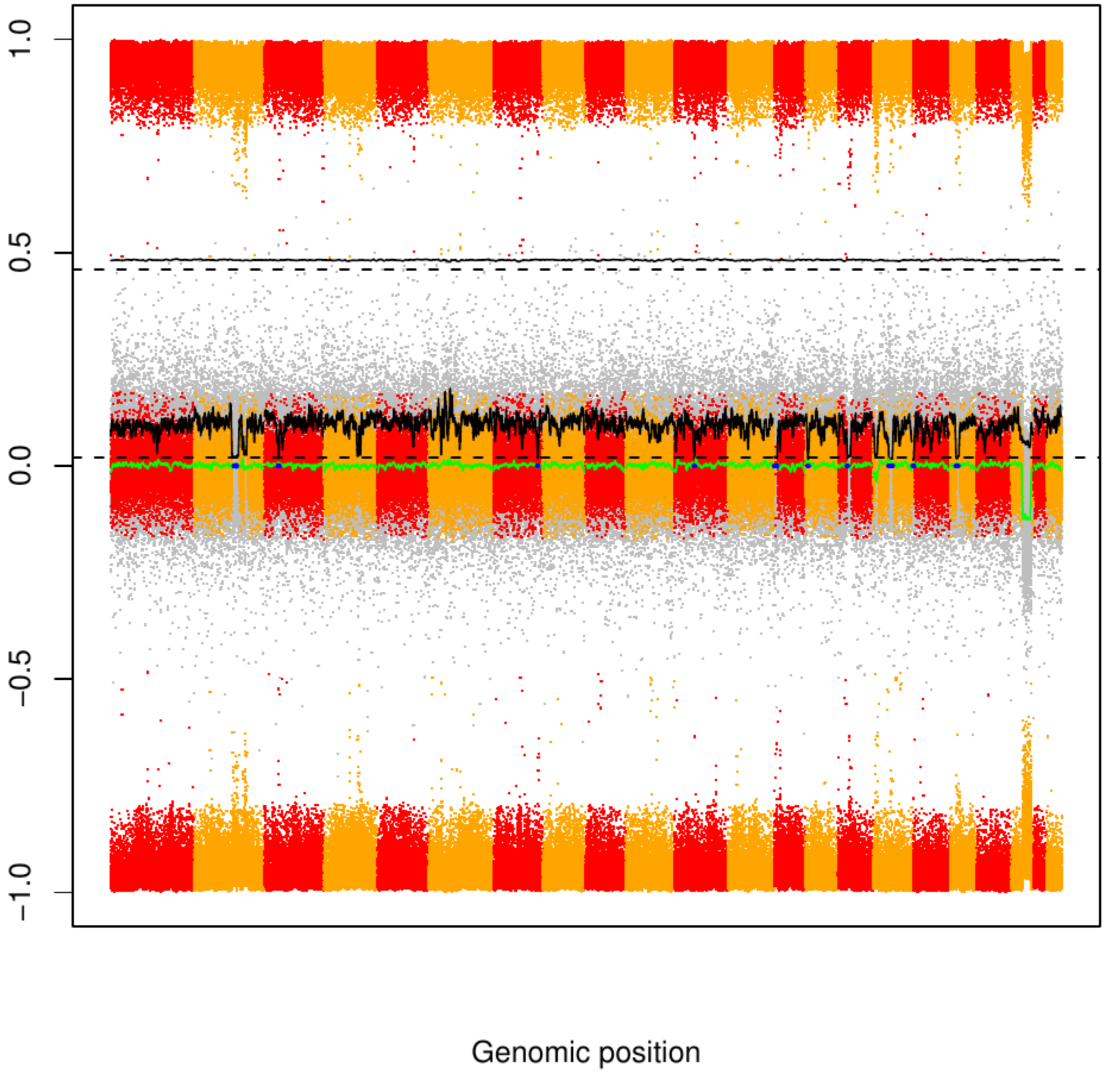

### CME76

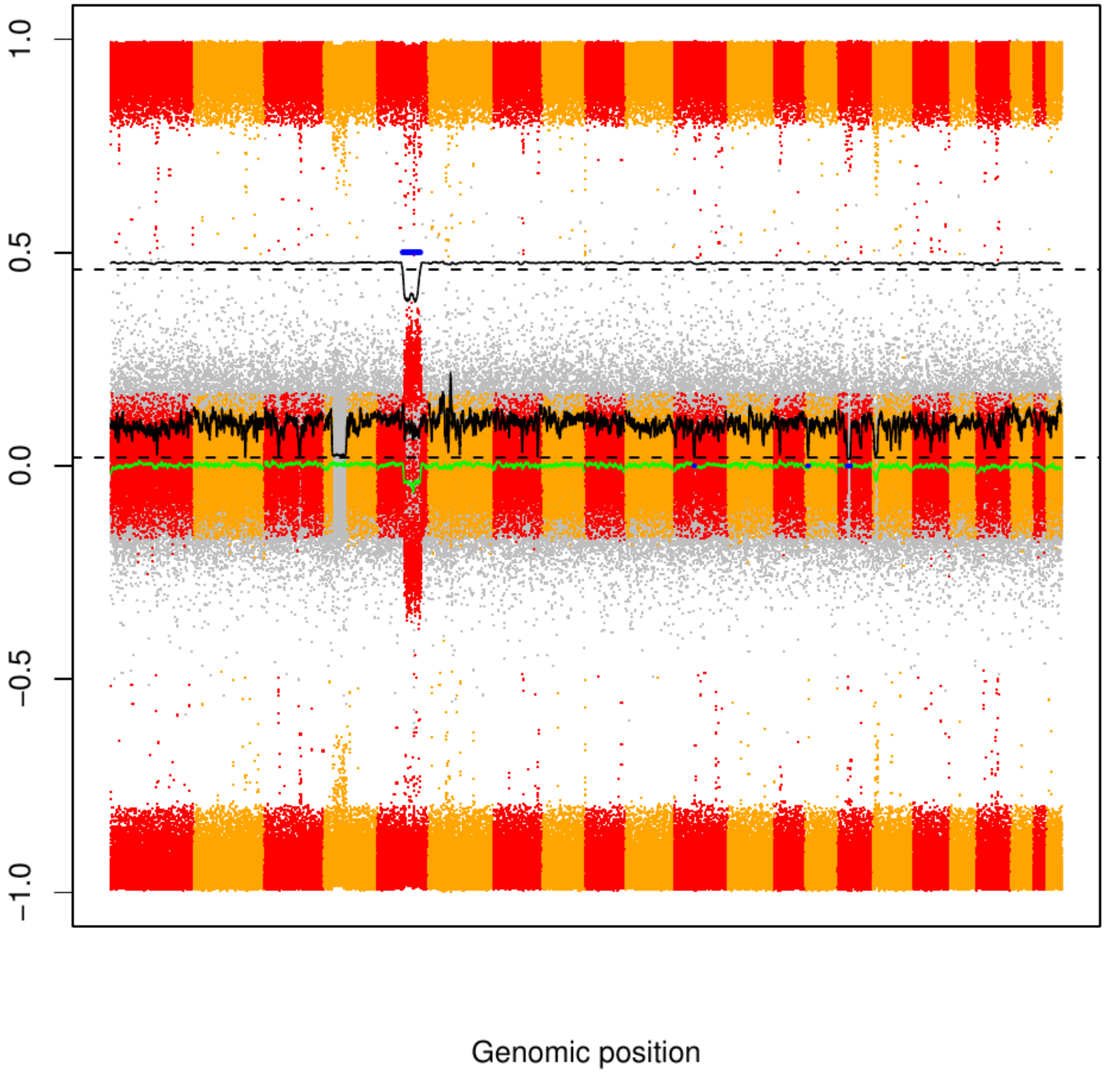

### CME77

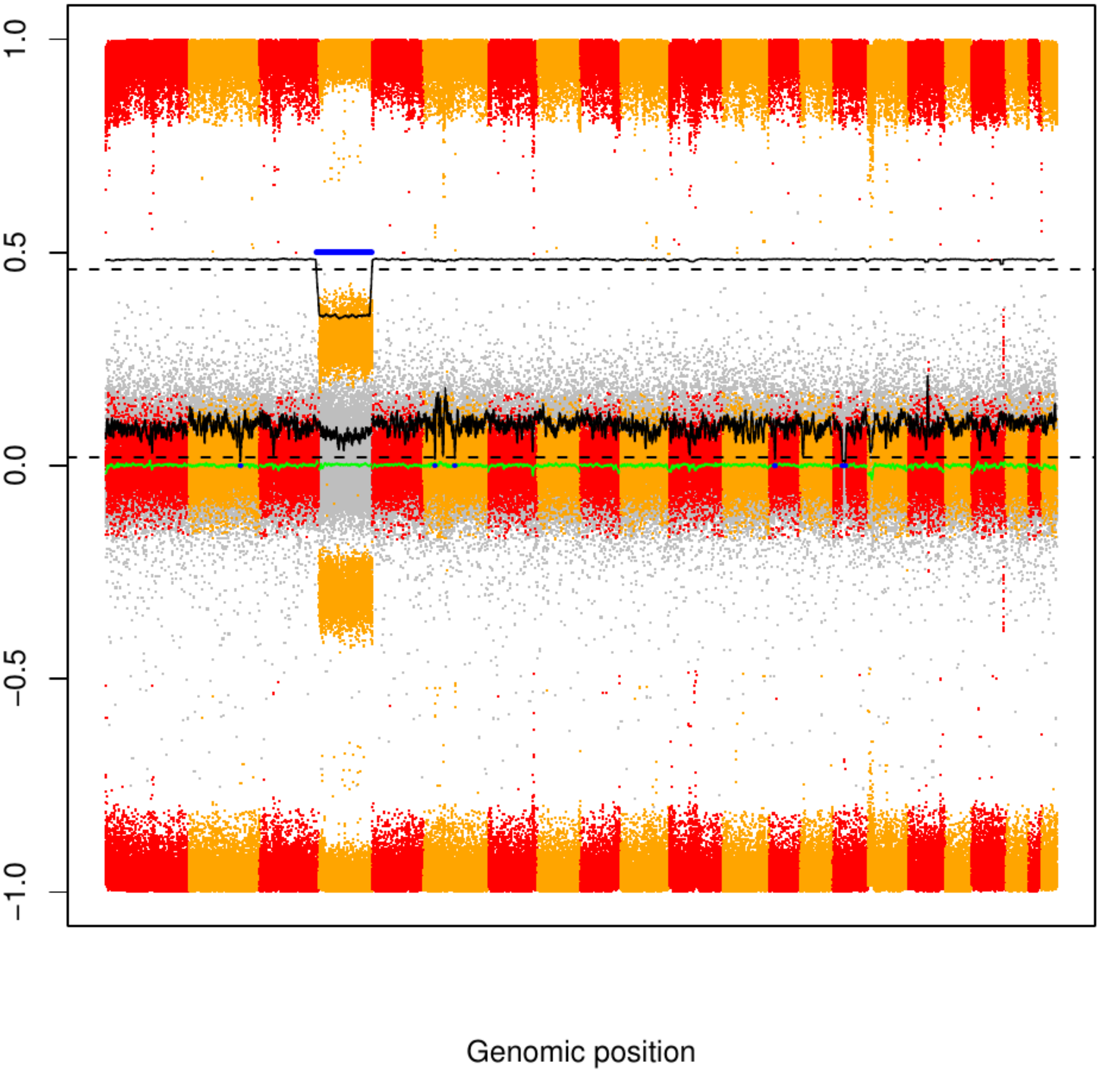

### CME78

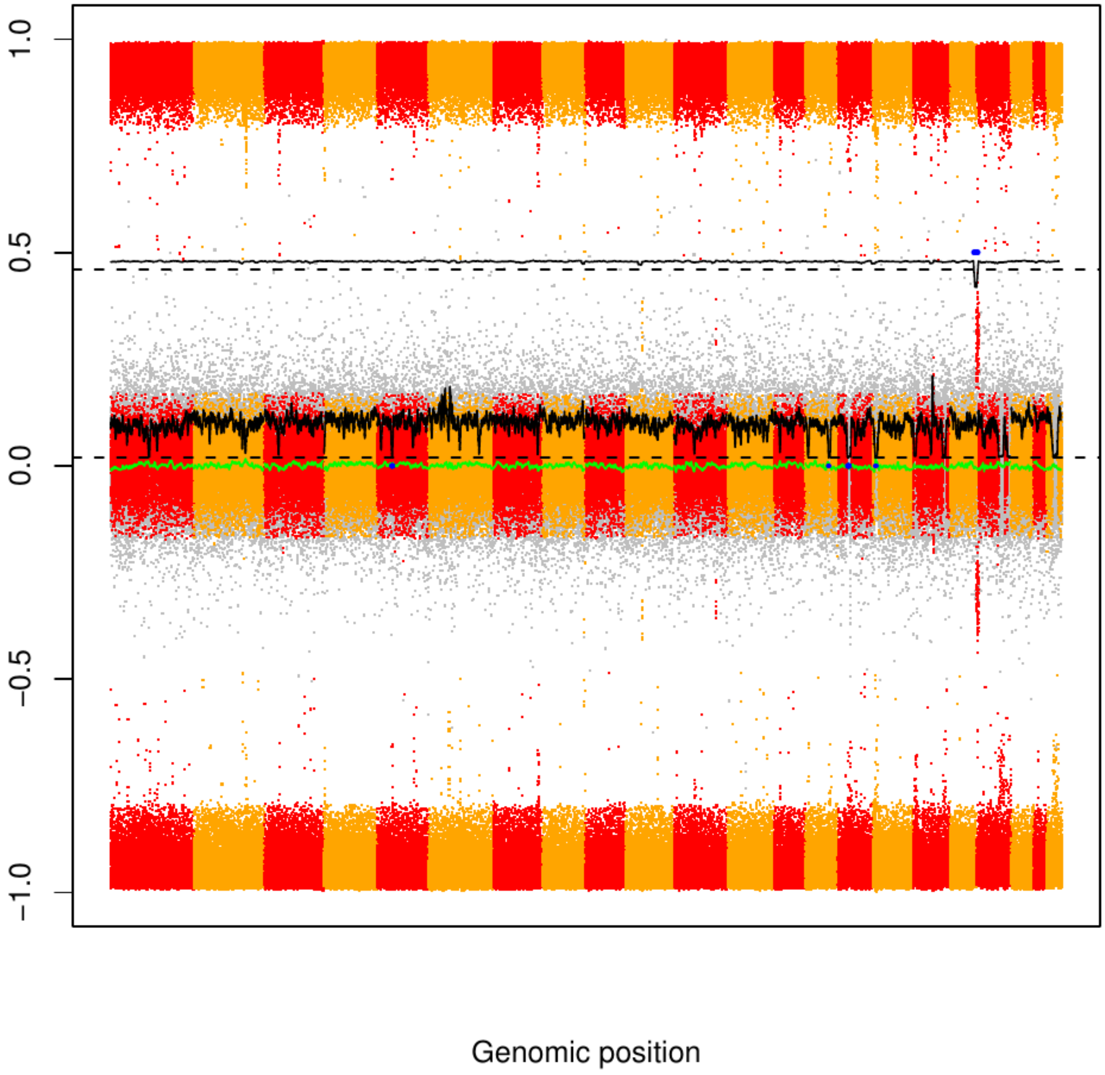

### CME79

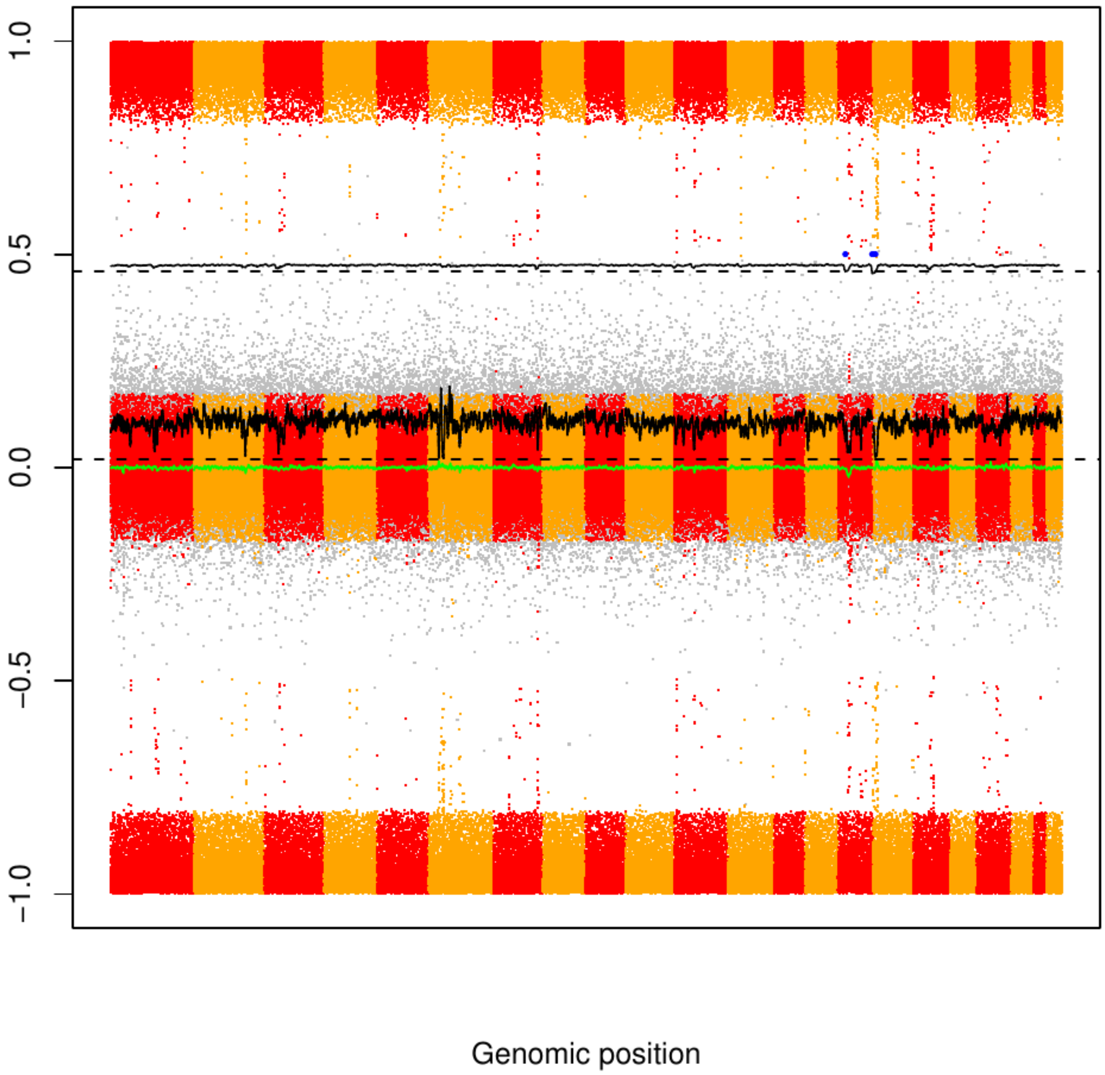

### CME80

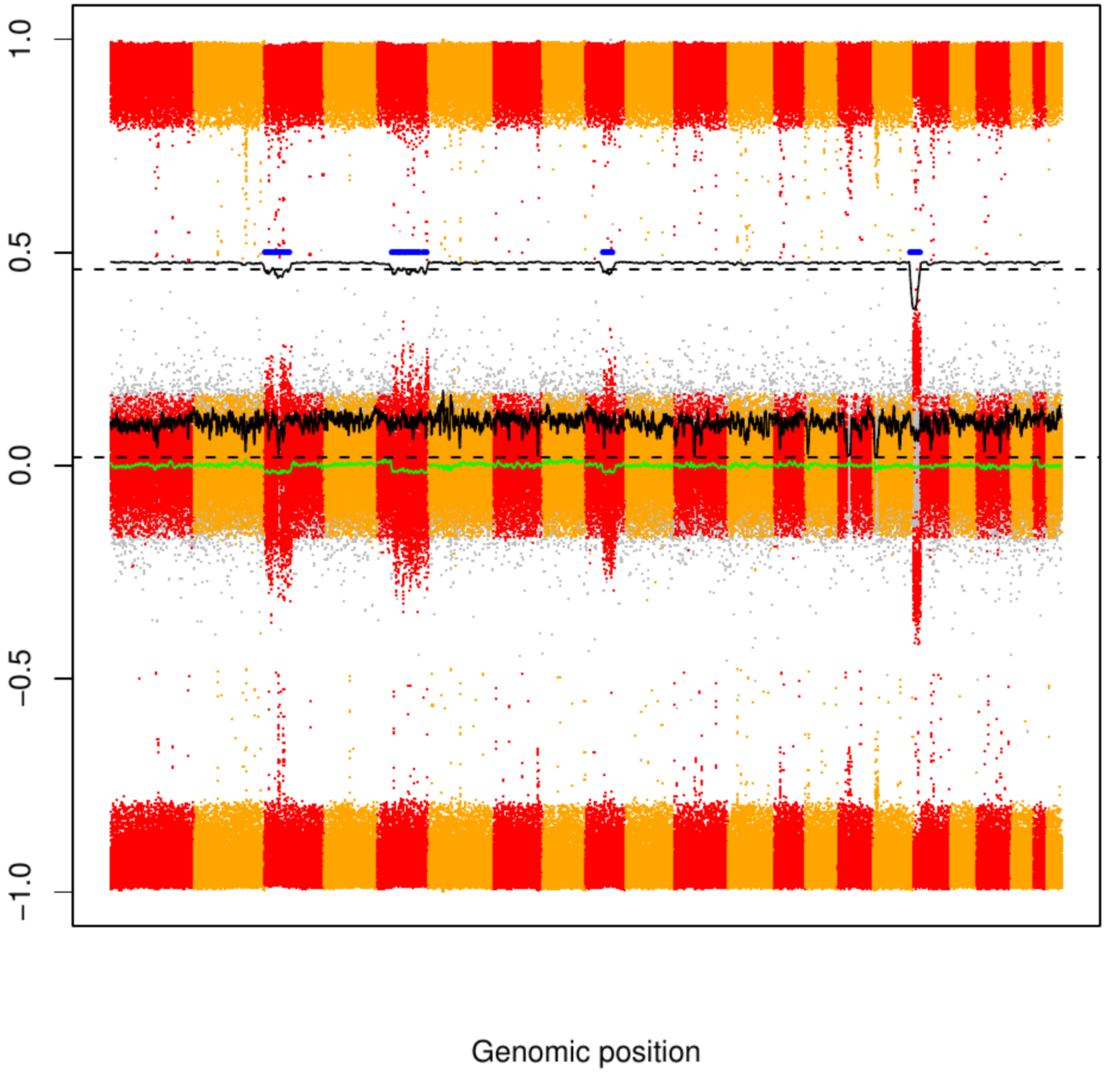

### CME81

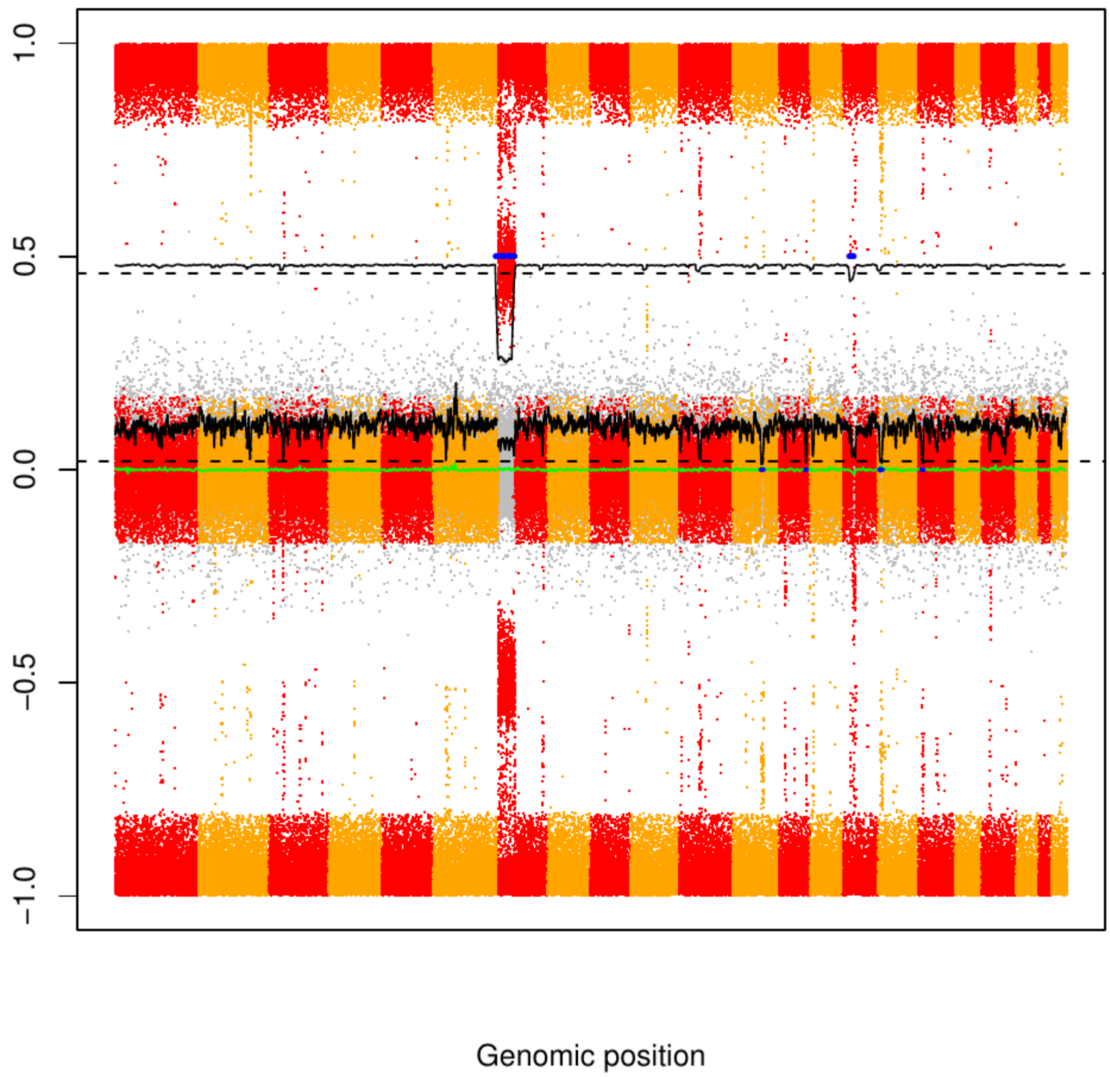

CME82

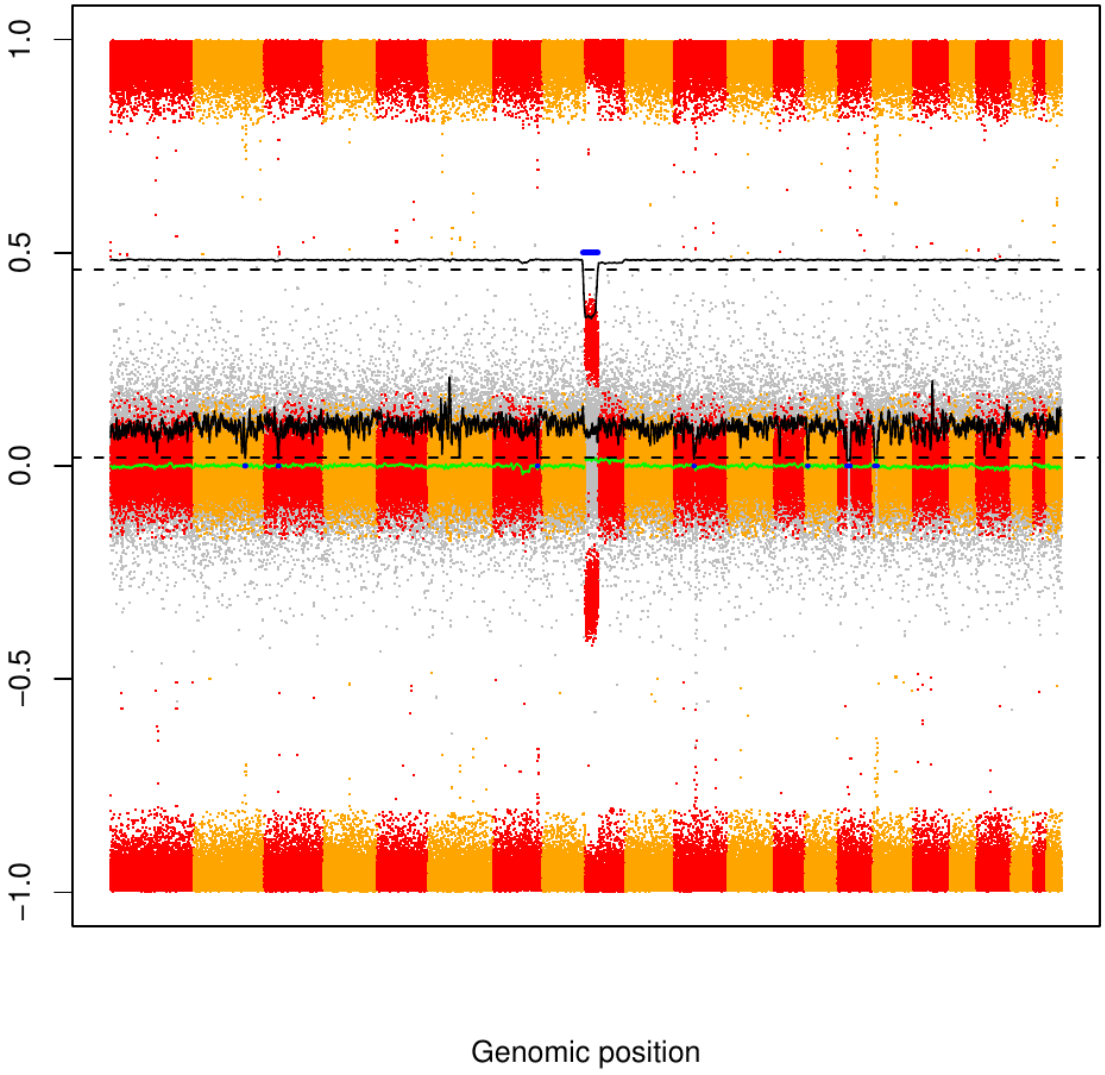

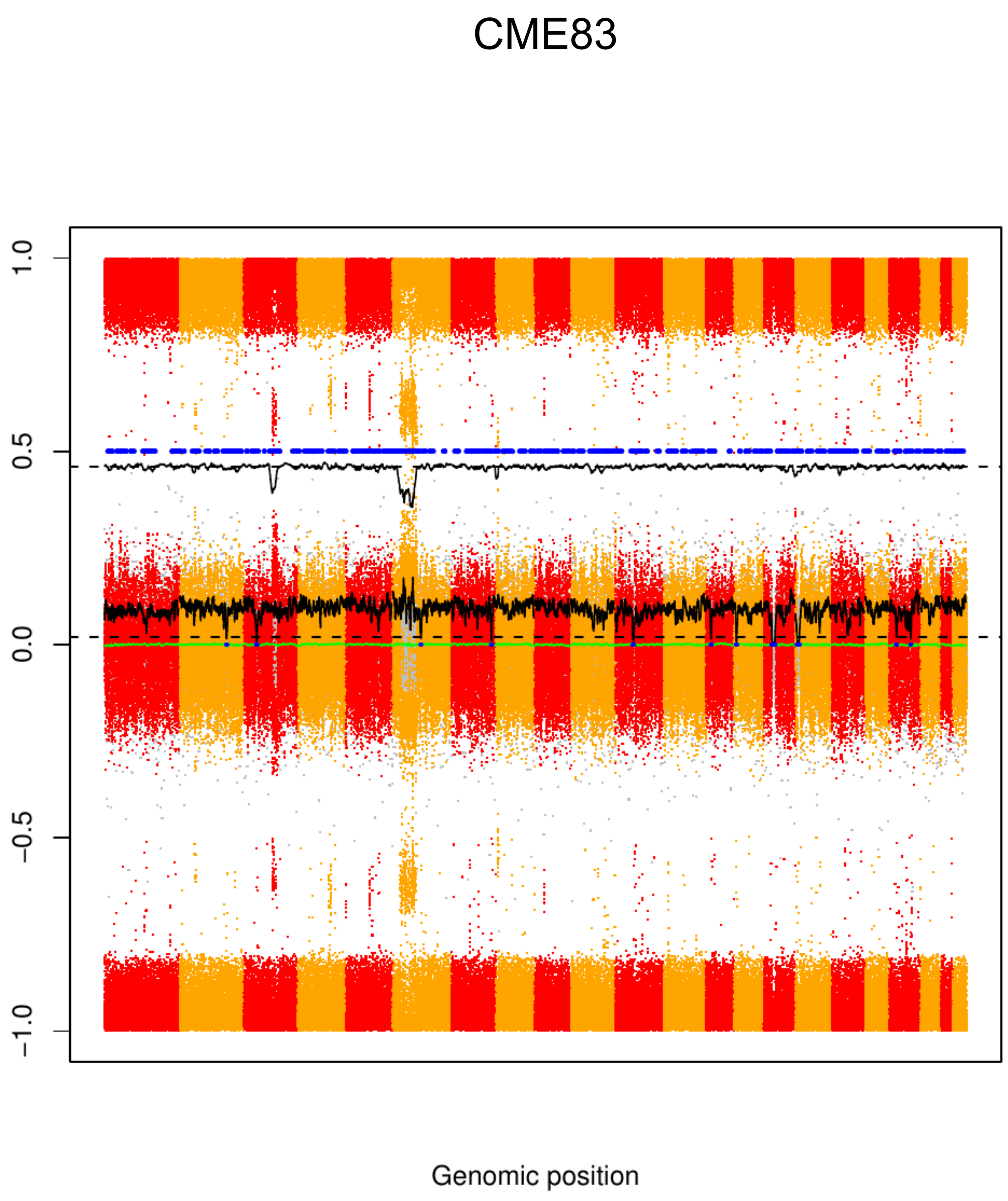
CME83
1.0
0.5
0.0
−0.5
−1.0
Genomic position

CME84

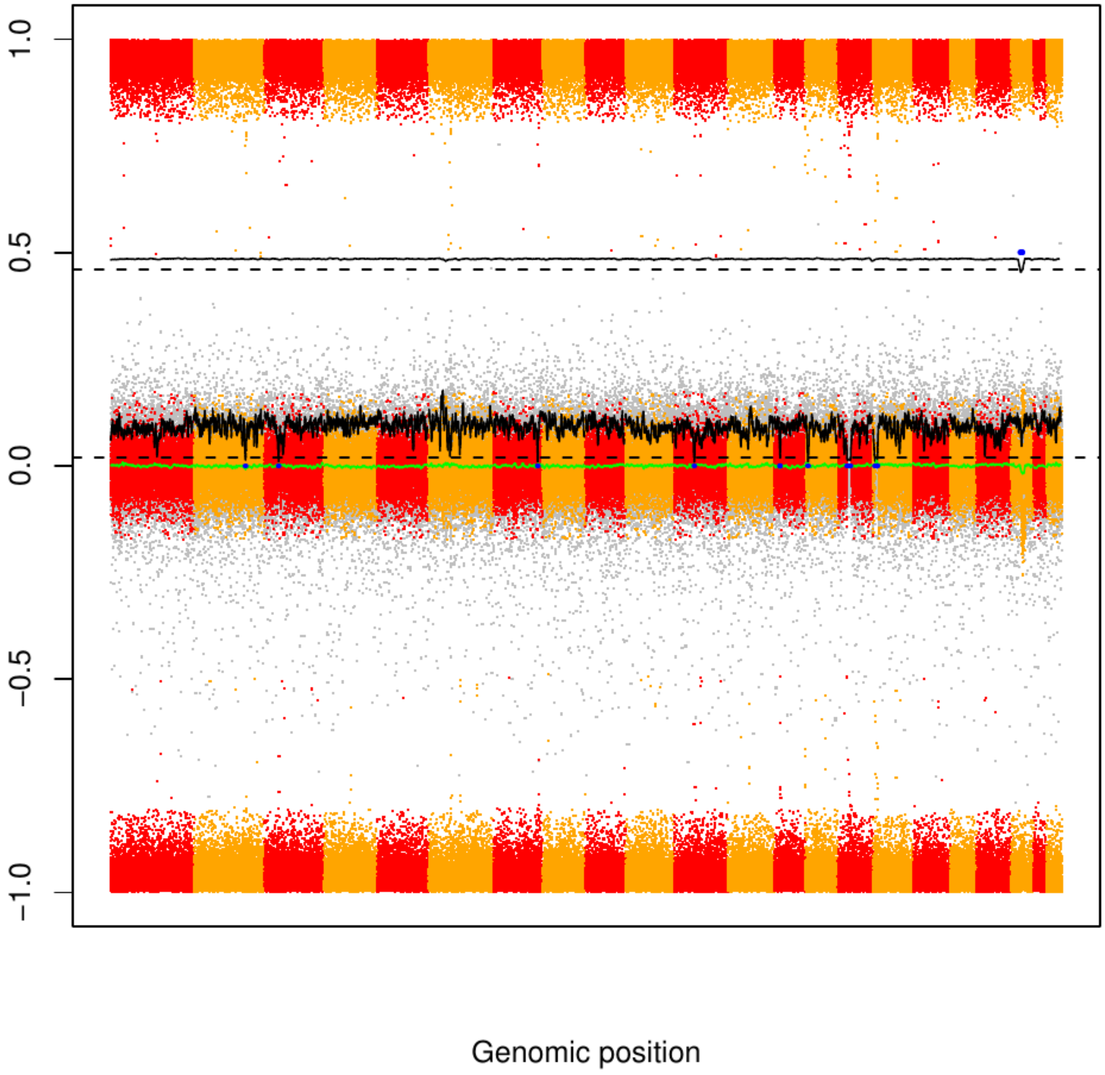

CME85

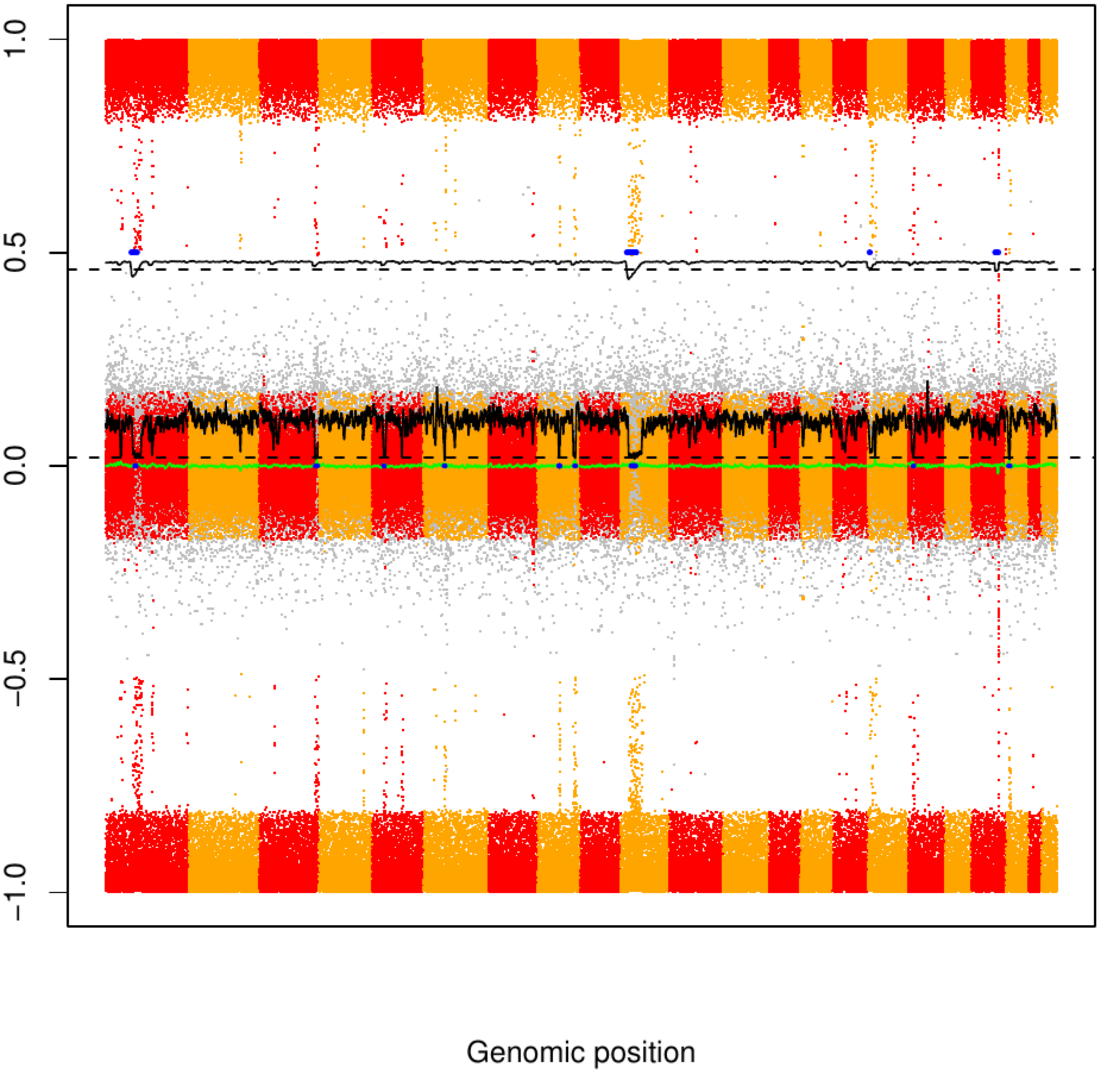

### CME86

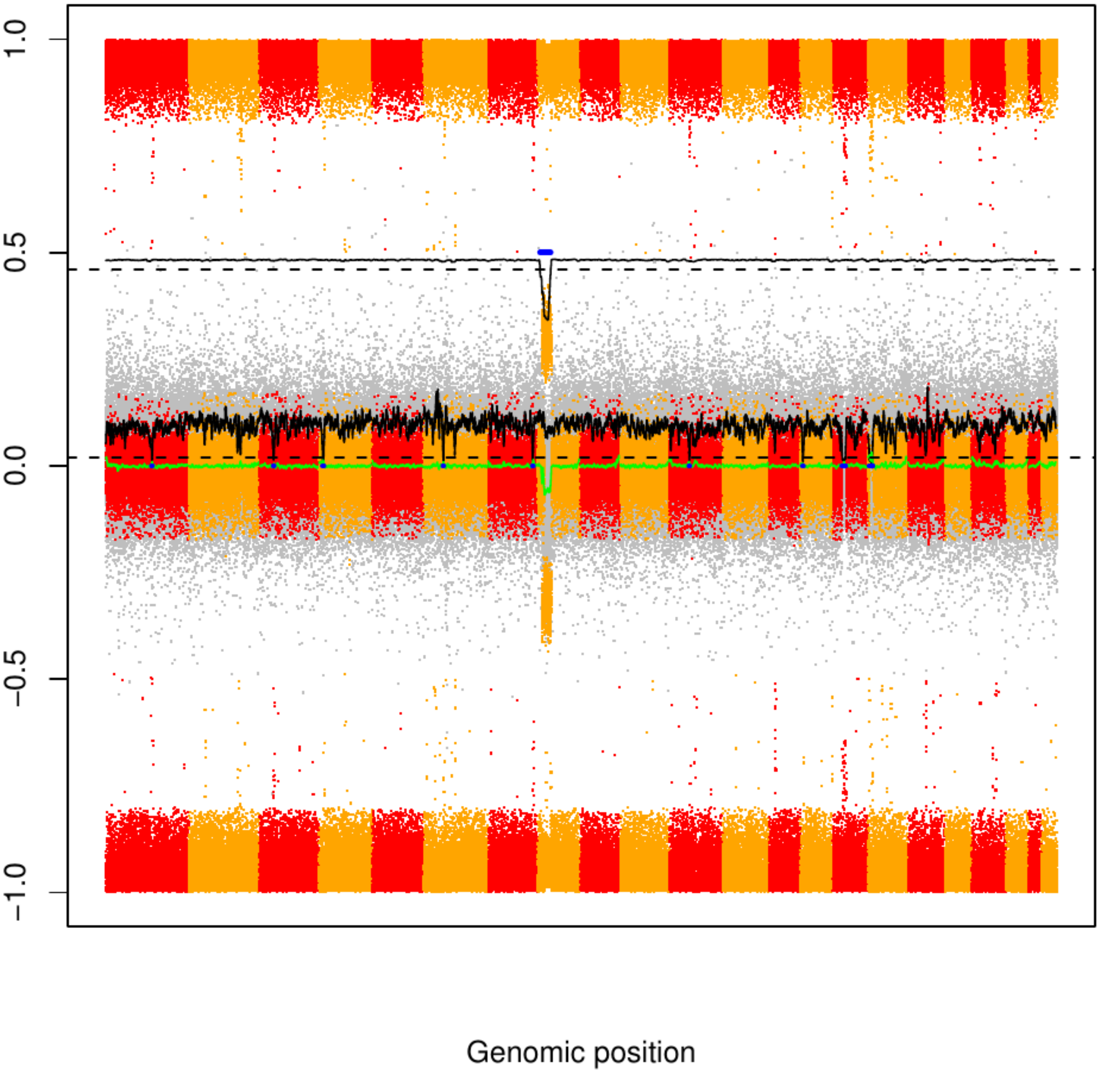

### CME87

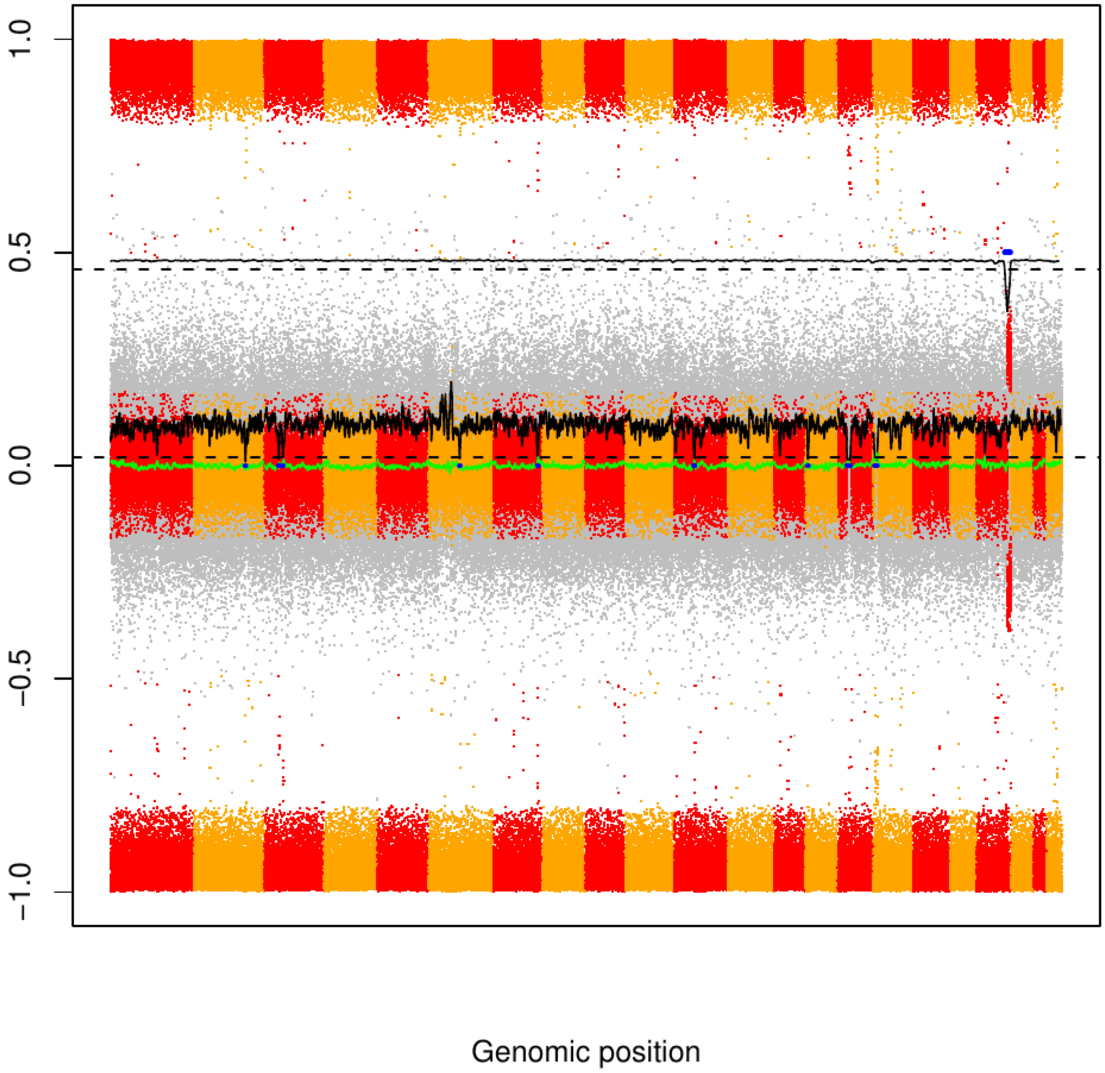

CME88

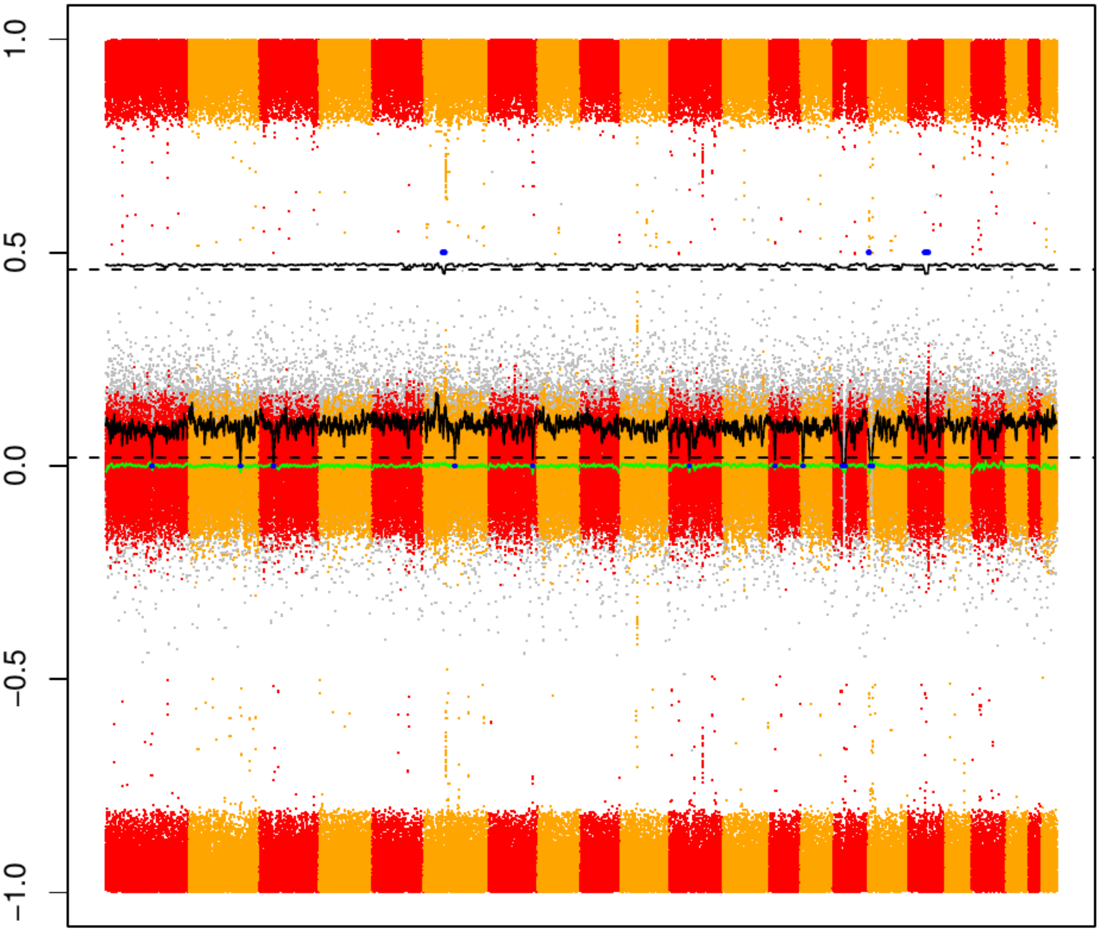

### CME89

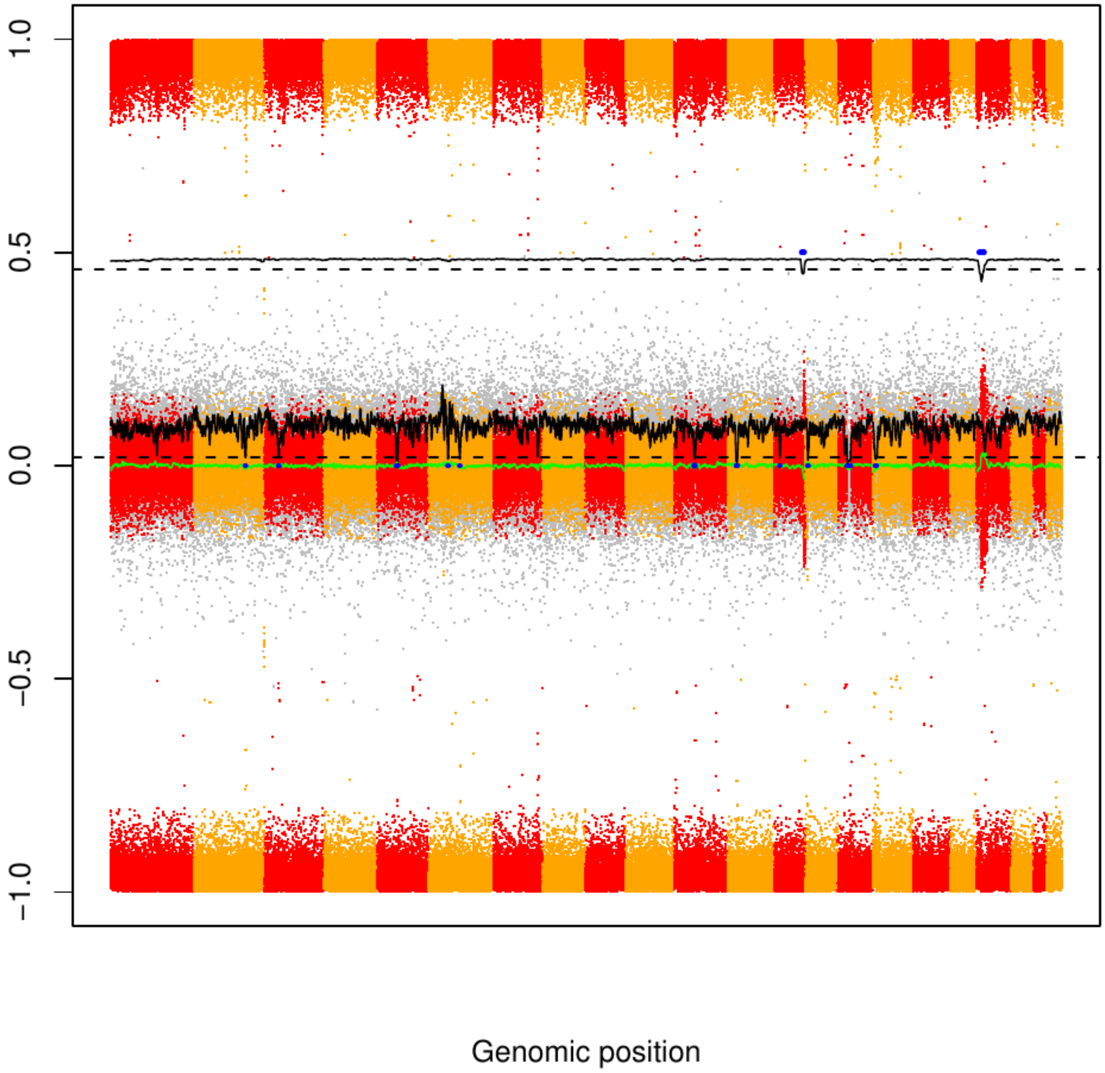

### CME90

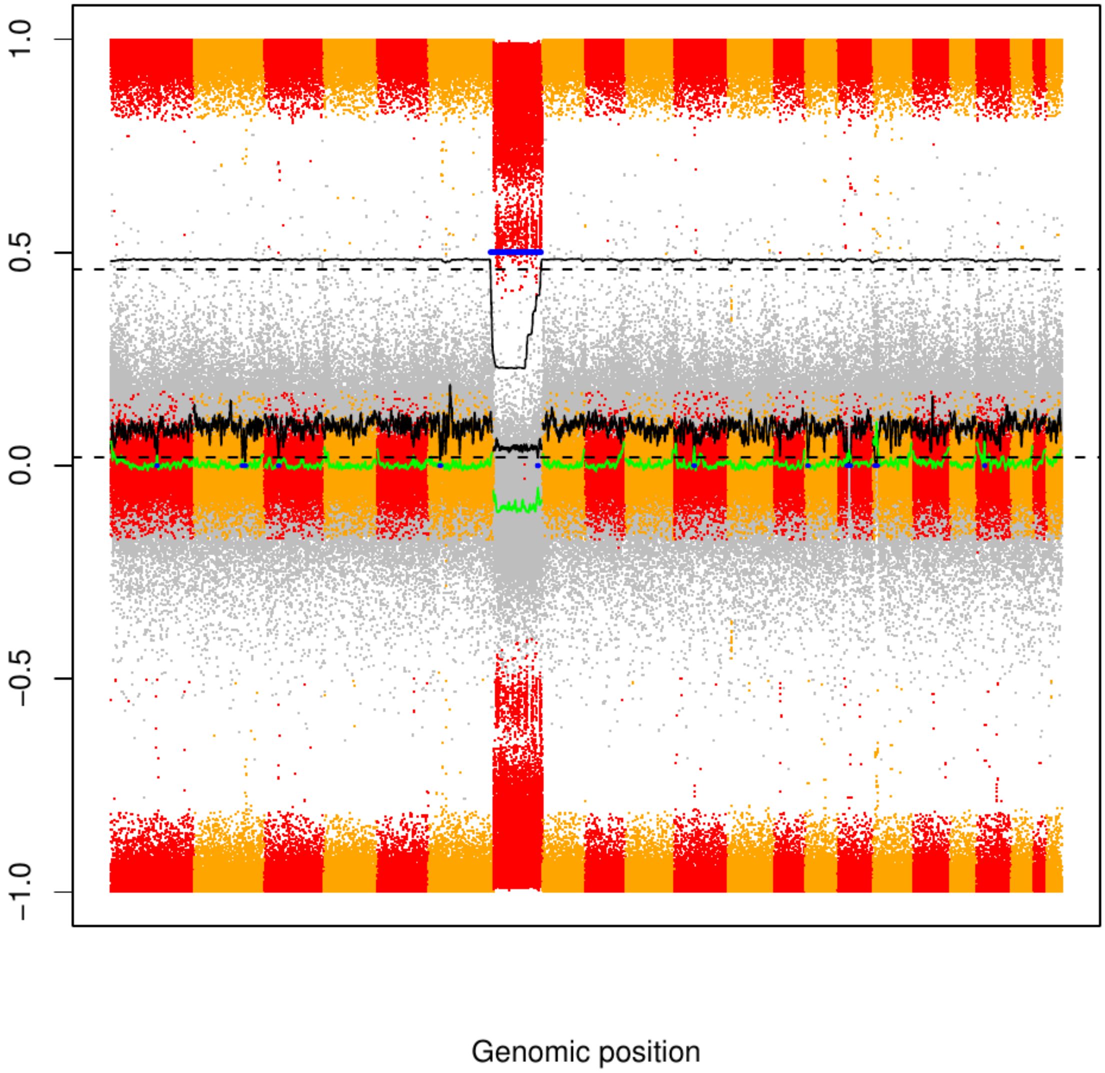

### CME91

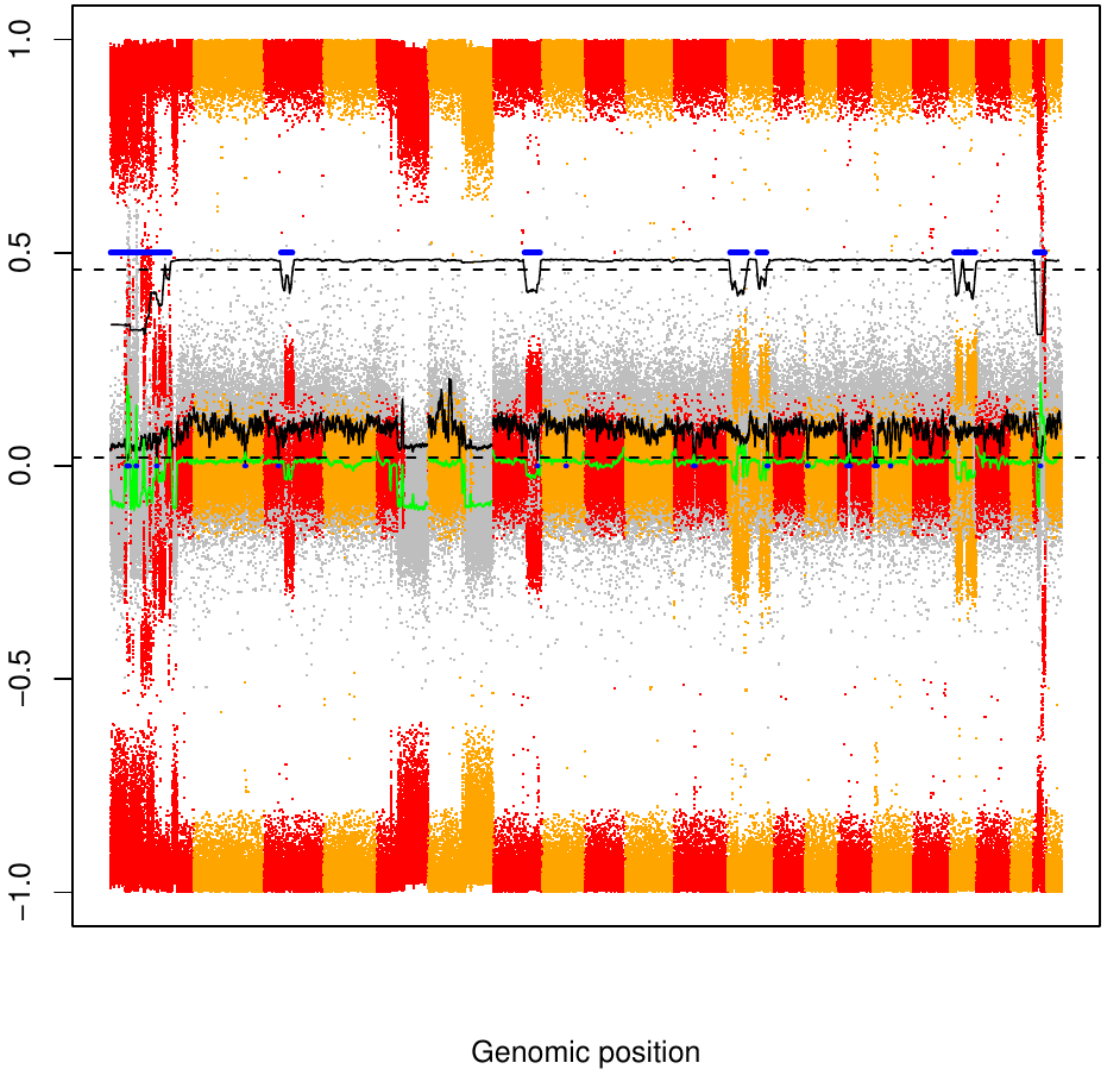

### CME92

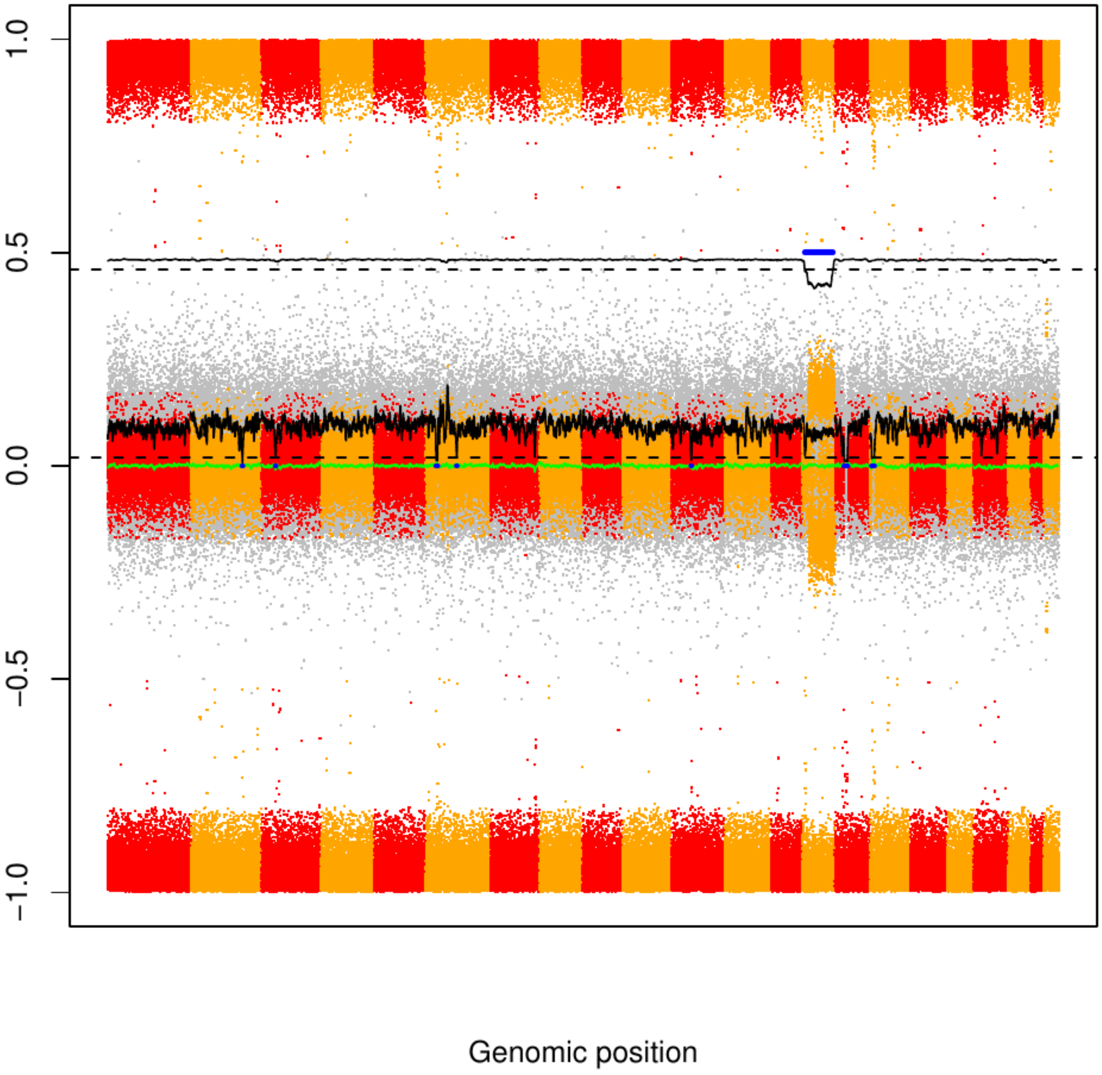

### CME93

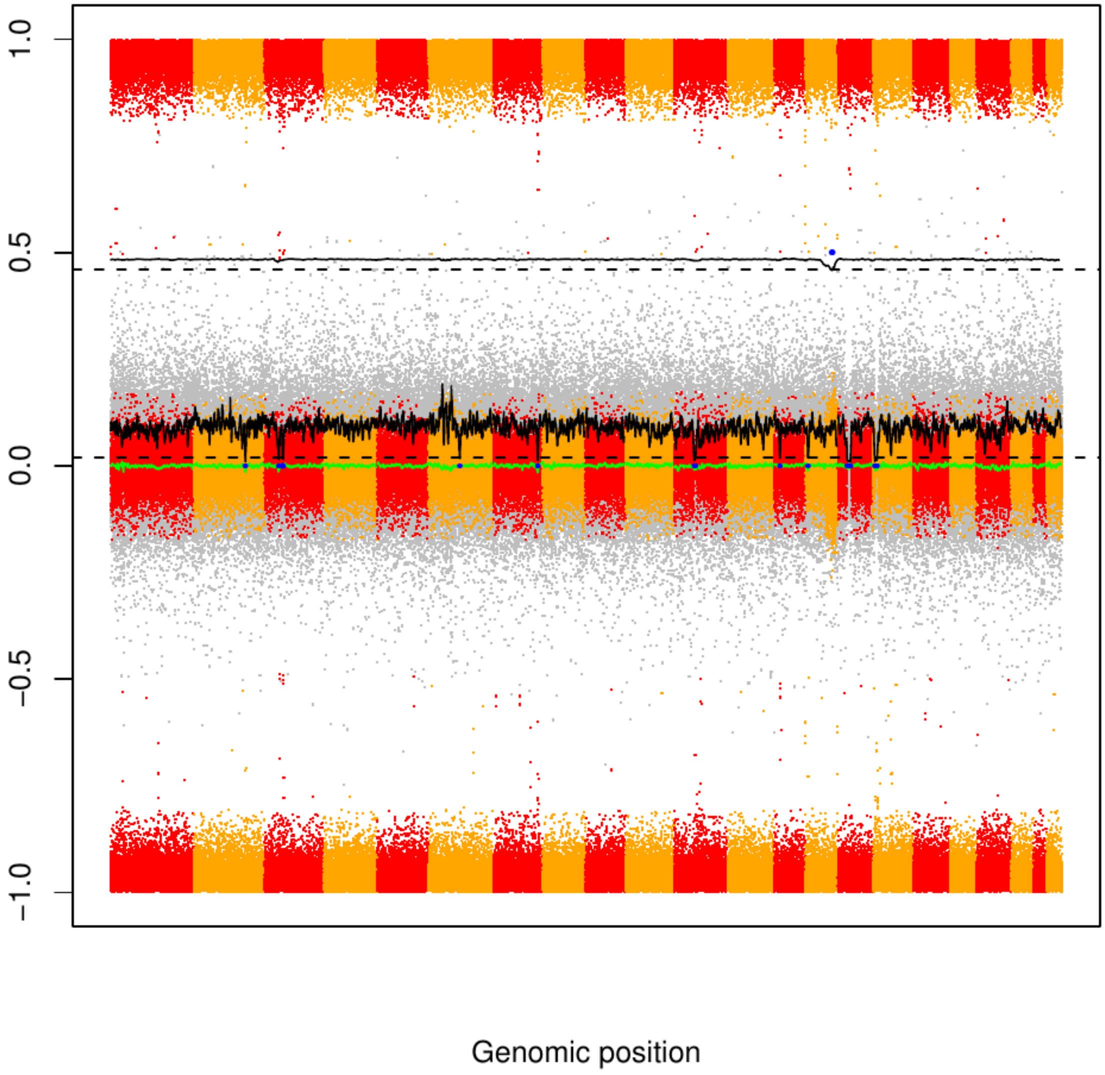

CME94

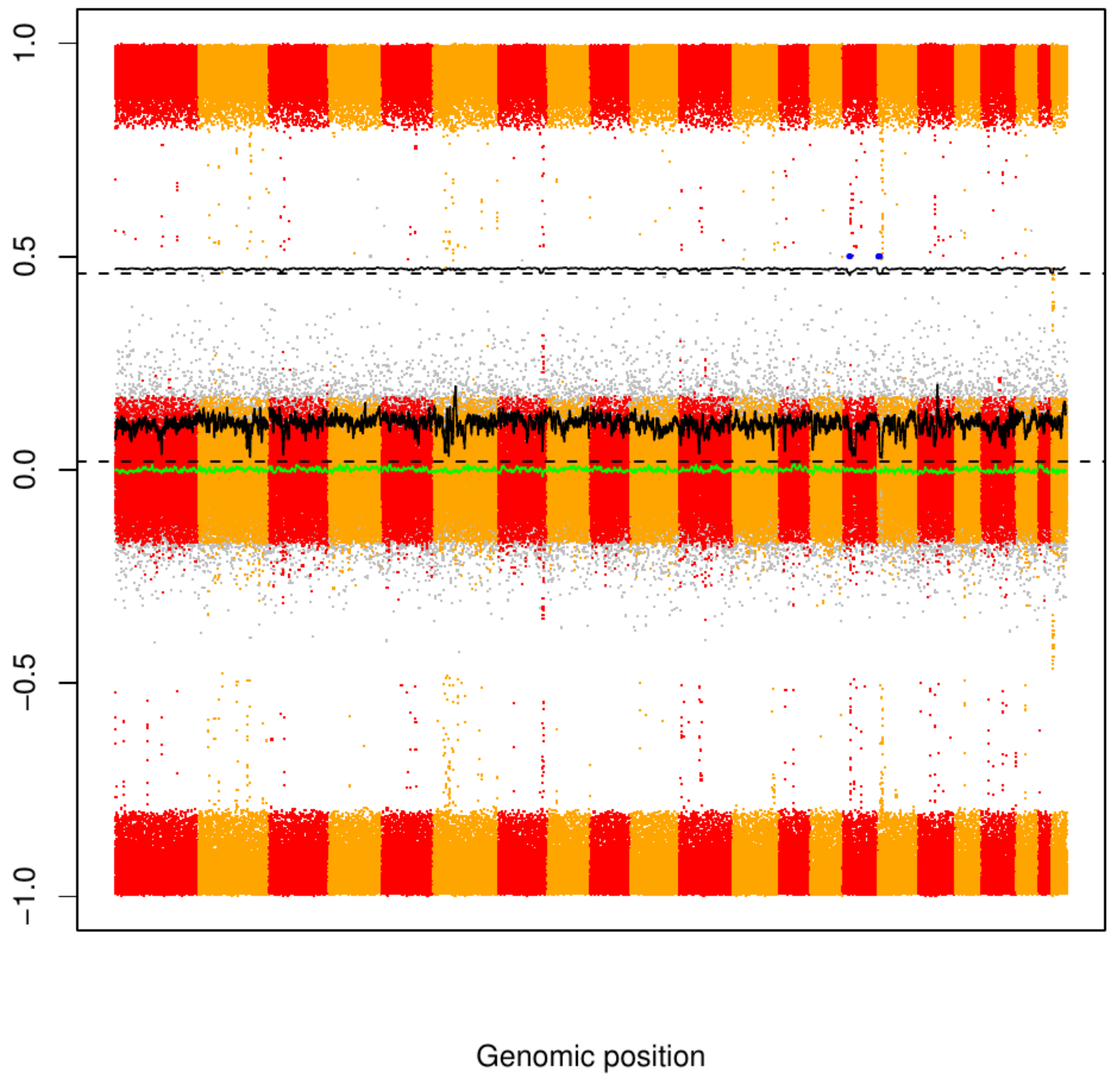

### CME95

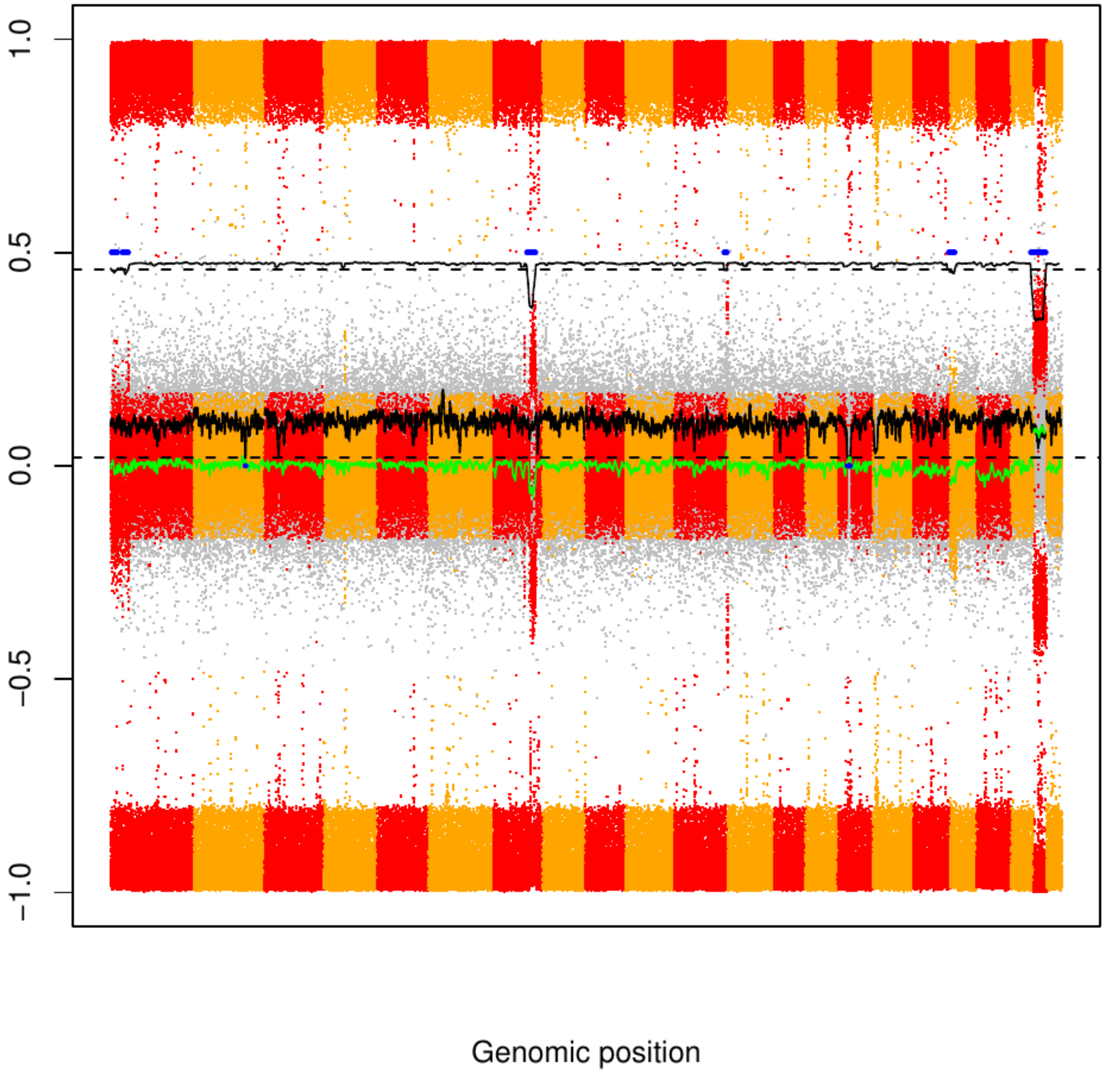

### CME96

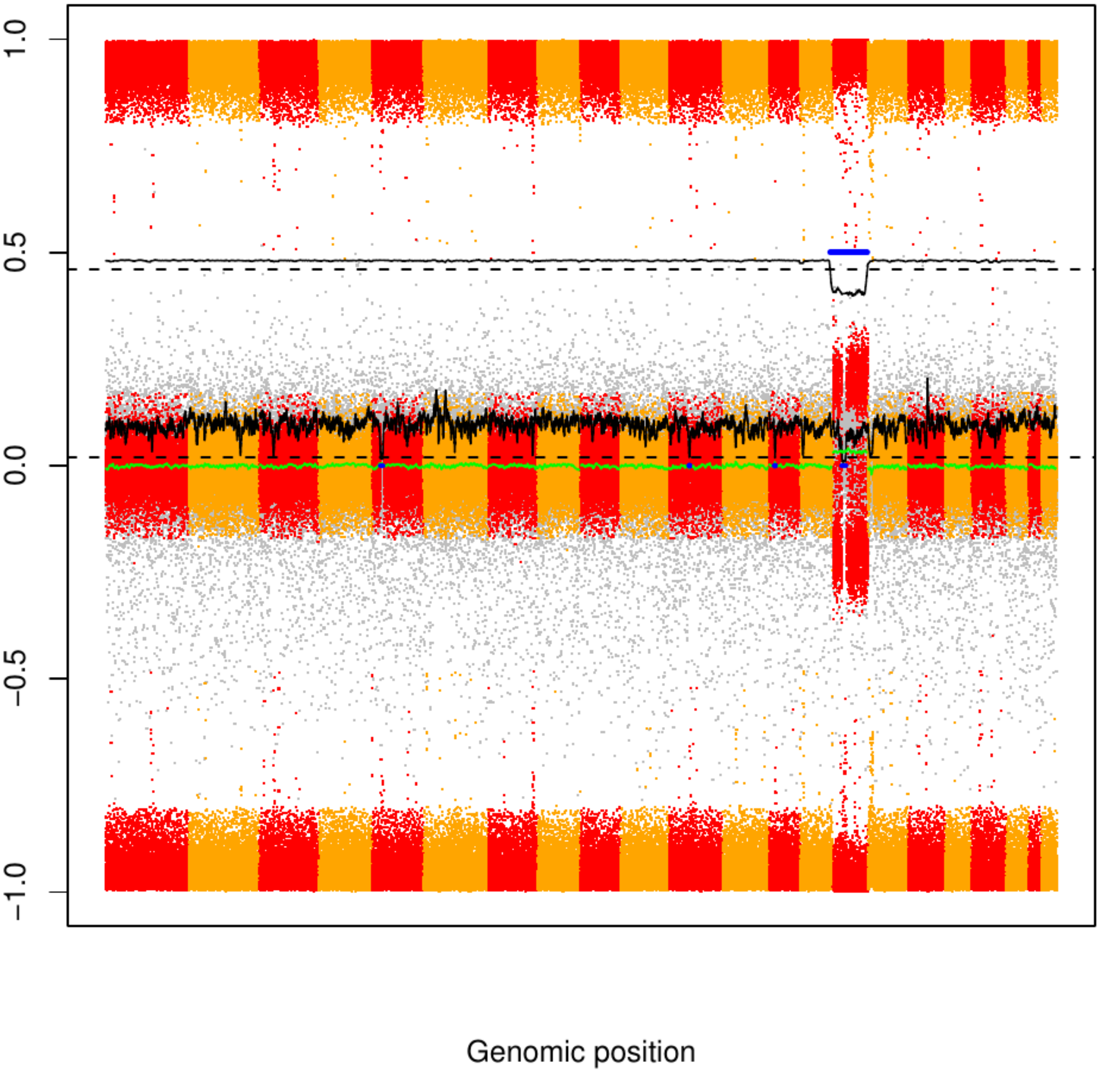

### CME97

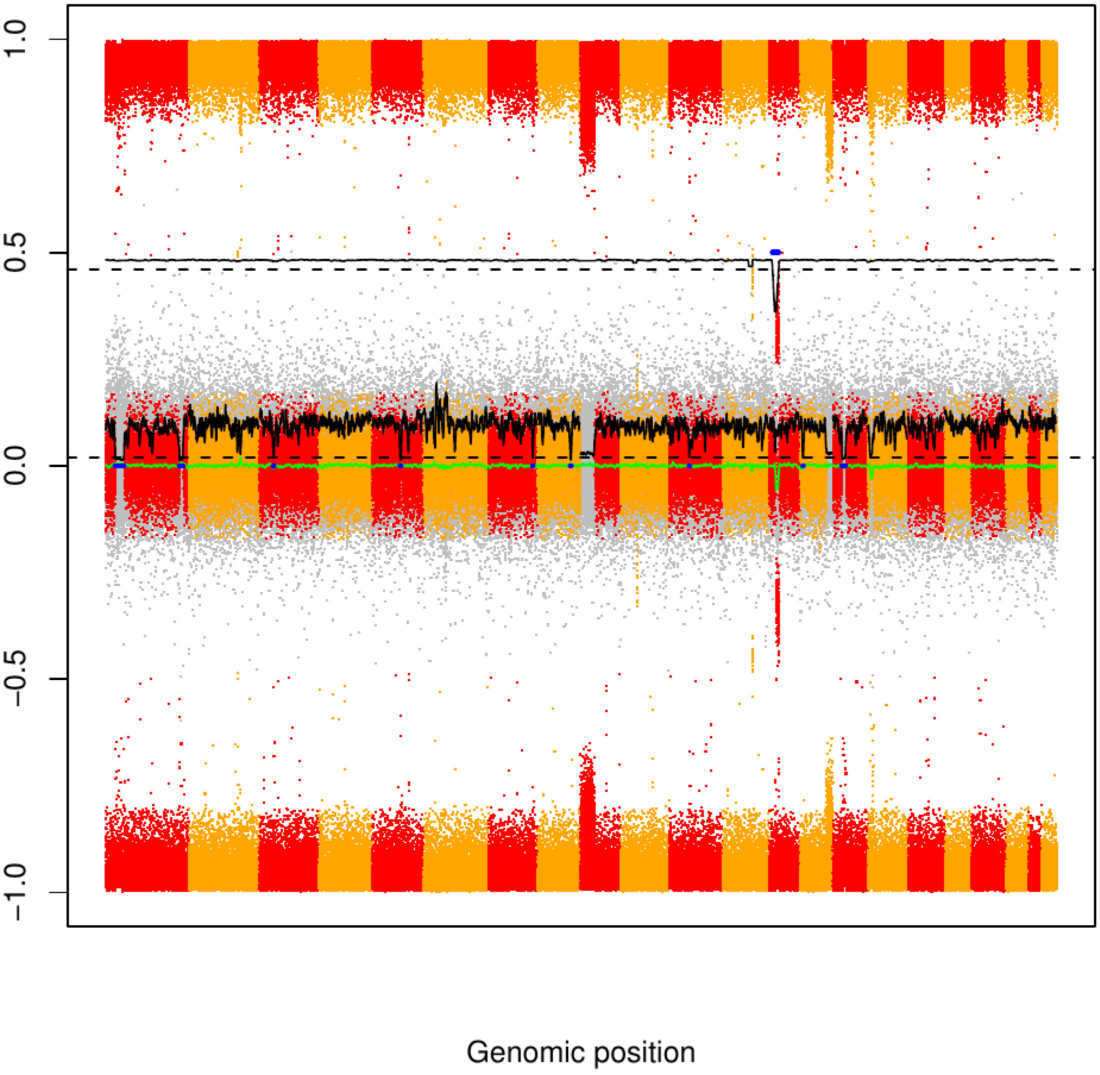

CME98

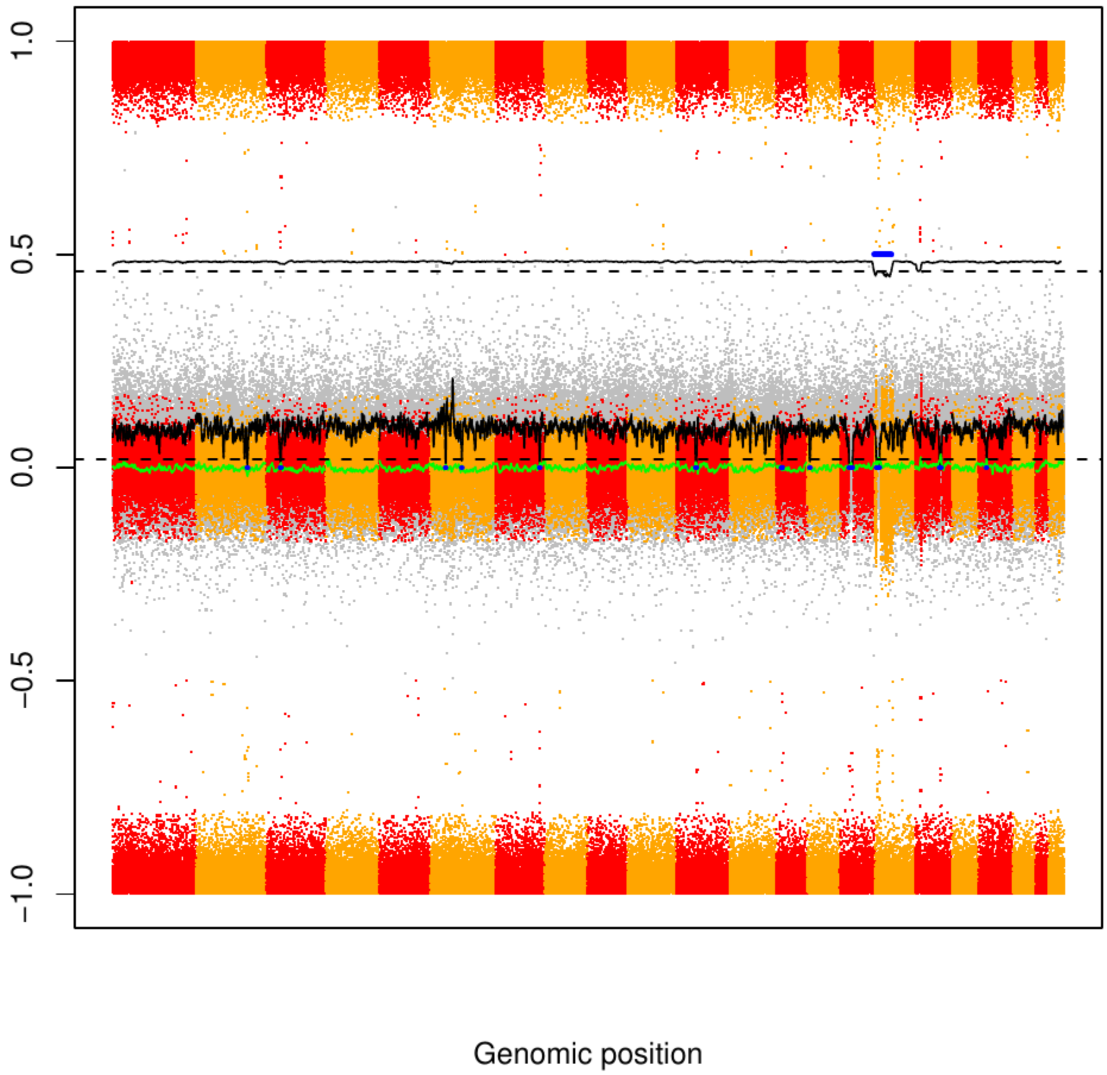

### CME99

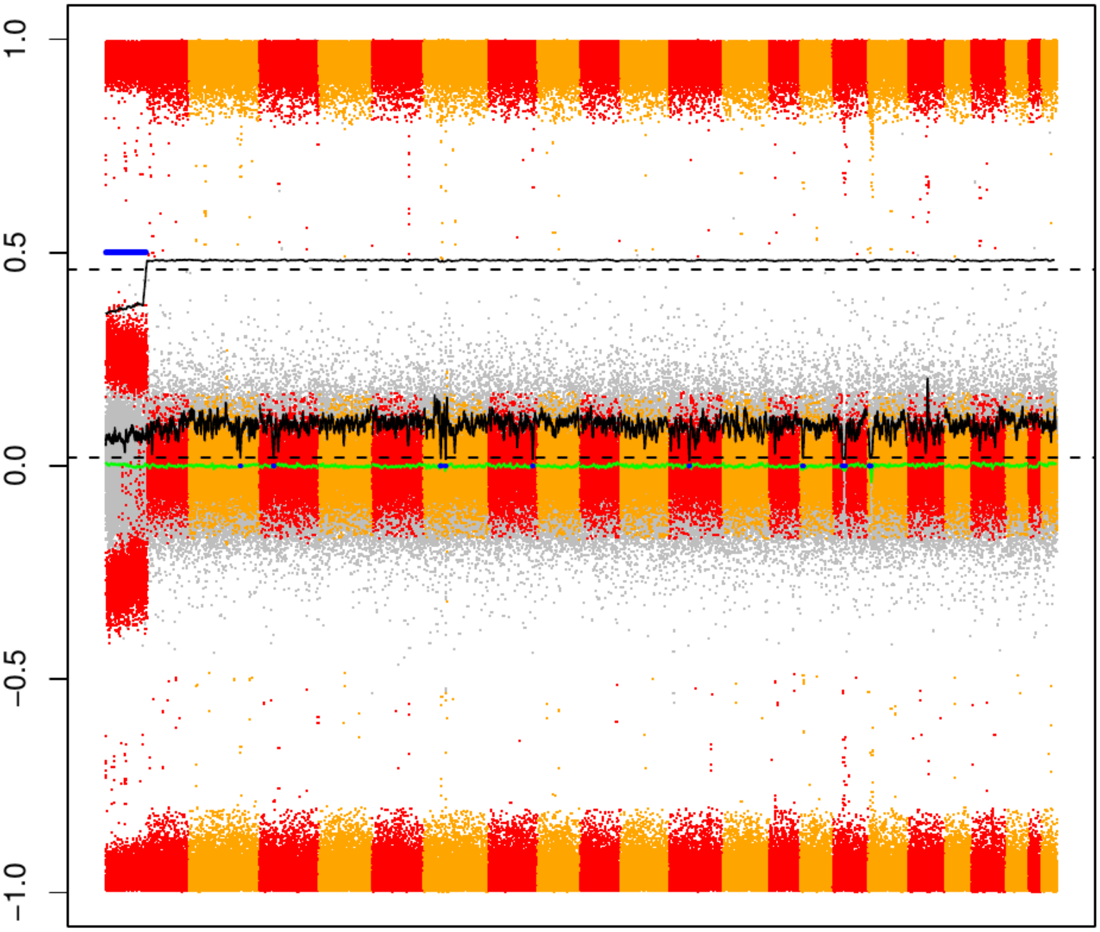

CME100

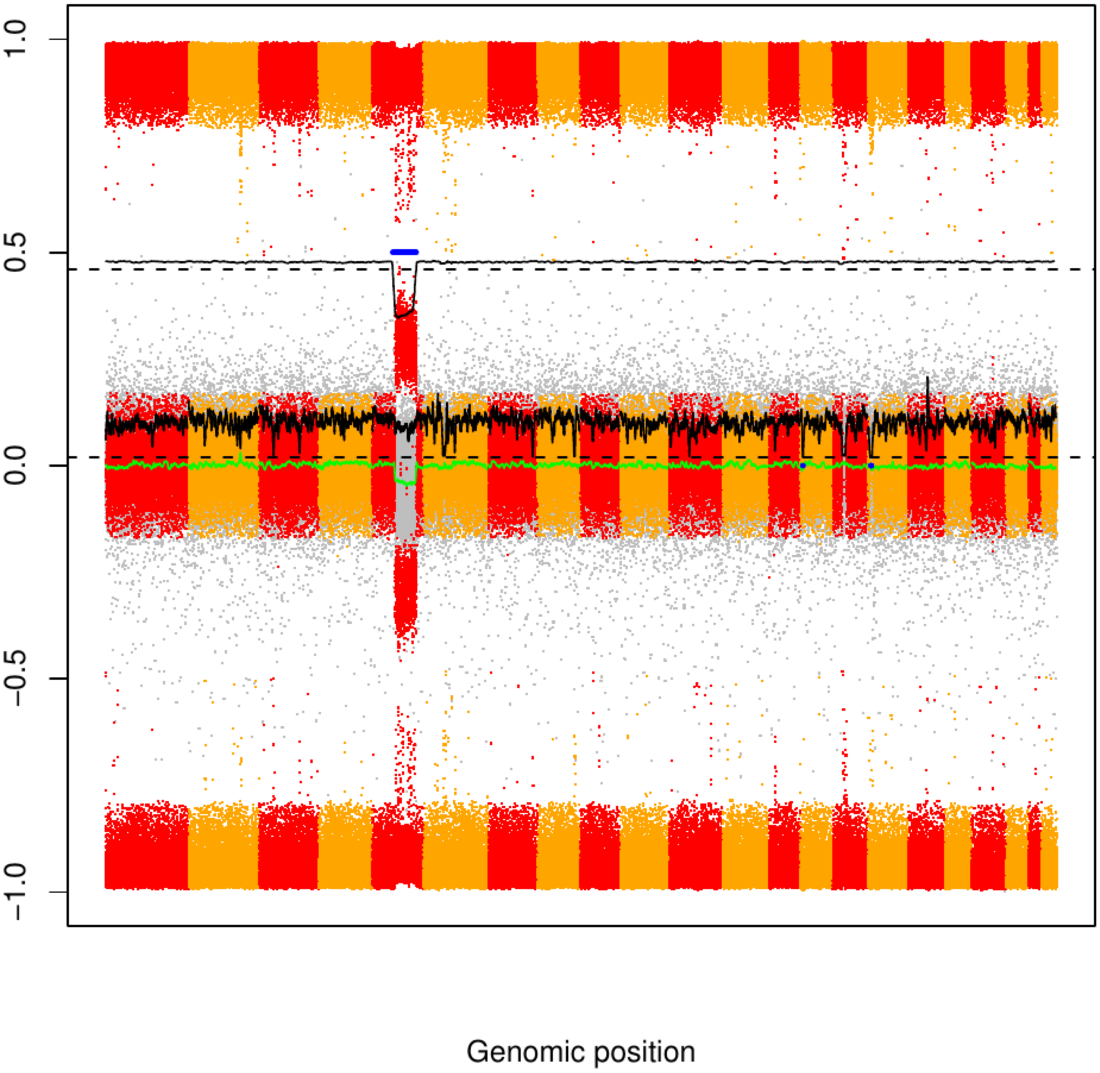

### CME101

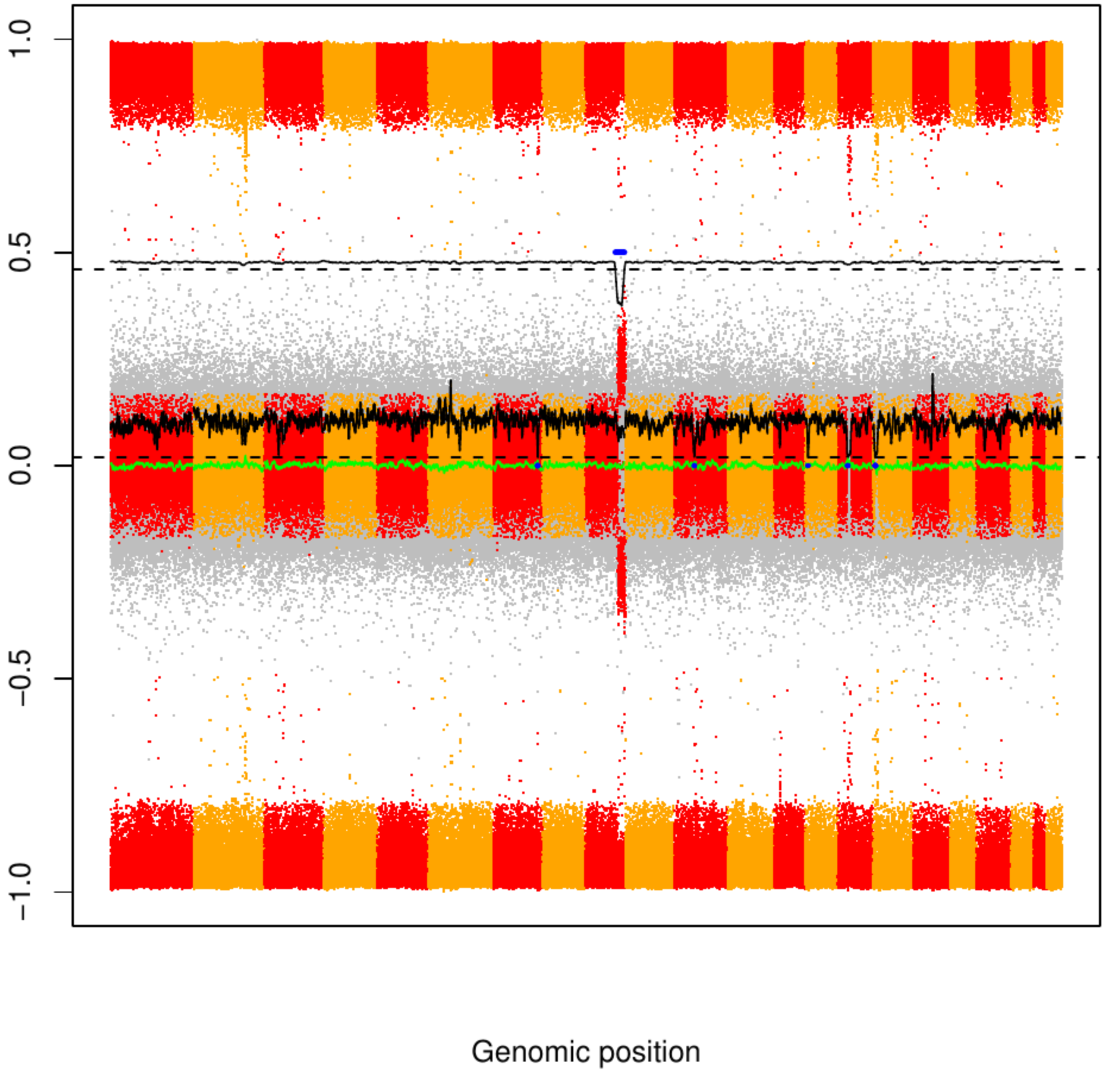

### CME102

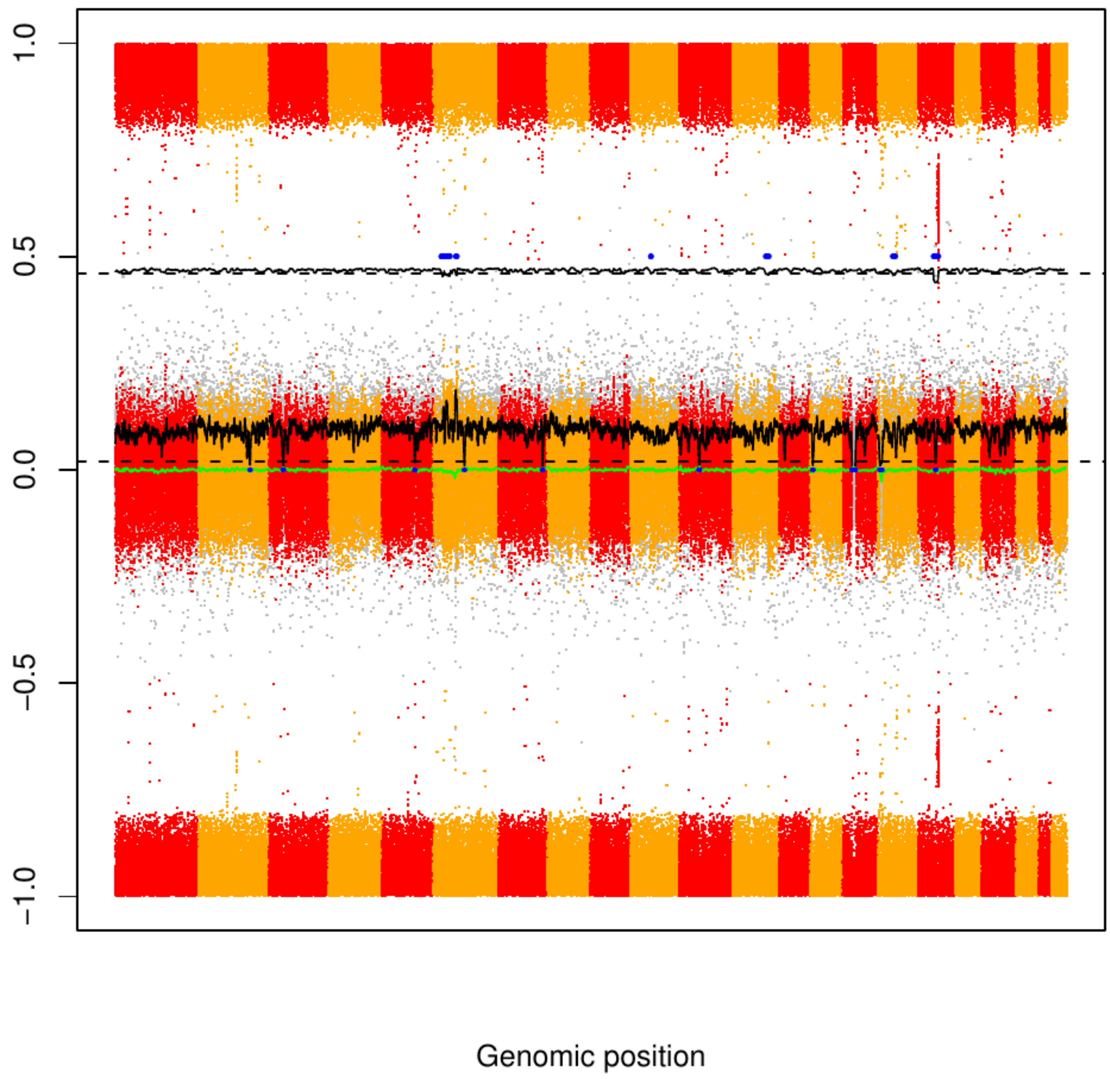

### CME103

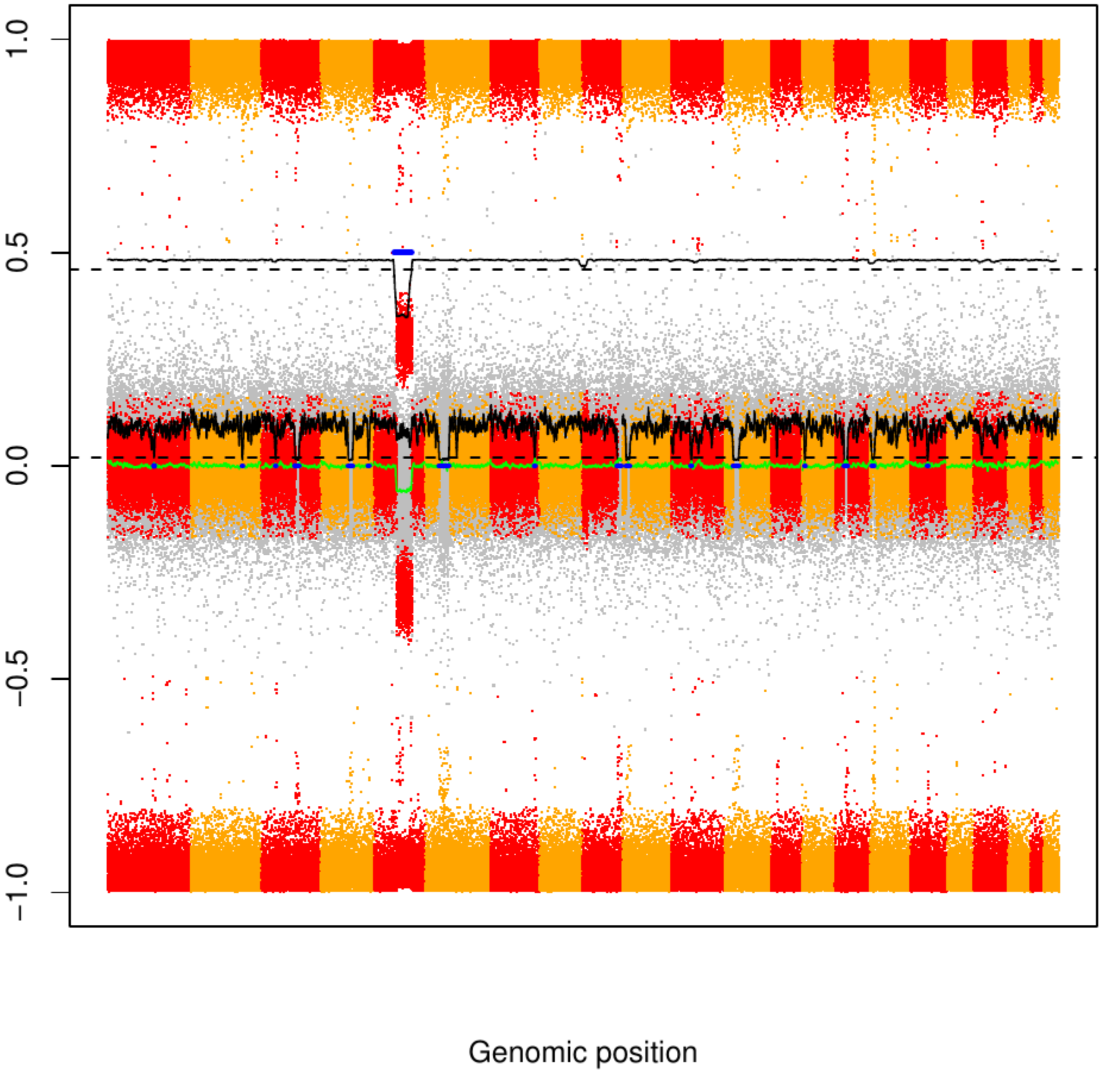

### CME104

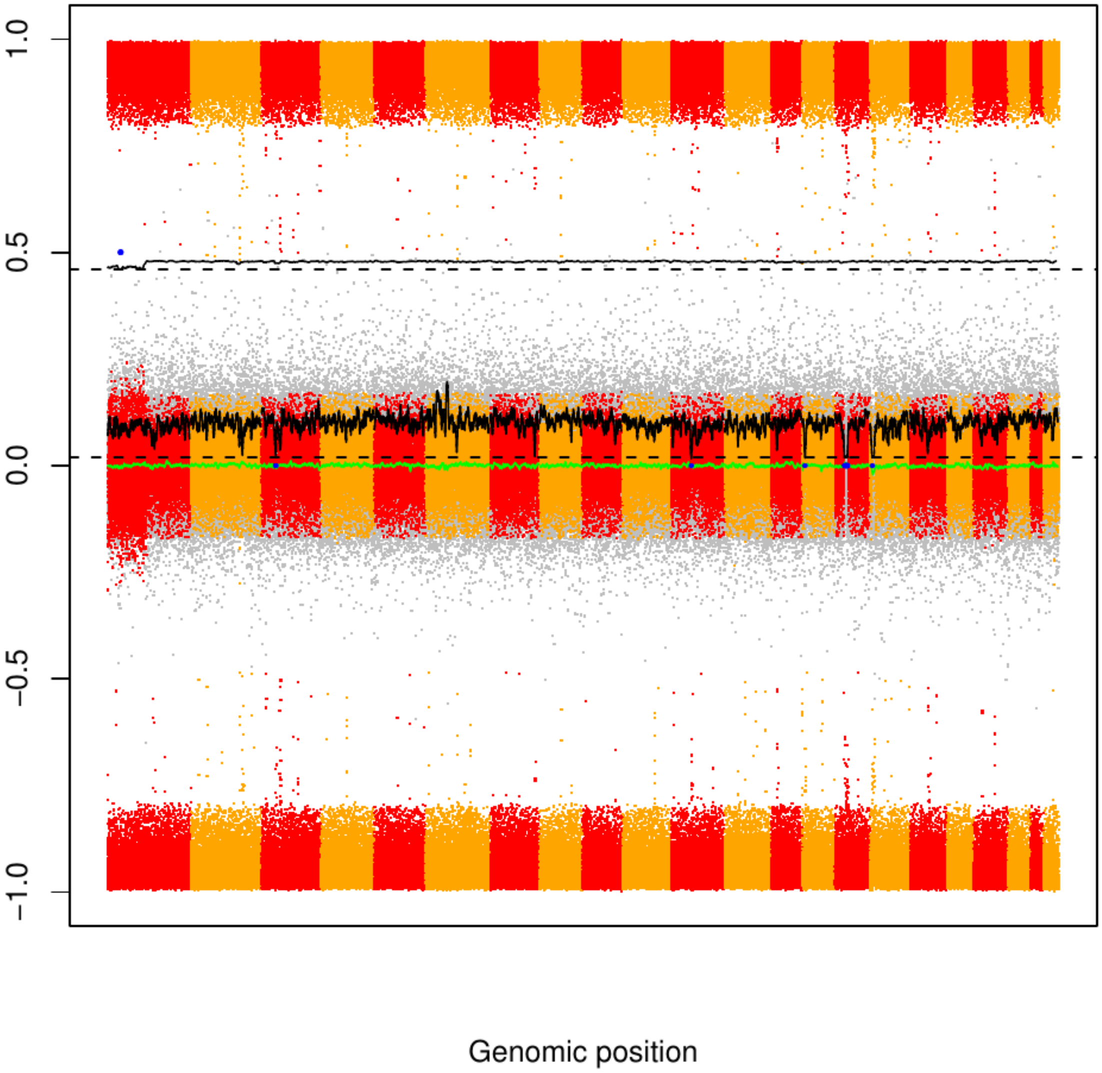

### CME105

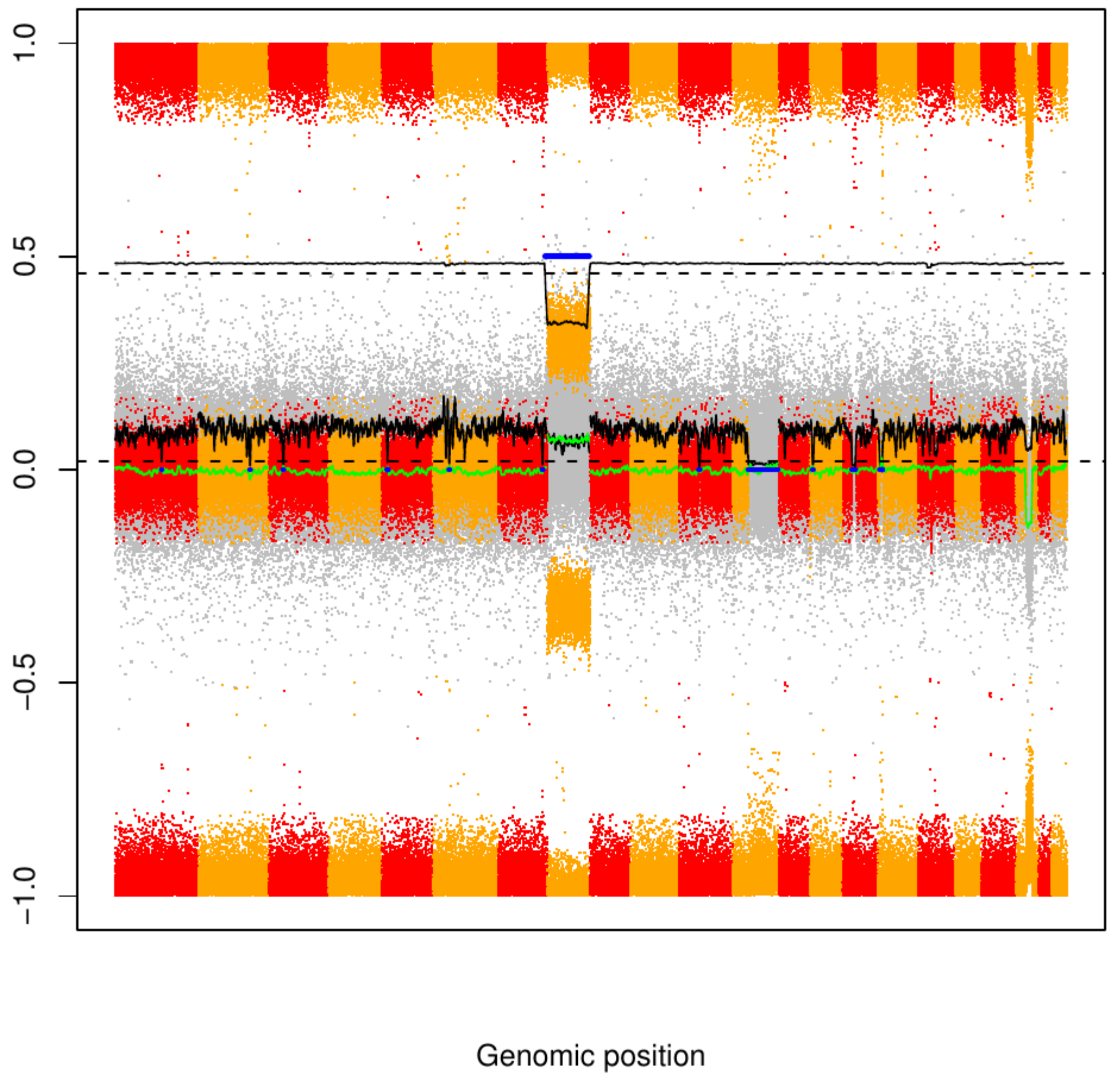

### CME106

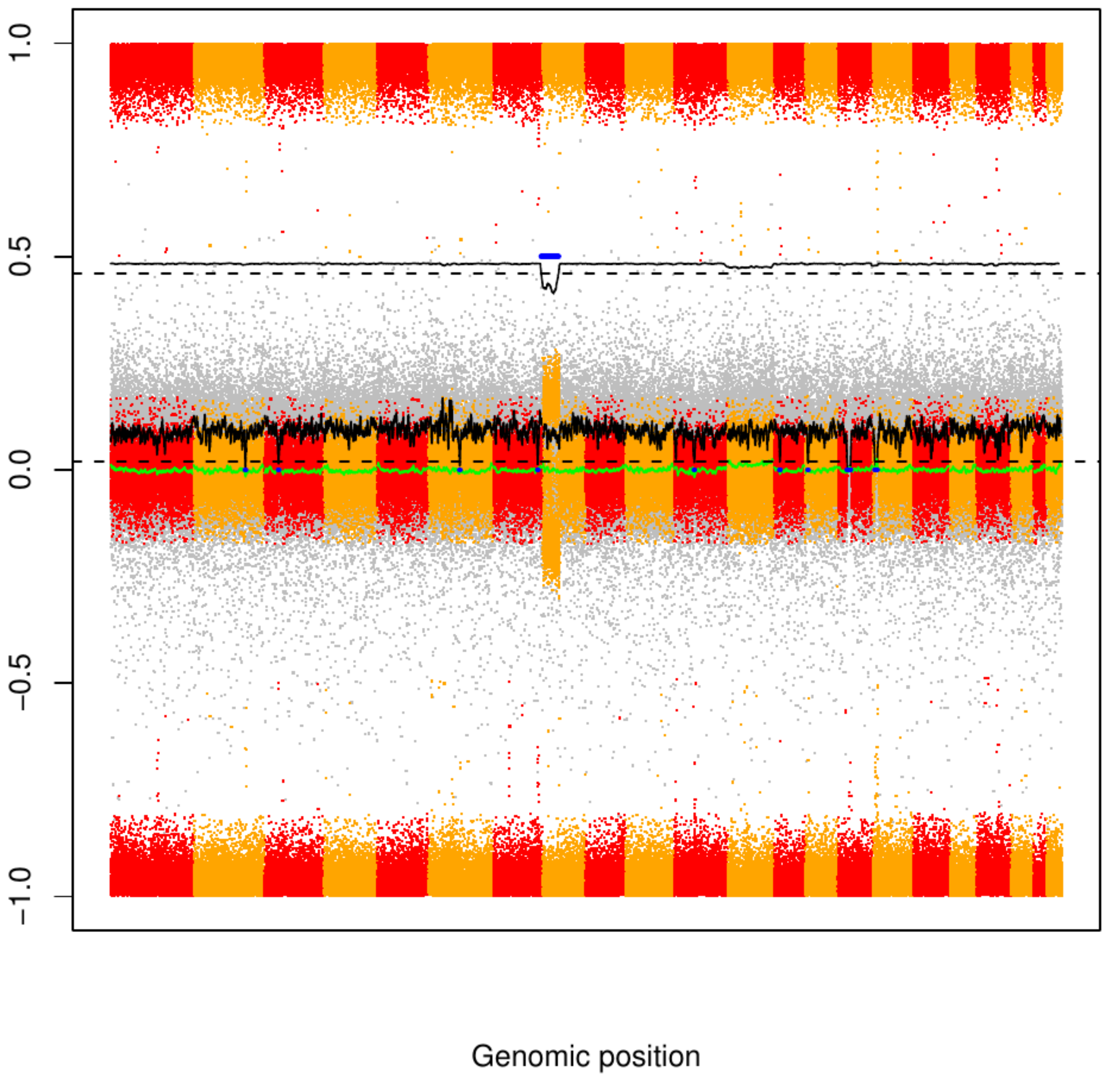

CME107

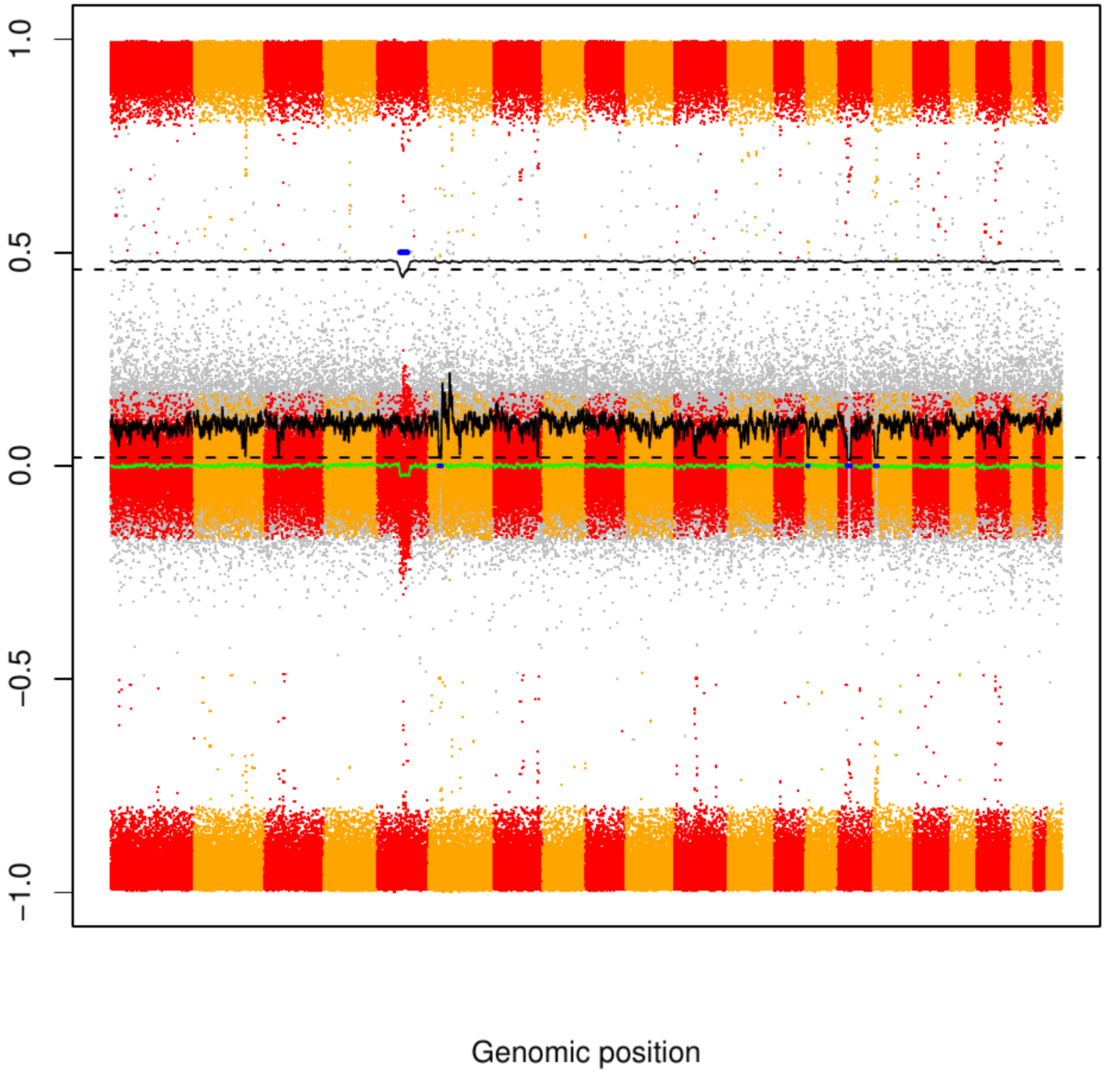

### CME108

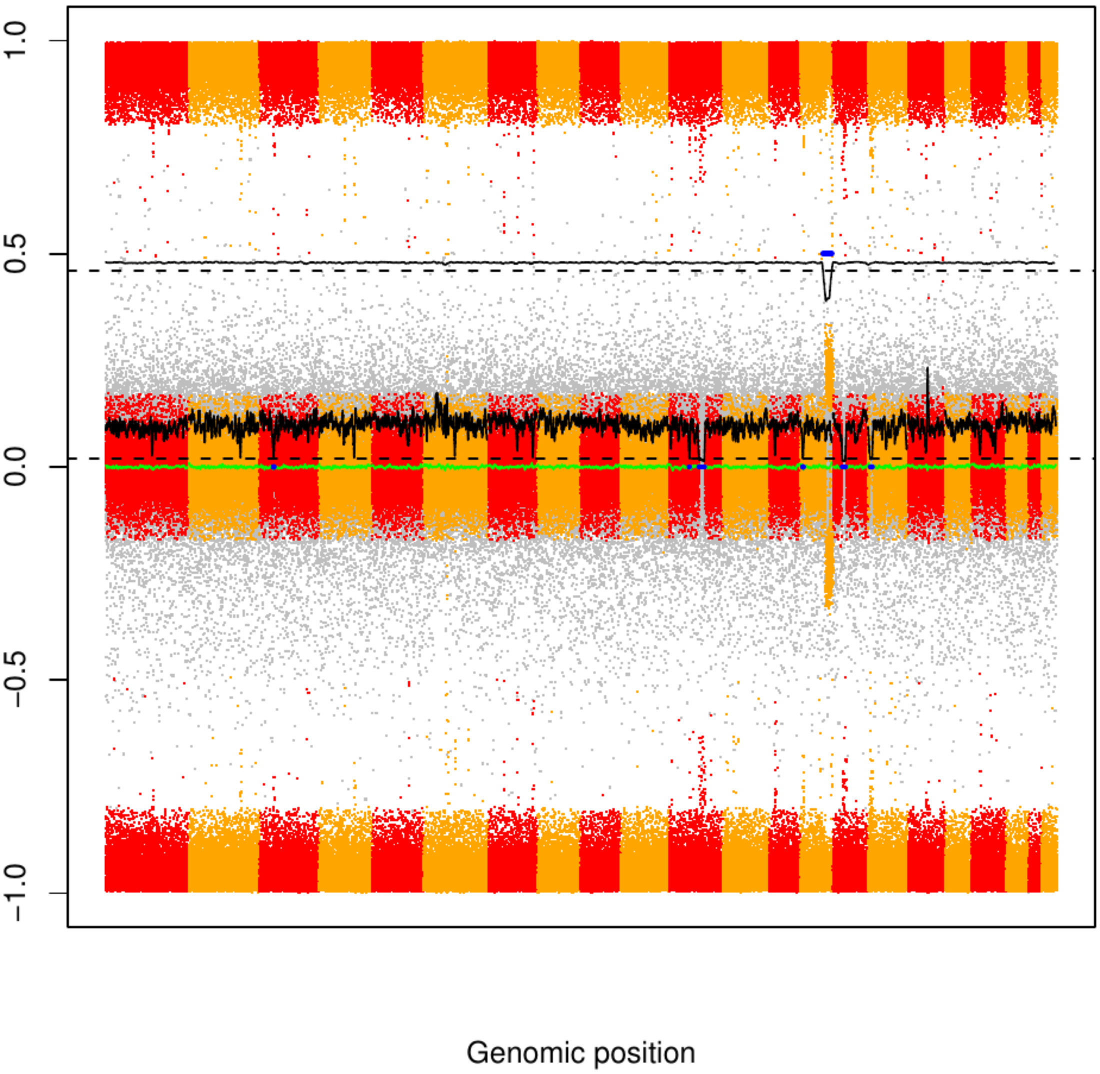

CME109

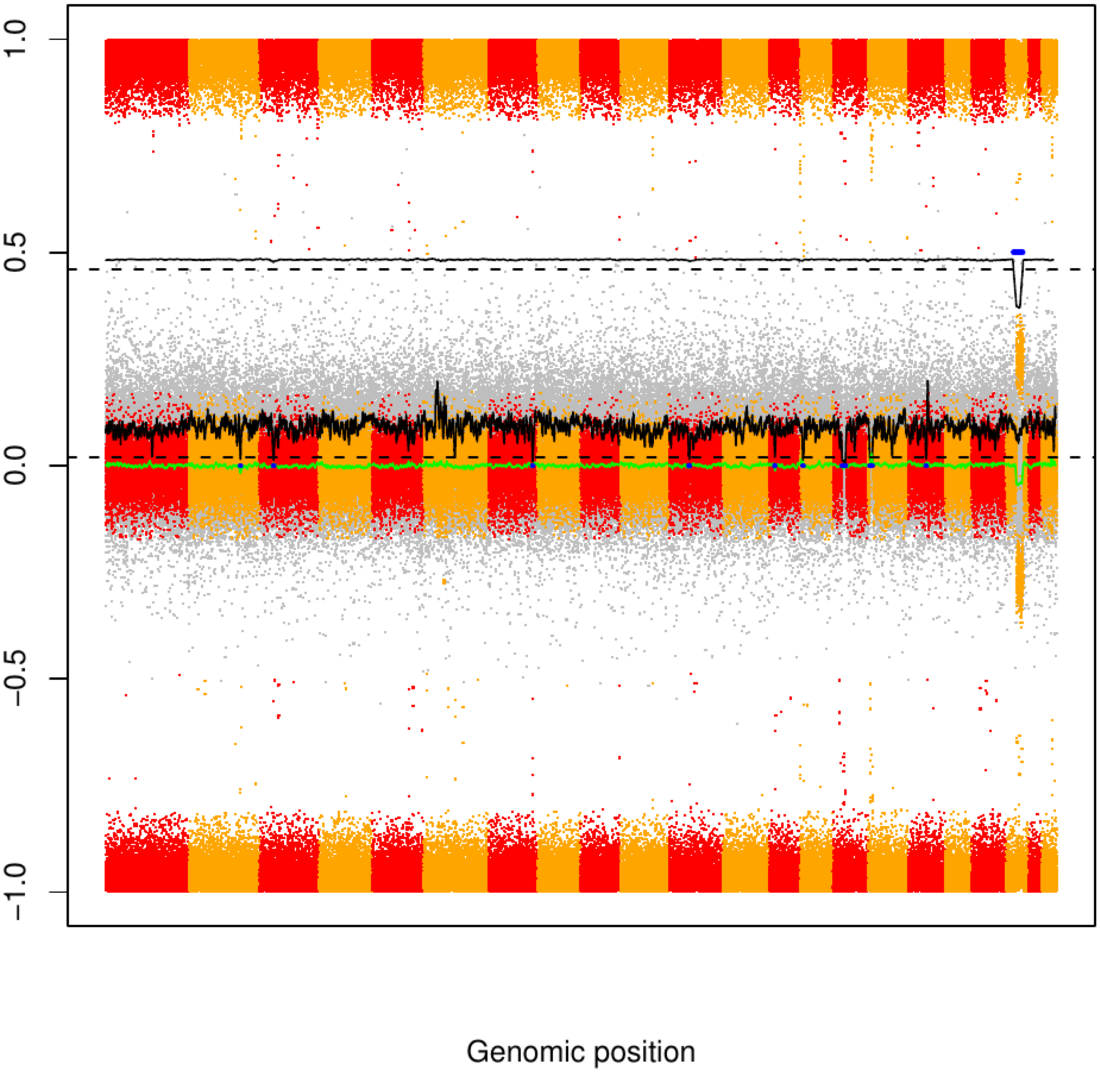

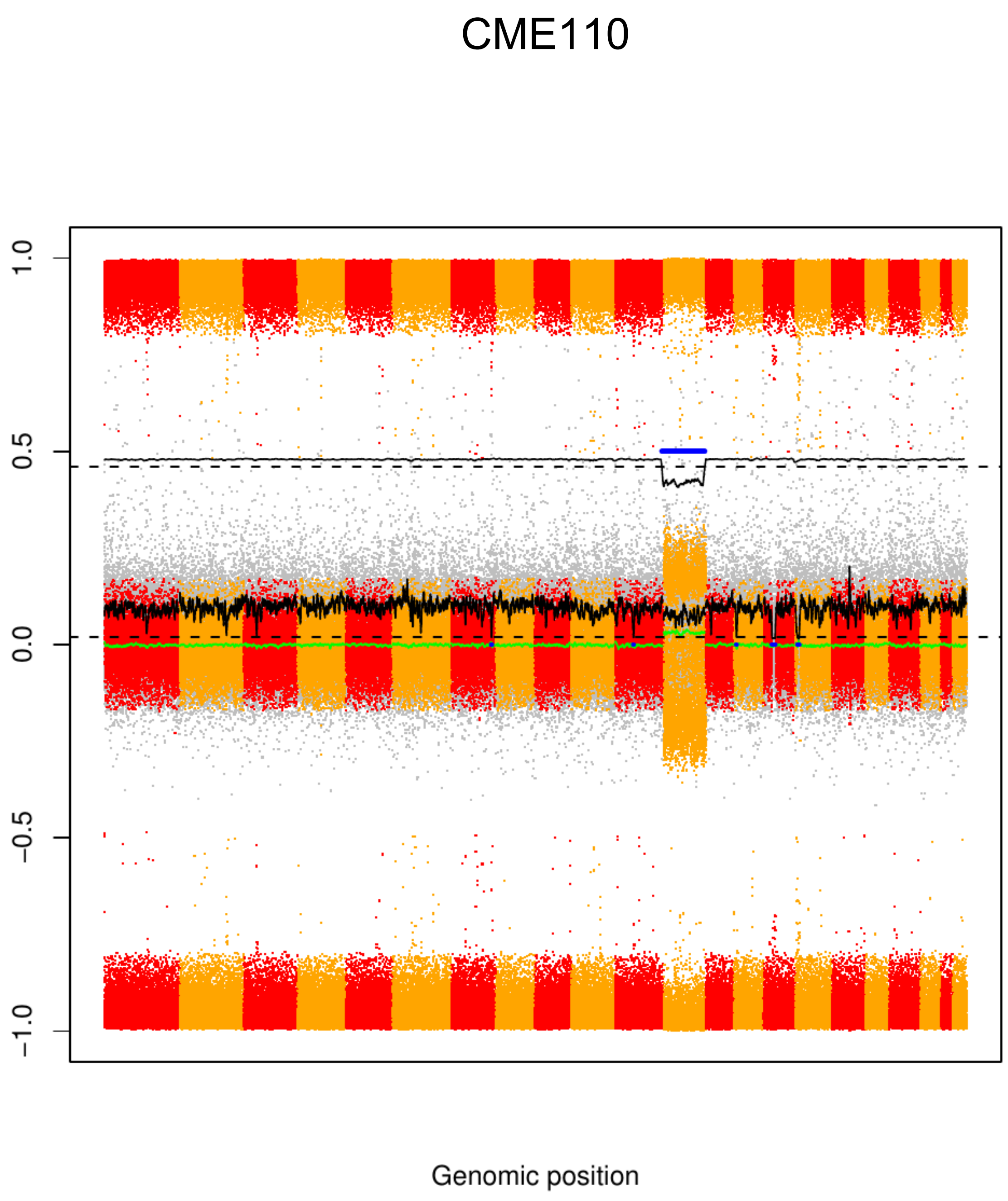
CME110
1.0
0.5
0.0
−0.5
−1.0
Genomic position

### CME111

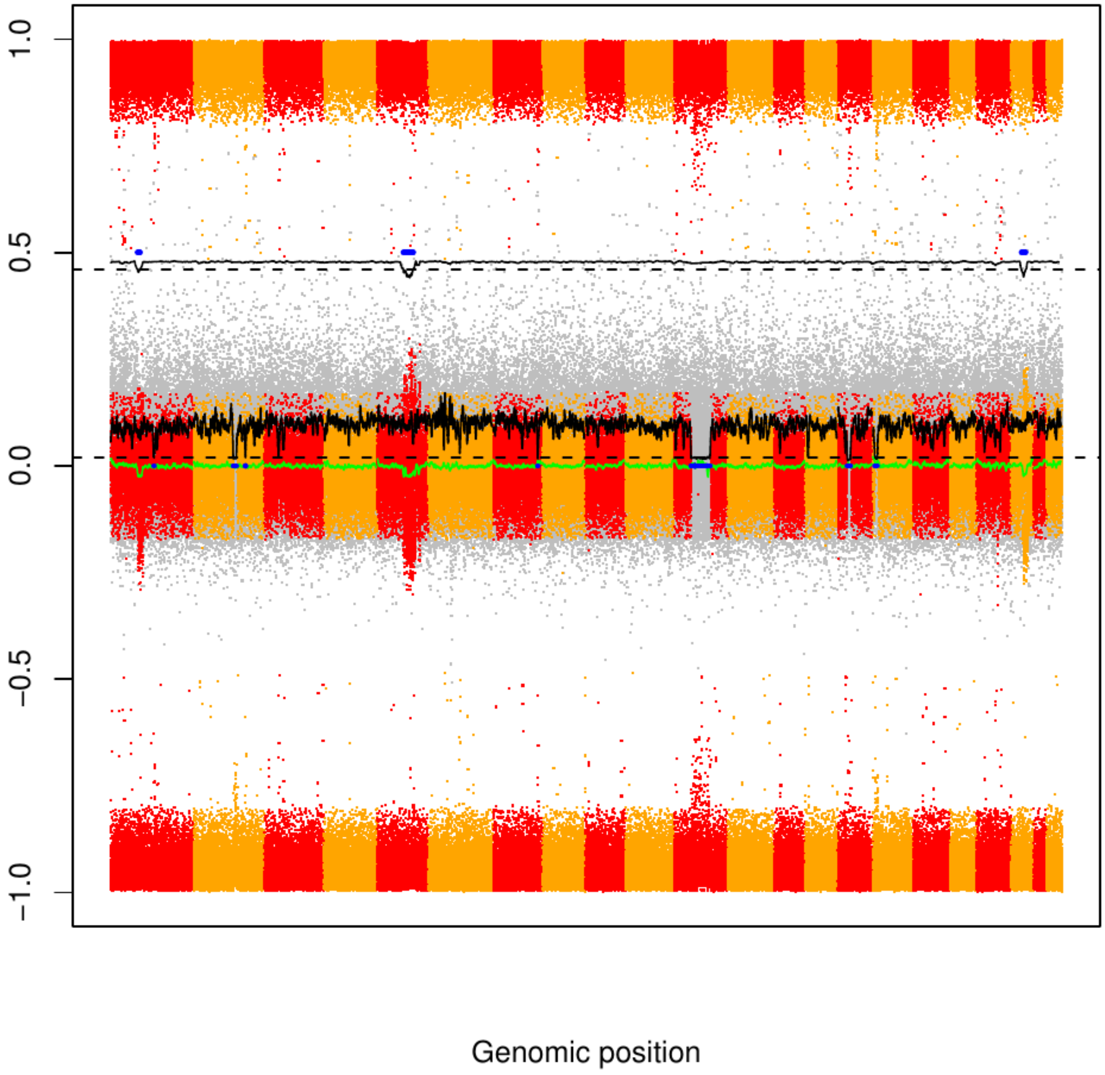

CME112

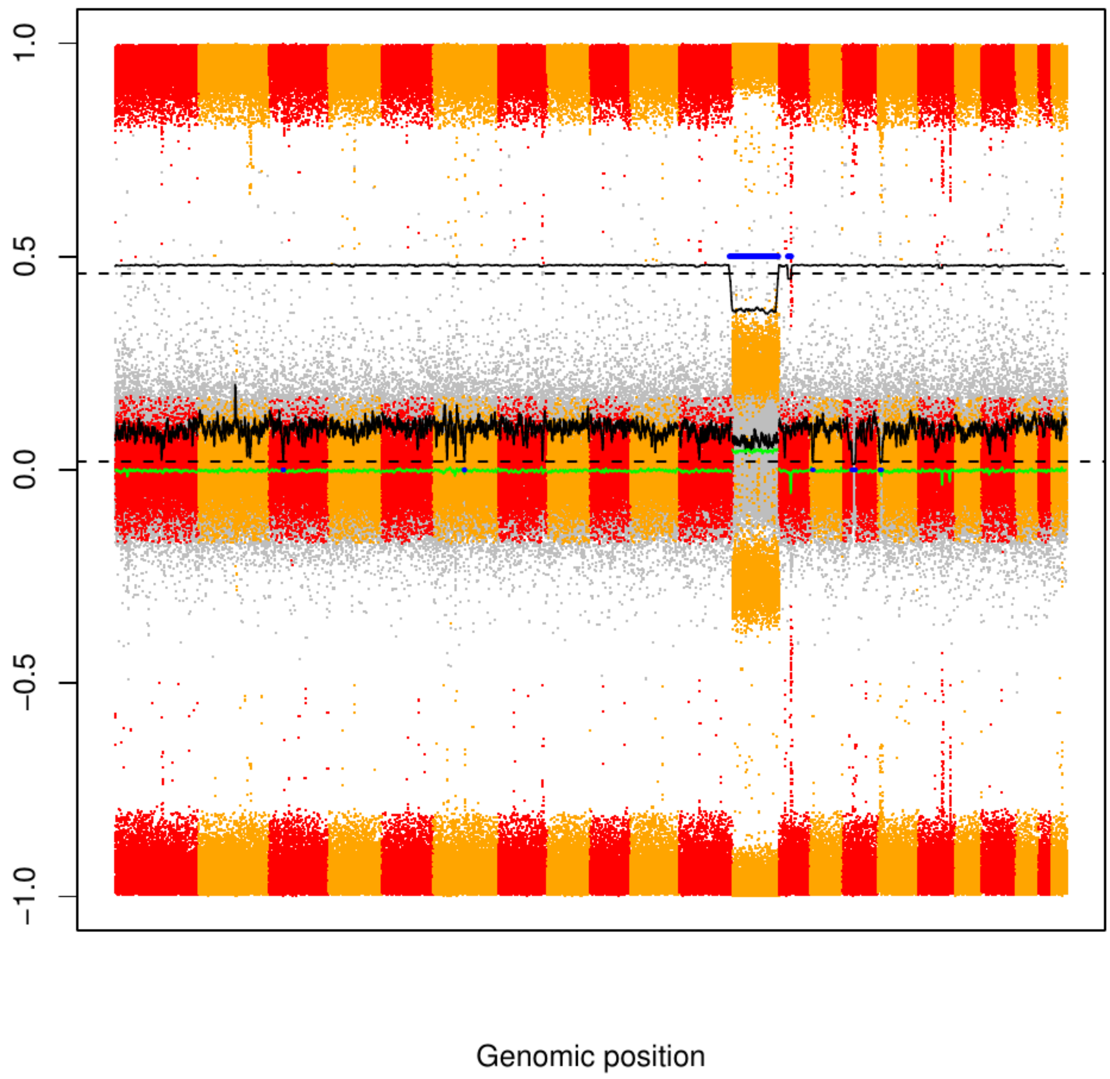

### CME113

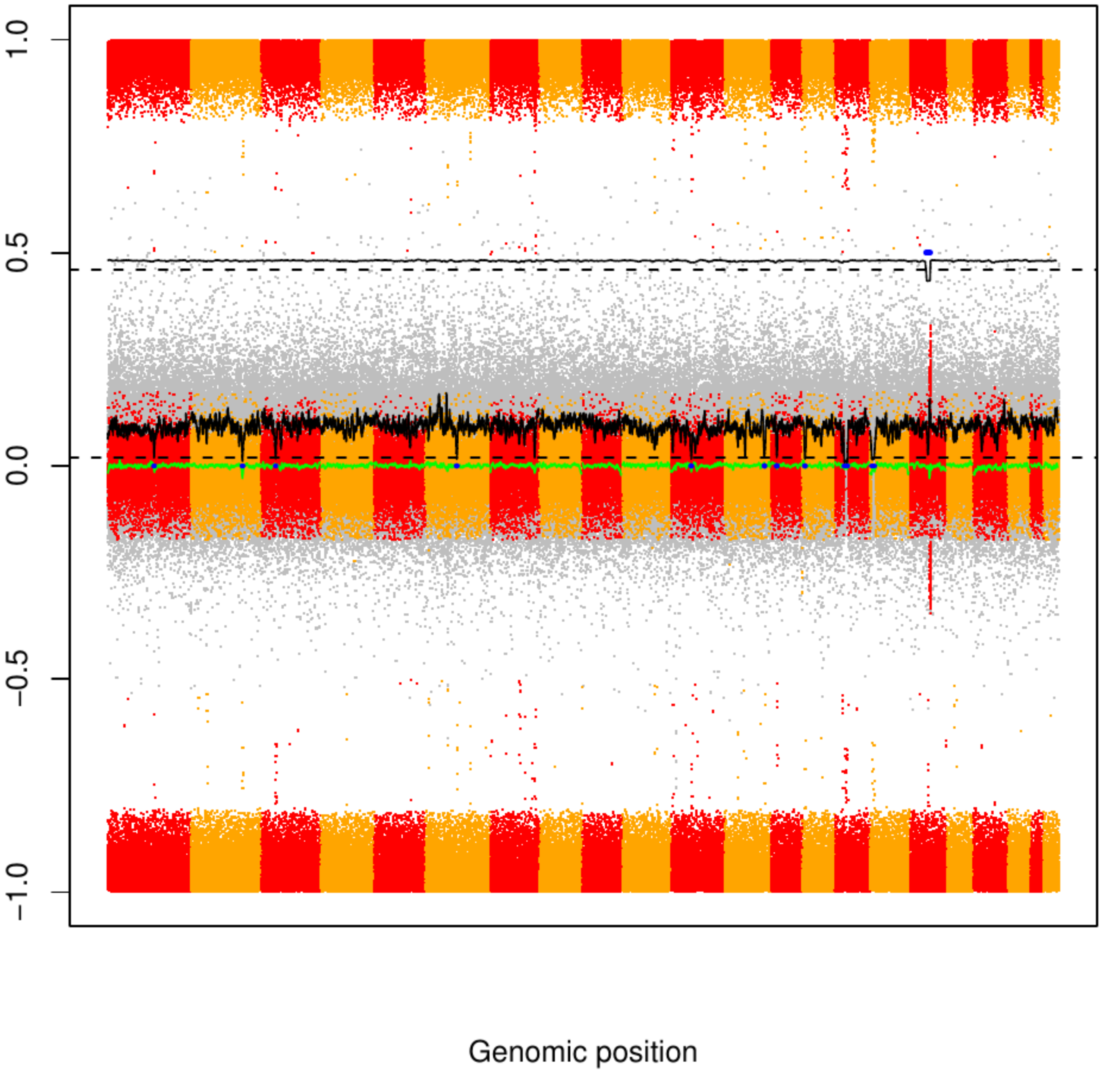

CME114

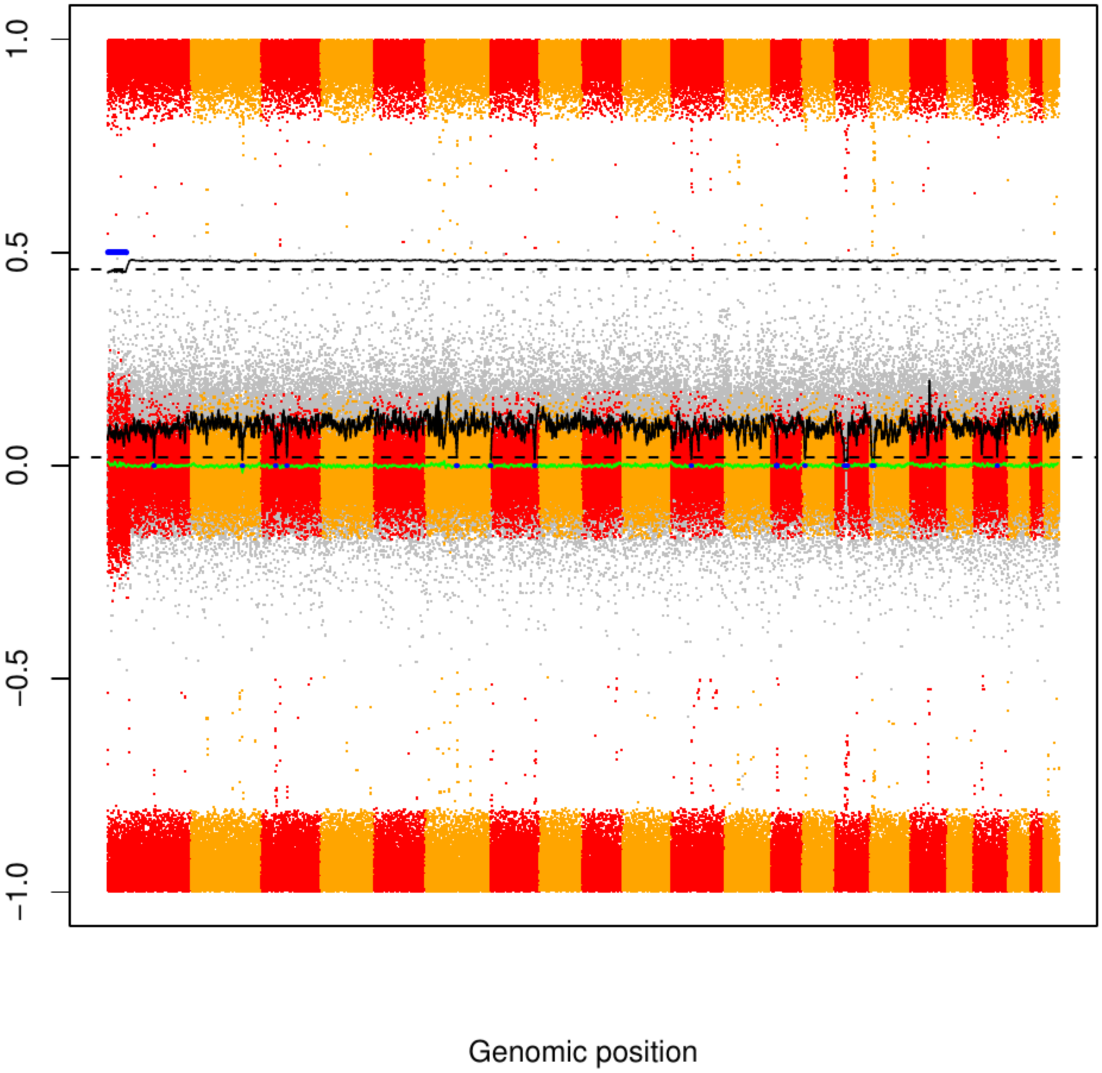

### CME115

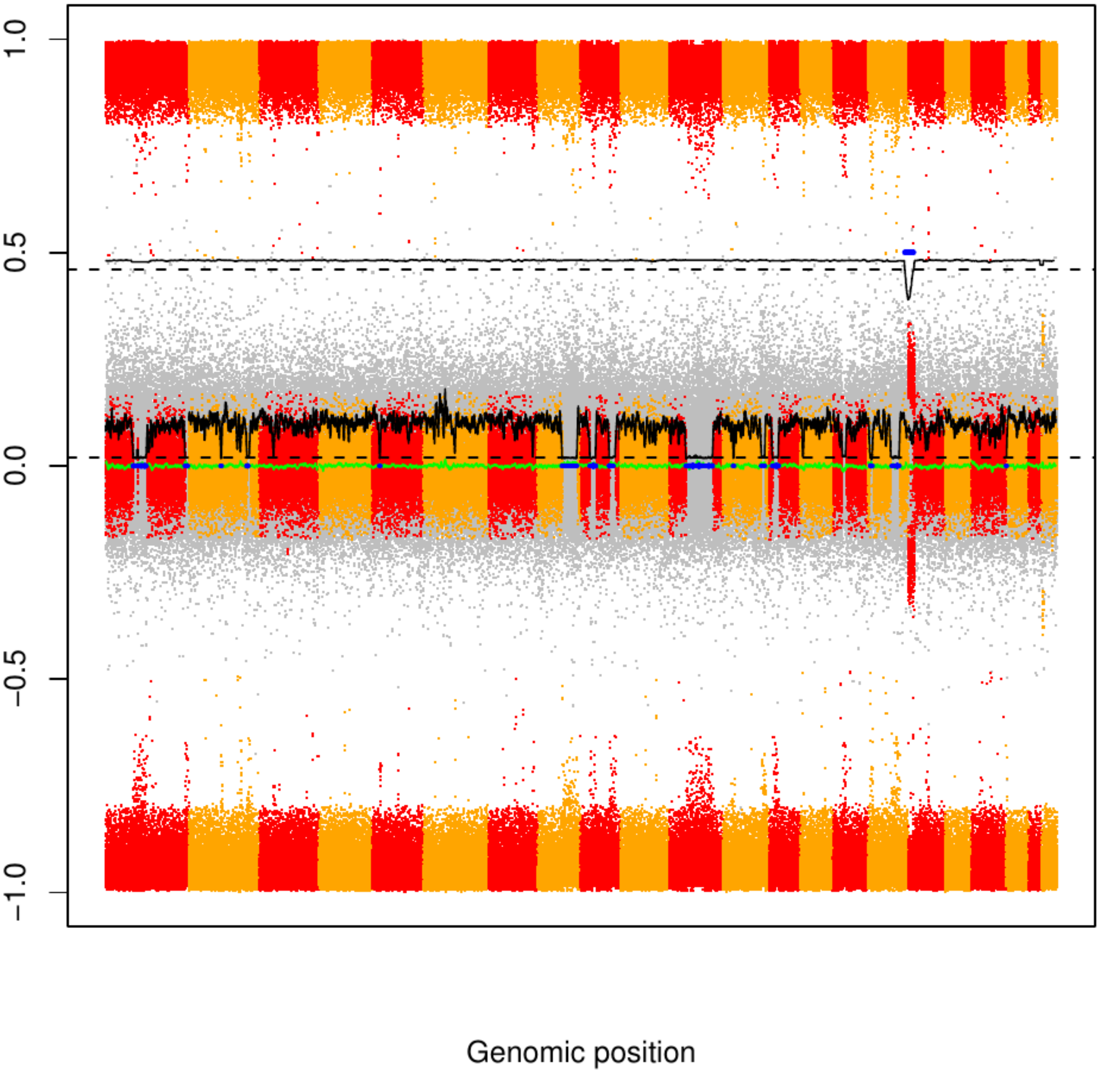

### CME116

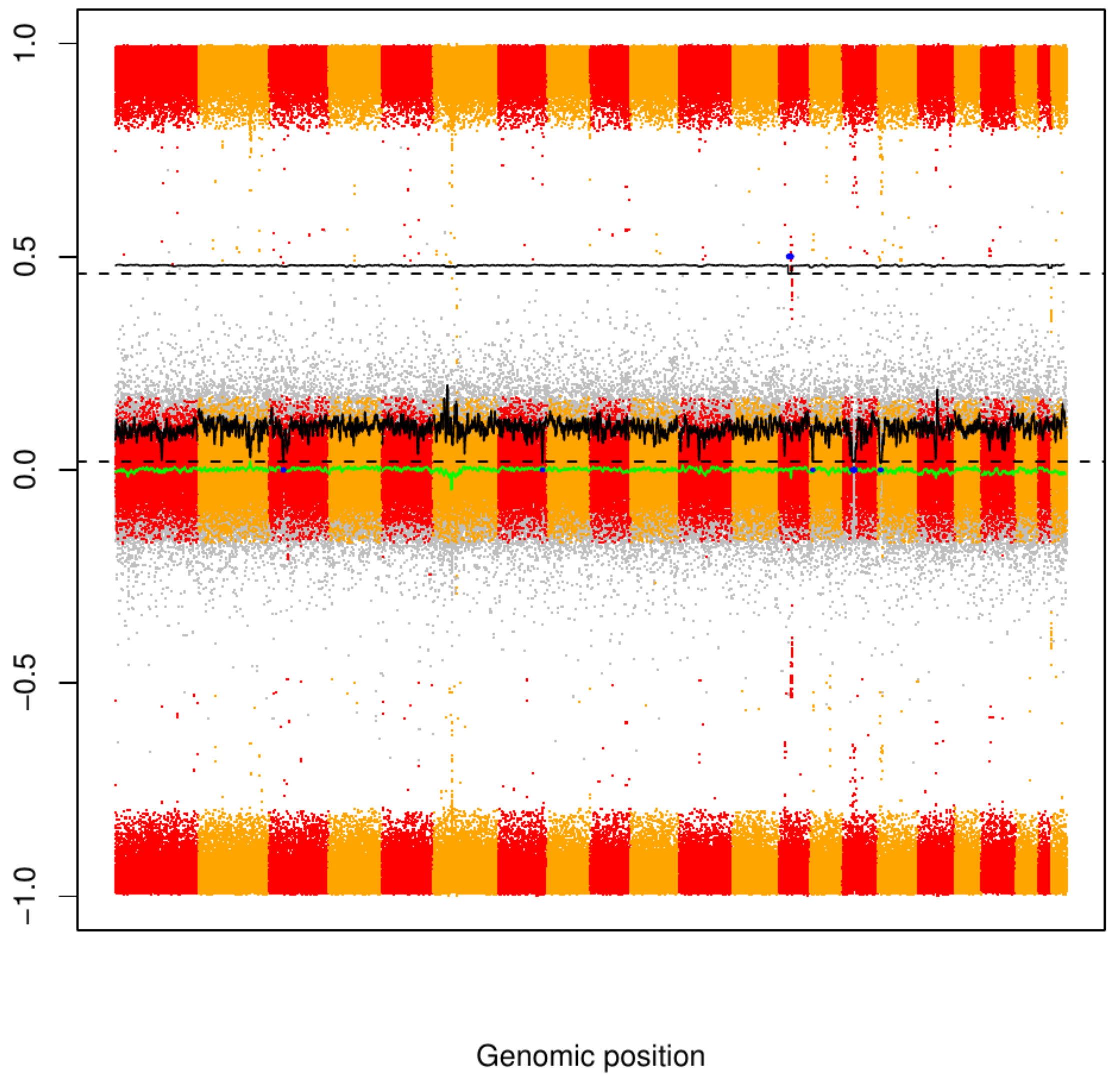

### CME117

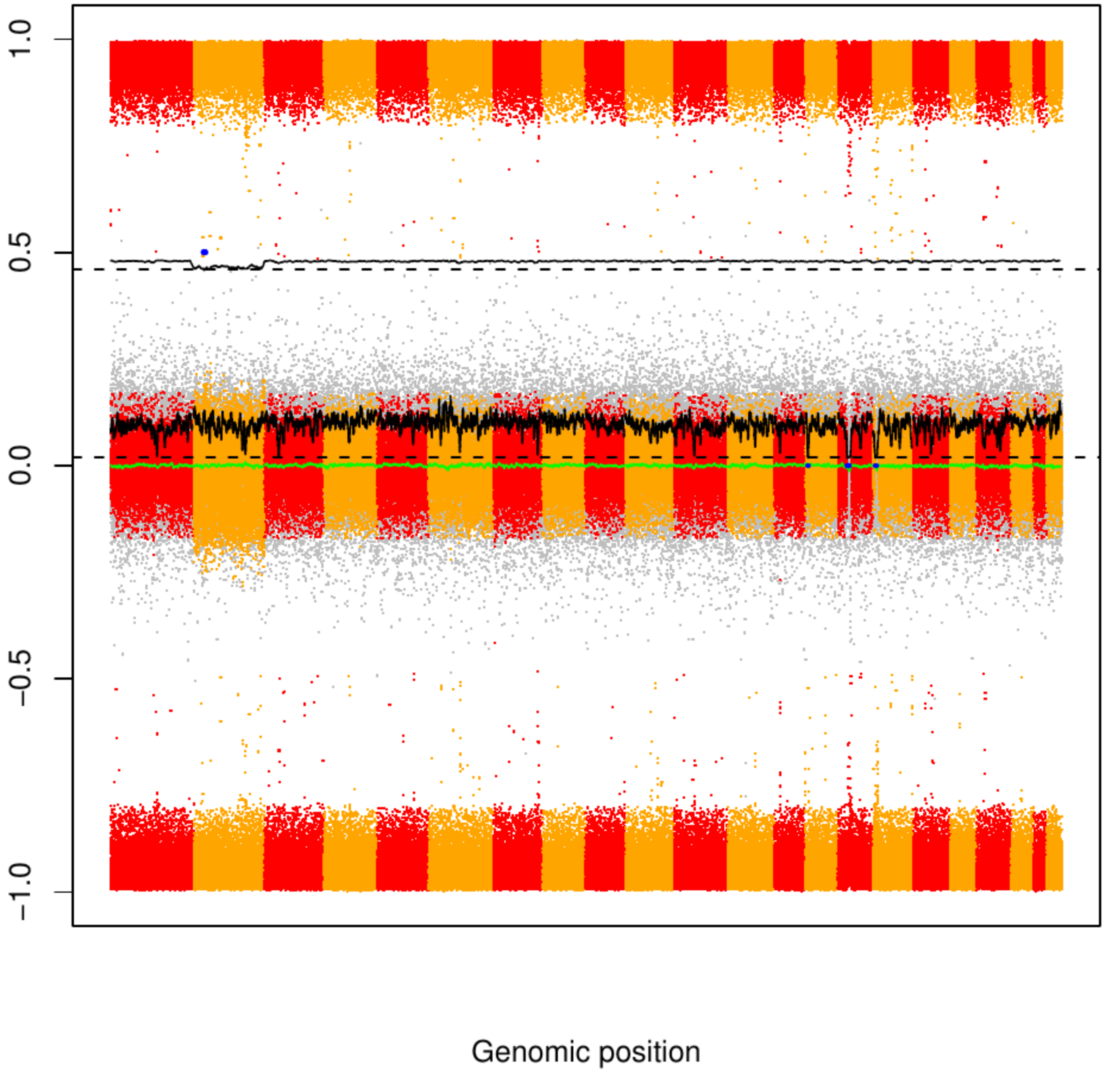

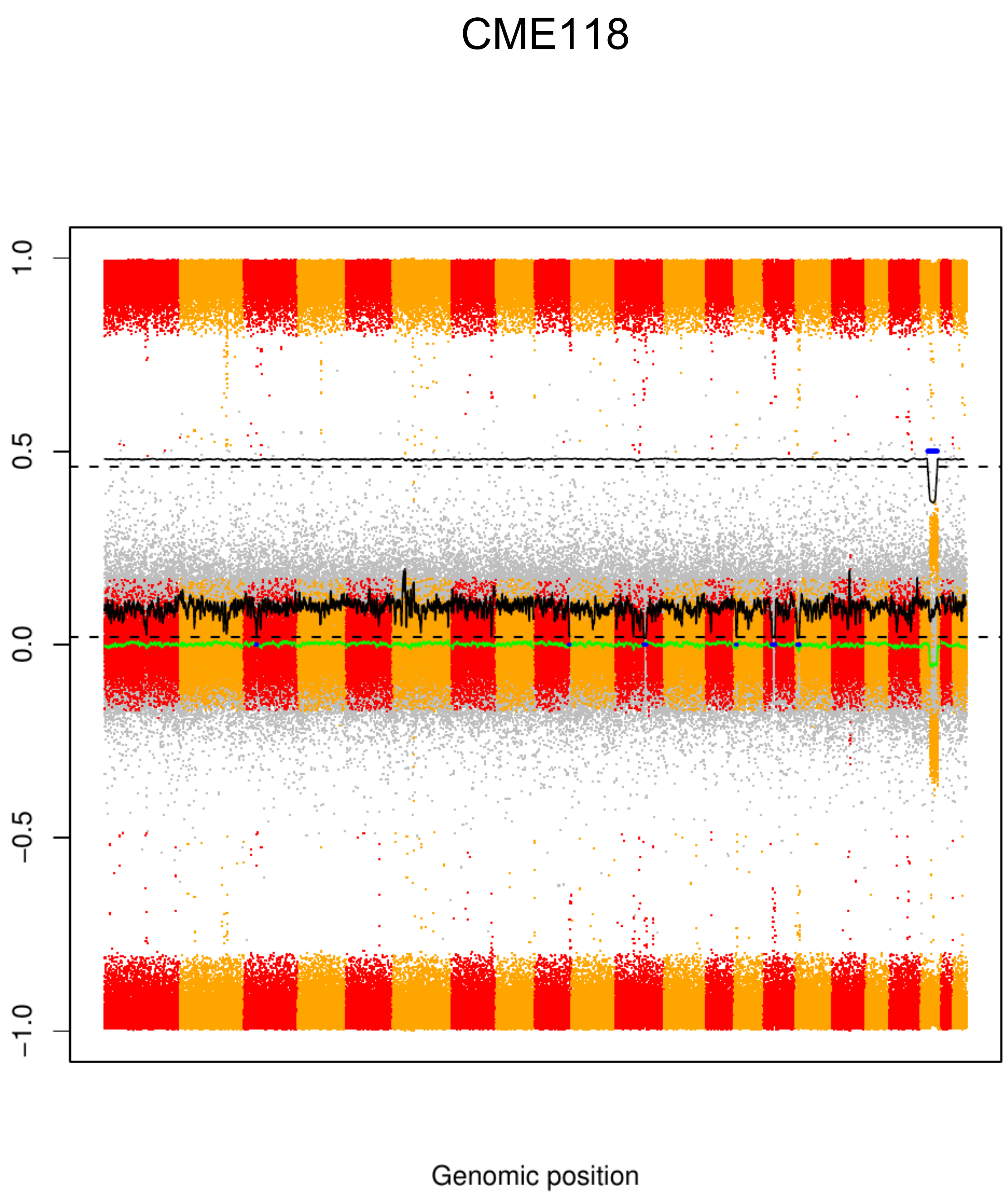
CME118
1.0
0.5
0.0
−0.5
−1.0
Genomic position

### CME119

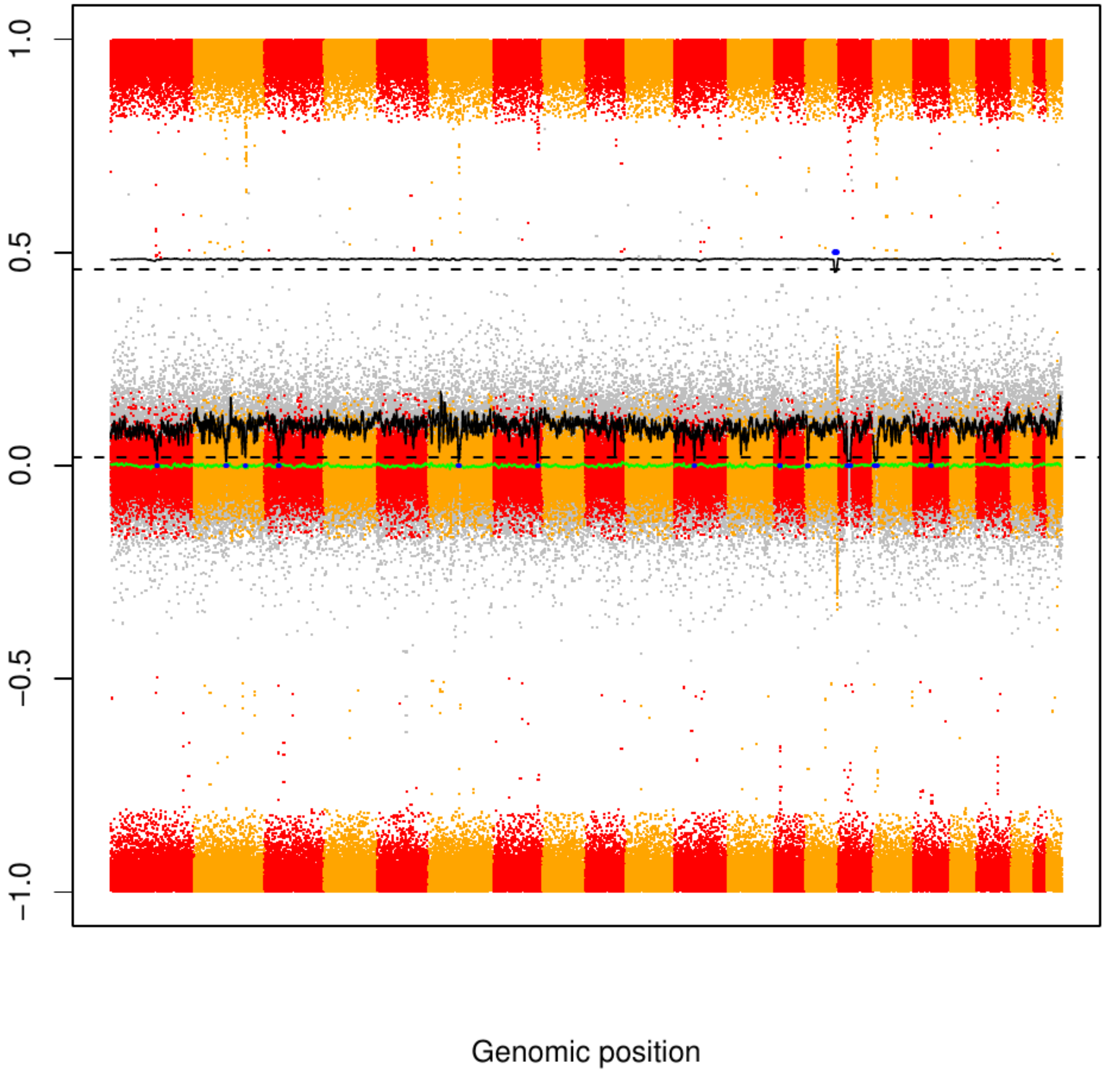

### CME120

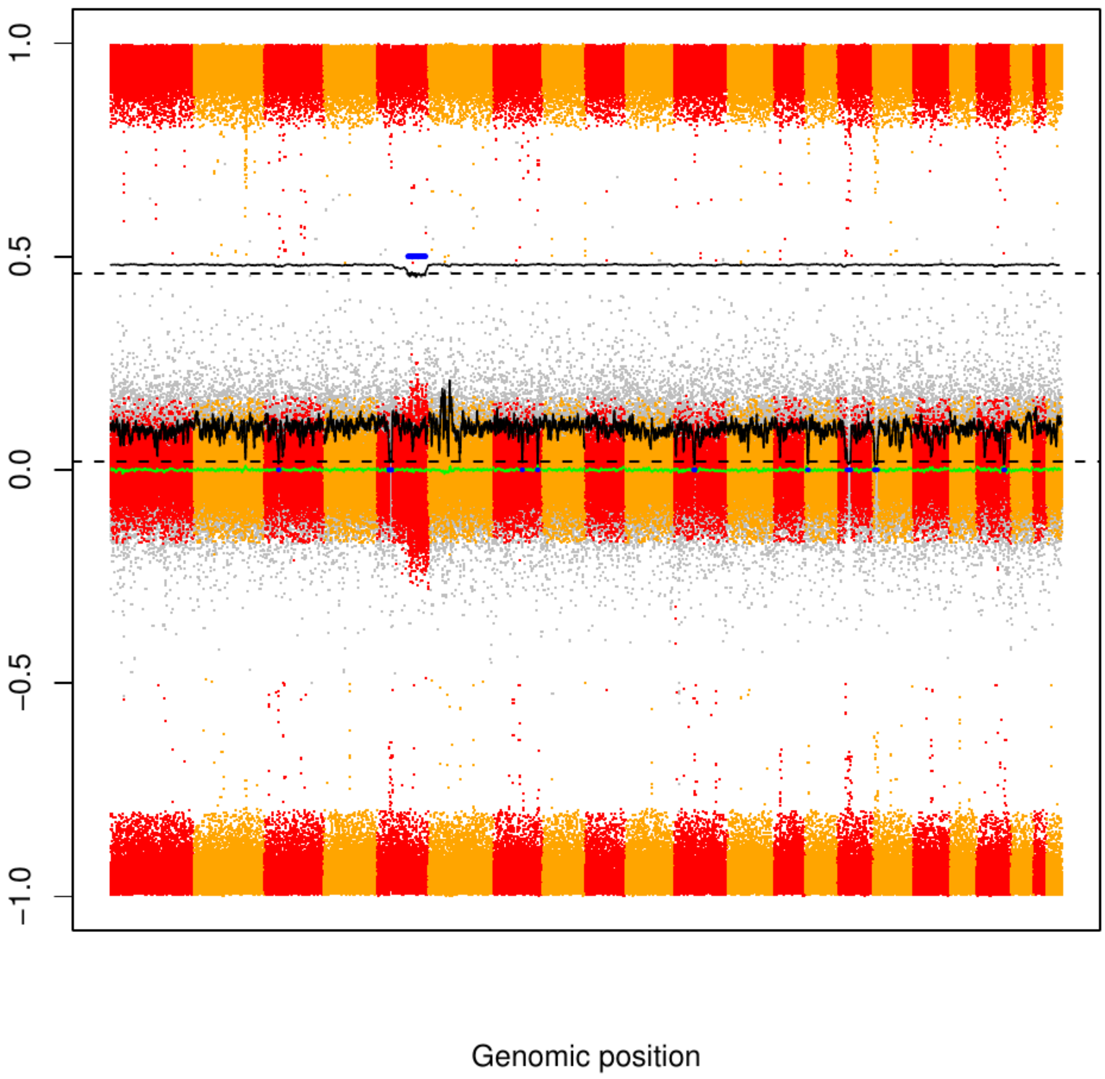

CME121

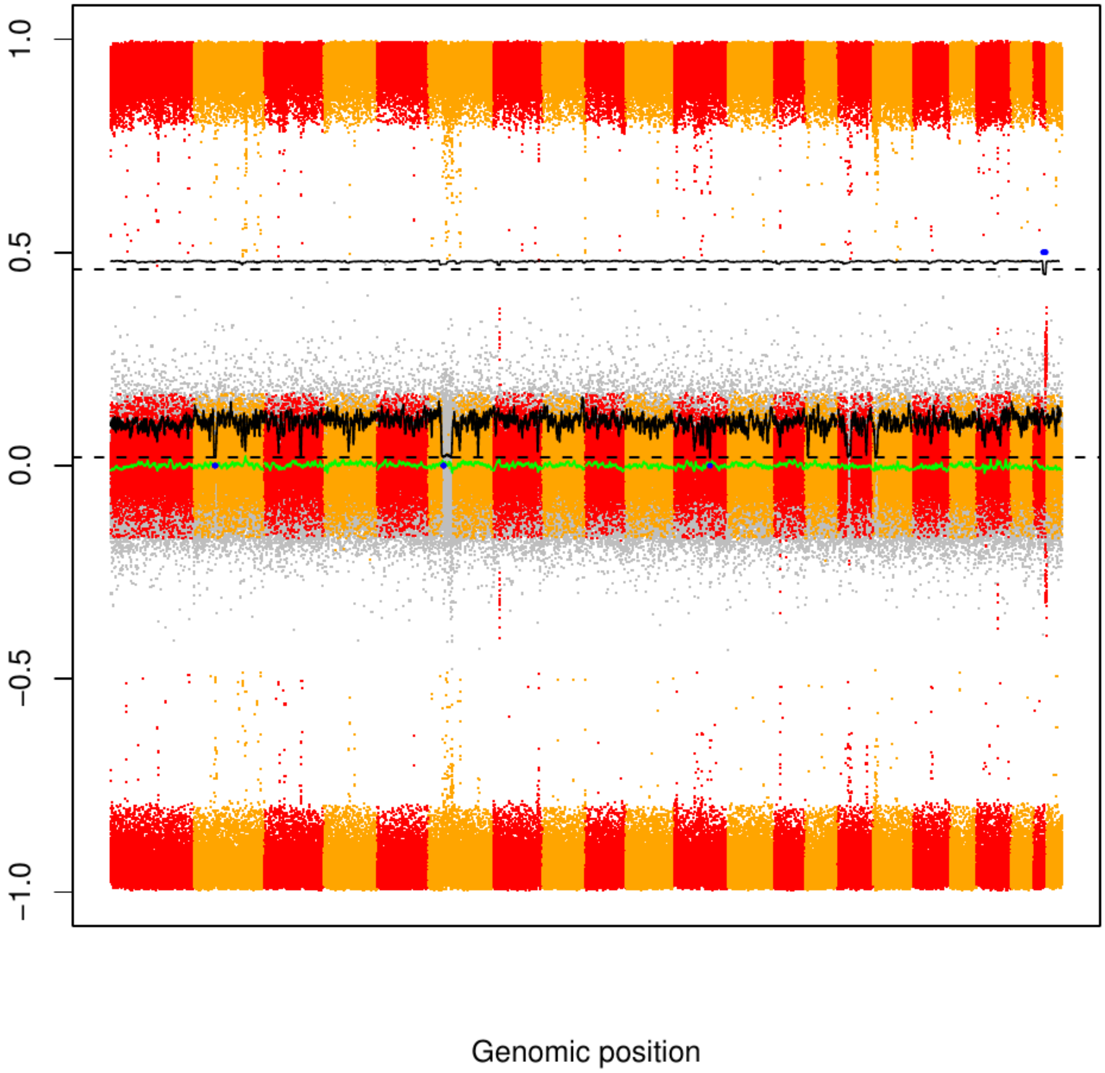

### CME122

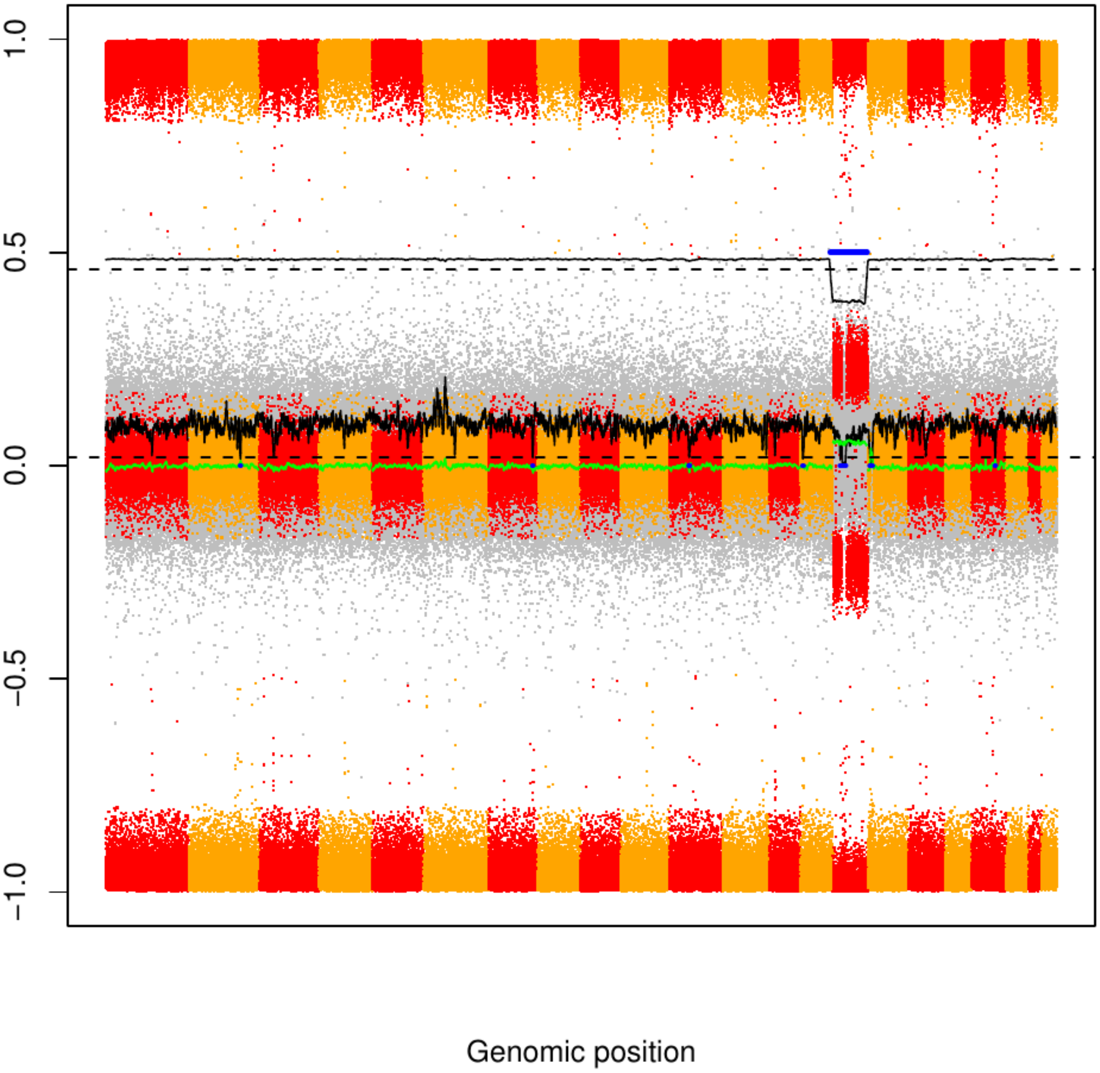

### CME123

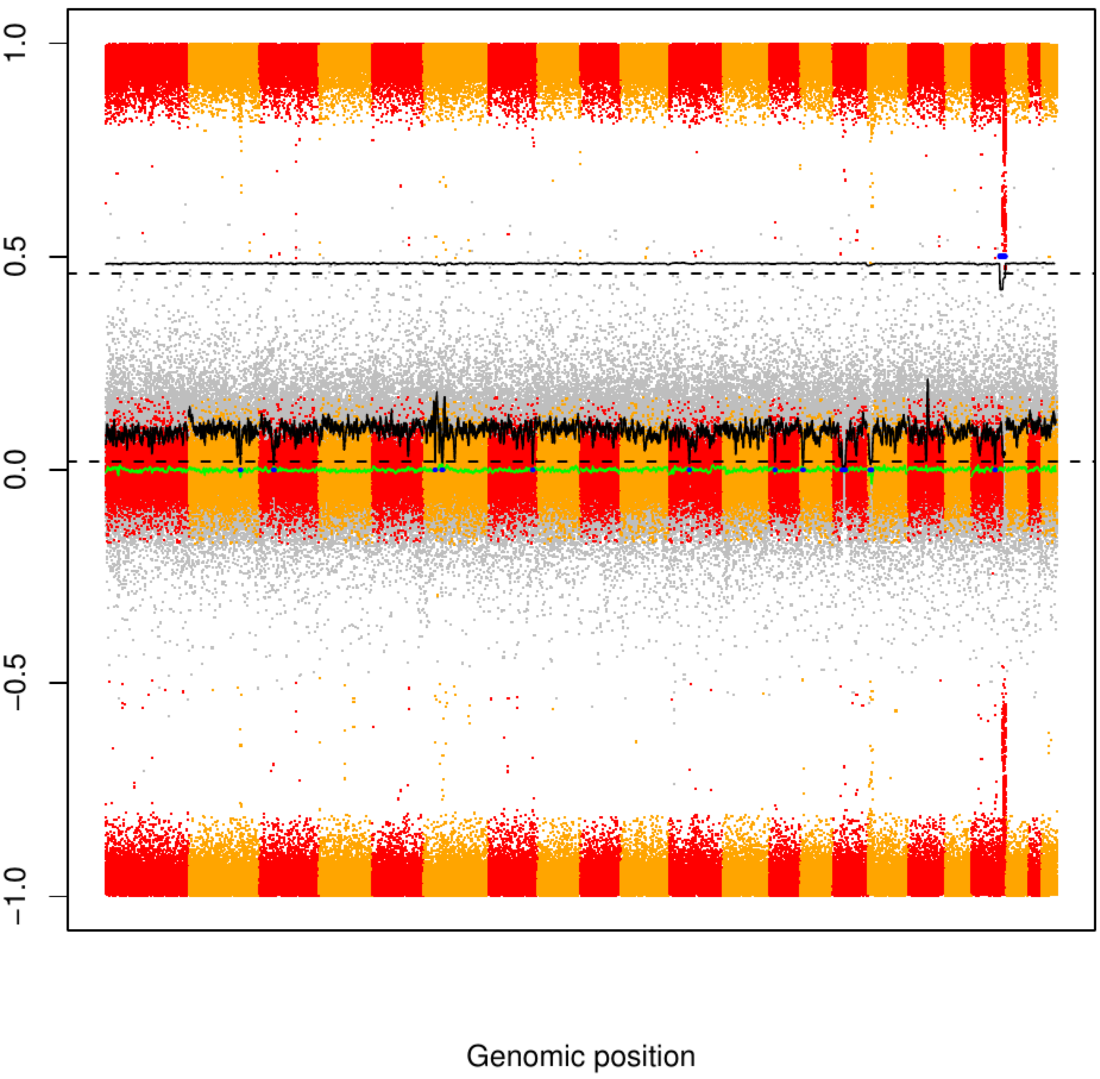

CME124

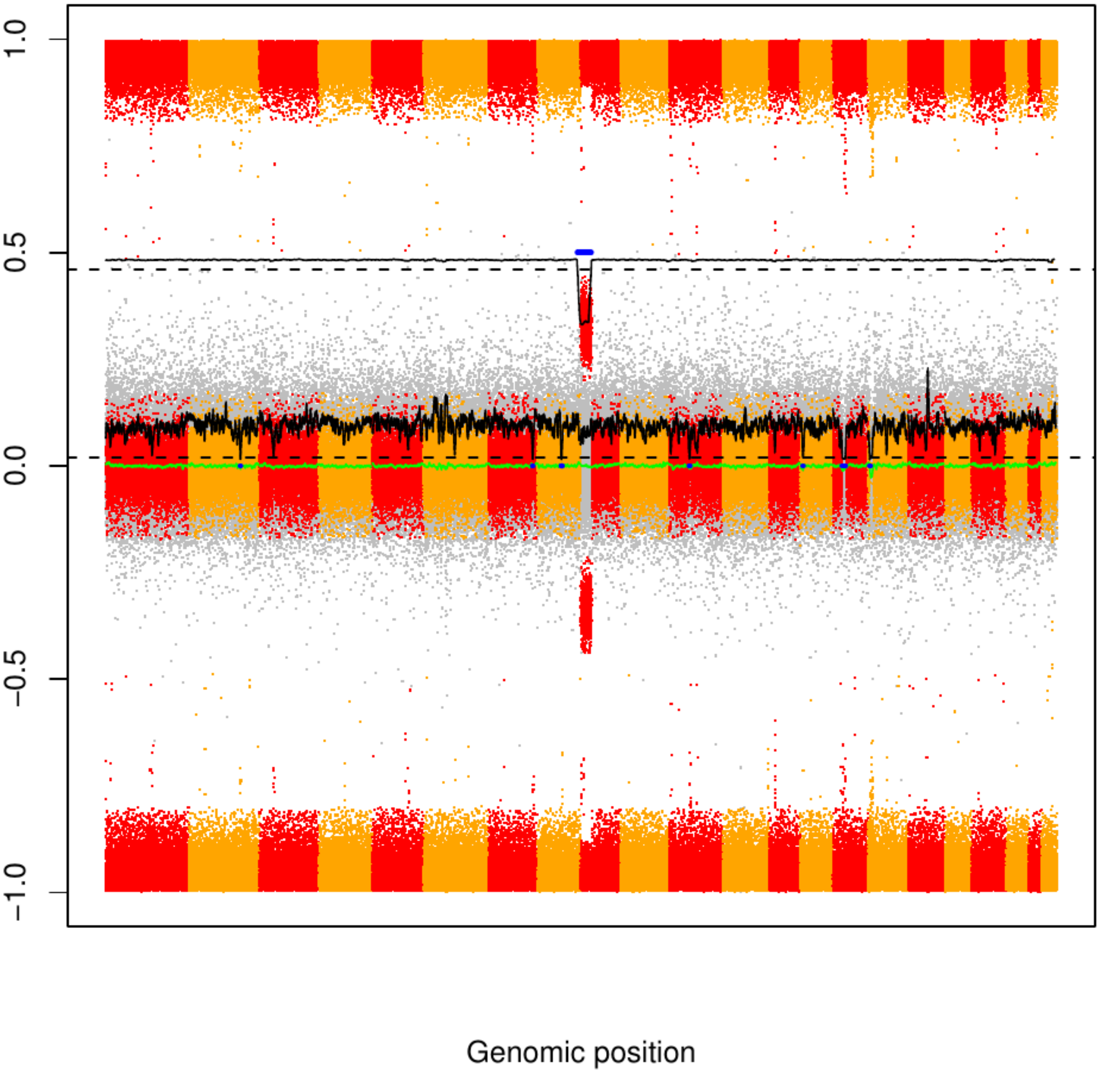

### CME125

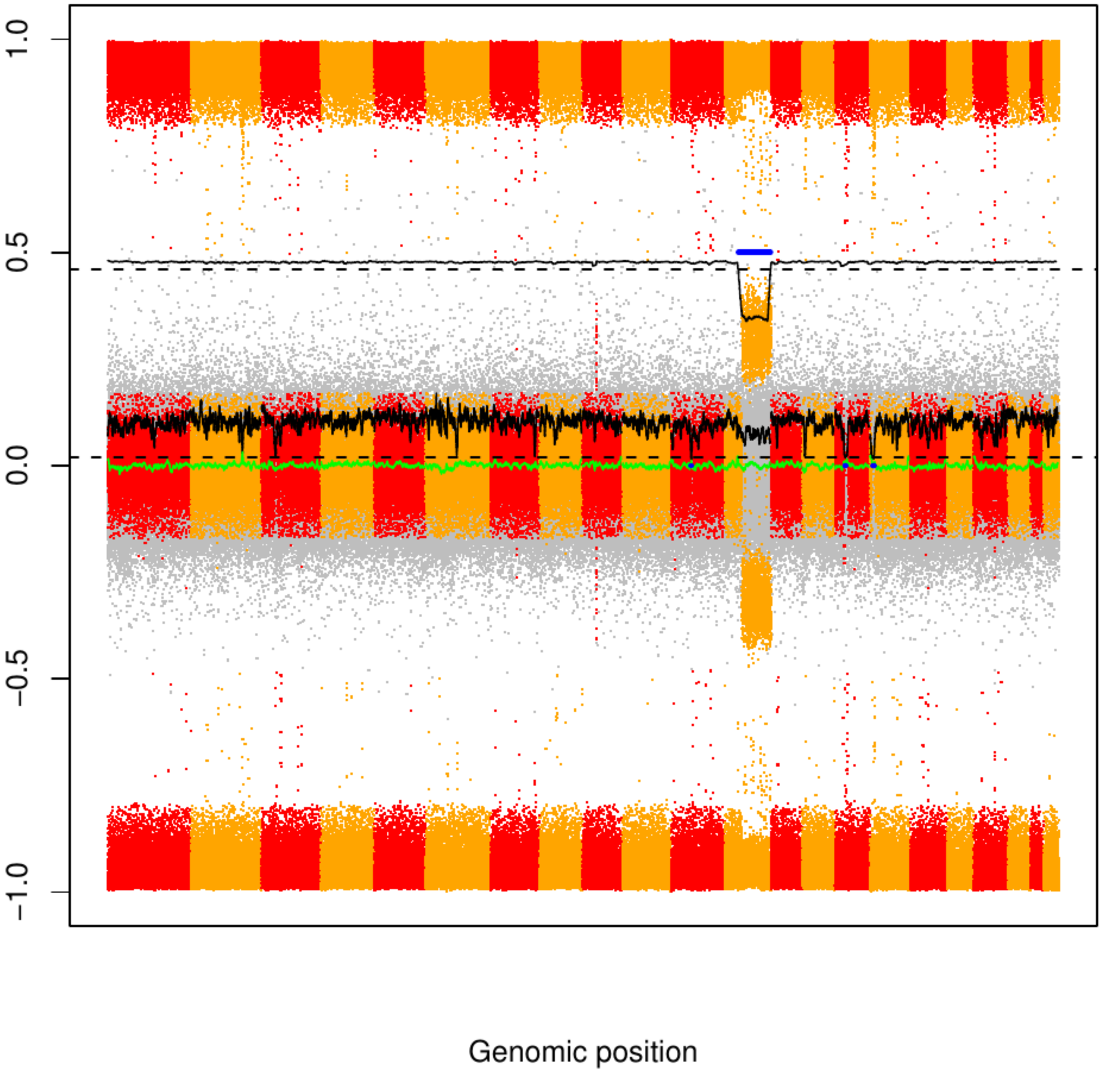

CME126

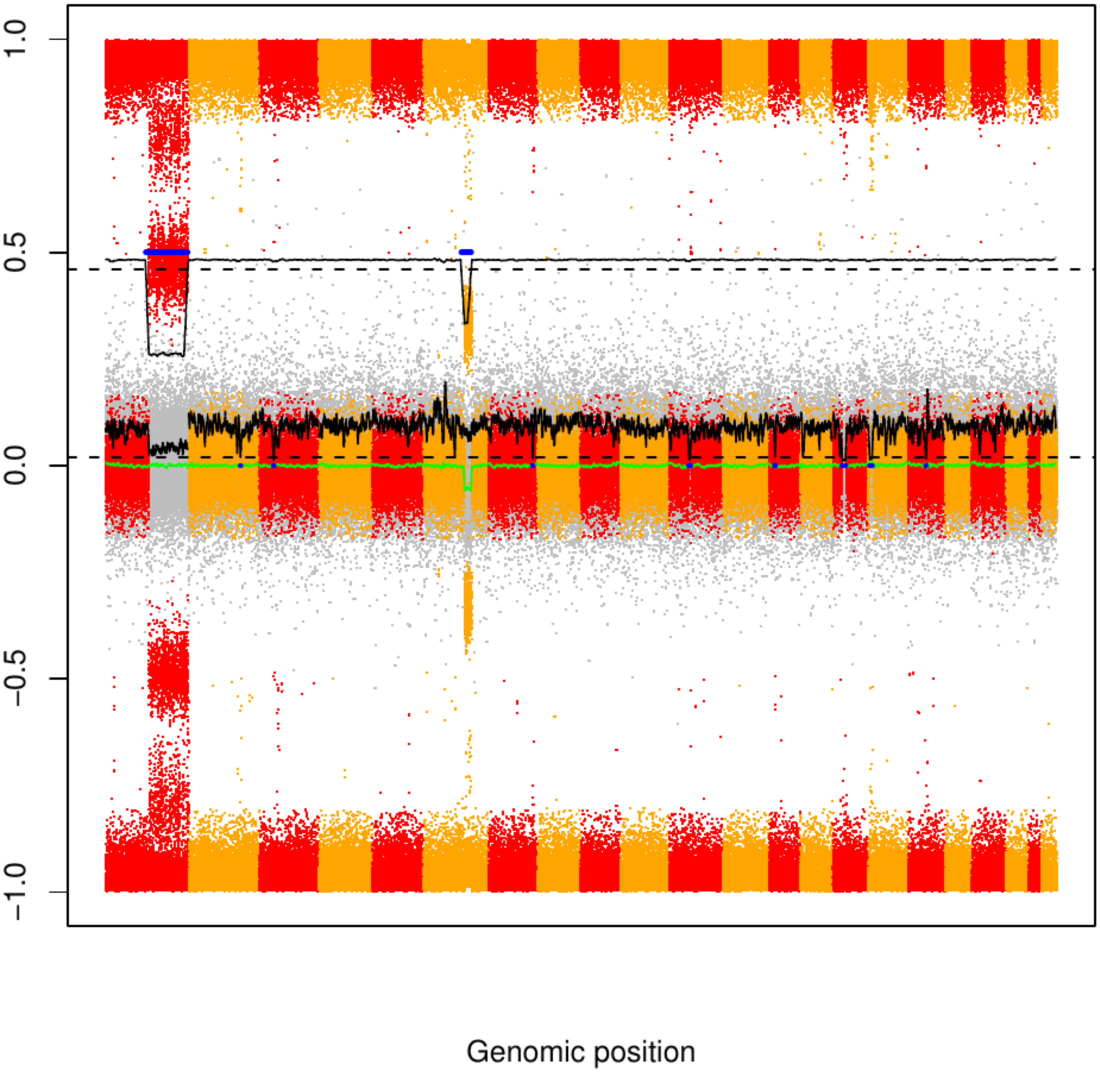

CME127

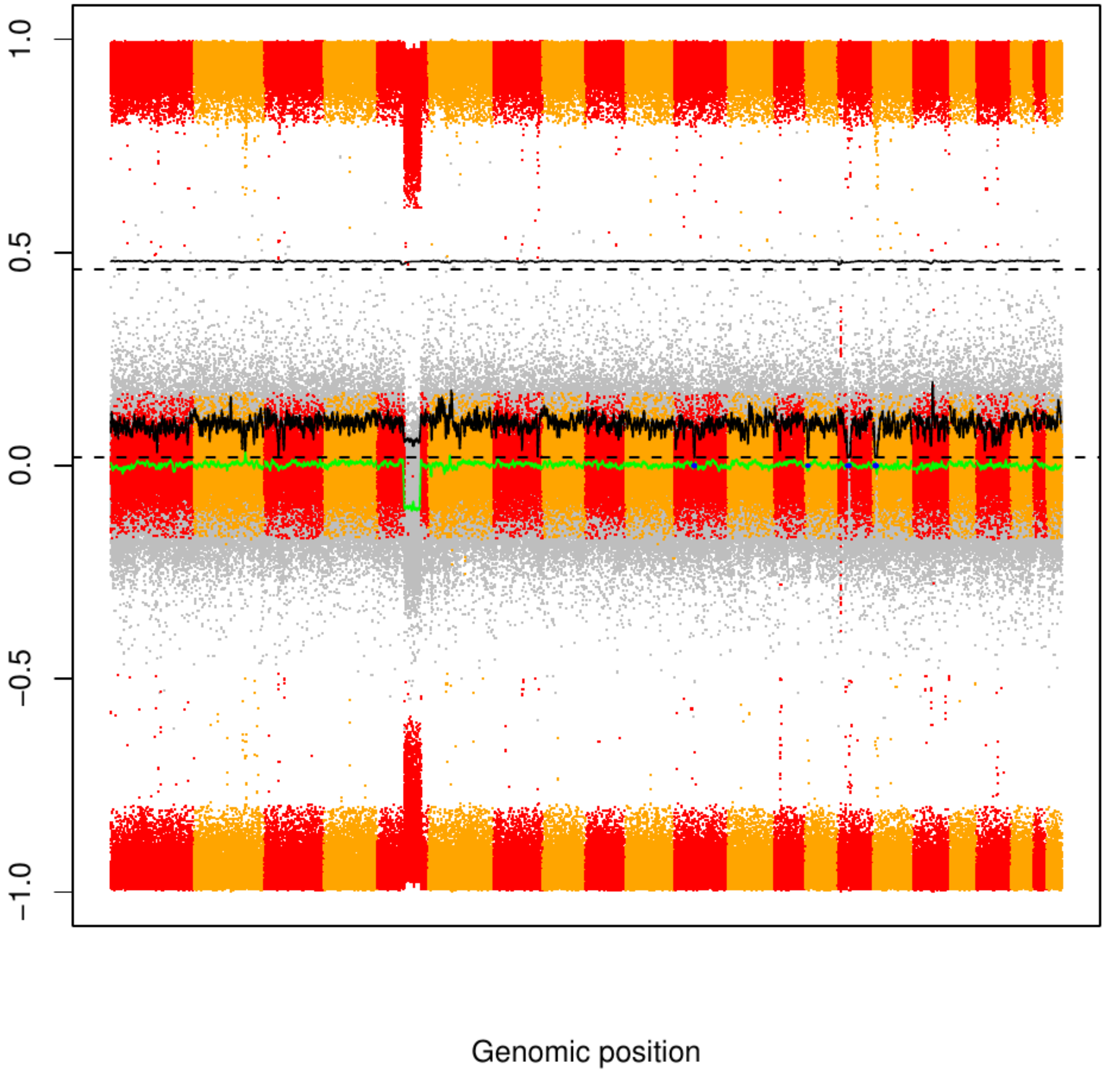

### CME128

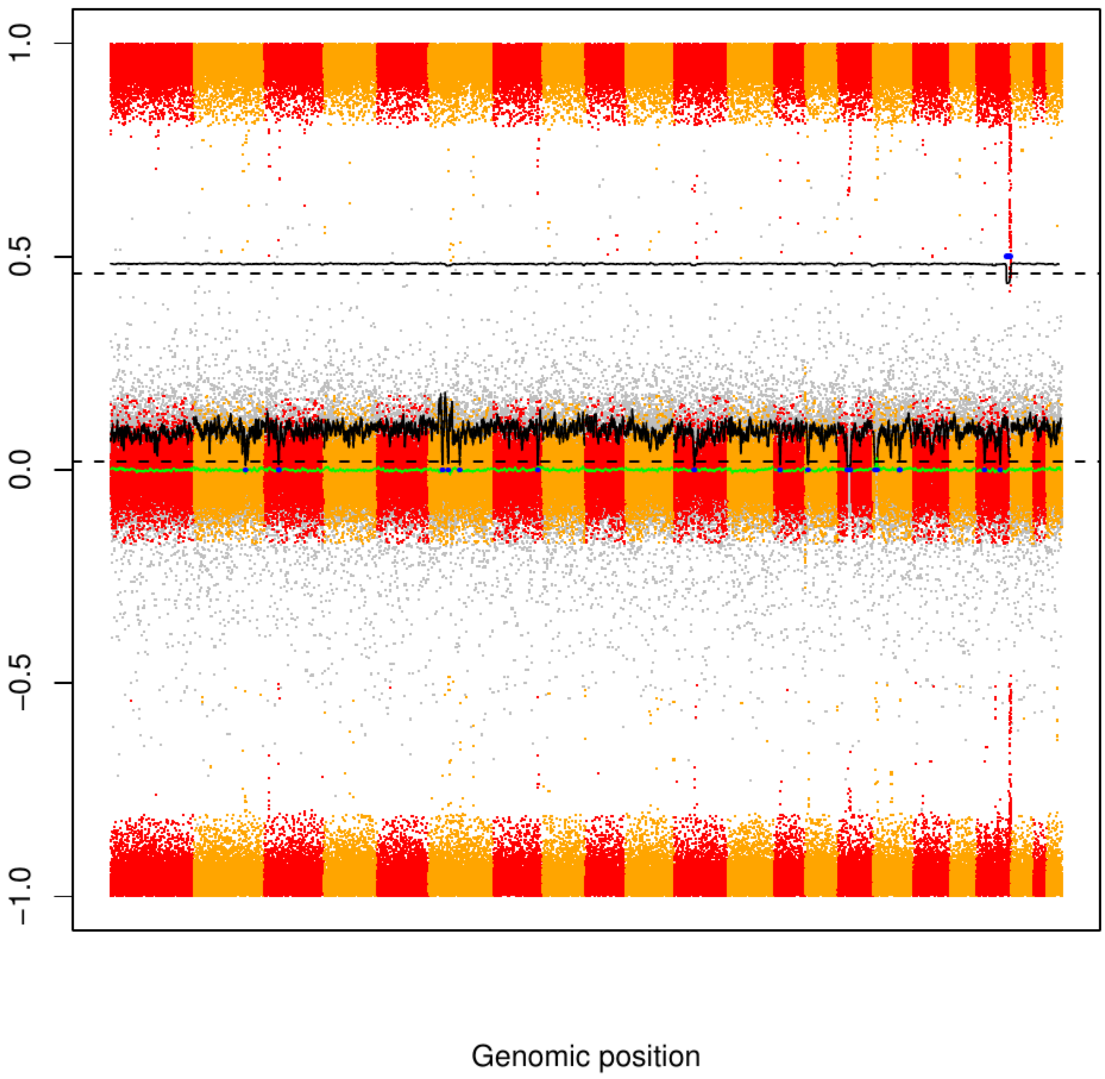

### CME129

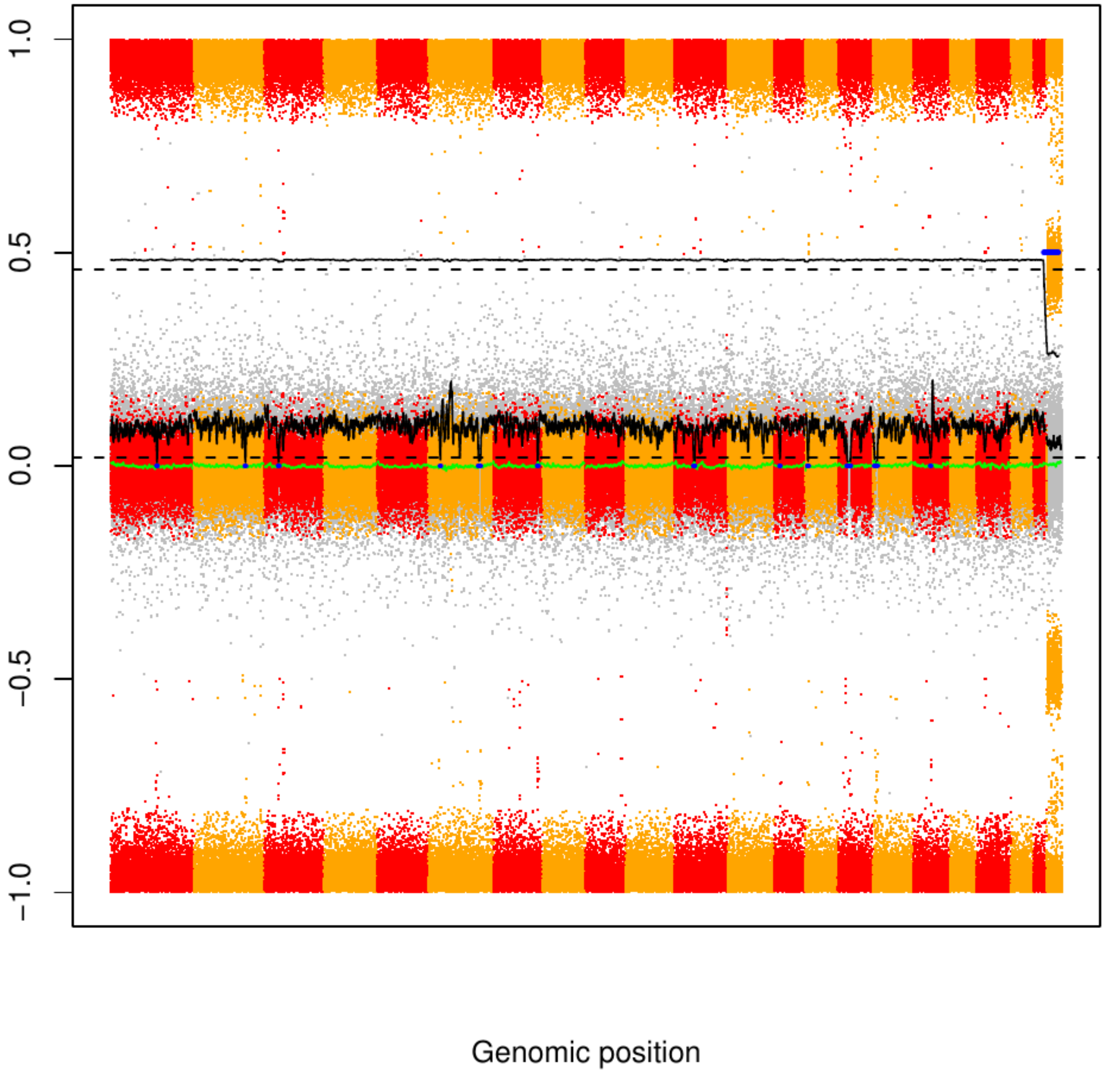

### CME130

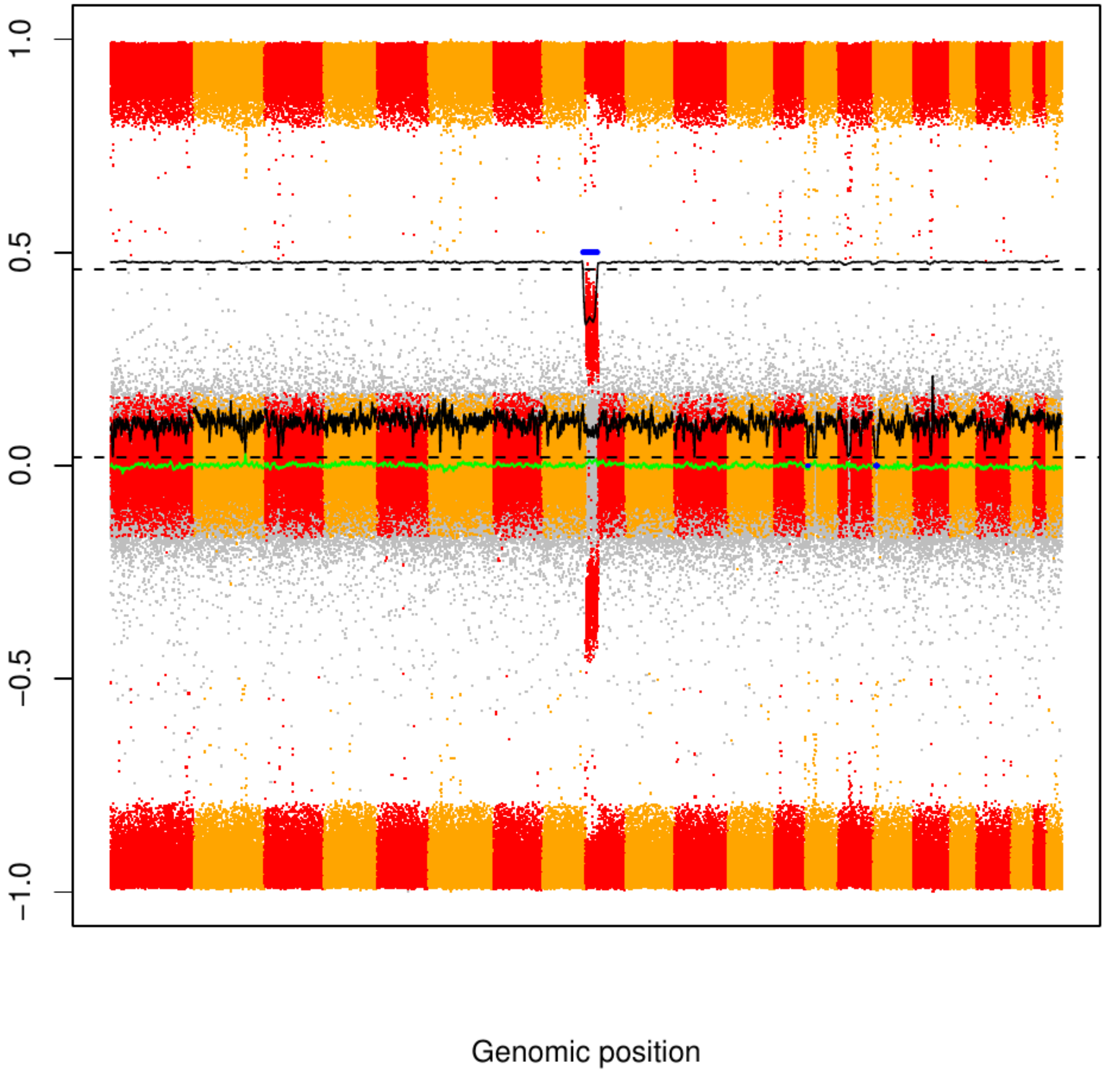

### CME131

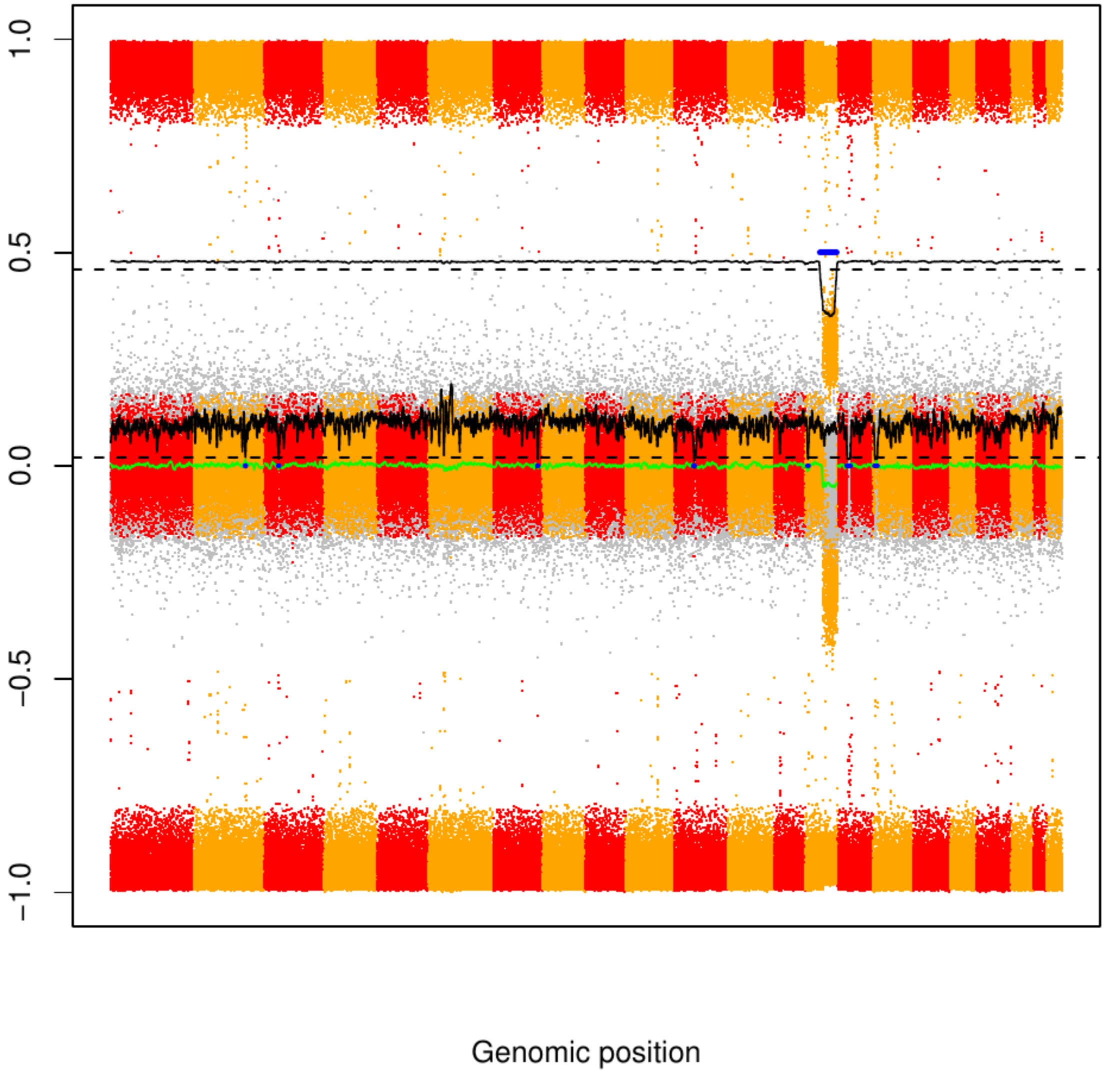

CME132

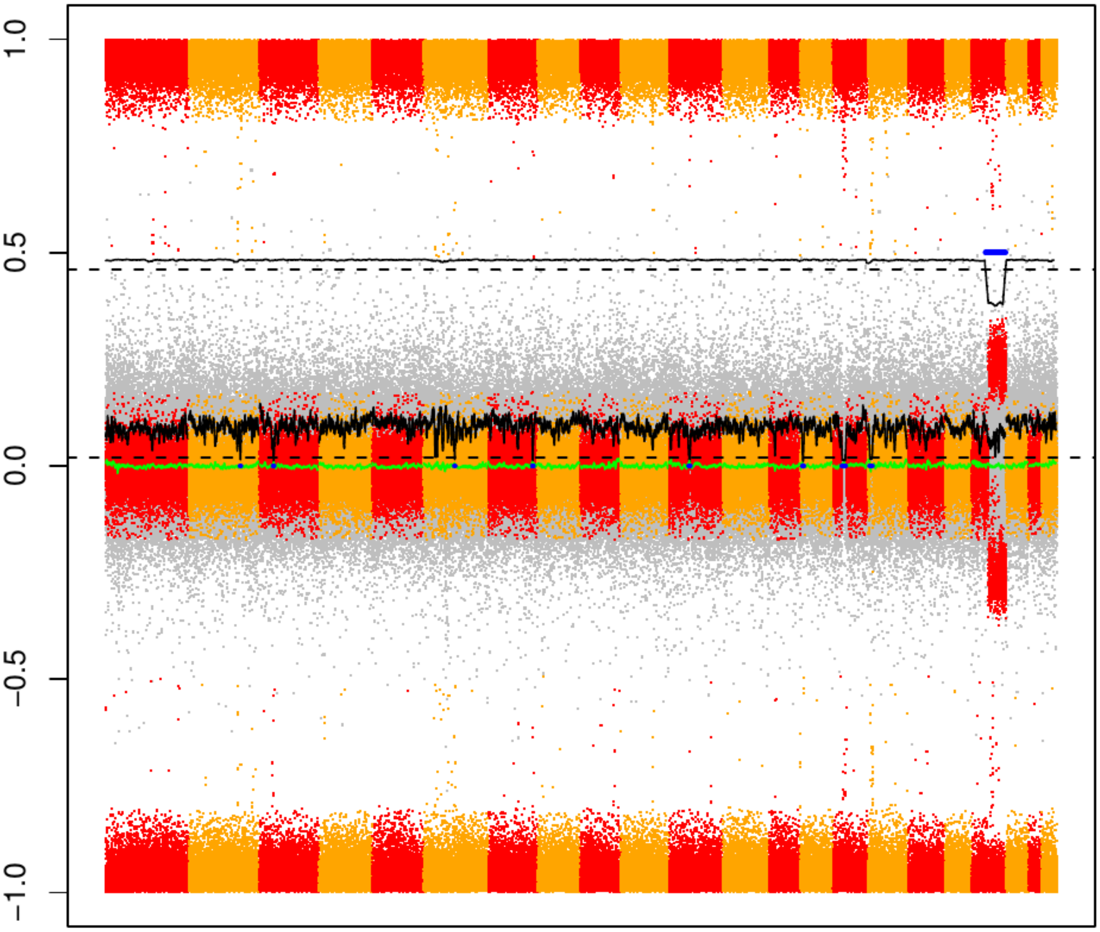

CME133

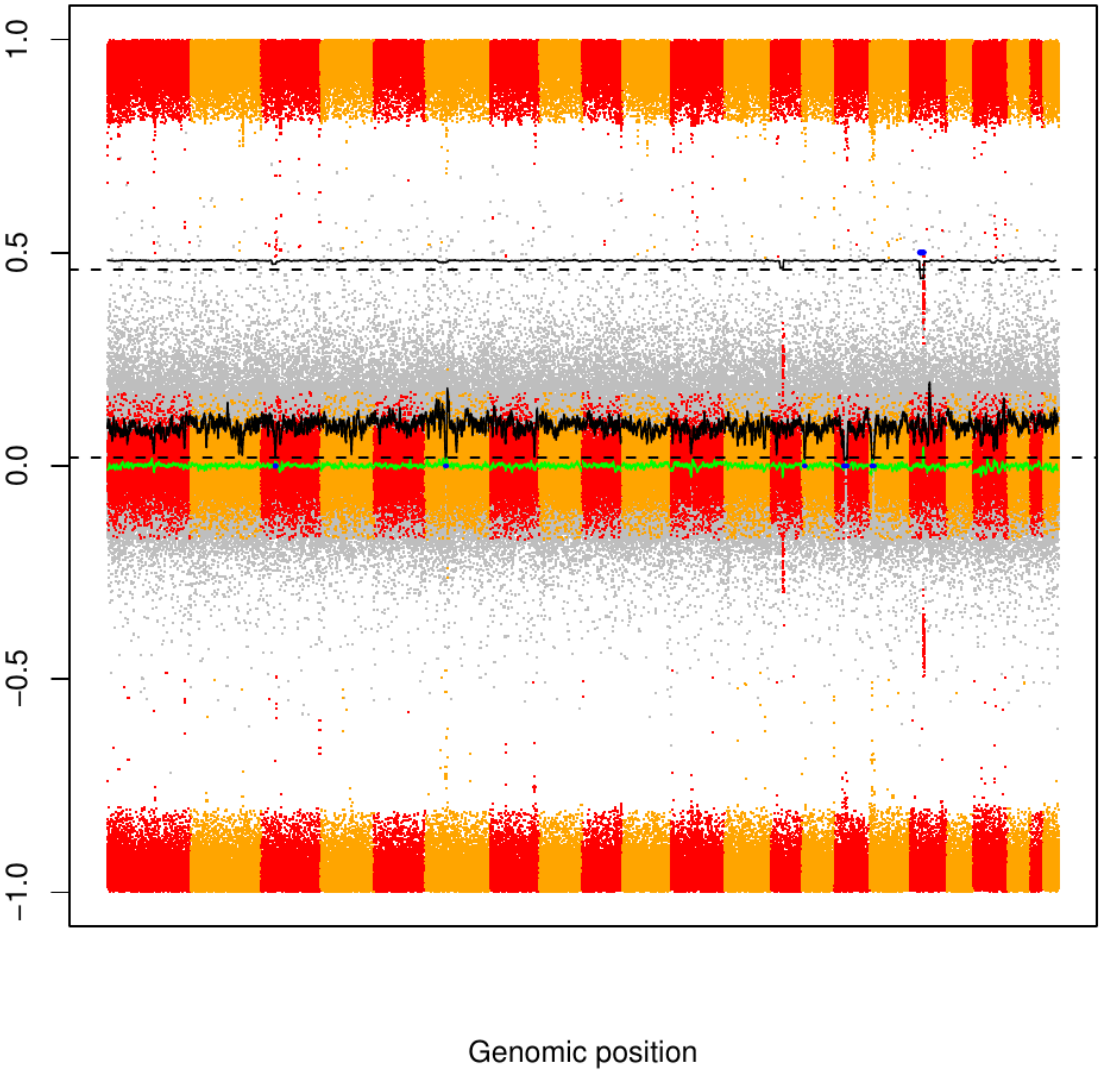
